# Efficacy of *Wolbachia*-mediated sterility to suppress adult *Aedes aegypti* populations

**DOI:** 10.1101/2023.11.24.23298996

**Authors:** Somya Bansal, Chee-Seng Chong, Borame Dickens, Jue Tao Lim, Youming Ng, Lu Deng, Caleb Lee, Li Yun Tan, Evidoxia Kakani, David Du Yu, Grace Chain, Pei Ma, Shuzhen Sim, Lee Ching Ng, Cheong Huat Tan

## Abstract

Incompatible insect technique coupled with sterile insect technique (IIT-SIT) via releases of sterile male *Wolbachia*-infected mosquitoes is a promising tool for dengue control. In a six-year trial from 2016 to 2022, comprising 10.35 km^2^ of high-rise housing estates and 607,872 residents, we designed a synthetic control study methodology to assess the efficacy of IIT-SIT in reducing adult female *Aedes aegypti* populations, using data from a large, routinely collected, nationwide surveillance system of 57,990 unique mosquito traps in public housing estates.

We demonstrated that *Wolbachia*-based IIT-SIT dramatically reduces wildtype *Aedes aegypti* populations by an average of 60.64% (95% CI: 59.59%–61.44%) and 79.37% (95% CI: 78.85%–79.87%) in 3, 6 months of releases and eventually 90.17% (95% CI: 89.92%–90.42%) in 12 and more months of releases. We further found a smaller but non-negligible suppression effect which gradually increased over time (47.91%, 95% CI: 47.29%–48.52%) in adjacent, non-intervention sites. Our results demonstrate the potential of IIT-SIT for strengthening dengue control in tropical cities, where dengue burden is the greatest.

## Introduction

Globally, *Aedes aegypti* is the primary vector responsible for the transmission of arboviruses, such as dengue. The lack of medical countermeasures for most of these arboviruses means that vector control remains the primary measure to stave arboviral transmission. Traditional vector control tools, such as application of insecticides, have drawbacks, such as the development of insecticide resistance and insecticide’s adverse effects on other species. This motivated interest in species-specific vector-control techniques.

Sterile insect technique (SIT) and incompatible insect technique (IIT) are promising approaches for the control of mosquito-borne disease transmission. IIT involves en-masse release of male mosquitoes infected with *Wolbachia* on the field and is expected to suppress wildtype mosquito populations and reduce disease transmission through cytoplasmic incompatibility (CI)^1^. Whereas SIT employs the release of sterile male mosquitoes that mate with wild-type females to prevent reproduction of offspring^2–5^. Both technologies have their drawbacks. SIT technologies have been hampered by their need for irradiation, which may affect male fitness and thus require larger release numbers to compensate for decline in competitiveness. The IIT approach require access to appropriate *Wolbachia-Aedes* strains, and risks abolishment of CI by any establishment of the released *Wolbachia* strain in the field due to imperfect sorting and unintentional releases of females^1,6^.

Specific to the *Ae. aegypti* vector, three recent pilot trials combined IIT with SIT to reduce the likelihood of stable establishment using low-dose irradiation to sterilize residual females during releases of *Wolbachia-*infected males. The IIT-SIT approach suppressed *Ae. aegypti* populations in semi-rural village settings in Thailand^7^, two suburban localities in Mexico^8^ and two high rise residential areas in Singapore^9^. Standalone IIT approaches using high-fidelity sex-sorting methodologies have demonstrated promising results in suburban Northern Australia^10^ and Fresno, California^11^ in short duration trials. Similarly, field trials involving SIT have successfully suppressed mosquito populations in several places, including Brazil^12^, Cuba^13^ and Italy^14^.

Compared to the low-rise, relatively isolated, or semi-rural study sites in the abovementioned pilots, the highly urbanized city-state of Singapore presents a more challenging landscape for IIT-SIT and dengue control in general. The contiguous urban landscape, combined with the tropical climate, is ideal for year-round *Ae. aegypti* breeding^15,16^ Almost 100% of the 5.6 million population lives in urban areas. While the general population density is around 7,688 people per square kilometre^17^ the local density in certain planning areas could be more than 40,000 residents/km^2^ in Singapore^18^. The over 19-fold growth in international visitor arrivals, from <100,000 in 1964 to >19 million in 2019^19^, demonstrated Singapore’s emergence as a regional and global travel hub which further facilitated the introduction and circulation of new *Aedes*-borne arbovirus strains. Decades of effective, integrated vector control, which focuses on source reduction has further lowered population immunity against dengue, leaving the population extremely vulnerable to large dengue outbreaks^15,16,20^.

Extensive field trials of IIT-SIT targeting *Ae. aegypti* have been ongoing in Singapore since 2016. As of Epidemiological Week (EW) 26 2022^21,22^, these field trials cover a total area of 10.35 km^2^ and a population of 607,872. Past phases of the field trials have already delineated the behaviour, efficacy to induce cytoplasmic incompatibility, competitiveness and fitness of *w*AlbB-SG, a localized *Ae. aegypti* line stably infected with *Wolbachia* in field condition^9^. Production and release strategies for SIT-IIT have also been fine-tuned to enable consistent releases over large and vertically distribution spatial scales. Four trial sites have had 6 years of rolling releases over geographically contiguous areas, covering and providing ample data to study its entomological efficacy and behaviour in large-scale field releases.

The primary objectives of our study are two-fold, to (**a**) characterize the efficacy of IIT-SIT interventions in suppressing wild-type *Ae. aegypti* populations (thereafter referred to as intervention efficacy) (**b**) characterize potential spillover and migration effects of IIT-SIT to non-release sites and their corresponding impact on the intervention efficacy of IIT-SIT in actual release sites. This was done by leveraging the large, routinely collected, nationwide surveillance system of 57,990 unique traps in public housing estates designed to capture gravid female *Aedes* mosquitoes, as well as a comprehensive spatio-temporal repository of potential confounders over 2019 to 2022. We took advantage of the staggered adoption of IIT-SIT interventions over 94 release sectors to further understand how duration of releases affected intervention efficacy. The staggered adoption setting also enabled us to understand how the gradual expansion of releases over geographically contiguous areas lead to the mitigation or enhancement of spillover/migration effects over time. By designing a robust synthetic control analysis framework which can account for the staggered, non-randomized nature of IIT-SIT field trials in Singapore, we further enabled appropriate estimation of intervention efficacies and their spillover/migration effects.

As demonstrated below, we observed (**1**) strong suppression of wild-type *Aedes aegypti* populations after three months of intervention in IIT-SIT release sectors, (**2**) drastic improvements in suppression of wild-type *Ae. aegypti* populations in release sectors as treatment time increased, and (**3**) a smaller but significant suppression of wild-type *Ae. aegypti* populations in non-release sites adjacent to release sectors.

## Results

### Baseline characteristics of release, non-release and synthetic control sites

We first compiled and harmonized spatio-temporally resolved mosquito abundance data from 57,990 Gravitraps deployed in all public housing apartment blocks, with a large set of sociodemographic and geographic characteristics. Gravitraps are cylindrical traps designed to catch ovipositing gravid female *Aedes* mosquitoes, and proxies for mosquito abundance. As part of the national Gravitrap surveillance system^23^, Gravitraps were placed in all public housing apartments in Singapore, where 85% of the Singaporean population resides. Data from Gravitraps were aggregated to form the Gravitrap *Ae. aegypti* Index (GAI), which was defined as the mean number of female adult *Ae. aegypti* caught per functional Gravitrap, and therefore adult female *Ae. aegypti* abundance. GAI data from EW8 2019 – EW26 2022 was considered for analysis. We considered sectors to be within the release arm if they ever adopt releases of *w*AlbB-SG males (thereafter referred to as *Wolbachia* releases) in the study period, and non-release sectors, which comprised all non-adjacent sectors which did not adopt *Wolbachia* releases in the study period.

Examination of baseline entomological, sociodemographic and geographic characteristics between release (intervention group) and the non-release (donor group) arms found significant differences in these characteristics in pre-and post-release period. Notably, minimum, mean temperature and maximum, mean windspeed had the most difference (standard mean difference > 0.15) on average in both pre-and post-release period between release and the non-release arm (Table 1). In addition, baseline differences in GAI were found in the pre-release period between the two arms. This motivated the use of synthetic control methods to generate synthetic control sites. Synthetic control sites match GAI and other confounders in the pre-release period between the release and non-release arms – thereby generating appropriate counterfactual sectors to which respective release sectors can be compared against, in the post-release period.

**Table 1.**
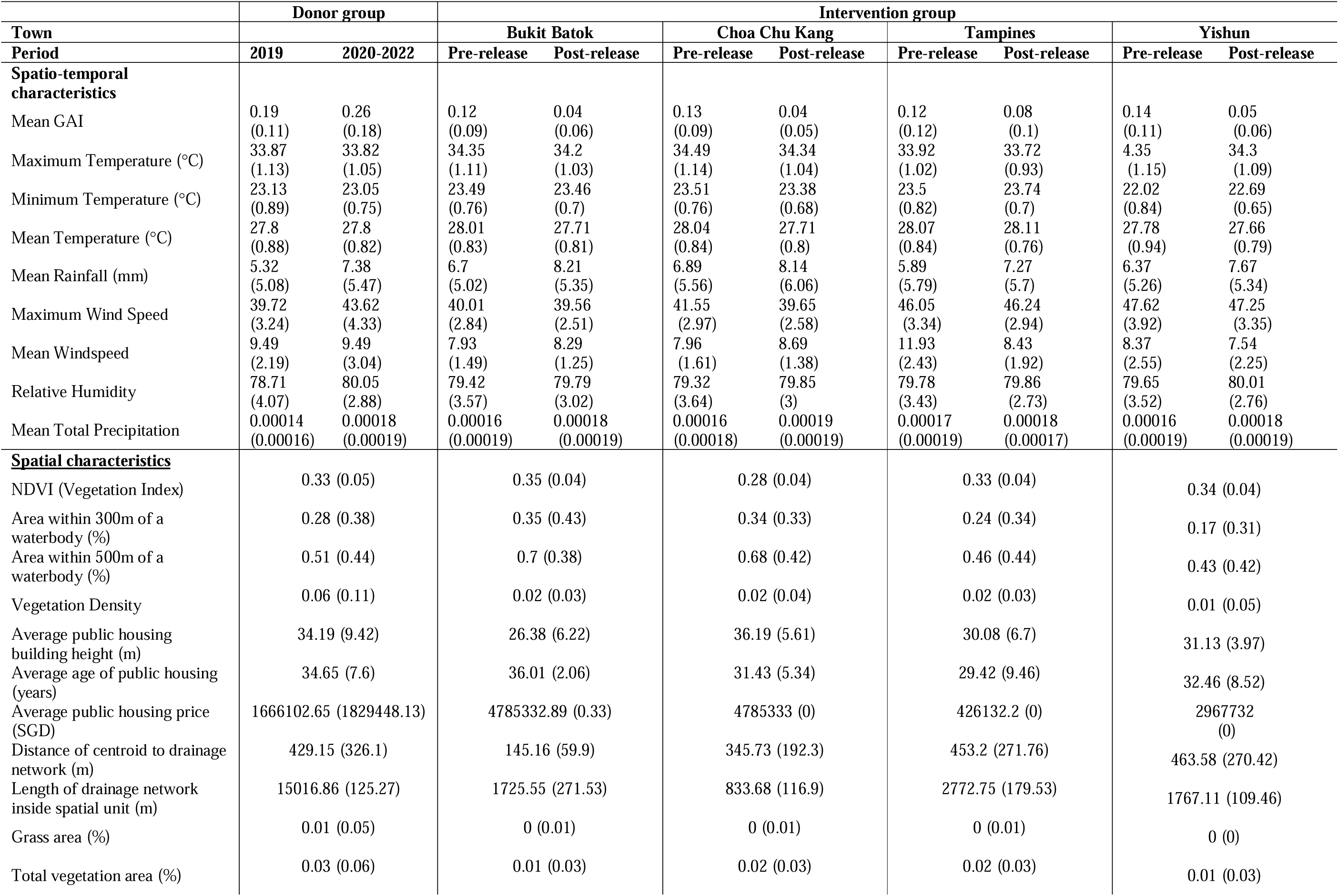
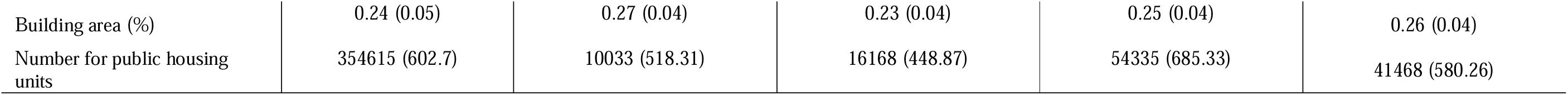
Baseline characteristics of study population pre- and post-*Wolbachia* releases in intervention and donor group. Here, for donor group, the pre-release period is EW8 – 52 2019 while post-release period is EW1 2020 – EW26 2022 as intervention towns experience *Wolbachia* releases from 2020 onwards. For the intervention group, the characteristics are aggregated based on sector-level pre- and post-*Wolbachia* release periods. GAI, rain and relative humidity are averaged across all sectors within intervention town or donor group. The “Minimum” (“Maximum”/“Mean”) characteristics were calculated by taking the minimum (maximum/mean) of the characteristics across all sectors within the respective groups. The spatial characteristics were calculated by averaging across sectors for within respective intervention towns/group. The values in brackets represent standard deviations.

|  | Donor group |  | Intervention group |  |  |  |  |  |  |  |
| --- | --- | --- | --- | --- | --- | --- | --- | --- | --- | --- |
| Town |  |  | Bukit Batok |  | Choa Chu Kang |  | Tampines |  | Yishun |  |
| Period | 2019 | 2020-2022 | Pre-release | Post-release | Pre-release | Post-release | Pre-release | Post-release | Pre-release | Post-release |
| **Spatio-temporal characteristics** |  |  |  |  |  |  |  |  |  |  |
| Mean GAI | 0.19<br>(0.11) | 0.26<br>(0.18) | 0.12<br>(0.09) | 0.04<br>(0.06) | 0.13<br>(0.09) | 0.04<br>(0.05) | 0.12<br>(0.12) | 0.08<br>(0.1) | 0.14<br>(0.11) | 0.05<br>(0.06) |
| Maximum Temperature (°C) | 33.87<br>(1.13) | 33.82<br>(1.05) | 34.35<br>(1.11) | 34.2<br>(1.03) | 34.49<br>(1.14) | 34.34<br>(1.04) | 33.92<br>(1.02) | 33.72<br>(0.93) | 4.35<br>(1.15) | 34.3<br>(1.09) |
| Minimum Temperature (°C) | 23.13<br>(0.89) | 23.05<br>(0.75) | 23.49<br>(0.76) | 23.46<br>(0.7) | 23.51<br>(0.76) | 23.38<br>(0.68) | 23.5<br>(0.82) | 23.74<br>(0.7) | 22.02<br>(0.84) | 22.69<br>(0.65) |
| Mean Temperature (°C) | 27.8<br>(0.88) | 27.8<br>(0.82) | 28.01<br>(0.83) | 27.71<br>(0.81) | 28.04<br>(0.84) | 27.71<br>(0.8) | 28.07<br>(0.84) | 28.11<br>(0.76) | 27.78<br>(0.94) | 27.66<br>(0.79) |
| Mean Rainfall (mm) | 5.32<br>(5.08) | 7.38<br>(5.47) | 6.7<br>(5.02) | 8.21<br>(5.35) | 6.89<br>(5.56) | 8.14<br>(6.06) | 5.89<br>(5.79) | 7.27<br>(5.7) | 6.37<br>(5.26) | 7.67<br>(5.34) |
| Maximum Wind Speed | 39.72<br>(3.24) | 43.62<br>(4.33) | 40.01<br>(2.84) | 39.56<br>(2.51) | 41.55<br>(2.97) | 39.65<br>(2.58) | 46.05<br>(3.34) | 46.24<br>(2.94) | 47.62<br>(3.92) | 47.25<br>(3.35) |
| Mean Windspeed | 9.49<br>(2.19) | 9.49<br>(3.04) | 7.93<br>(1.49) | 8.29<br>(1.25) | 7.96<br>(1.61) | 8.69<br>(1.38) | 11.93<br>(2.43) | 8.43<br>(1.92) | 8.37<br>(2.55) | 7.54<br>(2.25) |
| Relative Humidity | 78.71<br>(4.07) | 80.05<br>(2.88) | 79.42<br>(3.57) | 79.79<br>(3.02) | 79.32<br>(3.64) | 79.85<br>(3) | 79.78<br>(3.43) | 79.86<br>(2.73) | 79.65<br>(3.52) | 80.01<br>(2.76) |
| Mean Total Precipitation | 0.00014<br>(0.00016) | 0.00018<br>(0.00019) | 0.00016<br>(0.00019) | 0.00018<br>(0.00019) | 0.00016<br>(0.00018) | 0.00019<br>(0.00019) | 0.00017<br>(0.00019) | 0.00018<br>(0.00017) | 0.00016<br>(0.00019) | 0.00018<br>(0.00019) |
| **<u>Spatial characteristics</u>** |  |  |  |  |  |  |  |  |  |  |
| NDVI (Vegetation Index) | 0.33 (0.05) |  | 0.35 (0.04) |  | 0.28 (0.04) |  | 0.33 (0.04) |  | 0.34 (0.04) |  |
| Area within 300m of a waterbody (%) | 0.28 (0.38) |  | 0.35 (0.43) |  | 0.34 (0.33) |  | 0.24 (0.34) |  | 0.17 (0.31) |  |
| Area within 500m of a waterbody (%) | 0.51 (0.44) |  | 0.7 (0.38) |  | 0.68 (0.42) |  | 0.46 (0.44) |  | 0.43 (0.42) |  |
| Vegetation Density | 0.06 (0.11) |  | 0.02 (0.03) |  | 0.02 (0.04) |  | 0.02 (0.03) |  | 0.01 (0.05) |  |
| Average public housing building height (m) | 34.19 (9.42) |  | 26.38 (6.22) |  | 36.19 (5.61) |  | 30.08 (6.7) |  | 31.13 (3.97) |  |
| Average age of public housing (years) | 34.65 (7.6) |  | 36.01 (2.06) |  | 31.43 (5.34) |  | 29.42 (9.46) |  | 32.46 (8.52) |  |
| Average public housing price (SGD) | 1666102.65 (1829448.13) |  | 4785332.89 (0.33) |  | 4785333 (0) |  | 426132.2 (0) |  | 2967732 (0) |  |
| Distance of centroid to drainage network (m) | 429.15 (326.1) |  | 145.16 (59.9) |  | 345.73 (192.3) |  | 453.2 (271.76) |  | 463.58 (270.42) |  |
| Length of drainage network inside spatial unit (m) | 15016.86 (125.27) |  | 1725.55 (271.53) |  | 833.68 (116.9) |  | 2772.75 (179.53) |  | 1767.11 (109.46) |  |
| Grass area (%) | 0.01 (0.05) |  | 0 (0.01) |  | 0 (0.01) |  | 0 (0.01) |  | 0 (0) |  |
| Total vegetation area (%) | 0.03 (0.06) |  | 0.01 (0.03) |  | 0.02 (0.03) |  | 0.02 (0.03) |  | 0.01 (0.03) |  |

[**Table 1** Baseline characteristics of study population pre- and post-*Wolbachia* releases in intervention and donor group.
|  |  |  |  |  |  |
| --- | --- | --- | --- | --- | --- |
| Building area (%) | 0.24 (0.05) | 0.27 (0.04) | 0.23 (0.04) | 0.25 (0.04) | 0.26 (0.04) |
| Number for public housing units | 354615 (602.7) | 10033 (518.31) | 16168 (448.87) | 54335 (685.33) | 41468 (580.26) |

[**Table 1** Baseline characteristics of study population pre- and post-*Wolbachia* releases in intervention and donor group. Here, for donor group, the pre-release period is EW8 – 52 2019 while post-release period is EW1 2020 – EW26 2022 as intervention towns experience *Wolbachia* releases from 2020 onwards. For the intervention group, the characteristics are aggregated based on sector-level pre- and post-*Wolbachia* release periods.

GAI, rain and relative humidity are averaged across all sectors within intervention town or donor group. The “Minimum” (“Maximum”/“Mean”) characteristics were calculated by taking the minimum (maximum/mean) of the characteristics across all sectors within the respective groups. The spatial characteristics were calculated by averaging across sectors for within respective intervention towns/group. The values in brackets represent standard deviations.]

### Suppression of wild-type *Ae. aegypti* populations in release sites

Prior to adoption of *Wolbachia* releases, GAI for Bukit Batok, Choa Chu Kang, Yishun and Tampines averaged around 0.12 (Range 0.00 – 0.46), 0.13 (Range 0.00 – 0.65), 0.12 (Range 0.00 – 0.90) and 0.14 (Range 0.00 – 0.69), respectively. At the start of EW27 2018, designated release sites progressively adopted *Wolbachia* releases up until EW26 2022 (Figure 1). Approximately 1–7 *w*AlbB-SG males were released per study site resident per week in these locations. From this point, GAI in release sites plunged dramatically in the following two – four years following releases. On aggregate, town-level GAIs also decreased in conjunction with the increased adoption of *Wolbachia* releases over each township (Figure 1).

**Figure 1:**
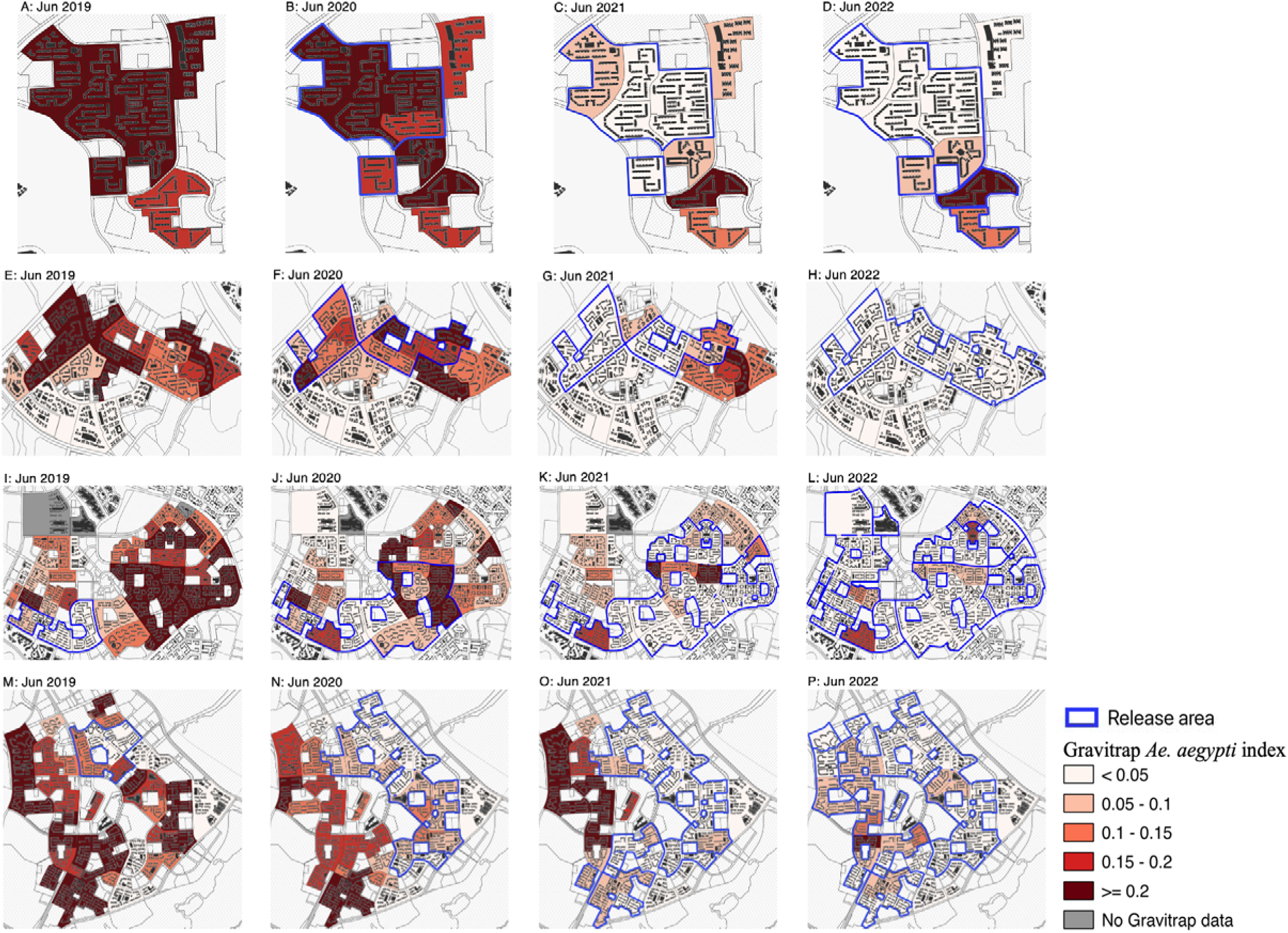
Average Gravitrap *Aedes aegypti* index per sector per year from EW1 2019 to EW26 2022 in **A –D** Bukit Batok, **E–H** Choa Chu Kang, **I–L** Tampines and **M–P** Yishun. Blue outlines represent release areas up to that time point.

### Efficacy of *Wolbachia* releases to suppress wild-type *Ae. aegypti* populations

To provide appropriate estimates of intervention efficacy in each release site, we developed a synthetic control framework to generating counterfactual sites to which release sites can be compared against in the post-release period.

As the effect of *Wolbachia* releases on GAI may vary with sites and time-since-release, we computed four separate measures of intervention efficacies. These were defined to be: *IE_total_*, the total percentage reduction in GAI across a specific time period, *IE_release_*, the percentage reduction in GAI by time-spent on release for release sector and *IE_sector_* the total percentage reduction in GAI in each sector. We also looked at intervention efficacies by calendar time (*IE_calendar_*) to examine whether there were year-on-year changes in intervention effects. All measures compared GAI reduction in release versus synthetic control sectors. Previous field trials also demonstrated the potential for wild-type *Ae. aegypti* migration from non-release locations to attenuate the potential upper limit of *Wolbachia* suppression in release sites. We therefore considered intervention efficacies based on separate classes of release sites to better examine this effect. Namely, buffer sectors which share borders with non-release sectors, and core sectors which do not share borders with non-release sectors.

Three months post-release, *IE_release_* was estimated to be around 59.61% (95% CI: 58.67%–60.50%). *IE_release_* dramatically improved and was maintained at a statistically significant 78.92% (95% CI: 78.41%–79.41%) six months post-release. This may be due to time required and multiple releases needed to achieve adequate suppression in each release location. Subsetting *IE_release_* to only comprise core or buffer sectors demonstrated a marginally larger intervention effect in core sectors surrounded by buffer sectors, especially when interventions just begun. Core sectors had *IE_release_* which ranged from 26.94% (95% CI: 25.77%–28.09%) to 80.28% (95% CI: 79.87%– 80.67%) in the first six months period of release and followed the same trend in improving IEs to 84.92% (95% CI: 84.46%–85.36%) six months post-release, in comparison to buffer sectors which had slightly lower intervention efficacies estimated in the same period (Figure 2). This suggests that core sectors surrounded by buffer sectors can maintain more effective suppression versus buffer sectors – possibly due to buffer sectors being able to counter wild-type female *Ae. aegypti* migration from adjacent non-release sectors.

**Figure 2:**
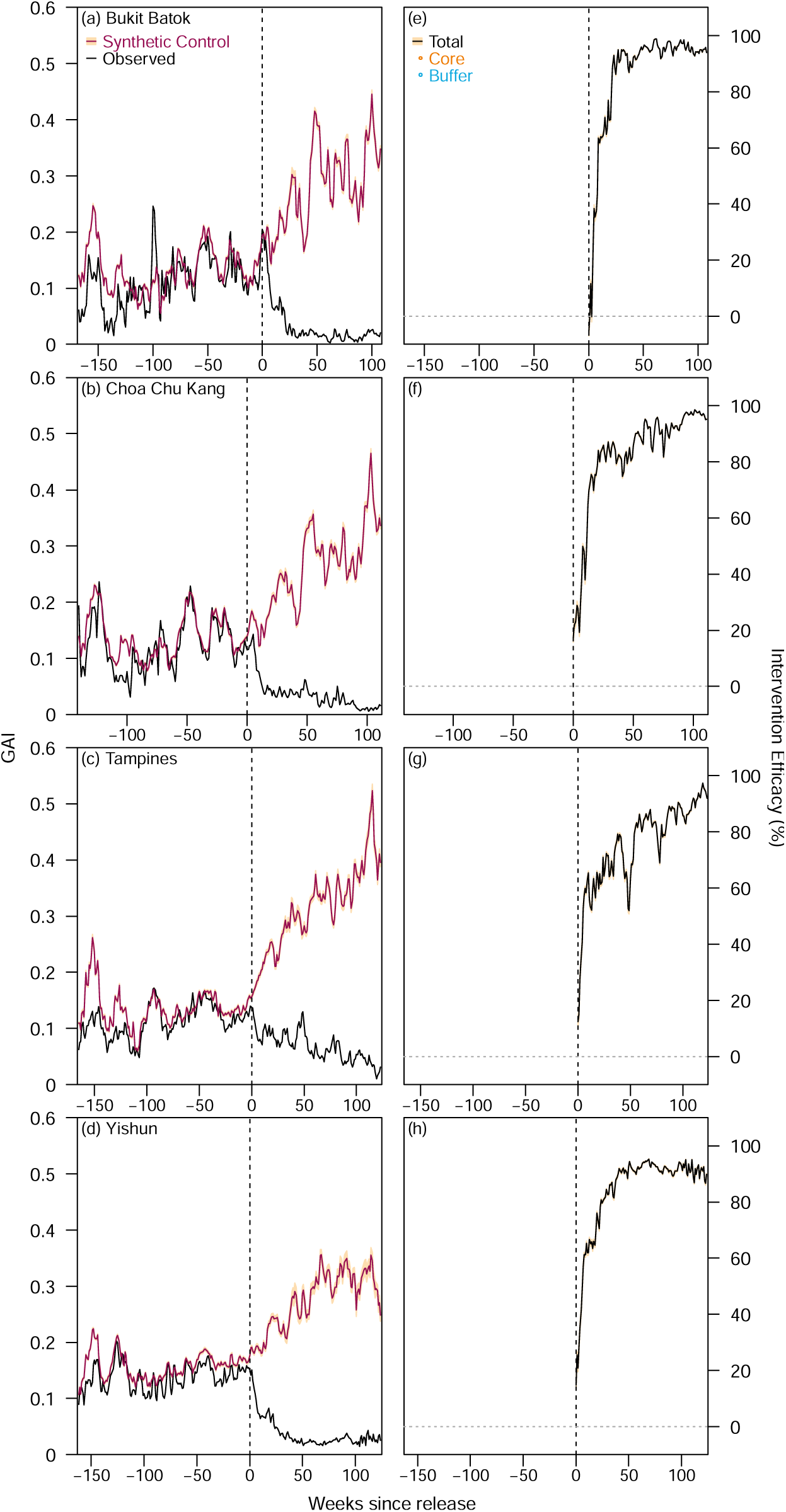
GAI for synthetic control and intervention sectors for (**a**) Bukit Batok (**b**) Choa Chu Kang (**c**) Tampines (**d**) Yishun. Intervention efficacies expressed as the percentage difference in GAI between synthetic control and intervention sectors for each week in post intervention time for (**e**) Bukit Batok (**f**) Choa Chu Kang (**g**) Tampines (**h**) Yishun.

Taken together, we estimated *IE_total_* to be 60.64% (95% CI: 59.59%–61.44%) and 79.37% (95% CI: 78.85%–79.87%) in the three and six months, respectively, and eventually 90.17% (95% CI: 89.92% – 90.42%) a year to end of study period. This highlights the need for around six months for mosquito suppression at the threshold of 80% to take place.

By calendar time, we note that *IE_year_* improved and was maintained since EW1 2021, indicating that intervention efficacies could be consistent despite any potential year-on-year changes in mosquito breeding patterns and/or abundance (Figure 3). In totality, we found an overall intervention efficacy of 70.27% (95% CI: 69.36% – 71.13%), with a similar value being noted in buffer sectors 70.57% (95% CI: 69.76% – 71.32%) and slightly higher in core sectors 74.20% (95% CI: 73.20% – 75.12%) by calendar time.

**Figure 3:**
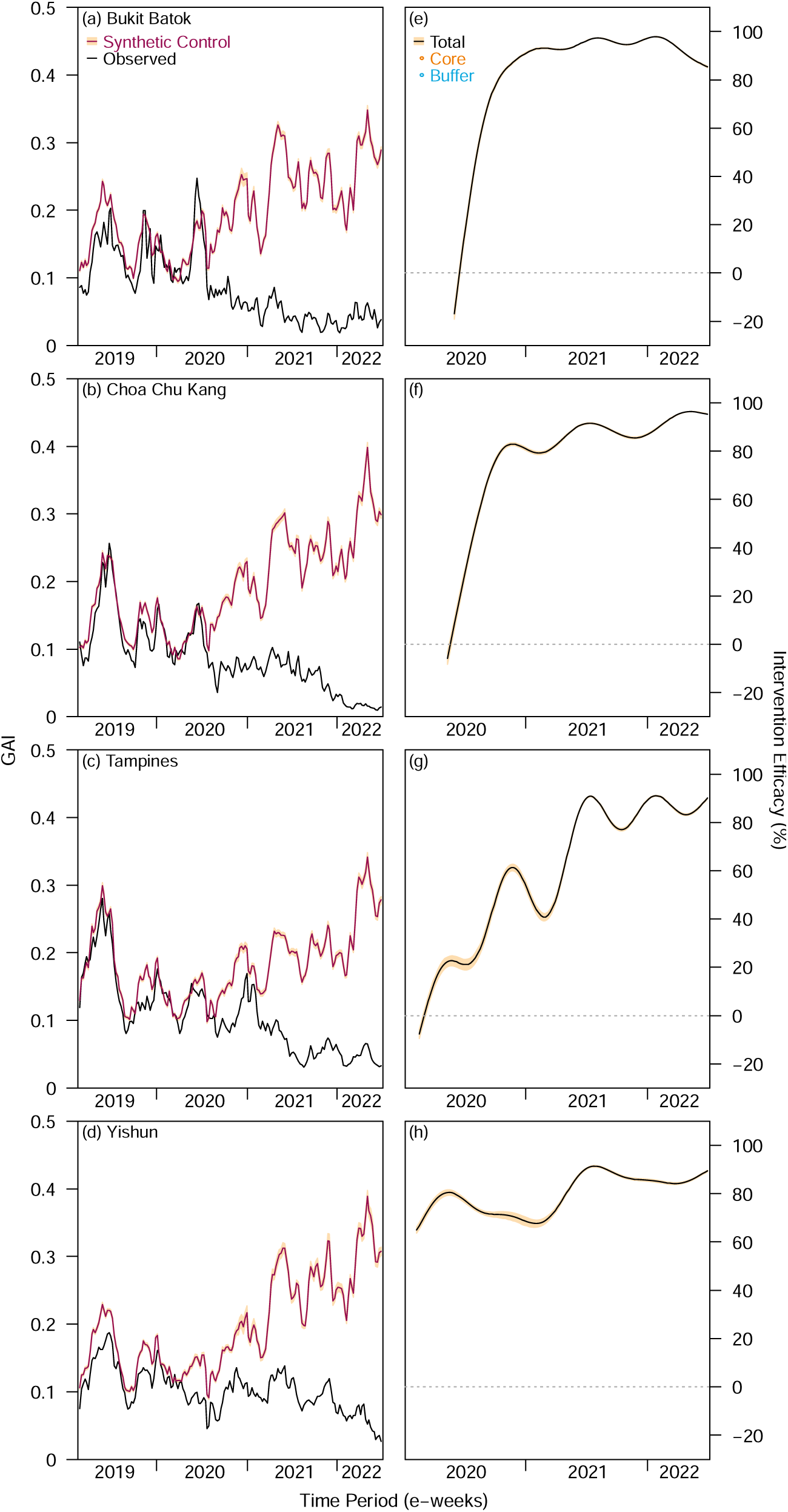
GAI for synthetic control and intervention sectors for ((**a**) Bukit Batok (**b**) Choa Chu Kang (**c**) Tampines (**d**) Yishun. Intervention efficacies expressed as the percentage difference in GAI between synthetic control and intervention sectors for each week in calendar time for (**e**) Bukit Batok (**f**) Choa Chu Kang (**g**) Tampines (**h**) Yishun.

Concomitant with the decrease in overall GAI across each town as *Wolbachia* releases were employed, sector specific intervention efficacies by event time similarly demonstrated the same pattern. To elaborate, intervention efficacies in the 1–6 months after release began ranged from - 8.39%–93.14% in each sector per town (Figure 4, A, F, K, P) but increased gradually to 10.34%– 99.44% 13 –18 months after releases (Figure 4, C, H, M, R) and finally to 78.03%–99.30% two or more years after releases (Figure 4, E, J, O, T). These effects were consistent across each township, and within each township. In sectors which were at the edge of townships and buffered by natural border/release sectors, intervention effects also appeared to be higher (Figure 4, A, F).

**Figure 4:**
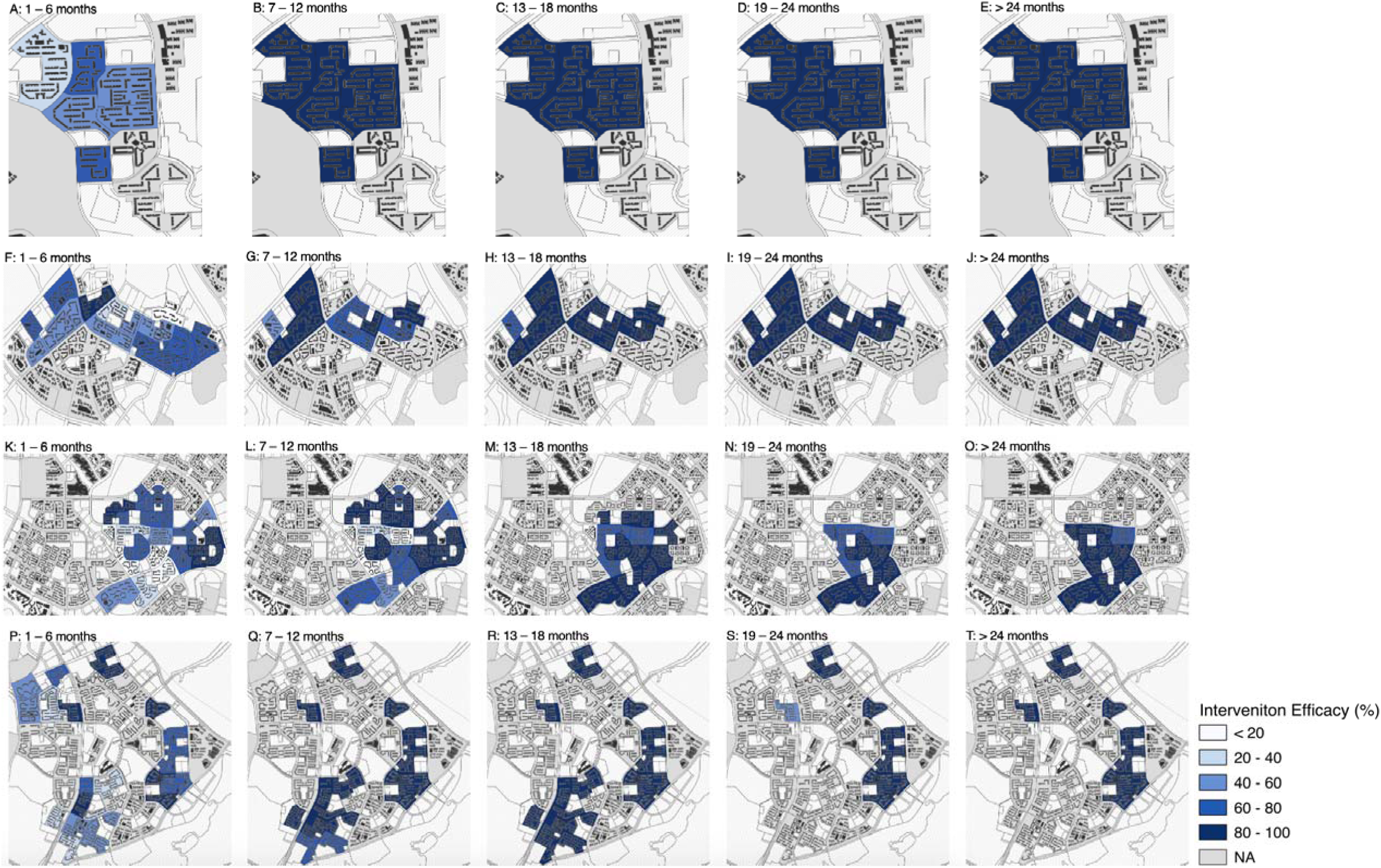
Intervention efficacies per sector per town by event time in **A**–**E**: Bukit Batok**, F – J**: Choa Chu Kang**, K – O**: Tampines, **P – T**: Yishun. Darker blue shades represent higher intervention efficacies. Unshaded areas in grey refer to locations which intervention efficacies cannot be estimated due to lack of pre-intervention observations to match intervention and donor pool trends to, or have less than the observed length of post-intervention time to estimate intervention efficacies by event time.

As interventions elapsed in each township, we also noticed a pattern of higher intervention efficacies for each sector. This pattern was apparent in Choa Chu Kang (Figure 5, D – F), Tampines (Figure 5, G – I) and Yishun (Figure 5, J – L) townships. In Choa Chu Kang township, the range of average intervention efficacies by calendar time increased from 58.79%–96.92% (Figure 5D) to 82.09%– 98.99% (Figure F) from EW1–26 2021 to EW1–26 2022, respectively. Whereas in Yishun township the range of average intervention efficacies by calendar time increased from 39.19%–95.61% (Figure 5G) to 15.64%–99.64% (Figure 5I) from EW1–26 2021 to EW1–26 2022, respectively. Notably, we found that intervention efficacies were high and at an average of 63.53% even for sectors where interventions were only adopted interventions for less than six months in EW27 2021 to EW26 2022. In these periods, 57.41%–72.95% of release coverage was achieved in each of these townships compared to only 41.04% – 72.95% in EW1–26 in 2021. This suggests that the efficacy of *Wolbachia* interventions in areas with less than six months of adoption time can be improved as overall coverage in each geographically contiguous area increased. Crucially, examination of intervention efficacies by both event time (Figure 4 above) and calendar time (Figure 5 below) in sectors revealed the interplay between the temporal and spatial components of *Wolbachia* interventions, where time required to effect considerable suppression to adult mosquito populations is positively influenced by the coverage of interventions in that location.

**Figure 5:**
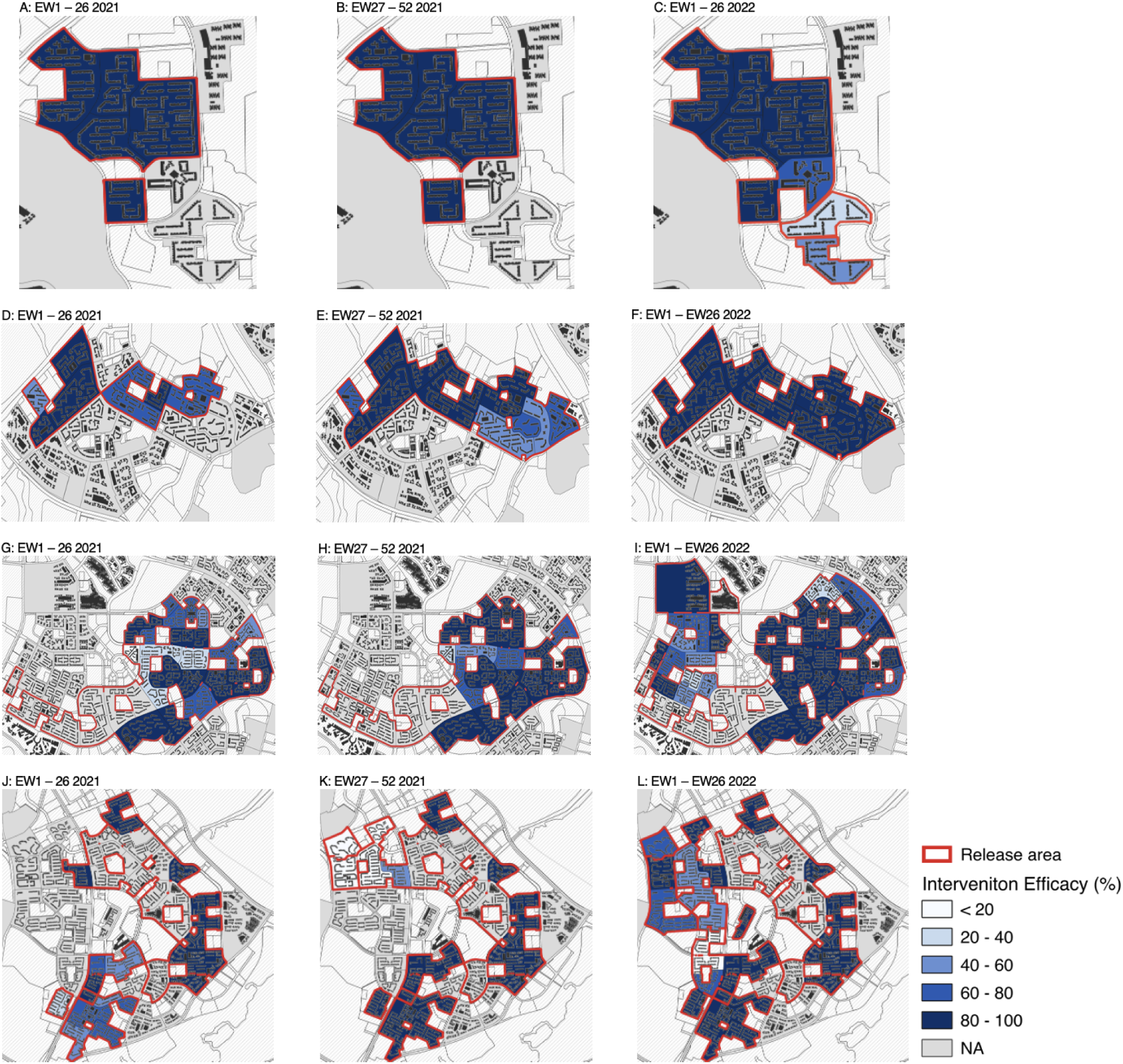
intervention efficacies per sector per town by calendar time in **A–C**: Bukit Batok, **D – F**: Choa Chu Kang, **G – I**: Tampines, **J – K**: Yishun. Darker blue shades represent higher intervention efficacies. Unshaded areas in grey refer to locations which intervention efficacies cannot be estimated due to lack of pre-intervention observations to match intervention and donor pool trends to, or did not implement interventions by that calendar date.

### Spillover and migration impacts of *Wolbachia* interventions

Motivated by our initial findings on the difference in intervention efficacies between core and buffer sectors, we examined whether there were any (**1**) spillover suppression effects on wild-type *Ae. aegypti* to surrounding sectors and (**2**) whether migration was affecting suppression effects on wild-type *Ae. aegypti* in buffer sectors, by examining the differences in GAI between traps close to edges versus those in the interior.

We re-employed the synthetic control method taking non-release sectors adjacent to buffer sectors as the intervention arm. The donor pool to generate the synthetic controls were similarly taken as control sectors in other townships which were never subject to *Wolbachia* interventions in the study period. Some non-release sectors gradually adopted releases and we only considered the period where they were not subjected to release to compute intervention efficacies. We found a small but non-negligible suppression effect which gradually increased over calendar time, which ranged from - 25.37% – 83.70% over EW3 2020 to EW26 2022 (Figure 6 below). In totality, over the release period, a spill-over intervention efficacy of 40.97% (95% CI: 39.57% – 42.30%) was estimated. Our estimates suggested that immigration of *w*AlbB-SG males from buffer sectors to adjacent non-release sectors may have led to suppression of wild-type female *Ae. aegypti* in the latter.

**Figure 6:**
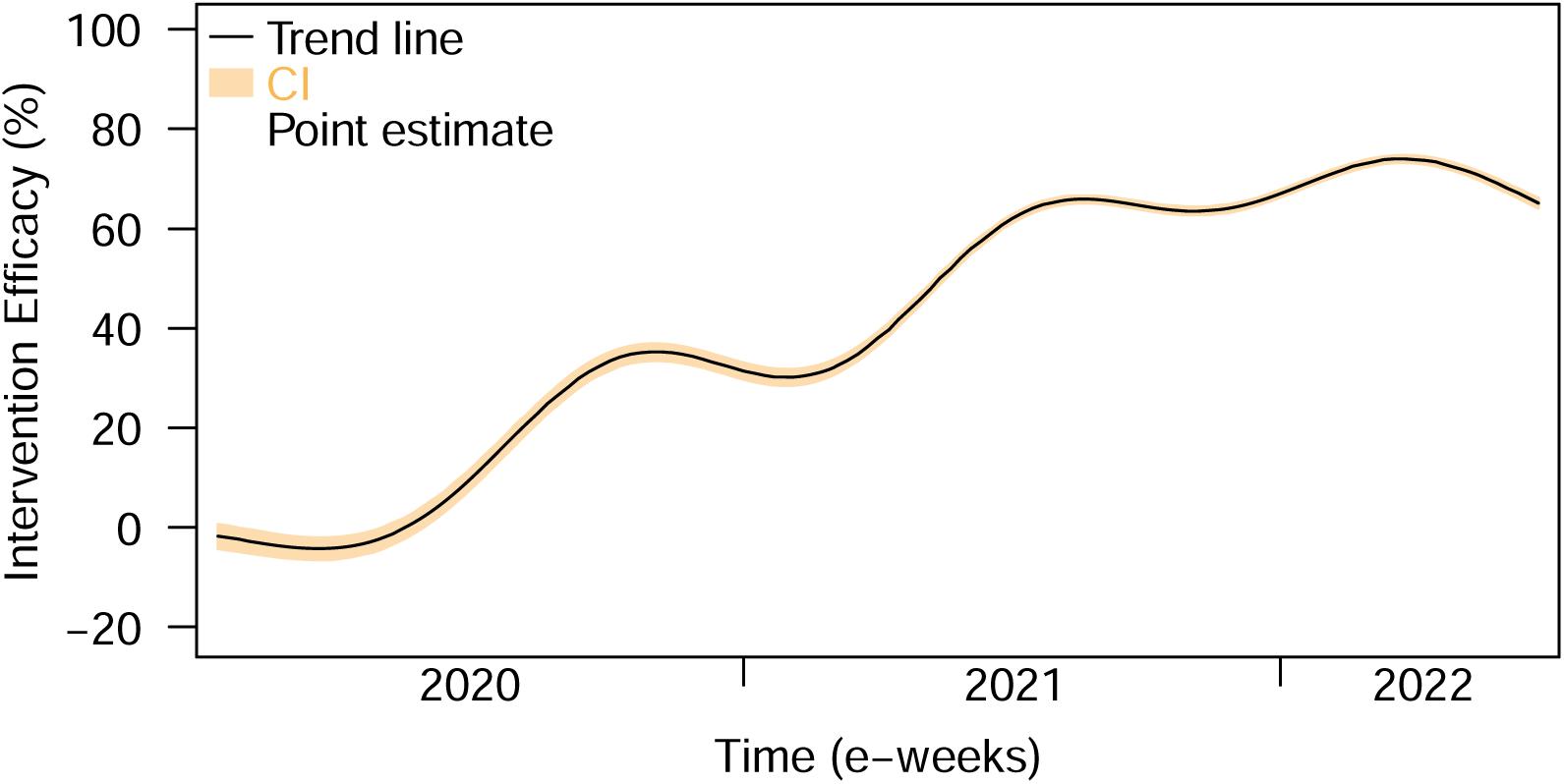
Estimated spillover intervention efficacies to non-release adjacent sectors from EW3 2020 to EW26 2022. For each time point, only sites which were non-release adjacent sectors at the same time point contributed to spillover intervention efficacy estimates.

Next, we examined whether edge effects were detected in buffer sectors. For traps in buffer sectors, we examined the overall GAI in these buffer sectors and if there was any correlation between individual trap GAI and their distance to the closest non-release adjacent sectors. Analysis indicated that individual level GAI values were found to be higher when they were closer to non-release adjacent sectors (Figure 7A). Additionally, we found over the course of releases that there was a significant, negative correlation between GAI and the closest distance to a non-release adjacent sector (Figure 7B). These findings suggest that within-site heterogeneity in GAI may be due to female mosquito migration from non-release sectors to buffer sectors and consequentially attenuate the upper limit of possible suppression in these locations.

**Figure 7:**
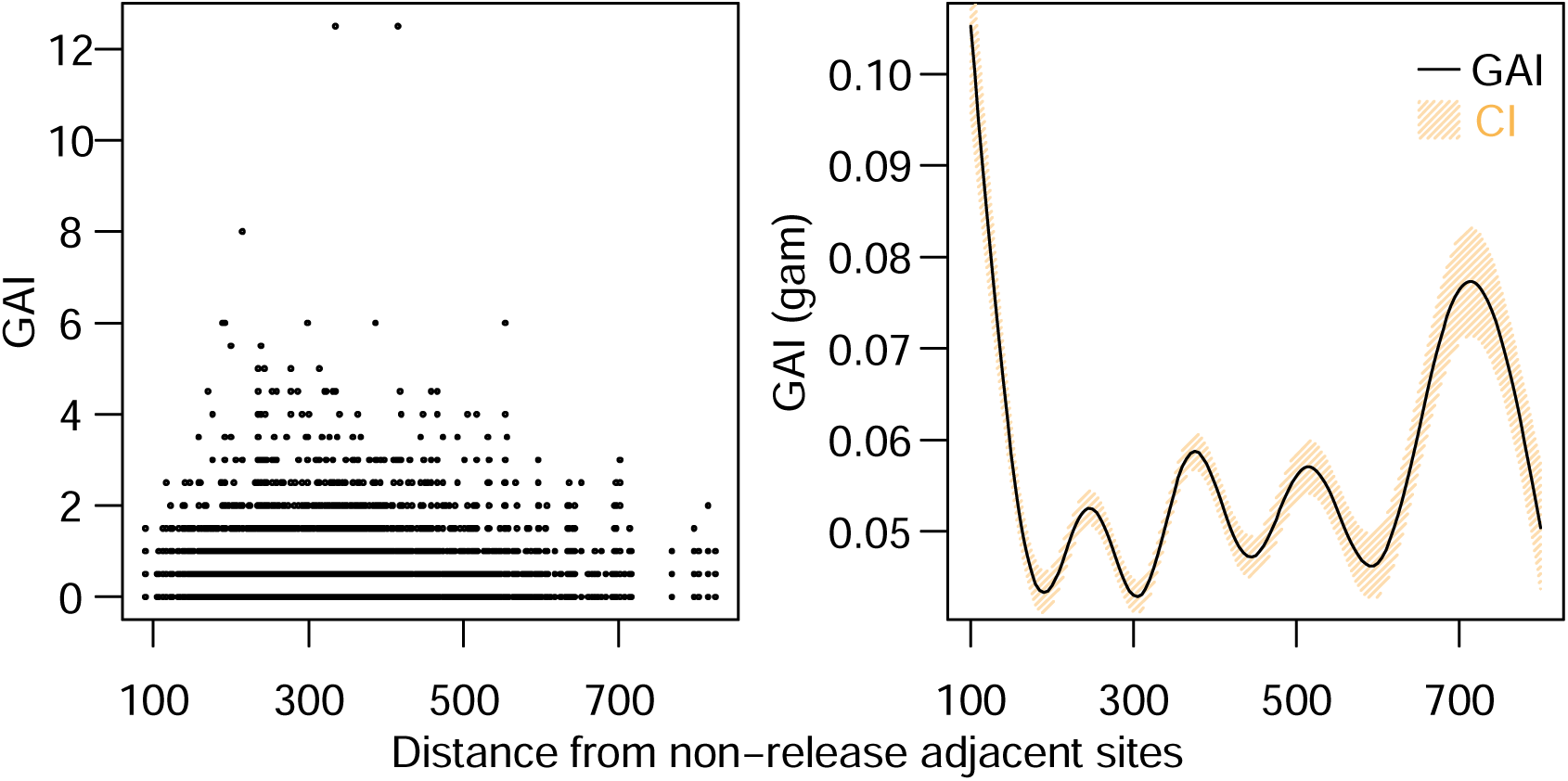
(**A**) Observed and (**B**) generalized additive model (GAM) predicted individual trap-level GAI located within buffer sectors plotted against the trap’s distance to the closest non-release adjacent sector boundary.

### Impact of *Wolbachia* interventions on adult *Aedes albopictus* populations

We constructed synthetic controls on all the release sectors with female adult *Ae. albopictus* abundance (*GAI_albo_*) taken as the outcome of interest using the best model. The average intervention efficacy across all release sectors was −19.63% (95% CI: −22.56% – −16.84%), indicating a small but non-negligible negative intervention effect of *Wolbachia* release on *Ae. albopictus* abundance with the intervention efficacy following a decreasing trend with respect to weeks post release (Figure 8). We however note noisy estimates from 120 weeks of post-release time, due to the small number of study sectors available here for analysis (N=12 of 94 study sectors).

**Figure 8:**
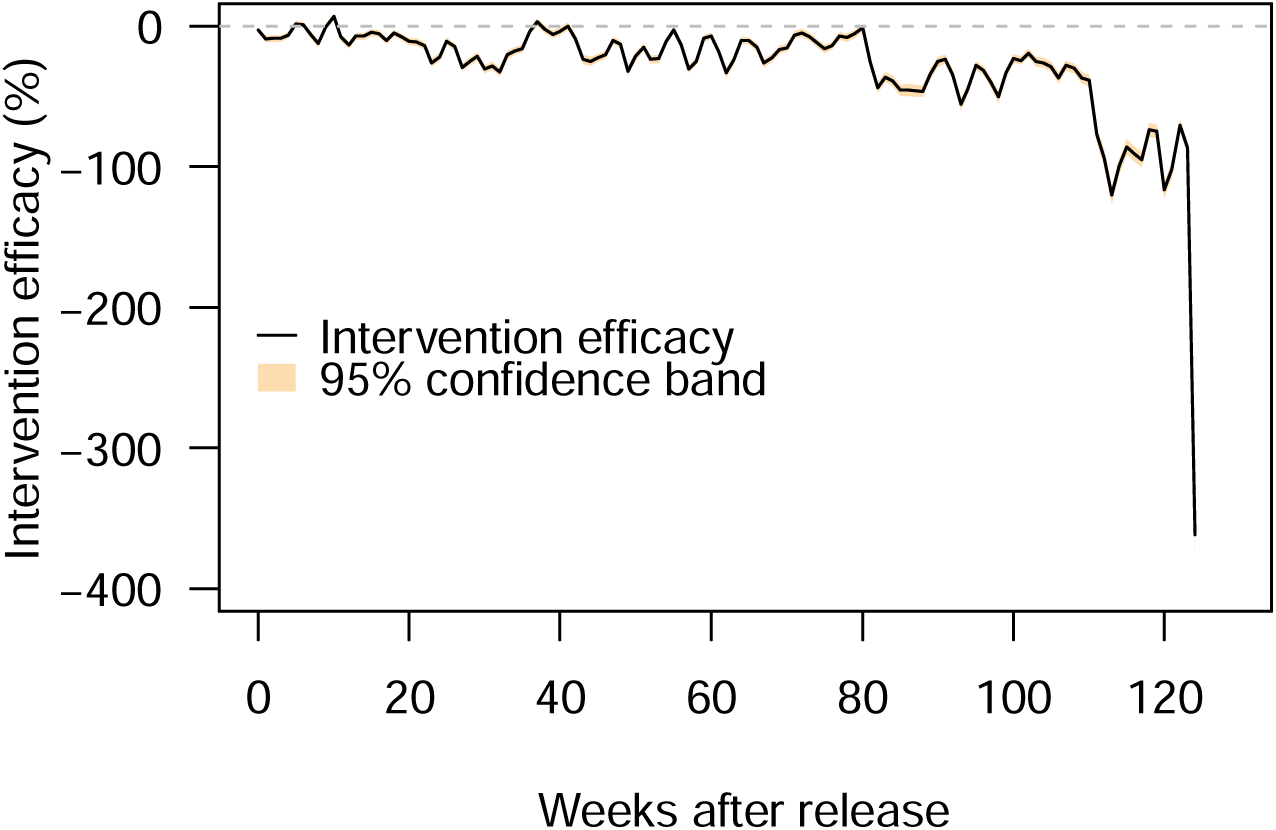
Overall intervention efficacy across all release sectors, expressed as the percentage difference in *GAI_albo_* between synthetic control and intervention sectors for each week in post intervention time.

By township, Figure 9 showed that for Bukit Batok and Tampines, *Wolbachia* release had little to no effect on *GAI_albo_* with average intervention efficacies of 1.99% (95% CI: −0.19% – 4 .09%) and 11.56% (95% CI: 9.71% - 13.32%), respectively. In contrast, an increase in *GAI_albo_* in Choa Chu Kang and Yishun, with an intervention efficacy −33.66% (95% CI: −36.92% – −30.56%) and −57.87% (95% CI: −62.57% – −53.44%), respectively was observed. While previous studies characterising factors associated to higher *Ae. albopictus* abundance in the study setting^24^ have suggested the important role of vegetation in influencing *Ae. albopictus* abundance, we found no drastic site-specific differences in these factors between all four intervention townships (Table 1).

**Figure 9:**
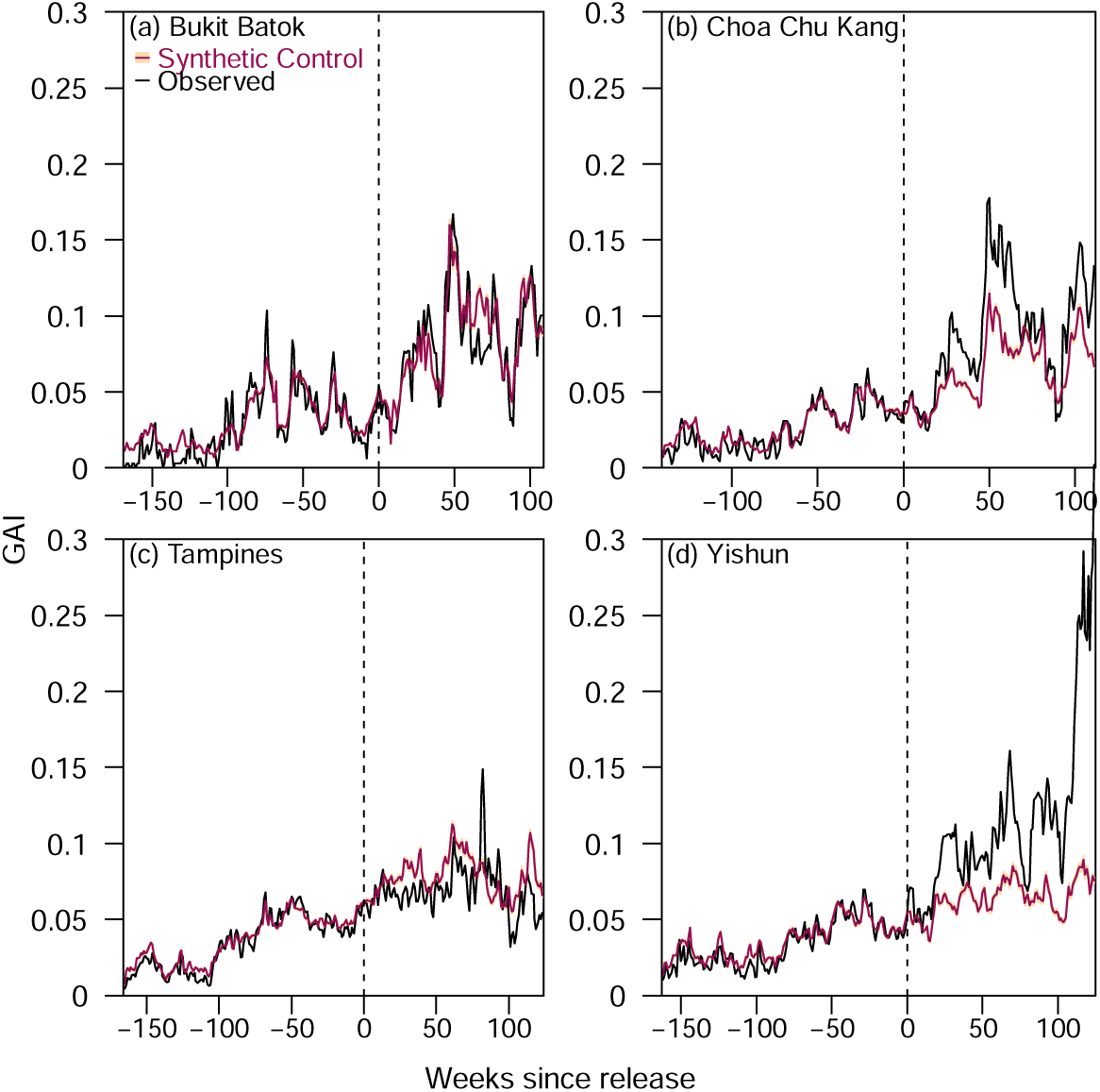
*GAI_albo_* for synthetic control and intervention sectors for (**a**) Bukit Batok, (**b**) Choa Chu Kang, (**c**) Tampines and (**d**) Yishun.

### Robustness checks

We ran several sensitivity analyses to ensure the validity of intervention efficacy estimates for *Wolbachia* interventions to suppress *Ae. aegypti* populations under our synthetic control framework. We found minimal pre-trends in GAI by testing for placebo effects in intervention sectors before *Wolbachia* releases occurred. This test, alongside a cross-validation procedure by splitting the preintervention time period of release sectors into training and validation periods, also showed that synthetic control methods reproduced the original observations in the pre-intervention period. The average root mean squared prediction error (RMSPE) ranged from 0.08 to 0.10 across the intervention group for 1 to 10 weeks before the actual intervention occurred, with RMSPE increasing with larger validation periods due to shorter training periods. This provided clear support that our estimated intervention effects were not estimated due to poor fit to the pre-release period.

Additionally, we conducted placebo tests where we iteratively took each control sector as the placebo-release sector and constructed respective synthetic controls using the remaining donor pool while excluding actual release sectors and examined the gap between synthetic control and observed GAI. If this placebo test showed that the gap estimated for release sector is unusually large relative to control sector, then the hypothesis that *Wolbachia* releases led to significant reductions in GAI is further supported. To conduct this placebo test, we examined the distribution of pre- and post-*Wolbachia* RMSPE for control and intervention sites (Figure 10), and noted that RMSPE measures the magnitude of the gap in GAI between each sector and its respective synthetic control^25^. Comparing iteratively the ratios of post versus pre-intervention RMPSEs between intervention sectors and the placebo-intervention sectors showed that the probability of obtaining a ratio which was as high as the actual release sectors, if we randomly assign a site as an intervention site, is 1/292 = 0.003. This supported the hypothesis that *Wolbachia* releases have led to significant reductions in *Ae. aegypti* populations.

**Figure 10:**
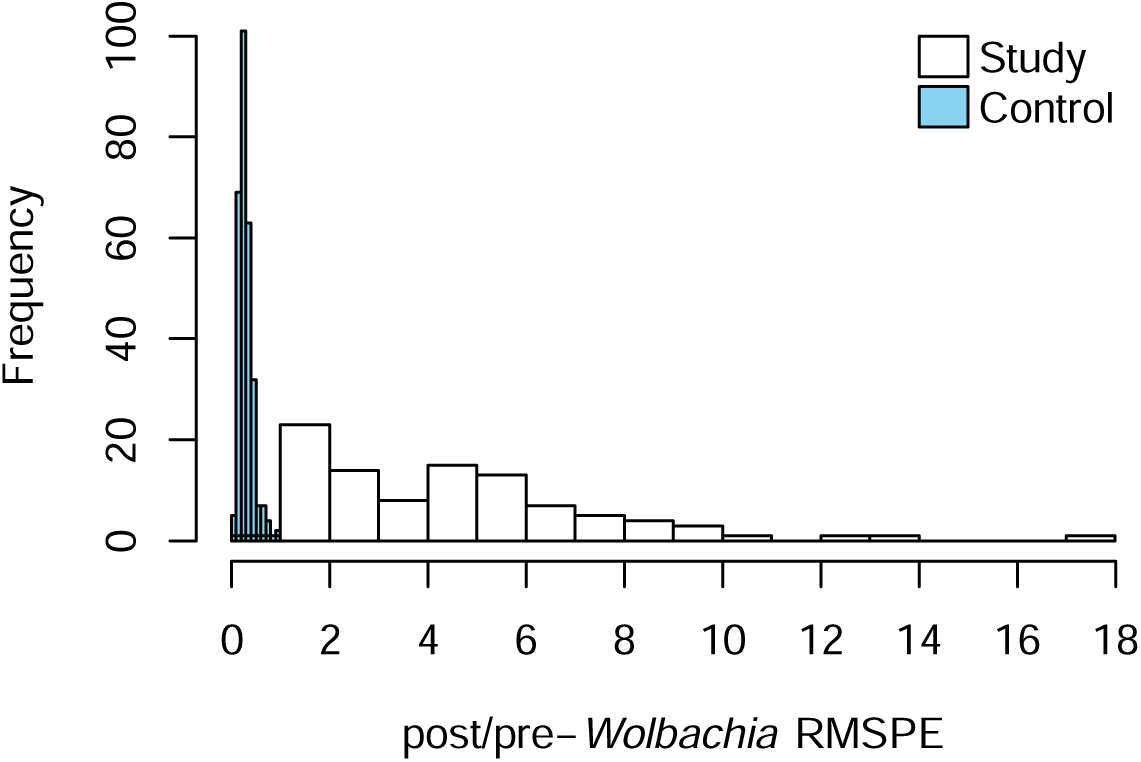
Ratio of post-*Wolbachia* RMSPE and pre-*Wolbachia* RMSPE for release (red) and control sectors (blue).

Lastly, we examined whether the three synthetic control approaches which could appropriately account for covariates reproduced significant intervention efficacies in all sectors post-release. Irrespective of the weighting strategy, positive and significant IEs were estimated for each model, and each IE estimate across each site had similar effect sizes (See supplementary material).

We also performed the above mentioned placebo checks on the synthetic control analysis for female adult *Ae. albopictus* abundance. The synthetic control *GAI_albo_* closely followed the intervention sites’ *GAI_albo_* in the pre-intervention validation periods (1 to 10 weeks), with RMSPE ranging from 0.02 to 0.03 across the intervention group, indicating good model fit in the pre-intervention period. Comparing iteratively the ratios of post versus pre-intervention RMPSEs between intervention sectors and the placebo-intervention sectors for *GAI_albo_*, the probability of obtaining a ratio as high as the actual release sectors, if we randomly assign a site as an intervention site, was below 0.05. (See supplementary information)

## Discussion

We demonstrated that IIT-SIT can lead to dramatic and consistent reductions in adult female *Ae. aegypti* populations. In particular, while initial intervention efficacies in the three months period after the start of interventions averaged around 60.54% (95% CI: 59.59% – 61.44%), this increased to around 90.17% (95% CI: 89.92% – 90.42%) for sites which experienced at least a year of intervention. Progressively, as more sectors within each township adopted intervention (Figure 1), we noticed that these new sites did not significantly dilute the overall intervention (Figure 3). This pattern was demonstrated across all four intervention townships. Our data suggested on average that an at least 6-months of IIT-SIT intervention was required to induce an 80% reduction in mosquito abundance, with further sector-specific reductions in the range of 90 – 100% possible if interventions are sustained in the 1.5 year or more period; averaging at around 91.43% (95% CI: 91.19% – 91.66%) intervention efficacy for adoption of at least 1.5 years of intervention.

However, the potential for elimination in intervention sites may be attenuated by potential migration of wild-type mosquitoes from non-release sites (Figure 7). Analysis of individual traps suggests moderately higher GAI values in traps which are closer to non-release areas, versus those which were at the interior. This suggests potentially more migration of wild-type female *Ae. aegypti* in locations closer to non-release sectors versus locations which are in the interior. We envision that migration issue would be attenuated as IIT-SIT programmes scale-up, with the number of intervention sectors gradually expanded to cover larger areas, and therefore reduce the potential number of sectors which experience migration issues.

Furthermore, spillover of interventions into non-intervention, adjacent sectors may potentially suppress wild-type mosquito populations in non-release locations. We found that estimated positive intervention effects were not limited to solely release sectors, demonstrating an additional advantage of the SIT-IIT approach. Rerunning the synthetic control analysis in neighbouring sectors suggests a smaller, but still significant level of intervention efficacy in non-release sites which are adjacent to intervention sectors which gradually increased over time. We estimated spillover interventions to have increased over EW3 2020 to EW26 2022 from −25.37% to 83.70%. These estimates suggest increasing immigration of *w*AlbB-SG males from buffer sectors to adjacent non-release sectors as intervention time increased in buffer sectors, which led to suppression of wild-type female *Ae. aegypti* in adjacent non-release sectors. These effects do not extend outside of intervention towns however, as we demonstrated that intervention sites do have a significantly larger dip in GAI in the post-intervention period when compared to pre-intervention GAI, compared to control (placebo) sites in almost all cases (Figure 10). The lack of consistent intervention efficacies of *Wolbachia* release on *Ae. albopictus* abundance among the four towns (Figure 9) suggests that changes in abundance may not be attributed to *Wolbachia* releases and warrants further investigation.

Our study has several strengths. (**1**) We used a large number of intervention sites which progressively adopted interventions over different timepoints to estimate the time required to generate a specified level of intervention efficacy. (**2**) Consequentially, the larger number of sites also allowed us to show that over calendar time, intervention efficacies were consistently high despite any potential spatio-temporal differences between sites. (**3**) We used a large and routinely maintained national mosquito surveillance system which comprehensively covered all intervention and control towns to evaluate intervention efficacies. This system, which has data up to the trap-level resolution, also enabled us to examine potential spillover and migration effects associated to the intervention. (**4**) We designed a comprehensive synthetic control framework to evaluate intervention efficacies. In particular, our synthetic control framework which was able to eliminate pre-intervention period biases in mosquito abundance between intervention and control sites, and thereby enabled us to obtain causal estimates of intervention efficacies in the post intervention period. Comprehensive placebo checks by sites and by time also further ensured that we are recovering the true *Wolbachia* intervention effect on mosquito abundance.

There are several limitations to our study. (**1**) The national mosquito surveillance system used for evaluating interventions only has data which started 2019 onwards, while *Wolbachia* interventions have started in 2018. This means that a potentially large number of sites with high time-spent in intervention cannot have intervention effects evaluated. (**2**) While we relied on trap-level data to examine potential wild-type migration into intervention sites, the actual source population of these mosquitoes cannot be determined if they are truly from non-adjacent locations. (**3**) The positive spillover intervention effect on adjacent sites may affect the fit of the synthetic control method for adjacent sites which eventually adopt intervention. This would eventually bias the intervention effects estimated from these locations downwards. However, care was taken to ensure that the overall fit in the pre-intervention period was reasonable (Figure 2, 3). (**4**) While the donor pool exclusively comprised sectors from non-release, never-adjacent townships, any potential spillover intervention effect to non-intervention sectors from intervention townships will bias estimated intervention efficacies in intervention townships downwards, thereby inducing conservative intervention efficacy estimates.

As dengue incidence increases globally and expands to non-endemic areas due to climate change, new tools for vector-control are required to reduce the transmission potential of vector-borne diseases. While our results demonstrate the high efficacy of IIT-SIT in reducing *Ae. aegypti* abundance, the strategy is likely to be enhanced, if complemented with other conventional vector-control measures which effectively target other stages of the mosquito lifecycle, such as source reduction efforts and community engagement to maintain high public vigilance against mosquito breeding and prevent complacency.

## Data Availability

All data produced in the present study are available upon reasonable request to the authors

## Acknowledgements

The authors thank Prof. Zhiyong Xi of Michigan State University for providing the *w*AlbB-infected *Aedes aegypti* line, which was used to generate the *w*AlbB-Sg line essential to this study, and staff of the Applied Entomology Department, National Environment Agency for consistent quality control, production, and releases of *w*AlbB-Sg. This study was supported by funding from Singapore’s Ministry of Finance, Ministry of Sustainability and the Environment, National Environment Agency, and National Robotics Program. JTL is supported by the Ministry of Education (MOE), Singapore Start-up Grant. SB and ARC are supported by an MOE Tier 2 grant.

## Methods

### Monitoring

Adult *Aedes aegypti* and *Ae. albopictus* populations in release and control sectors were monitored weekly using an average of six Gravitraps^23^ per public housing apartment block. Gravitraps were placed in public spaces along corridors and were evenly vertically distributed throughout the block, corresponding to a ratio of approximately one trap for every 20 households. A total of 57,990 traps were deployed in Singapore public housing estates. Donor sites used to construct synthetic controls were all other sectors in towns which did not have *Wolbachia* releases up till EW26 2022, and comprise 30 towns with a population of 3,225,130 individuals. Eventually-treated sectors were not included in the donor pool.

All mosquito samples were sexed and morphologically identified. Female *Aedes* mosquitoes that cannot be speciated morphologically were identified using a multiplex *Ae. aegypti*- and *Ae. albopictus*-specific real-time qPCR assay targeting the *COI* gene (*Aedes* forward 5’-TCC CGC CTT CRG TGC GCG G-3’, *Aedes* reverse 5’-CGC GGG ATG TAY TCA TCA ACC-3’, probe AEG Cy5 – 5’-TAG TCA GAC GTG GTG GTG ACA CAC C-3’– BHQ2, and ALBO HEX – 5’-ACG GTG GCC GGC GTG CCA GTC GT-3’ – BHQ1). The multiplex qPCR reactions were performed in 20µl total volume containing 1X SensiFAST No-Rox Probe Mastermix (Bioline, USA); 0.15µM and 0.05µM COI primers and probes, respectively; 0.4 µM and 0.15µM wsp primers and probes, respectively; and 2µl. Cycling was carried out in a Rotorgene Q (Qiagen, Germany) using the following conditions: 95°C for 5 min, 40 cycles at 95°C for 10s, and 60°C for 30 sec.

Quality controls are regularly carried out in our *Ae. aegypti* colonies that are infected with *Wolbachia* (*w*AlbB-SG). *Wolbachia* in our mosquito colonies were quantified using a multiplex real-time qPCR assay targeting the *w*AlbB *wsp* gene (using primers and probe sequences mentioned above) and the *Aedes aegypti RpS17* gene (forward primer 5’-TCC GTG GTA TCT CCA TCA AGC T-3’, reverse primer 5’-CAC TTC CGG CAC GTA GTT GTC-3’, probe Texas Red – 5’-CAG GAG GAG GAA CGT GAG CGC AG-3 – BHQ2). qPCR reactions were performed in 20ul total volume containing 1X SensiFAST No-Rox Probe Mastermix (Bioline, USA); 0.4µM and 0.2µM wsp primers and probes, respectively; 0.2µM and 0.1µM RpS17 primers and probes, respectively; and 5µl of DNA template. Cycling was carried out in a LightCycler®96 (Roche, USA) using the following conditions: 95°C for 5 min, 45 cycles at 95°C for 10s, and 60°C for 30 sec. Amplification of the *wsp* and *RpS17* genes from individual mosquitoes was compared against a standard curve generated from a 10-fold serial dilution of DNA standard for each gene.

### Production

Eggs were submerged in hatching broth (0.071g baker’s yeast (MP Biomedicals, France), 0.357g nutrient media (CM0001 Nutrient broth, Oxoid, England) and 1000ml deionised water) to induce hatching. After 24 hours, larvae were counted using an automated larvae counter (Orinno Technology Pte Ltd, Singapore) and transferred to rearing trays measuring (53 x 40.5 x 10 cm) or (103 x 63 x 3 cm) at a density of one or four larvae per ml of deionized water, respectively. Larvae were fed with ground Tetramin Flakes (Tetra, Germany) in either slurry or capsule form. Six days post-hatching, male pupae were separated from female pupae and larvae using the Fay-Morlan glass plate separation method (Wolbaki, China) 2018-2020) or the Pupae Separation System (Orinno Technology Pte Ltd, Singapore) (2020-2023) prior irradiation for IIT-SIT releases. Pupae were allowed to eclose inside release containers. For releases in Tampines, the automated, multi-step sex separation process described in Crawford et al.^11^ was followed with pupae collected six-days post hatching. In Tampines town SIT protocols were adopted and irradiated mosquitoes were released from Jan 2020 (at a small location). Irradiated mosquitoes were released in the rest of Tampines town from Aug 2020 onwards.

Irradiation was performed with either the RS2000 (2018 – Jan 2019) or RS2400V irradiators (After Jan 2019) (Rad Source Technologies Inc., USA). For IIT-SIT irradiation (female sterilization), pupae were irradiated in plastic petri dishes with diameter 15.0 cm or in plastic containers measuring 6.5 cm in diameter and 7.0 cm in height (for release). For SIT irradiation (male sterilization), pupae were irradiated in petri dishes with diameter 9.4 cm. Petri dishes or containers were placed at the center of the irradiation chamber of the X-ray irradiator. The effective dose was 30 Gy to 40 Gy, depending on pupae density, irradiator model and desired sterility.

### Release Approach

Releases were conducted twice weekly on weekdays (0630–1030 hrs) at designated locations in high-rise public housing estates covering 607,872 individuals as of Epidemiological Week (EW) 26 2022. Bukit Batok, Choa Chu Kang and Yishun towns were subjected to interventions which combined IIT with SIT. Tampines relied on a high-fidelity sex-sorting approach which increasingly adopted SIT protocols over the trial duration (See Methods Table 1 for summary, supplementary information for details). To trial whether *Ae. aegypti* population suppression could be sustained over increasingly larger areas, Yishun and Tampines were two large towns which were selected to adopt an expanding release strategy, where release sites were gradually expanded to adjacent neighbourhoods. Whereas Bukit Batok and Choa Chu Kang towns were selected based on their smaller size to adopt a targeted release approach, which focused releases on areas with high *Ae. aegypti* abundance and persistent dengue transmission. (Methods Table 1). Towns were demarcated planning areas used by government ministries and departments for administrative purposes. To ensure an even distribution of mosquitoes, releases were conducted at 6–12 equally spaced release locations per apartment block; half of the mosquitoes were released at the ground, and the other half at upper floors, alternating between middle (levels 5–6) and high floors (levels 10–11).

**Methods Table 1:** Summary of *Wolbachia* intervention approaches over 4 townships.

| Township | Bukit Batok | Choa Chu Kang | Tampines | Yishun |
| --- | --- | --- | --- | --- |
| Intervention start date | EW23 2020 | EW20 2020 | EW39 2018 | EW27 2018 |
| Study end date | EW26 2022 | EW26 2022 | EW26 2022 | EW26 2022 |
| Intervention time (weeks) | 109 | 112 | 197 | 209 |
| Total township size (m<sup>2</sup>)<sup>*</sup> | 627,441 | 1,145,559 | 5,088,046 | 3,473,690 |
| Production Approach<sup>**</sup> | IIT-SIT | IIT-SIT | High fidelity sex-sorting | IIT-SIT |
| Frequency of release | Twice weekly | Twice weekly | Twice weekly | Twice weekly |
| Release strategy<sup>***</sup> | Targeted<sup>#</sup> | Targeted | Expanding<sup>##</sup> | Expanding |
| Number of mosquitoes released | 1–7 wAlbB-SG males were released per study site resident per week |  |  |  |
| Total number of mosquitoes released (rounded to thousands) | 17,139,000 | 14,598,000 | 109,432,000 | 77,659,000 |
| Township population covered by release over study period | 40,132 | 64,672 | 272,048 | 231,020 |

<sup>\*</sup>Total area of public housing estates subject to release in respective townships in EW26 2022
<sup>\*\*</sup>The IIT-SIT approach and high-fidelity sex-sorting were detailed above and has been previously characterized<sup>9,11</sup>.
<sup>\*\*\*</sup>Denotes approach to releasing *Wolbachia*-infected *Ae. aegypti* males
<sup>#</sup>Targeted approach which focused releases on areas with high *Aedes aegypti* abundance and persistent dengue transmission
<sup>##</sup>Expanding (“rolling”) approach where release sites were gradually expanded to adjacent neighbourhoods

### Spatio-temporal covariates data

We extracted a comprehensive set of spatio-temporally explicit variables to represent environmental heterogeneity across sectors. Data sources and processing procedures were explicitly detailed in the Supplementary Information. Data includes:

**(1)** 10m vegetation map with areas classified across multiple vegetation types including grass, forest and managed vegetation to signify availability of natural breeding sites and nectar availability for male mosquitoes. The percentage cover of each vegetation type was calculated within each sector as mosquitoes often show preferential areas to breed and rest.
**(2)** The averaged Normalized Difference Vegetation Index per sector was also utilised as an alternative measure of vegetation cover.
**(3)** To represent both host density and urban breeding habitat availability, data on the locations of all public housing estates where over 80% of Singapore’s resident population reside were obtained. Utilising residential location and resale data, the average age of public housing residences was collected as older age is a well-established risk factor for higher Gravitrap indices^24^.
**(4)** Average residence price over the study duration as a proxy for household income and socioeconomic status.
**(5)** Building height, which has been correlated to adult *Aedes* abundance, was calculated according to the number of floors and average height of each level of 3m.
**(6)** The number of condominiums and landed properties was additionally collected within each sector representing additional hosts being available. The percentage cover of built area was calculated as a sum of all residential, commercial and industrial buildings, representing the level of urbanicity, which has been associated with *Ae. aegpyti* presence.
**(7)** The major open drainage network for Singapore was obtained from the Public Utilities Board and has been previously shown as a key breeding site for mosquitoes around HDBs^26^. The average distance of each HDB block within a sector to a drain was measured as well as the length of the network within the sector.
**(8)** Well-established meteorological variables which are known to affect mosquito survival or fecundity were collected. These included daily mean, maximum, and minimum temperature, total rainfall, maximum rainfall falling within a 30-minute, 60-minute and 120-minute window, and wind speed, which were obtained from 21 local weather stations. Hourly dewpoint and ambient ground air temperature were also taken from remote sensing measurements to estimate relative humidity over the time period using standard formula. These values were aggregated at a weekly level to correspond with the dengue case data.

### Estimation of intervention efficacy

We designed a synthetic control framework to examine the impact of releasing male *Ae. aegypti* infected with *Wolbachia* on female *Ae. aegypti* abundance in four large-scale release towns (Bukit Batok, Choa Chu Kang, Tampines and Yishun) from EW8 2019 to EW26 2022. *Ae. aegypti* is the primary vector responsible for dengue transmission in Singapore. The spatial resolution used for analysis were sectors, which comprise 10 – 20 public housing apartment blocks each and measured around 0.088 km^2^ on average. Sectors were used for planning of surveillance and control for environmental infectious diseases in Singapore. The potentially different impacts on *Ae. albopictus* abundance and non-release sectors were also explored.

We used the canonical synthetic control method^27^ to generate synthetic controls for each sector which experienced *Wolbachia* releases. Synthetic controls were a weighted average of donor pool sectors which never experienced releases. The method re-weighs control sectors which were never-treated in the study period such that the average pre-treatment trend of the Gravitrap index (the outcome variable) and other covariates is similar to the outcome variables’ trend and covariates of the treated-sector of interest. The Gravitrap *Ae. aegypti* index proxied for female *Ae. aegypti* abundance. This was done by first calculating the importance of included covariates for the outcome variable and subsequently computing weights which minimised the difference between the treated sector and the synthetic control sector in the importance-weighted covariates. The use of synthetic control methods (SCM) was motivated by potential pre-intervention temporal and spatial differences in mosquito abundance between release and non-release sectors, which renders direct comparison inappropriate. SCM alleviates this by using pre-intervention information on the endpoint of interest to find an optimal set of weights for the non-release group to generate a respective synthetic control for each release sector. This enables the generation of a data-driven counterfactual (synthetic control sector) to the release sector, and consequentially enables causal identification of the intervention effect after weight estimation, by comparing the differences in *Ae. aegypti* abundance between the release sector and its respective synthetic control sector, in the post-intervention period.

We formally define 3 aggregate measures of intervention efficacy (IE), (**A**) the total percentage reduction in female adult *Ae. aegypti* abundance across all time periods (**B**) the percentage reduction in female adult *Ae. aegypti* abundance by time-spent on release for release sectors (**C**) the percentage reduction female adult *Ae. aegypti* abundance by calendar time for release sectors (**D**) the total percentage reduction in female adult *Ae. aegypti* abundance in each sector. All measures compare reduction female adult *Ae. aegypti* abundance in release versus synthetic control sectors. (**A**) Provides a measure of total intervention effect despite year-on-year variations in vector abundance, and only considers comparisons for sectors which adopted releases versus synthetic control sectors (**B**) provides a measure of intervention effect by time spent on releases, to help understand the release duration required to achieve an estimated level of mosquito suppression in a specific sector (**C**) provides a measure of intervention effect by calendar time, to help understand how there may be temporal variations in intervention effect across calendar time (**D**) provides a sector-specific measure of intervention efficacy, and can be compared graphically to treatment time and adjacent sectors to further understand spillover and migration effects of the intervention, as described below.

We constructed 95% confidence intervals by generating 100 bootstrap samples of GAI values using ‘meboot’ package in R, which can create bootstrap timeseries ensembles without assuming stationarity. We then employed the synthetic control method to all bootstrapped timeseries using the same fitting procedure as the full sample. Bootstrap samples were then used to construct the empirical distribution of the synthetic control weights and thereafter used to compute 95% confidence intervals for intervention efficacies.

### Spill-over and migration effects

While treatment sectors were progressively adopted or targeted such that migration can be minimized through the use of roads and geographic areas, immigration of wild-type females from nearby non-release sectors can hamper the upper limit on achievable suppression. We examined potential spill-over and migration effects in four ways. Namely, we recomputed aggregates of intervention efficacies as estimated under the synthetic control method for sectors which were (**1**) at the edge of release zones and considered them buffer sectors (**2**) non-release sectors which were adjacent to the buffer sites and (**3**) core sectors, which were surrounded by buffer sectors and/or existing geographical features such as four-lane roads. (**4**) We additionally examined trap-level Gravitrap data in buffer sectors and estimated correlations between wild-type *Ae, aegypti* abundance and the distance of GAI values to the boundary of the closest non-release adjacent sectors. The correlation was then examined using a generalized additive model, taking the cumulative GAI post release as the dependent variables of interest, and the distance to the boundary of the closest non-release adjacent sector as the explanatory variable of interest.

### Impact on *Aedes albopictus* populations

*Ae. albopictus* is the secondary vector for dengue transmission in the study setting, but *Wolbachia* releases exclusively targets and suppresses adult *Ae. aegypti* populations. While it has been demonstrated that *Ae. albopictus* and *Ae. aegypti* occupy somewhat different ecological niches and their abundance were associated to different drivers^24^, a reduction in *Ae. aegypti* abundance due to *Wolbachia* interventions may reduce interspecies competition, such as larval competition in places with limited aquatic carrying capacity. Using the optimal weighting scheme, we re-estimated synthetic control weights for release sectors using female adult *Ae. albopictus* abundance as the outcome variable. The different measures of intervention efficacy for this endpoint was then estimated by similarly computing the differences in synthetic control and release sectors post-intervention, using the different aggregation schemes.

### Accounting for covariates

Inclusion of both the dependent variable of interest (female adult *Ae. aegypti/albopictus* abundance) together with covariates in the optimization process of standard SCM would render covariates being ignored^28^. This may lead to a weighting scheme where important covariates were not balanced between synthetic controls and release sectors and potentially biases intervention efficacy estimates. To obviate this risk, we considered four alternative estimators for generating weights for each synthetic control. Namely, using pre-release values of the dependent variable and the covariates, we calculated SCM weights using all pre-intervention covariates and female adult *Ae. aegypti* abundance values (**M1**), only the latest pre-intervention value of all covariates and female adult *Ae. aegypti* abundance values (**M2**), the pre-intervention average of all covariates and female adult *Ae. aegypti* abundance values (**M3**) and using all pre-release observations of female adult *Ae. aegypti* abundance without covariates (**M4**). **M1** and **4** represents the standard SCM with and without covariates respectively. **M2** is motivated by arguing that weights estimated based on the final value enables us to achieve a good fit at the pre-release cut-off time. **M3** is motivated by past work arguing that averaging both covariate and dependent variable values lead to constant weightage between both variable times in SCM weight estimation^25,29^. We used mean differences or standardized mean differences to assess covariate balance between release and synthetic-control sectors. Good balance was achieved across the pre- and post-intervention periods for **M2** and **3** (See supplementary information).

We considered maximum temperature (°C), mean temperature (°C), minimum temperature (°C), rainfall (mm), mean wind speed, maximum wind speed, mean relative humidity, NDVI (vegetation index), area within 300m of a waterbody (%), area within 500m of a waterbody (%), vegetation density, average age of public housing (years), average price of housing ($s), distance of centroid to drainage network (m), length of drainage network inside spatial unit (m), grass area (%), total vegetation area (%), dewpoint temperature, vapor pressure, large-scale precipitation, 2 meter temperature, max. 2 meter temperature, surface pressure, soil temperature, total precipitation and number of public housing units. Several covariates were previously found to be associated to both *Ae aegypti* and *Ae. albopictus* abundance in the study setting^18^.

As **M1** failed to take into account any covariates in the weighting process, it was precluded from all other assessments (See supplementary information).

### Robustness check on synthetic control method

We examined the bias of our proposed estimators for the synthetic control method in the period prior to releases of *Wolbachia*. Negligible bias in the pre-release period can ensure that the proposed synthetic control estimator is generating valid synthetic control sectors in the post-release period for comparison to actual release sectors. In each sector, we split female adult *Ae. aegypti* abundance time series in intervention sites into initial training (2019, 10 weeks prior to intervention) and validation (10 weeks prior to intervention) sets. In the initial time step, we estimated weighting schemes using **M2–4** by incorporating the control sectors to generate synthetic controls for the release sectors. Thereafter, we computed the forecast error between the synthetic control group and the release group by employing a cross-validation approach. If the weighting scheme is valid, the differences in female adult *Ae. aegypti* abundance between synthetic control and release sectors in the pre-release period should be similar. After the initial time-step, we reiterated this procedure by iteratively adding 1 more epidemiological week of data and re-estimating the weighting scheme, with forecast errors recomputed and stored for this time step. This procedure is repeated till the validation set is empty. We thereafter compute the differences in forecast errors between the synthetic control and release groups in the pre-release period examine potential biases in the synthetic control estimators. **M4** was the estimator which demonstrated the smallest forecast error across most forecast horizons and interventions. It was thus taken as the estimator for the weighting scheme used to generate synthetic controls for release sectors. Supplementary information section 3 provides full details of weighting scheme and our assessment approaches.

### Placebo checks on synthetic control method

We subjected our synthetic control framework to a battery of sensitivity analyses to further ascertain the validity of our intervention efficacy estimates. First, we constructed placebo intervention events 13 and 24 weeks before the earliest actual release occurred in each sector. These were conducted to determine whether measured intervention effects were attributable to treatment anticipation of *Wolbachia* interventions occurring before actual interventions, which may mediate individual-level behaviour to vector control and potentially confound mosquito abundance in the pre-release period. Using the optimal weighting scheme, we constructed synthetic controls before the actual release occurs and measured the treatment effect of the placebo intervention.

Thereafter, we employed another placebo test to (**1**) examine whether there are any potential intervention spillover effects to control regions and (**2**) whether the observed effect on dengue incidence rates in intervention sites was truly related to the implementation of *Wolbachia* interventions rather than being influenced by other processes^25,29^. We constructed synthetic controls for all the control sites and did permutation test to check if the results are credible by comparing the distribution of ratio of post-to pre-*Wolbachia* RMPSE for control and release sectors.

All robustness and placebo checks were also repeated, taking *Ae. albopictus* abundance as the outcome variable of interest. This was done to assess the validity of the synthetic control method in examining the impact of *Wolbachia* interventions on *Ae. albopictus* abundance. We also verified the robustness of our results of spillover effects, by repeating all assessment checks on the outcome of *Ae. aegypti* abundance in sites which were eventually adjacent and never treated or not-yet treated sites.

