## Supplementary material for "Efficacy of *Wolbachia*-mediated sterility to suppress adult *Aedes aegypti* populations": SI

### Supplementary Information for "*Efficacy of Wolbachia-mediated sterility to suppress Aedes aegypti populations*"

October 2023

6.3 Estimated intervention efficacies on *Aedes albopictus* abundance by event time for best model 42

6.4 Estimated intervention efficacies on *Aedes aegypti* abundance in non-release, adjacent sectors by event time for best model . . . . . 44

### 1 Data

We extracted a set of spatially explicit variables to represent environmental heterogeneity across sectors.

#### 1.1 Vegetation data

A 10m vegetation map (Gaw et al., 2019) with areas classified across multiple vegetation types including grass, forest and managed vegetation based on Sentinel-2 satellite data, was utilised to signify the availability of natural breeding sites and nectar availability for mosquito males. The percentage cover of each vegetation type was calculated within each sector as mosquitoes often show preferential areas to breed and rest (USGS, 2023a) Similarly, the averaged Landsat Normalized Difference Vegetation Index per sector was also utilised for this purpose (USGS, 2023b).

#### 1.2 Residential data

To represent both host density and urban breeding habitat availability, data on the locations of public housing estates named Housing Development Board (HDBs) where over 80% of Singapore's resident population reside was obtained from Onemap (OneMap, 2023) Utilising the HDB location and HDB resale data, the average age of HDB buildings was collected as older age is a well-established risk factor for higher Gravitrap indices (OGP, 2023.) This is due to building deterioration providing additional breeding habitats in cracks and design features such as laundry poles which are no longer built due to the pooling of water within the supports. Average HDB house price from 2015–2022, a proxy for household income and socioeconomic status, was calculated based on an XGBoost model previously employed (Park et al., 2020). Building height, which has also been correlated to Gravitrap indices, was calculated according to the number of floors and average height of each level of 3m. The number of condominiums and landed properties was additionally collected within each sector representing additional hosts being available. The percentage cover of built area was calculated as a sum of all residential, commercial and industrial buildings, representing the level of urbanicity, which has been associated with *Ae. aegpyti* presence (Kolimenakis et al., 2021) The major open drainage network for Singapore was obtained from the Public Utilities Board as a key breeding site for mosquitoes around HDBs. The average distance of each HDB block within a sector to a drain was measured as well as the length of the network within the sector (Fernandez et al., 2023)

#### 1.3 Meteorological data

For meteorological data, well-established variables which are known to affect mosquito survival or fecundity were collected. These included daily mean, maximum, and minimum temperature, total rainfall, maximum rainfall falling within a 30-minute, 60-minute and 120-minute window, and wind speed, which were obtained from a total of 21 weather stations installed by the National Environment Agency. We created daily complete raster maps through inverse distance weighting interpolation, which was carried out using cross validation of leave-one-out for the fitting of the inverse distancing power to minimise the error in observation on the raster surface of the test point. Hourly dewpoint and ambient ground air temperature were taken from ERA5, published by ECMWF (ECMWF, 2023), to estimate relative humidity over the time period using standard formula (Lawrence, 2005) These values were aggregated at a weekly level to correspond with the dengue case data (DoS, 2023)

#### 1.4 Entomological data

Gravitraps were placed along the common corridors of public housing apartments in all intervention sites, at the ratio of 1 trap for every 20 households, i.e. two to three traps on each of three floors per apartment block: lower floor (2nd), mid floor (5th or 6th floor) and high floor (10th or 11th floor) from

EW8 2019 to EW26 2022. This corresponded to an average of six Gravitraps per apartment block. This trap-to-household ratio and deployment was based on logistic considerations for long term monitoring, and it provided an assessment of the density as well as vertical distribution of the mosquito population. Mosquito data was collected from all Gravitraps on a bi-weekly basis.

#### 2 Study setting

117 sectors grouped into 4 intervention towns comprising of an at-risk population of 607,872 residents have been subject to *Wolbachia* releases under two separate approaches. First, the phased release approach where releases were gradually expanded to encompass the sectors in Yishun and Tampines townships (See Figure 1,2). Secondly, the targeted approach where large areas of the intervention site were subject to *Wolbachia* releases from the start date of intervention. This was employed in Bukit Batok (Figure 4) and Choa Chu Kang (Figure 3) townships. Out of the 117 sectors, 94 sectors received *Wolbachia* release from EW1 2020 onward with at least a year pre-intervention data available and are analysed using the synthetic control method (described below). The exclusion of the 20 sectors was due to the need for a sufficient pre-intervention period of the outcome variable to generate synthetic controls as counterfactuals for each release site to which intervention efficacies could be obtained. A 1-year period was thus taken as the minimum length used to obtain synthetic control weights for the 94 sectors.

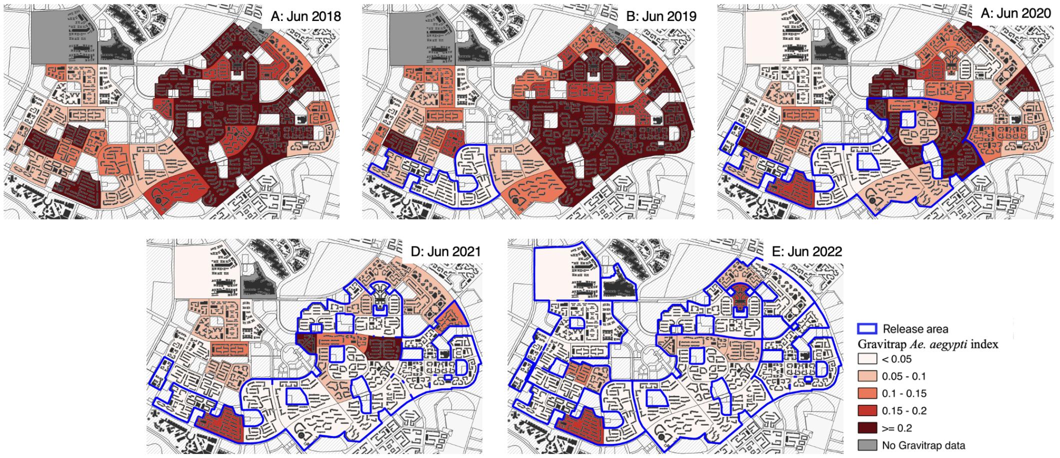

Figure 1: Visualization of the phased release approach in Tampines township from Sep 2018 to March 2022. Regions which experienced releases are shaded in the map.

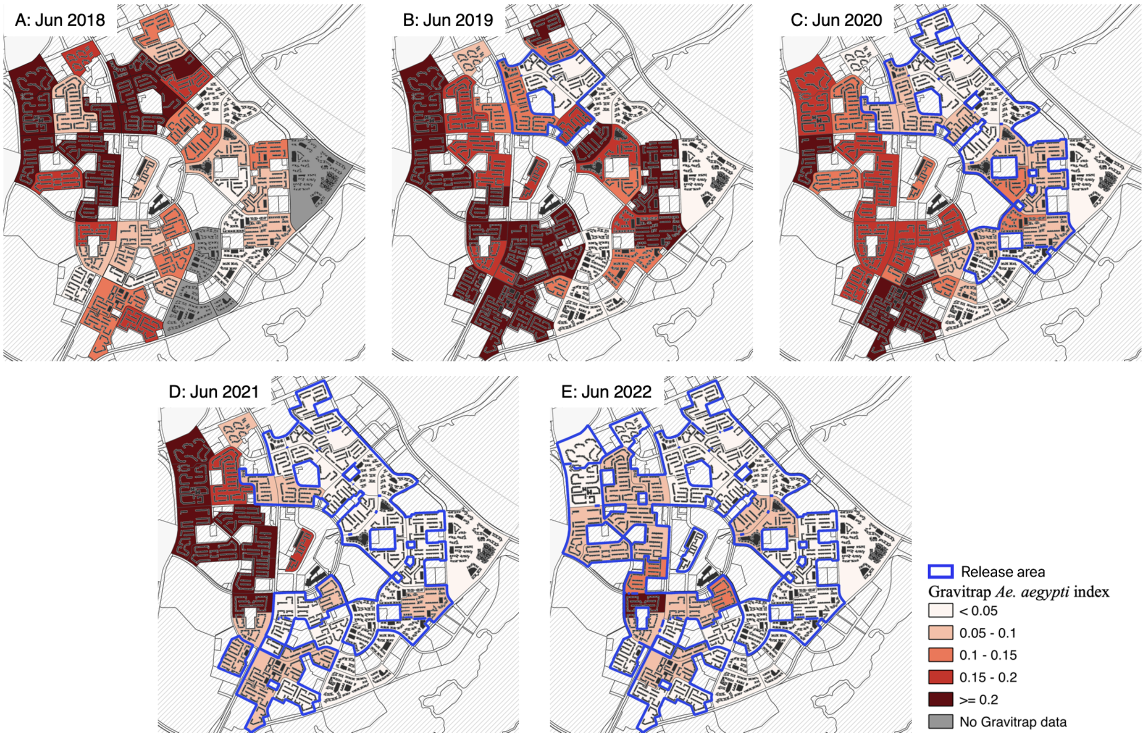

Figure 2: Visualization of the phased release approach in Yishun township from July 2018 to March 2022. Regions which experienced releases are shaded in the map.

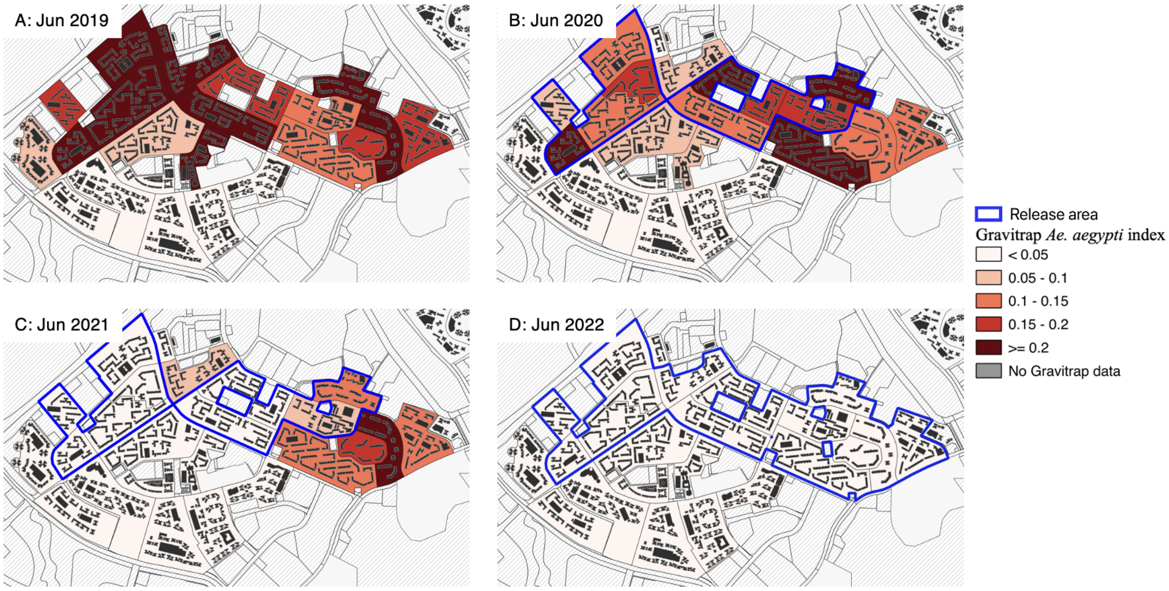

Figure 3: Visualization of the targeted release approach in Choa Chu Kang township from May 2020 to March 2022. Regions which experienced releases are shaded in the map.

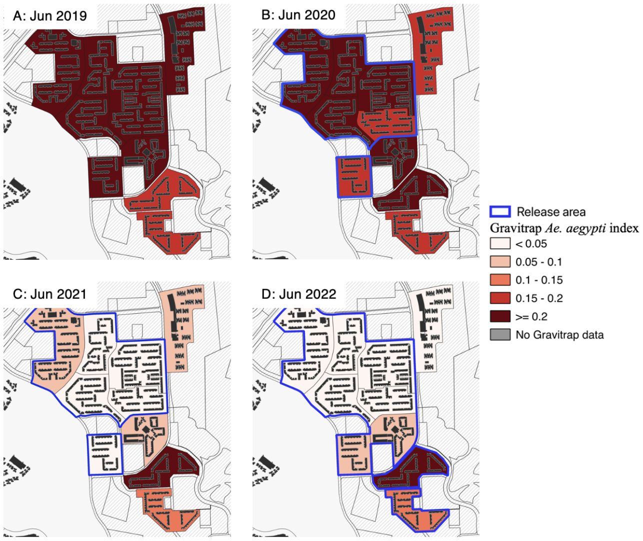

Figure 4: Visualization of the targeted release approach in Bukit Batok township from June 2020 to March 2022. Regions which experienced releases are shaded in the map.

### 3 Synthetic control framework to estimate intervention efficacy

Trends in *Aedes* abundance as proxied by the Gravitrap index (GAI) between different release areas versus control areas may be different, along with differences in other geographical characteristics which may confound estimates of intervention efficacy. This makes the estimation of intervention efficacies for *Wolbachia* incompatible insect technique on *Aedes* abundance difficult. We therefore employed the synthetic control method (Abadie et al., 2010) while appropriately accounting for potential inbalance between covariates to estimate the intervention efficacies in four trial sites (Bukit Batok, Choa Chu Kang, Tampines, Yishun).

#### 3.1 Construction of synthetic controls

Our study comprises of $j = \{1, 2, \dots, J, J+1, \dots, J+N_{tr}\}$ units, where $j = J+1, \dots, J+N_{tr}$ are study units and rest $J$ units are control units which comprise the donor pool to construct synthetic controls. Both intervention and control units are observed over time periods $t_j = \{1, 2 \dots, T_{0,j}, T_{0,j}+1 \dots T_j\}$. Let $T_{0,j}$ be the number of time periods before intervention where both control and intervention sectors never experienced *Wolbachia* releases. $T_{1,j} = T_j - T_{0,j}$ is the number of time periods after intervention for a respective sector $j$. In the time period after $T_{0,j}$, control units are not subject to *Wolbachia* releases.

We construct synthetic control for each of the intervention sites individually. Abadie et al., 2010 defines synthetic control implicitly as the weighted average of the donor pool such that the weights $w$ sum to 1, i.e., $W = (w_1, \dots, w_J)'$, with $0 \leq w_j \leq 1$ for $j = 1, \dots, J$ and $w_1 + \dots + w_J = 1$. $W^*$ is chosen as the value of $W$ that minimizes the pre-intervention period difference in intervention and control sites for both the dependent variable of interest (GAI) and other covariates as described in the preceding section:

$$(X_1 - X_0 W)' V (X_1 - X_0 W) \tag{1}$$

where $X_1$ is $k \times 1$ matrix of $k$ predictors of a treated sector and $X_0$ is $k \times J$ matrix with $k$ for each of the units in the donor pool. We omit the sector subscript $j$ for clarity. $V$ is a $k \times k$ diagonal matrix with $v_m$ signifying the importance of $m^{th}$ variable when measuring the difference between $X_1$ and $X_0 W$.

Only recently the SCM optimization problem was given explicitly by Malo et al., 2020 as a bi-level optimization problem:

$$\min_{V,W} (Y_1^{\text{pre}} - Y_0^{\text{pre}} W(V))'(Y_1^{\text{pre}} - Y_0^{\text{pre}} W(V)) \tag{2}$$

$$\text{such that } W(V) = \text{argmin}(X_1 - X_0 W)'V(X_1 - X_0 W) \tag{3}$$

$$1'W = 1$$

$$1'V = 1$$

$$V, W >= 0$$

where $Y_1^{\text{pre}}$ is a $T_0 \times 1$ matrix of pre-intervention outcomes for treated unit and $Y_0^{\text{pre}}$ is a $T_0 \times J$ matrix of pre-intervention outcomes for donor pool.

Accounting for covariates inclusion of both the dependent variable of interest (GAI) together with spatial confounders in the optimization process of standard synthetic control methods would lead to spatial confounders being ignored in the weighting process. We obviate this risk by exploring four separate approaches to construct synthetic controls. Namely: using pre-intervention values of the dependent variable, as well as observed values of the covariates, we calculated SCM weights using all pre-intervention covariates and GAI (**M1**), only the latest pre-intervention value of all covariates and GAI (**M2**), the pre-intervention average of all covariates and GAI (**M3**) and using all pre-intervention GAI observations without covariates (**M4**). **M1** and **4** represents the standard synthetic control method with and without covariates respectively. **M2** is motivated by arguing that weighting based on the final value enables us to achieve a good fit at the pre-intervention cut-off time. **M3** is motivated by past work arguing that averaging both covariate and dependent variable values lead to constant weightage between both variable times in SCM weight estimation (Abadie and Gardeazabal, 2003; Abadie et al., 2015).

Following Sections 3.3,4, **M4** dominates on each of the assessment criterion and was therefore chosen as the weighting framework to estimate the effect of *Wolbachia* releases on GAI in release sites.

#### 3.2 Estimating using all pre-intervention outcomes as predictors

When we take all lagged values of the dependent variable (GAI) as the predictor, the optimal solution theoretically is to choose the predictor weights such that the covariates are zero and only the lagged values of dependent variable are included for weighting. We rehash the results of Kaul et al., 2022:

We partition the predictor data matrices $X_1$ and $X_0$ as

$$X_1 = \begin{pmatrix} C_1 \\ Z_1 \end{pmatrix}, \quad X_0 = \begin{pmatrix} C_0 \\ Z_0 \end{pmatrix}$$

with $C_0$ and $C_1$ containing the values of the covariates of the control and treated sectors, respectively; and $Z_1 = Y_1^{pre}$ and $Z_0 = Y_0^{pre}$. We partition $V$ correspondingly as,

$$V = \begin{pmatrix} V_C & 0 \\ 0 & V_Z \end{pmatrix}$$

where $V_C$ contains the weights for the covariates, while $V_Z$ collects the weights of the pre-treatment values of the dependent variable. The inner optimization (3) becomes –

$$(C_1 - C_0 W)'V_C(C_1 - C_0 W) + (Z_1 - Z_0 W)'V_Z(Z_1 - Z_0 W) \tag{4}$$

Denoting $W^{**}$ as the weights $W$ that produce the best fit with respect to the outcome variable alone,

$$W^{**} := \text{argmin}_W (Z_1 - Z_0 W)'(Z_1 - Z_0 W)$$

then by definition of $W^{**}$, we have –

$$(Z_1 - Z_0W^*)'(Z_1 - Z_0W^*) \leq (Z_1 - Z_0W^*(V)'(Z_1 - Z_0W^*(V)) \textit{ for all } V$$

where $W^*(V)$ is the solution to the outer optimization problem (2).

On the other hand, if we choose $V^*$ with $V_C^*$ equal to zero and $V_Z^*$ equal to the identity matrix, then inspection of Equation (2) gives

$$W^*(V^*) = W^{**}$$

Therefore, although the inner objective function (3) explicitly takes the covariates into account, the synthetic control resulting after optimizing the outer objective function (2) will be calculated by ignoring the covariates completely.

We ran **M1** to verify the above theoretical results in all intervention areas. That is, used all observations of GAI and geographical characteristics to generate synthetic controls for intervention townships in the pre-intervention period using the donor group. In all townships, all covariate weights were estimated to be 0. Since **M1** fails to take covariates into account, subsequent analysis is only performed for **M2–4**, to explicitly take into account potential confounders of *Aedes* abundance in both the intervention sites and donor pool.

##### 3.3 Cross-validation to assess weighting scheme validity

To assess the validity of our weighting scheme, we employed a battery of robustness checks. Placebo checks are detailed in succeeding subsections after estimation of intervention efficacies.

First, we employed cross-validation by splitting the pre-intervention time period into separate training and validation periods. The validation period is iteratively increased from 1 to 10 weeks and forecast error is calculated for the validation period for each study site. This was computed for all considered weighting schemes for all intervention sectors. The average RMSPE increased as validation period was longer since the model was trained on smaller period of time. Model 4 in general gives the smallest average RMSPE across the validation periods (See Table 1).

Continuing the notation above (and also in subsequent sections) with $J+1, J+2, J+3, ..., J+N_{tr}$ as treated units and $j = 1 \ldots J$ as control units, forecast error for each treated unit $i = J+1, J+2, J+3, ..., J+N_{tr}$ is calculated as:

$$\left( \frac{1}{cv} \sum_{t=t_{cv}}^{T} (Y_{it} - \sum_{j=1}^{J} w_j^* Y_{jt})^2 \right)^{1/2}, \ t_{cv} = T_0 - cv$$

where $cv = \{1, 2 \ldots 8\}$ denotes the validation period and $Y_{it}$, $Y_{jt}$ are the GAI of control unit $j$, intervention unit $i$ at time $t$.

| Validation period | Model 2 | Model 3 | Model 4 |
|---|---|---|---|
| 1 | 0.085 | 0.102 | 0.088 |
| 2 | 0.089 | 0.091 | 0.083 |
| 3 | 0.087 | 0.088 | 0.083 |
| 4 | 0.090 | 0.093 | 0.084 |
| 5 | 0.088 | 0.092 | 0.086 |
| 6 | 0.087 | 0.092 | 0.087 |
| 7 | 0.091 | 0.094 | 0.089 |
| 8 | 0.090 | 0.093 | 0.090 |
| 9 | 0.090 | 0.095 | 0.091 |
| 10 | 0.093 | 0.096 | 0.093 |

Table 1: Average root mean squared prediction error (RMSPE) for female *Aedes aegypti* abundance across all sites calculated during validation period of 1 to 10 weeks.

##### 3.4 Estimation of *Wolbachia* intervention effect on *Aedes* abundance

The effect of *Wolbachia* releases on intervention sectors at time $t$ is given by the comparison of post-intervention outcomes in the intervention sectors and post-intervention outcome had the intervention sectors not experienced the intervention, with the latter estimated as $w_j^* Y_{jt}$, the synthetic control sector:

$$\alpha_{it} = Y_{it} - \sum_{j=1}^{J} w_j^* Y_{jt}, \text{ where } T_0 < t \leq T$$

We further define the intervention efficacy ($IE$) of *Wolbachia* releases on intervention sectors for each week as the weekly percentage change in observed and synthetic GAI in the post-intervention period, from the start time.

$$IE_{y,w} \text{ (\% change in week } w \text{ of year } y) = \frac{-(S.Obs_{y,w} - S.Syn_{y,w})}{S.Syn_{y,w}} \times 100 \quad (5)$$

where $S.Obs_{y.w}$ and $S.Syn_{y.w}$ are the sum of observed and synthetic GAI in epidemiological week $w$ of year $y$. Note that a positive IE implies decrease in GAI due do intervention.

We additionally estimated the effect of *Wolbachia* release by different spatial (core, buffer, non-release but adjacent) and temporal (event time, calendar time) aggregates as described in the main text. The synthetic control method was also employed taking *Aedes albopictus* abundance as the endpoint of interest, to examine potential effects of *Wolbachia* intervention on other mosquito specieses, using the different aggregates.

##### 3.5 Weights of donor pool

Tables 3–15 show the contribution of control sectors for synthetic control of all the intervention sectors across all weighting schemes. The tables only show sectors which contribute to synthetic control weights for each respective intervention sectors.

Table 2: Donor weights for Bukit Batok sectors for model 2. Only sectors with any contribution to synthetic control weights for intervention sectors' synthetic controls are shown

| Donor | | Model 2 | | | | | | | | |
|---|---|---|---|---|---|---|---|---|---|---|
| Town | Sector | FL424 | FL425 | FL426 | FL633 | FL634 | FL641 | FL655 | FL833 | FL833b |
| Ang Mo Kio | FL386 | 0 | 0 | **0.30** | 0 | 0 | 0 | 0 | 0 | 0 |
| | FL455 | 0 | **0.30** | 0 | 0 | 0 | **0.20** | 0 | 0 | 0 |
| | FL472 | 0 | 0 | **0.08** | 0 | 0 | 0 | 0 | 0 | 0 |
| | FL499 | 0 | **0.07** | 0 | 0 | 0 | 0 | 0 | 0 | 0 |
| | FL500 | 0 | 0 | 0 | 0 | 0 | 0 | 0 | **0.14** | 0 |
| Bedok | FL237 | 0 | 0 | 0 | **0.02** | 0 | 0 | 0 | 0 | 0 |
| Bishan | FL34 | 0 | 0 | 0 | **0.07** | **0.08** | 0 | 0 | 0 | **0.05** |
| Bukit Batok | FL435 | **0.23** | 0 | 0 | 0 | 0 | 0 | 0 | **0.15** | 0 |
| | FL681 | **0.10** | 0 | 0 | 0 | 0 | 0 | **0.22** | 0 | 0 |
| | FL682 | 0 | 0 | 0 | 0 | **0.20** | 0 | 0 | 0 | 0 |
| | FL686 | 0 | 0 | 0 | 0 | **0.26** | 0 | 0 | 0 | 0 |
| | FL699 | **0.21** | **0.29** | 0 | **0.46** | 0 | **0.25** | 0 | 0 | 0 |
| | FL702 | 0 | 0 | 0 | 0 | 0 | **0.16** | 0 | 0 | 0 |
| | FL731 | 0 | 0 | 0 | **0.37** | **0.18** | 0 | 0 | 0 | **0.23** |
| Bukit Panjang | FL647 | 0 | 0 | 0 | 0 | 0 | 0 | **0.17** | 0 | 0 |
| Choa Chu Kang | FL729 | 0 | 0 | **0.05** | 0 | 0 | 0 | 0 | 0 | 0 |
| | FL811 | 0 | 0 | 0 | 0 | 0 | 0 | 0 | 0 | **0.08** |
| | FL814 | **0.01** | 0 | 0 | 0 | 0 | 0 | 0 | 0 | 0 |
| Clementi | FL423 | 0 | 0 | 0 | 0 | 0 | **0.06** | 0 | 0 | 0 |
| | FL640 | 0 | 0 | 0 | 0 | 0 | **0.11** | 0 | 0 | **0.06** |
| Geylang | FL83 | 0 | 0 | 0 | 0 | 0 | 0 | 0 | 0 | **0.02** |
| Hougang | FL405 | **0.04** | 0 | 0 | 0 | 0 | 0 | 0 | 0 | 0 |
| | FL409 | 0 | 0 | 0 | **0.01** | 0 | 0 | 0 | 0 | 0 |
| | FL414 | 0 | 0 | 0 | 0 | **0.12** | 0 | 0 | 0 | 0 |
| | FL464 | 0 | 0 | 0 | 0 | 0 | **0.13** | 0 | 0 | 0 |
| Jurong East | FL665 | 0 | 0 | **0.01** | 0 | 0 | 0 | 0 | 0 | 0 |
| | FL701 | 0 | 0 | 0 | 0 | 0 | 0 | 0 | **0.47** | **0.35** |
| Jurong West | FL431 | 0 | 0 | 0 | 0 | 0 | 0 | 0 | 0 | **0.05** |
| | FL621 | 0 | 0 | 0 | 0 | **0.15** | 0 | 0 | 0 | 0 |
| | FL625 | 0 | 0 | **0.07** | 0 | 0 | 0 | 0 | 0 | 0 |
| | FL638 | 0 | **0.17** | 0 | 0 | 0 | 0 | **0.12** | 0 | 0 |
| | FL694 | 0 | 0 | 0 | 0 | 0 | 0 | **0.10** | 0 | 0 |
| | FL709 | 0 | **0.10** | 0 | 0 | 0 | 0 | 0 | 0 | 0 |
| | FL769 | 0 | 0 | 0 | 0 | 0 | 0 | **0.07** | 0 | 0 |
| | FL777 | 0 | 0 | 0 | 0 | 0 | 0 | 0 | 0 | **0.11** |
| Kallang | FL13 | 0 | 0 | 0 | 0 | 0 | 0 | 0 | 0 | **0.06** |
| | FL185 | 0 | 0 | 0 | **0.05** | 0 | 0 | 0 | 0 | 0 |
| Marine Parade | FL311 | **0.03** | 0 | 0 | 0 | 0 | 0 | 0 | 0 | 0 |
| Novena | FL68 | 0 | 0 | 0 | **0.02** | 0 | 0 | 0 | **0.15** | 0 |
| Pasir Ris | FL217 | **0.18** | 0 | 0 | 0 | 0 | 0 | 0 | 0 | 0 |
| | FL228 | 0 | 0 | 0 | 0 | 0 | 0 | **0.23** | 0 | 0 |
| Queenstown | FL352 | 0 | 0 | 0 | 0 | 0 | 0 | **0.10** | **0.06** | 0 |
| Rochor | FL73 | 0 | 0 | **0.19** | 0 | 0 | 0 | 0 | 0 | 0 |
| Serangoon | FL480 | 0 | 0 | **0.20** | 0 | 0 | 0 | 0 | 0 | 0 |
| Toa Payoh | FL59 | 0 | **0.06** | 0 | 0 | 0 | 0 | 0 | 0 | 0 |
| Woodlands | FL587 | 0 | 0 | 0 | 0 | 0 | 0 | 0 | **0.03** | 0 |
| | FL591 | **0.22** | 0 | 0 | 0 | 0 | **0.10** | 0 | 0 | 0 |
| | FL600 | 0 | 0 | **0.11** | 0 | 0 | 0 | 0 | 0 | 0 |

Table 3: Donor weights for Bukit Batok sectors for models 3 and 4. Only sectors with any contribution to synthetic control weights for intervention sectors' synthetic controls are shown

| Donor | | Model 3 | | | | | | | | | Model 4 | | | | | | | | |
|---|---|---|---|---|---|---|---|---|---|---|---|---|---|---|---|---|---|---|---|
| Town | Sector | FL424 | FL425 | FL426 | FL633 | FL634 | FL641 | FL655 | FL833 | FL833b | FL424 | FL425 | FL426 | FL633 | FL634 | FL641 | FL655 | FL833 | FL833b |
| | FL372 | 0 | 0 | 0 | 0 | 0 | 0 | 0 | 0 | 0 | 0 | 0 | 0 | 0 | 0 | 0 | 0 | 0 | **0.01** |
| | FL386 | 0 | 0 | 0 | 0 | 0 | 0 | 0 | 0 | 0 | 0 | 0 | **0.01** | 0 | 0 | 0 | 0 | 0 | 0 |
| | FL455 | **0.14** | 0 | 0 | 0 | 0 | **0.33** | 0 | 0 | 0 | **0.06** | 0 | 0 | 0 | 0 | **0.01** | 0 | 0 | 0 |
| Ang Mo Kio | FL471 | 0 | 0 | **0.01** | 0 | 0 | 0 | 0 | 0 | 0 | 0 | 0 | 0 | 0 | 0 | 0 | 0 | 0 | 0 |

Continued on next page

**Table 3 – continued from previous page**

| Donor | | Model 3 | | | | | | | | | Model 4 | | | | | | | | |
|---|---|---|---|---|---|---|---|---|---|---|---|---|---|---|---|---|---|---|---|
| Town | Sector | FL424 | FL425 | FL426 | FL633 | FL634 | FL641 | FL655 | FL833 | FL833b | FL424 | FL425 | FL426 | FL633 | FL634 | FL641 | FL655 | FL833 | FL833b |
| | FL472 | 0 | 0 | 0 | 0 | 0 | 0 | 0 | 0 | 0 | 0 | 0 | **0.01** | 0 | 0 | 0 | 0 | 0 | 0 |
| | FL499 | 0 | **0.29** | 0 | 0 | 0 | 0 | 0 | 0 | 0 | 0 | 0 | 0 | 0 | 0 | 0 | 0 | 0 | 0 |
| | FL500 | 0 | **0.17** | 0 | 0 | 0 | 0 | 0 | 0 | 0 | 0 | 0 | 0 | 0 | 0 | 0 | 0 | 0 | 0 |
| Bedok | FL211 | 0 | 0 | 0 | 0 | 0 | 0 | 0 | 0 | 0 | 0 | 0 | 0 | 0 | 0 | 0 | **0.11** | 0 | 0 |
| | FL231 | 0 | 0 | 0 | 0 | 0 | 0 | **0.01** | 0 | 0 | 0 | 0 | 0 | 0 | 0 | 0 | 0 | 0 | 0 |
| | FL237 | 0 | 0 | 0 | 0 | 0 | 0 | 0 | 0 | 0 | **0.04** | 0 | 0 | 0 | 0 | 0 | 0 | 0 | 0 |
| Bishan | FL34 | 0 | 0 | 0 | **0.31** | **0.14** | 0 | 0 | 0 | 0 | 0 | 0 | 0 | **0.33** | **0.28** | 0 | 0 | **0.05** | 0 |
| | FL36 | 0 | 0 | 0 | 0 | 0 | 0 | 0 | 0 | 0 | 0 | 0 | 0 | 0 | 0 | 0 | 0 | **0.01** | 0 |
| | FL47 | 0 | 0 | 0 | 0 | 0 | 0 | 0 | 0 | 0 | 0 | 0 | 0 | 0 | 0 | 0 | 0 | **0.11** | 0 |
| Bukit Batok | FL434 | 0 | 0 | 0 | **0.05** | **0.20** | 0 | 0 | 0 | **0.09** | 0 | 0 | 0 | **0.03** | **0.03** | 0 | 0 | 0 | **0.15** |
| | FL435 | 0 | 0 | 0 | 0 | 0 | 0 | 0 | 0 | 0 | 0 | 0 | 0 | 0 | 0 | 0 | 0 | 0 | 0 |
| | FL681 | 0 | 0 | 0 | 0 | 0 | 0 | 0 | 0 | 0 | 0 | 0 | 0 | 0 | 0 | 0 | 0 | 0 | 0 |
| | FL682 | **0.05** | 0 | 0 | 0 | **0.29** | 0 | 0 | 0 | 0 | 0 | 0 | 0 | 0 | **0.01** | 0 | 0 | 0 | 0 |
| | FL686 | 0 | 0 | 0 | 0 | **0.24** | 0 | 0 | 0 | 0 | 0 | 0 | 0 | **0.07** | **0.16** | 0 | 0 | 0 | 0 |
| | FL699 | **0.24** | **0.27** | 0 | 0 | 0 | 0 | 0 | 0 | 0 | 0 | 0 | 0 | **0.16** | 0 | 0 | 0 | 0 | 0 |
| | FL702 | 0 | 0 | **0.21** | 0 | 0 | **0.13** | 0 | 0 | 0 | 0 | 0 | 0 | 0 | 0 | 0 | 0 | 0 | 0 |
| | FL731 | 0 | 0 | 0 | **0.16** | **0.06** | 0 | 0 | 0 | **0.16** | 0 | 0 | 0 | **0.16** | 0 | 0 | 0 | 0 | **0.03** |
| Bukit Merah | FL174 | 0 | 0 | 0 | 0 | 0 | 0 | 0 | 0 | 0 | 0 | **0.01** | 0 | 0 | 0 | 0 | 0 | 0 | 0 |
| | FL193 | 0 | 0 | 0 | 0 | 0 | **0.12** | 0 | 0 | 0 | 0 | 0 | 0 | 0 | 0 | **0.07** | **0.34** | **0.06** | 0 |
| | FL194 | 0 | 0 | 0 | 0 | 0 | 0 | 0 | 0 | 0 | 0 | 0 | **0.05** | 0 | 0 | 0 | 0 | 0 | 0 |
| | FL7 | 0 | 0 | 0 | 0 | 0 | 0 | 0 | 0 | 0 | **0.03** | 0 | 0 | 0 | 0 | 0 | 0 | 0 | 0 |
| Bukit Panjang | FL647 | **0.13** | 0 | 0 | 0 | 0 | 0 | 0 | 0 | 0 | 0 | 0 | 0 | 0 | 0 | 0 | 0 | 0 | 0 |
| | FL648 | 0 | 0 | 0 | 0 | 0 | 0 | 0 | 0 | 0 | 0 | 0 | 0 | 0 | 0 | 0 | **0.19** | 0 | 0 |
| | FL659 | 0 | 0 | 0 | 0 | 0 | 0 | 0 | **0.03** | 0 | 0 | 0 | 0 | 0 | 0 | 0 | 0 | 0 | 0 |
| | FL829 | 0 | 0 | 0 | 0 | 0 | 0 | 0 | 0 | 0 | **0.06** | 0 | 0 | 0 | 0 | 0 | 0 | 0 | 0 |
| Choa Chu Kang | FL729 | 0 | 0 | 0 | 0 | 0 | 0 | 0 | 0 | 0 | 0 | 0 | 0 | 0 | 0 | 0 | 0 | **0.04** | **0.01** |
| | FL764 | 0 | 0 | 0 | 0 | 0 | 0 | **0.20** | 0 | 0 | 0 | 0 | 0 | 0 | 0 | 0 | 0 | 0 | 0 |
| | FL811 | 0 | 0 | 0 | 0 | 0 | 0 | 0 | 0 | 0 | 0 | 0 | 0 | 0 | **0.12** | 0 | 0 | 0 | 0 |
| | FL814 | 0 | 0 | 0 | 0 | 0 | 0 | 0 | 0 | 0 | 0 | 0 | 0 | 0 | 0 | 0 | **0.04** | 0 | 0 |
| | FL818 | **0.08** | 0 | 0 | 0 | 0 | **0.04** | 0 | 0 | 0 | **0.05** | 0 | 0 | 0 | 0 | **0.07** | 0 | 0 | 0 |
| Clementi | FL423 | 0 | 0 | 0 | 0 | 0 | 0 | 0 | 0 | 0 | 0 | 0 | 0 | 0 | 0 | 0 | 0 | 0 | 0 |
| | FL428 | 0 | **0.18** | 0 | 0 | 0 | 0 | 0 | 0 | 0 | 0 | 0 | 0 | 0 | 0 | 0 | 0 | 0 | 0 |
| | FL640 | 0 | 0 | 0 | 0 | 0 | 0 | 0 | 0 | 0 | 0 | 0 | 0 | 0 | 0 | 0 | 0 | 0 | 0 |
| | FL667 | 0 | 0 | 0 | 0 | 0 | 0 | 0 | **0.21** | 0 | 0 | 0 | 0 | 0 | 0 | 0 | 0 | **0.07** | 0 |
| Geylang | FL200 | 0 | 0 | 0 | **0.01** | 0 | 0 | 0 | 0 | **0.10** | 0 | **0.07** | 0 | 0 | 0 | 0 | 0 | 0 | **0.06** |
| | FL204 | 0 | 0 | 0 | 0 | 0 | 0 | 0 | 0 | 0 | 0 | 0 | 0 | 0 | 0 | 0 | **0.04** | 0 | 0 |
| | FL83 | 0 | **0.06** | 0 | 0 | 0 | 0 | 0 | 0 | 0 | 0 | **0.02** | 0 | 0 | **0.01** | 0 | **0.08** | **0.05** | 0 |
| Hougang | FL375 | 0 | 0 | 0 | 0 | 0 | 0 | 0 | 0 | 0 | 0 | **0.24** | 0 | 0 | 0 | 0 | 0 | 0 | 0 |
| | FL405 | 0 | 0 | 0 | 0 | 0 | 0 | 0 | 0 | 0 | 0 | 0 | 0 | 0 | 0 | 0 | 0 | 0 | 0 |
| | FL409 | 0 | 0 | 0 | 0 | 0 | 0 | 0 | 0 | 0 | 0 | 0 | 0 | 0 | 0 | 0 | 0 | 0 | 0 |
| | FL414 | 0 | 0 | 0 | 0 | 0 | 0 | 0 | 0 | 0 | 0 | 0 | 0 | 0 | **0.09** | 0 | 0 | 0 | 0 |
| | FL451 | 0 | 0 | 0 | 0 | 0 | 0 | 0 | 0 | 0 | 0 | **0.17** | 0 | **0.06** | 0 | 0 | **0.01** | 0 | 0 |
| | FL452 | 0 | 0 | 0 | 0 | 0 | 0 | 0 | 0 | 0 | 0 | 0 | 0 | 0 | 0 | 0 | 0 | 0 | **0.04** |
| | FL464 | 0 | 0 | 0 | 0 | 0 | 0 | 0 | 0 | 0 | 0 | 0 | 0 | 0 | 0 | **0.14** | 0 | 0 | 0 |
| | FL483 | 0 | 0 | 0 | 0 | 0 | 0 | 0 | 0 | 0 | **0.12** | 0 | **0.05** | 0 | 0 | 0 | 0 | 0 | 0 |
| | FL495 | 0 | 0 | 0 | 0 | 0 | **0.03** | 0 | 0 | 0 | 0 | 0 | 0 | 0 | 0 | 0 | 0 | 0 | 0 |
| Jurong East | FL665 | 0 | 0 | 0 | 0 | 0 | 0 | 0 | **0.28** | 0 | 0 | 0 | 0 | 0 | 0 | 0 | 0 | **0.09** | 0 |
| | FL669 | 0 | 0 | 0 | 0 | 0 | 0 | 0 | 0 | 0 | 0 | 0 | **0.01** | 0 | 0 | 0 | 0 | 0 | 0 |
| | FL697 | 0 | 0 | 0 | 0 | 0 | 0 | 0 | **0.20** | **0.13** | 0 | 0 | 0 | 0 | 0 | 0 | 0 | 0 | **0.06** |
| | FL698 | 0 | 0 | 0 | 0 | 0 | 0 | 0 | 0 | 0 | 0 | 0 | **0.01** | 0 | 0 | 0 | 0 | 0 | 0 |
| | FL701 | 0 | 0 | 0 | 0 | 0 | 0 | 0 | 0 | **0.16** | 0 | 0 | 0 | 0 | 0 | 0 | 0 | 0 | 0 |
| | FL711 | 0 | 0 | **0.15** | 0 | 0 | 0 | 0 | 0 | 0 | 0 | 0 | **0.16** | 0 | 0 | 0 | 0 | 0 | 0 |
| | FL724 | 0 | 0 | 0 | 0 | 0 | 0 | 0 | 0 | 0 | 0 | 0 | **0.08** | 0 | 0 | 0 | 0 | 0 | 0 |
| | FL725 | 0 | 0 | **0.14** | 0 | 0 | 0 | 0 | 0 | 0 | 0 | 0 | **0.12** | 0 | 0 | 0 | 0 | 0 | 0 |
| Jurong West | FL431 | 0 | 0 | 0 | 0 | 0 | 0 | 0 | 0 | 0 | 0 | **0.03** | 0 | 0 | 0 | 0 | 0 | 0 | 0 |
| | FL441 | 0 | 0 | 0 | **0.14** | 0 | **0.23** | 0 | 0 | **0.23** | 0 | 0 | 0 | 0 | 0 | 0 | 0 | 0 | **0.16** |
| | FL621 | 0 | 0 | 0 | **0.15** | **0.06** | 0 | 0 | 0 | 0 | 0 | 0 | 0 | **0.18** | **0.23** | 0 | 0 | 0 | 0 |
| | FL625 | 0 | 0 | **0.30** | 0 | 0 | 0 | 0 | 0 | 0 | 0 | 0 | **0.07** | 0 | 0 | **0.01** | 0 | 0 | 0 |
| | FL638 | 0 | 0 | 0 | 0 | 0 | 0 | **0.14** | 0 | 0 | 0 | 0 | 0 | 0 | 0 | 0 | 0 | 0 | 0 |
| | FL652 | 0 | 0 | 0 | 0 | 0 | 0 | 0 | 0 | 0 | 0 | **0.09** | 0 | 0 | 0 | **0.17** | 0 | 0 | **0.03** |
| | FL662 | 0 | 0 | 0 | 0 | 0 | 0 | 0 | 0 | 0 | **0.10** | **0.10** | 0 | 0 | 0 | **0.04** | 0 | **0.09** | **0.02** |
| | FL694 | 0 | 0 | 0 | 0 | 0 | 0 | **0.19** | 0 | 0 | 0 | 0 | 0 | 0 | 0 | 0 | 0 | 0 | 0 |
| | FL709 | 0 | 0 | 0 | 0 | 0 | 0 | 0 | 0 | 0 | 0 | 0 | 0 | 0 | 0 | 0 | 0 | 0 | 0 |
| | FL710 | 0 | 0 | 0 | 0 | 0 | 0 | 0 | 0 | 0 | 0 | 0 | 0 | 0 | 0 | 0 | 0 | **0.03** | 0 |
| | FL769 | 0 | **0.03** | 0 | 0 | 0 | 0 | 0 | 0 | 0 | 0 | 0 | 0 | 0 | 0 | 0 | 0 | 0 | 0 |
| | FL777 | 0 | 0 | 0 | 0 | 0 | 0 | 0 | 0 | **0.13** | 0 | 0 | 0 | 0 | 0 | 0 | 0 | 0 | 0 |
| Kallang | FL13 | 0 | 0 | 0 | 0 | 0 | 0 | 0 | 0 | 0 | 0 | 0 | 0 | 0 | 0 | 0 | 0 | 0 | 0 |
| | FL16 | 0 | 0 | 0 | 0 | 0 | 0 | 0 | 0 | 0 | 0 | 0 | 0 | 0 | 0 | 0 | 0 | **0.04** | 0 |
| Kallang | FL185 | 0 | 0 | 0 | 0 | 0 | 0 | 0 | 0 | 0 | 0 | **0.06** | 0 | 0 | 0 | 0 | 0 | 0 | **0.04** |
| | FL85 | 0 | 0 | 0 | 0 | 0 | 0 | **0.05** | 0 | 0 | 0 | 0 | 0 | 0 | 0 | 0 | 0 | **0.01** | 0 |
| Marine Parade | FL310 | 0 | 0 | 0 | 0 | 0 | 0 | 0 | 0 | 0 | 0 | 0 | 0 | 0 | 0 | 0 | 0 | 0 | **0.07** |
| | FL311 | **0.02** | 0 | 0 | 0 | 0 | 0 | 0 | 0 | 0 | 0 | 0 | 0 | 0 | 0 | 0 | 0 | 0 | 0 |

Continued on next page

**Table 3 – continued from previous page**

| Donor | | Model 3 | | | | | | | | | Model 4 | | | | | | | | |
|---|---|---|---|---|---|---|---|---|---|---|---|---|---|---|---|---|---|---|---|
| Town | Sector | FL424 | FL425 | FL426 | FL633 | FL634 | FL641 | FL655 | FL833 | FL833b | FL424 | FL425 | FL426 | FL633 | FL634 | FL641 | FL655 | FL833 | FL833b |
| | FL318 | 0 | 0 | 0 | 0 | 0 | 0 | 0 | 0 | 0 | **0.15** | 0 | 0 | 0 | 0 | 0 | 0 | 0 | 0 |
| Novena | FL68 | 0 | 0 | 0 | 0 | 0 | 0 | 0 | **0.21** | 0 | **0.18** | 0 | 0 | 0 | 0 | 0 | 0 | **0.03** | 0 |
| Pasir Ris | FL166 | 0 | 0 | 0 | 0 | 0 | 0 | **0.02** | 0 | 0 | 0 | 0 | 0 | 0 | **0.01** | 0 | 0 | **0.05** | 0 |
| | FL217 | 0 | 0 | 0 | 0 | 0 | 0 | 0 | 0 | 0 | 0 | 0 | 0 | 0 | 0 | 0 | 0 | 0 | 0 |
| | FL228 | 0 | 0 | 0 | 0 | 0 | 0 | 0 | 0 | 0 | 0 | 0 | 0 | 0 | 0 | 0 | 0 | 0 | 0 |
| | FL256 | 0 | 0 | 0 | 0 | 0 | 0 | 0 | 0 | 0 | 0 | 0 | 0 | 0 | 0 | 0 | 0 | 0 | **0.03** |
| | FL288 | 0 | 0 | 0 | 0 | 0 | 0 | 0 | 0 | 0 | 0 | 0 | 0 | 0 | 0 | **0.06** | 0 | 0 | 0 |
| | FL293 | 0 | 0 | 0 | 0 | **0.01** | 0 | 0 | 0 | 0 | 0 | 0 | 0 | 0 | 0 | 0 | 0 | 0 | 0 |
| | FL294 | 0 | 0 | 0 | 0 | 0 | 0 | 0 | 0 | 0 | 0 | 0 | 0 | 0 | 0 | **0.01** | 0 | 0 | 0 |
| | FL336 | 0 | 0 | 0 | 0 | 0 | 0 | 0 | 0 | 0 | 0 | 0 | **0.08** | 0 | 0 | 0 | 0 | 0 | 0 |
| | FL348 | 0 | 0 | 0 | 0 | 0 | 0 | 0 | 0 | 0 | 0 | 0 | 0 | 0 | **0.01** | 0 | 0 | 0 | 0 |
| Queenstown | FL114 | 0 | 0 | 0 | 0 | 0 | 0 | 0 | 0 | 0 | 0 | **0.01** | 0 | 0 | 0 | 0 | **0.15** | 0 | 0 |
| | FL31 | 0 | 0 | 0 | 0 | 0 | 0 | **0.38** | **0.07** | 0 | 0 | 0 | 0 | 0 | 0 | 0 | 0 | **0.01** | 0 |
| | FL352 | 0 | 0 | 0 | 0 | 0 | 0 | 0 | 0 | 0 | 0 | 0 | 0 | 0 | 0 | 0 | **0.04** | 0 | 0 |
| Rochor | FL73 | 0 | 0 | 0 | 0 | 0 | **0.12** | 0 | 0 | 0 | 0 | 0 | **0.09** | 0 | 0 | **0.01** | 0 | **0.03** | 0 |
| Sembawang | FL527 | 0 | 0 | 0 | 0 | 0 | 0 | 0 | 0 | 0 | 0 | 0 | 0 | 0 | 0 | 0 | 0 | 0 | **0.15** |
| Sengkang | FL133 | 0 | 0 | 0 | 0 | 0 | 0 | 0 | 0 | 0 | 0 | 0 | 0 | 0 | 0 | **0.03** | 0 | 0 | 0 |
| | FL368 | 0 | 0 | 0 | 0 | 0 | 0 | 0 | 0 | 0 | 0 | 0 | 0 | 0 | 0 | **0.10** | 0 | 0 | 0 |
| | FL369 | 0 | 0 | 0 | 0 | 0 | 0 | 0 | 0 | 0 | **0.02** | 0 | 0 | 0 | 0 | **0.20** | 0 | 0 | 0 |
| | FL497 | 0 | 0 | 0 | 0 | 0 | 0 | 0 | 0 | 0 | 0 | **0.02** | 0 | 0 | 0 | 0 | 0 | 0 | 0 |
| | FL504 | 0 | 0 | 0 | 0 | 0 | 0 | 0 | 0 | 0 | 0 | 0 | 0 | 0 | 0 | 0 | 0 | **0.06** | 0 |
| | FL538 | **0.08** | 0 | 0 | 0 | 0 | 0 | 0 | 0 | 0 | 0 | 0 | 0 | 0 | 0 | 0 | 0 | 0 | 0 |
| | FL733 | 0 | 0 | 0 | 0 | 0 | 0 | 0 | 0 | 0 | 0 | **0.09** | 0 | 0 | 0 | 0 | 0 | 0 | 0 |
| Serangoon | FL392 | 0 | 0 | **0.16** | 0 | 0 | 0 | 0 | 0 | 0 | 0 | 0 | 0 | 0 | 0 | 0 | 0 | 0 | 0 |
| | FL393 | 0 | 0 | 0 | 0 | 0 | 0 | 0 | 0 | 0 | 0 | 0 | **0.05** | 0 | 0 | 0 | 0 | 0 | 0 |
| | FL480 | 0 | 0 | 0 | 0 | 0 | 0 | 0 | 0 | 0 | 0 | 0 | **0.06** | 0 | 0 | 0 | 0 | 0 | 0 |
| Toa Payoh | FL164 | 0 | 0 | 0 | **0.18** | 0 | 0 | 0 | 0 | 0 | 0 | 0 | 0 | **0.01** | **0.03** | 0 | 0 | 0 | 0 |
| | FL319 | 0 | 0 | 0 | 0 | 0 | 0 | 0 | 0 | 0 | 0 | **0.09** | 0 | 0 | 0 | 0 | 0 | 0 | 0 |
| | FL40 | 0 | 0 | 0 | 0 | 0 | 0 | 0 | 0 | 0 | 0 | 0 | 0 | 0 | 0 | 0 | 0 | **0.03** | 0 |
| | FL44 | 0 | 0 | 0 | 0 | 0 | 0 | 0 | 0 | 0 | 0 | 0 | 0 | 0 | 0 | **0.05** | 0 | 0 | 0 |
| | FL58 | 0 | 0 | 0 | 0 | 0 | 0 | 0 | 0 | 0 | 0 | 0 | 0 | 0 | 0 | 0 | 0 | **0.13** | 0 |
| | FL59 | 0 | 0 | 0 | 0 | 0 | 0 | 0 | 0 | 0 | 0 | 0 | 0 | 0 | 0 | 0 | 0 | 0 | **0.13** |
| | FL61 | 0 | 0 | 0 | 0 | 0 | 0 | 0 | 0 | 0 | **0.03** | 0 | 0 | 0 | 0 | **0.02** | 0 | 0 | 0 |
| Woodlands | FL247 | 0 | 0 | **0.02** | 0 | 0 | 0 | 0 | 0 | 0 | 0 | 0 | **0.03** | 0 | 0 | 0 | 0 | 0 | 0 |
| | FL253 | 0 | 0 | 0 | 0 | 0 | 0 | 0 | 0 | 0 | 0 | 0 | 0 | 0 | 0 | 0 | 0 | 0 | **0.01** |
| | FL254 | 0 | 0 | 0 | 0 | 0 | 0 | 0 | 0 | 0 | **0.05** | 0 | **0.04** | 0 | 0 | 0 | 0 | 0 | 0 |
| | FL262 | 0 | 0 | 0 | 0 | 0 | 0 | 0 | 0 | 0 | 0 | 0 | 0 | 0 | **0.02** | 0 | 0 | 0 | 0 |
| | FL508 | 0 | 0 | 0 | 0 | 0 | 0 | 0 | 0 | 0 | 0 | 0 | 0 | 0 | 0 | 0 | 0 | 0 | **0.03** |
| | FL572 | 0 | 0 | 0 | 0 | 0 | 0 | 0 | 0 | 0 | **0.01** | 0 | 0 | 0 | 0 | 0 | 0 | 0 | 0 |
| | FL575 | 0 | 0 | 0 | 0 | 0 | 0 | 0 | 0 | 0 | 0 | 0 | **0.03** | 0 | 0 | 0 | 0 | 0 | 0 |
| | FL580 | 0 | 0 | 0 | 0 | 0 | 0 | 0 | 0 | 0 | **0.02** | 0 | 0 | 0 | 0 | 0 | 0 | 0 | 0 |
| | FL587 | 0 | 0 | 0 | 0 | 0 | 0 | 0 | 0 | 0 | 0 | 0 | 0 | 0 | 0 | 0 | 0 | 0 | 0 |
| | FL591 | **0.27** | 0 | 0 | 0 | 0 | 0 | 0 | 0 | 0 | **0.08** | 0 | 0 | 0 | 0 | 0 | 0 | 0 | 0 |
| | FL600 | 0 | 0 | 0 | 0 | 0 | 0 | 0 | 0 | 0 | 0 | 0 | **0.05** | 0 | 0 | **0.03** | 0 | 0 | 0 |

Table 4: Donor weights for Choa Chu Kang sectors for model 2. Only sectors with any contribution to synthetic control weights for intervention sectors' synthetic controls are shown

| Donor | | Model 2 | | | | | | | | | | | | | | |
|---|---|---|---|---|---|---|---|---|---|---|---|---|---|---|---|---|
| Town | Sector | FL635 | FL636 | FL637 | FL650 | FL718 | FL740 | FL742 | FL748 | FL765 | FL768 | FL803 | FL804 | FL806 | FL807 | FL826 |
| Ang Mo Kio | FL454 | 0 | **0.18** | 0 | 0 | 0 | 0 | 0 | 0 | 0 | 0 | 0 | 0 | 0 | 0 | 0 |
| | FL499 | 0 | 0 | 0 | 0 | 0 | **0.28** | 0 | 0 | 0 | 0 | 0 | 0 | 0 | 0 | **0.16** |
| Bedok | FL237 | 0 | 0 | 0 | 0 | 0 | 0.0 | 0 | 0 | 0 | 0 | 0 | **0.12** | 0 | 0 | 0 |
| Bishan | FL34 | 0 | 0 | 0 | **0.21** | 0 | 0 | 0 | 0 | 0 | 0 | 0 | **0.26** | 0 | 0 | 0 |
| Bukit Batok | FL434 | 0 | 0 | 0 | 0 | 0 | 0 | 0 | **0.06** | 0 | 0 | 0 | **0.22** | 0 | 0 | 0 |
| | FL686 | 0 | 0 | 0 | 0 | 0 | 0 | 0 | **0.01** | 0 | 0 | 0 | 0 | 0 | 0 | 0 |
| | FL699 | 0 | 0 | 0 | **0.26** | 0 | 0 | 0 | **0.40** | 0 | 0 | 0 | **0.06** | **0.40** | 0 | 0 |
| | FL731 | 0 | 0 | 0 | 0 | 0 | 0 | 0 | **0.20** | 0 | 0 | 0 | 0 | 0 | 0 | 0 |
| Bukit Merah | FL193 | **0.16** | 0 | 0 | 0 | 0 | 0 | 0 | 0 | 0 | 0 | 0 | 0 | 0 | 0 | 0 |
| Bukit Panjang | FL647 | 0 | 0 | 0 | 0 | 0 | 0 | 0 | 0 | 0 | 0 | 0 | 0 | 0 | **0.04** | 0 |
| | FL659 | 0 | 0 | 0 | 0 | **0.24** | 0 | 0 | 0 | **0.42** | **0.04** | 0 | 0 | 0 | 0 | 0 |
| Choa Chu Kang | FL729 | 0 | **0.09** | **0.14** | **0.33** | 0 | 0 | **0.04** | 0 | **0.07** | 0 | 0 | **0.04** | 0 | 0 | **0.04** |
| | FL764 | **0.26** | 0 | **0.20** | 0 | 0 | 0 | 0 | 0 | 0 | 0 | 0 | 0 | 0 | 0 | **0.11** |

Continued on next page

**Table 4 – continued from previous page**

| Donor | | Model 2 | | | | | | | | | | | | | | |
|---|---|---|---|---|---|---|---|---|---|---|---|---|---|---|---|---|
| Town | Sector | FL635 | FL636 | FL637 | FL650 | FL718 | FL740 | FL742 | FL748 | FL765 | FL768 | FL803 | FL804 | FL806 | FL807 | FL826 |
| | FL811 | 0 | **0.06** | 0 | 0 | 0 | 0 | **0.06** | 0 | 0 | 0 | 0 | 0 | **0.02** | 0 | 0 |
| | FL814 | **0.01** | 0 | **0.10** | 0 | 0 | 0 | 0 | 0 | 0 | 0 | 0 | 0 | 0 | 0 | 0 |
| | FL818 | 0 | 0 | 0 | 0 | **0.38** | 0 | 0 | 0 | 0 | 0 | 0 | 0 | 0 | **0.06** | 0 |
| | FL823 | 0 | 0 | 0 | 0 | 0 | 0 | **0.36** | 0 | 0 | 0 | 0 | 0 | **0.19** | 0 | 0 |
| Clementi | FL428 | 0 | 0 | 0 | 0 | 0 | 0 | 0 | **0.01** | 0 | 0 | 0 | 0 | 0 | 0 | 0 |
| | FL640 | **0.12** | 0 | 0 | 0 | 0 | 0 | 0 | 0 | **0.15** | 0 | 0 | 0 | 0 | 0 | 0 |
| Geylang | FL19 | 0 | 0 | 0 | 0 | 0 | 0 | 0 | 0 | 0 | 0 | **0.13** | 0 | 0 | 0 | 0 |
| | FL200 | 0 | **0.21** | **0.26** | 0 | 0 | 0 | 0 | **0.03** | 0 | 0 | 0 | 0 | 0 | 0 | 0 |
| Hougang | FL375 | 0 | 0 | 0 | 0 | 0 | **0.18** | 0 | 0 | 0 | 0 | 0 | 0 | 0 | 0 | 0 |
| | FL451 | 0 | 0 | 0 | 0 | 0 | 0 | 0 | 0 | 0 | 0 | 0 | 0 | 0 | 0 | **0.23** |
| | FL464 | 0 | 0 | 0 | 0 | 0 | 0 | 0 | 0 | 0 | 0 | **0.40** | 0 | 0 | 0 | 0 |
| | FL489 | 0 | 0 | 0 | 0 | **0.09** | 0 | 0 | 0 | 0 | 0 | 0 | 0 | 0 | 0 | 0 |
| | FL495 | 0 | 0 | 0 | 0 | 0 | 0 | **0.01** | 0 | 0 | 0 | 0 | 0 | 0 | 0 | 0 |
| Jurong East | FL2 | 0 | 0 | 0 | 0 | 0 | 0 | 0 | 0 | **0.18** | 0 | 0 | 0 | 0 | 0 | 0 |
| | FL665 | 0 | 0 | 0 | 0 | 0 | 0 | 0 | **0.07** | 0 | **0.06** | **0.47** | 0 | 0 | 0 | 0 |
| | FL697 | 0 | 0 | **0.14** | 0 | 0 | **0.15** | 0 | 0 | 0 | 0 | 0 | 0 | 0 | 0 | 0 |
| | FL725 | 0 | 0 | **0.12** | 0 | 0 | 0 | 0 | 0 | 0 | 0 | 0 | 0 | 0 | 0 | 0 |
| Jurong West | FL430 | 0 | **0.05** | 0 | 0 | 0 | 0 | 0 | 0 | 0 | 0 | 0 | 0 | 0 | 0 | 0 |
| | FL431 | 0 | 0 | 0 | 0 | 0 | 0 | 0 | 0 | 0 | **0.11** | 0 | 0 | 0 | 0 | 0 |
| | FL438 | 0 | 0 | **0.02** | 0 | 0 | 0 | 0 | 0 | 0 | 0 | 0 | 0 | 0 | 0 | 0 |
| | FL442 | 0 | **0.08** | 0 | 0 | 0 | 0 | 0 | 0 | 0 | 0 | 0 | 0 | 0 | 0 | 0 |
| | FL621 | 0 | 0 | 0 | **0.20** | 0 | 0 | 0 | **0.07** | 0 | 0 | 0 | 0 | 0 | 0 | 0 |
| | FL652 | 0 | **0.13** | 0 | 0 | 0 | 0 | 0 | 0 | 0 | 0 | 0 | 0 | 0 | 0 | 0 |
| | FL662 | 0 | 0 | 0 | 0 | 0 | 0 | 0 | 0 | 0 | **0.14** | 0 | 0 | 0 | 0 | 0 |
| | FL694 | **0.27** | 0 | 0 | 0 | 0 | 0 | 0 | 0 | 0 | 0 | 0 | 0 | 0 | 0 | 0 |
| | FL709 | 0 | 0 | 0 | 0 | 0 | **0.20** | 0 | 0 | 0 | 0 | 0 | 0 | 0 | 0 | 0 |
| | FL710 | 0 | 0 | 0 | 0 | 0 | 0 | 0 | 0 | 0 | **0.05** | 0 | 0 | 0 | 0 | 0 |
| | FL769 | 0 | 0 | 0 | 0 | 0 | 0 | 0 | 0 | 0 | **0.11** | 0 | 0 | 0 | 0 | 0 |
| Kallang | FL14 | 0 | 0 | 0 | 0 | 0 | 0 | 0 | 0 | 0 | 0 | 0 | **0.20** | 0 | 0 | 0 |
| | FL85 | 0 | 0 | 0 | 0 | 0 | 0 | 0 | 0 | 0 | 0 | 0 | 0 | **0.13** | 0 | 0 |
| | FL93 | 0 | 0 | 0 | 0 | 0 | 0 | 0 | 0 | **0.01** | 0 | 0 | 0 | 0 | 0 | 0 |
| Marine Parade | FL310 | 0 | 0 | 0 | 0 | 0 | 0 | 0 | 0 | 0 | 0 | 0 | 0 | 0 | **0.09** | 0 |
| | FL318 | 0 | **0.10** | 0 | 0 | 0 | 0 | 0 | 0 | 0 | 0 | 0 | 0 | 0 | 0 | 0 |
| Pasir Ris | FL227 | 0 | 0 | 0 | 0 | 0 | 0 | 0 | 0 | 0 | **0.13** | 0 | 0 | 0 | 0 | 0 |
| | FL321 | 0 | 0 | 0 | 0 | 0 | 0 | **0.12** | 0 | 0 | 0 | 0 | 0 | 0 | 0 | 0 |
| Queenstown | FL339 | 0 | **0.09** | 0 | 0 | 0 | 0 | 0 | 0 | 0 | 0 | 0 | 0 | 0 | 0 | 0 |
| | FL352 | 0 | 0 | 0 | 0 | 0 | 0 | **0.01** | 0 | 0 | 0 | 0 | 0 | **0.01** | 0 | 0 |
| Rochor | FL72 | 0 | 0 | 0 | 0 | 0 | 0 | 0 | 0 | 0 | **0.16** | 0 | **0.10** | 0 | 0 | 0 |
| Sembawang | FL527 | 0 | 0 | 0 | 0 | 0 | 0 | 0 | 0 | 0 | **0.13** | 0 | 0 | 0 | **0.08** | 0 |
| Sengkang | FL132 | 0 | 0 | 0 | 0 | 0 | 0 | 0 | 0 | **0.05** | 0 | 0 | 0 | 0 | 0 | 0 |
| | FL135 | 0 | 0 | 0 | 0 | 0 | 0 | 0 | 0 | 0 | 0 | 0 | 0 | 0 | **0.06** | 0 |
| | FL368 | 0 | 0 | 0 | 0 | 0 | 0 | **0.11** | 0 | 0 | 0 | 0 | 0 | 0 | 0 | 0 |
| | FL492 | **0.01** | 0 | 0 | 0 | 0 | 0 | 0 | 0 | 0 | 0 | 0 | 0 | 0 | 0 | 0 |
| | FL497 | **0.17** | 0 | 0 | 0 | 0 | 0 | 0 | 0 | 0 | 0 | 0 | 0 | 0 | 0 | 0 |
| | FL733 | 0 | 0 | 0 | 0 | 0 | 0 | 0 | 0 | 0 | 0 | 0 | 0 | 0 | **0.15** | 0 |
| Serangoon | FL461 | 0 | 0 | 0 | 0 | 0 | 0 | 0 | 0 | 0 | **0.04** | 0 | 0 | 0 | 0 | 0 |
| Toa Payoh | FL59 | 0 | 0 | 0 | 0 | 0 | **0.18** | 0 | 0 | 0 | 0 | 0 | 0 | 0 | 0 | 0 |
| Woodlands | FL226 | 0 | 0 | 0 | 0 | 0 | 0 | 0 | 0 | 0 | 0 | 0 | 0 | **0.14** | 0 | **0.02** |
| | FL232 | 0 | 0 | 0 | 0 | 0 | 0 | 0 | 0 | 0 | 0 | 0 | 0 | 0 | 0 | **0.11** |
| | FL252 | 0 | 0 | 0 | 0 | 0 | 0 | 0 | 0 | 0 | 0 | 0 | 0 | **0.08** | 0 | 0 |
| | FL253 | 0 | 0 | 0 | 0 | 0 | 0 | 0 | 0 | **0.13** | 0 | 0 | 0 | 0 | 0 | 0 |
| | FL262 | 0 | 0 | 0 | 0 | 0 | 0 | 0 | 0 | 0 | 0 | 0 | 0 | 0 | **0.29** | 0 |
| | FL572 | 0 | **0.01** | 0 | 0 | 0 | 0 | 0 | 0 | 0 | 0 | 0 | 0 | 0 | 0 | 0 |
| | FL587 | 0 | 0 | 0 | 0 | 0 | 0 | 0 | 0 | 0 | 0 | 0 | 0 | **0.04** | 0 | 0 |
| | FL597 | 0 | 0 | 0 | 0 | 0 | 0 | 0 | 0 | 0 | 0 | 0 | 0 | 0 | 0 | **0.15** |
| | FL598 | 0 | 0 | 0 | 0 | 0 | 0 | 0 | **0.16** | 0 | 0 | 0 | 0 | 0 | 0 | **0.18** |
| | FL600 | 0 | 0 | 0 | 0 | **0.06** | 0 | 0 | 0 | 0 | **0.03** | 0 | 0 | 0 | 0 | 0 |
| | FL607 | 0 | 0 | 0 | 0 | **0.22** | 0 | 0 | 0 | 0 | 0 | 0 | 0 | 0 | 0 | 0 |
| | FL619 | 0 | 0 | 0 | 0 | **0.02** | 0 | 0 | 0 | 0 | 0 | 0 | 0 | 0 | **0.22** | 0 |
| | FL830 | 0 | 0 | 0 | 0 | 0 | 0 | **0.29** | 0 | 0 | 0 | 0 | 0 | 0 | 0 | 0 |

Table 5: Donor weights for Choa Chu Kang sectors for models 3 and 4. Only sectors with any contribution to synthetic control weights for intervention sectors' synthetic controls are shown

| Donor | | Model 3 | | | | | | | | | Model 4 | | | | | | | | |
|---|---|---|---|---|---|---|---|---|---|---|---|---|---|---|---|---|---|---|---|
| Town | Sector | FL635 | FL636 | FL637 | FL650 | FL718 | FL740 | FL742 | FL748 | FL765 | FL635 | FL636 | FL637 | FL650 | FL718 | FL740 | FL742 | FL748 | FL765 |
| Ang Mo Kio | FL17 | 0 | 0 | 0 | 0 | 0 | 0 | 0 | 0 | 0 | 0 | 0 | 0 | 0 | 0 | 0 | **0.01** | 0 | 0 |
| | FL372 | 0 | 0 | 0 | 0 | 0 | 0 | 0 | 0 | 0 | 0 | 0 | 0 | 0 | 0 | **0.07** | 0 | 0 | 0 |
| | FL454 | 0 | **0.14** | 0 | 0 | 0 | 0 | 0 | 0 | 0 | 0 | 0 | 0 | 0 | 0 | 0 | 0 | 0 | 0 |
| | FL456 | 0 | 0 | 0 | 0 | 0 | 0 | 0 | 0 | 0 | 0 | 0 | 0 | 0 | **0.06** | 0 | 0 | 0 | **0.05** |
| | FL499 | **0.02** | 0 | 0 | 0 | 0 | 0 | 0 | 0 | 0 | 0 | 0 | 0 | 0 | 0 | **0.16** | 0 | 0 | 0 |
| Bedok | FL131 | 0 | 0 | 0 | 0 | 0 | 0 | 0 | 0 | 0 | **0.07** | 0 | 0 | 0 | 0 | 0 | 0 | **0.01** | 0 |
| | FL225 | 0 | 0 | 0 | 0 | 0 | 0 | 0 | 0 | 0 | 0 | 0 | 0 | 0 | **0.03** | 0 | 0 | 0 | 0 |
| | FL237 | 0 | 0 | 0 | 0 | 0 | 0 | 0 | 0 | 0 | 0 | 0 | 0 | 0 | 0 | 0 | 0 | **0.06** | **0.02** |
| | FL238 | 0 | 0 | 0 | 0 | 0 | 0 | 0 | 0 | 0 | 0 | 0 | **0.04** | 0 | 0 | 0 | 0 | 0 | **0.01** |
| Bishan | FL34 | 0 | 0 | 0 | **0.14** | 0 | 0 | 0 | 0 | 0 | 0 | 0 | 0 | **0.26** | 0 | 0 | 0 | 0 | 0 |
| | FL47 | 0 | 0 | 0 | 0 | 0 | **0.14** | 0 | 0 | **0.04** | 0 | 0 | 0 | 0 | 0 | **0.06** | 0 | 0 | 0 |
| | FL51 | 0 | 0 | 0 | 0 | 0 | 0 | 0 | 0 | 0 | 0 | 0 | **0.01** | 0 | 0 | 0 | 0 | 0 | **0.01** |
| Bukit Batok | FL434 | 0 | 0 | 0 | 0 | 0 | 0 | 0 | **0.09** | 0 | 0 | **0.33** | **0.06** | 0 | 0 | 0 | 0 | **0.07** | 0 |
| | FL680 | 0 | 0 | 0 | 0 | **0.13** | 0 | 0 | 0 | 0 | 0 | 0 | 0 | 0 | **0.05** | 0 | 0 | 0 | 0 |
| | FL686 | 0 | 0 | 0 | 0 | 0 | 0 | 0 | 0 | 0 | 0 | 0 | **0.18** | 0 | 0 | 0 | 0 | 0 | 0 |
| | FL699 | 0 | 0 | 0 | **0.19** | 0 | 0 | 0 | **0.43** | 0 | 0 | 0 | 0 | **0.14** | 0 | 0 | 0 | **0.42** | 0 |
| | FL731 | 0 | 0 | 0 | 0 | 0 | 0 | 0 | **0.33** | 0 | 0 | 0 | 0 | 0 | 0 | 0 | 0 | **0.30** | 0 |
| Bukit Merah | FL147 | 0 | 0 | 0 | 0 | 0 | 0 | 0 | 0 | 0 | 0 | **0.18** | 0 | 0 | 0 | 0 | 0 | 0 | 0 |
| | FL160 | 0 | 0 | 0 | 0 | 0 | 0 | 0 | 0 | 0 | 0 | 0 | 0 | 0 | 0 | 0 | **0.04** | 0 | 0 |
| | FL174 | 0 | 0 | 0 | 0 | 0 | 0 | 0 | 0 | 0 | 0 | **0.02** | 0 | 0 | 0 | 0 | 0 | 0 | 0 |
| | FL193 | 0 | 0 | 0 | 0 | 0 | 0 | 0 | 0 | 0 | 0 | 0 | **0.05** | 0 | 0 | 0 | 0 | 0 | **0.06** |
| | FL194 | 0 | 0 | 0 | 0 | 0 | 0 | 0 | 0 | 0 | 0 | 0 | 0 | 0 | 0 | 0 | **0.14** | 0 | 0 |
| | FL7 | 0 | 0 | 0 | 0 | 0 | 0 | 0 | 0 | 0 | 0 | **0.01** | 0 | 0 | 0 | 0 | 0 | 0 | 0 |
| | FL8 | 0 | 0 | 0 | 0 | 0 | 0 | 0 | 0 | 0 | **0.09** | 0 | **0.02** | 0 | 0 | 0 | 0 | 0 | 0 |
| Bukit Panjang | FL648 | **0.08** | 0 | 0 | 0 | 0 | 0 | 0 | 0 | 0 | 0 | 0 | 0 | 0 | 0 | 0 | 0 | 0 | 0 |
| | FL659 | **0.01** | 0 | 0 | 0 | **0.18** | 0 | 0 | 0 | **0.20** | 0 | 0 | 0 | 0 | 0 | 0 | 0 | 0 | **0.11** |
| | FL829 | 0 | 0 | **0.03** | 0 | 0 | 0 | 0 | 0 | 0 | 0 | 0 | 0 | 0 | 0 | 0 | 0 | 0 | 0 |
| Choa Chu Kang | FL729 | 0 | **0.21** | **0.12** | **0.23** | **0.02** | 0 | **0.03** | 0 | 0 | 0 | 0 | 0 | **0.20** | 0 | 0 | 0 | 0 | 0 |
| | FL764 | 0 | **0.01** | **0.21** | 0 | 0 | 0 | 0 | 0 | 0 | 0 | 0 | **0.04** | 0 | 0 | 0 | 0 | 0 | 0 |
| | FL811 | 0 | **0.06** | **0.03** | 0 | 0 | 0 | **0.22** | 0 | 0 | 0 | 0 | **0.02** | 0 | 0 | 0 | 0 | 0 | 0 |
| | FL814 | 0 | 0 | 0 | 0 | 0 | 0 | 0 | 0 | 0 | **0.01** | 0 | 0 | 0 | 0 | 0 | 0 | 0 | 0 |
| | FL818 | 0 | 0 | **0.02** | 0 | **0.61** | 0 | 0 | 0 | **0.10** | 0 | 0 | 0 | 0 | **0.49** | 0 | 0 | 0 | 0 |
| | FL823 | 0 | **0.04** | 0 | 0 | 0 | 0 | **0.27** | 0 | 0 | 0 | 0 | 0 | 0 | 0 | 0 | **0.10** | 0 | 0 |
| Geylang | FL200 | 0 | 0 | **0.27** | 0 | 0 | 0 | 0 | **0.04** | 0 | 0 | 0 | **0.03** | 0 | 0 | 0 | 0 | 0 | 0 |
| | FL81 | 0 | 0 | 0 | 0 | 0 | 0 | 0 | 0 | 0 | 0 | 0 | 0 | 0 | 0 | 0 | **0.16** | 0 | 0 |
| | FL83 | 0 | 0 | 0 | 0 | 0 | 0 | 0 | 0 | 0 | **0.01** | 0 | 0 | 0 | 0 | 0 | 0 | 0 | 0 |
| Hougang | FL375 | **0.14** | 0 | 0 | 0 | 0 | 0 | 0 | 0 | **0.13** | **0.06** | 0 | 0 | 0 | 0 | 0 | 0 | 0 | 0 |
| | FL451 | **0.11** | 0 | 0 | 0 | 0 | 0 | 0 | **0.06** | 0 | **0.13** | 0 | 0 | 0 | 0 | 0 | 0 | **0.02** | 0 |
| | FL452 | 0 | 0 | 0 | 0 | 0 | 0 | 0 | 0 | **0.14** | 0 | **0.03** | **0.04** | 0 | 0 | 0 | 0 | 0 | **0.05** |
| | FL495 | 0 | 0 | 0 | 0 | **0.01** | 0 | 0 | 0 | 0 | 0 | 0 | 0 | 0 | 0 | 0 | 0 | 0 | 0 |
| Jurong East | FL665 | 0 | 0 | 0 | 0 | 0 | 0 | 0 | 0 | 0 | 0 | 0 | 0 | 0 | 0 | 0 | 0 | 0 | **0.19** |
| | FL668 | 0 | **0.13** | 0 | 0 | 0 | 0 | 0 | 0 | 0 | 0 | **0.17** | 0 | 0 | 0 | 0 | 0 | 0 | 0 |
| | FL697 | 0 | 0 | **0.28** | **0.07** | 0 | **0.04** | 0 | 0 | 0 | 0 | 0 | **0.19** | **0.06** | 0 | **0.03** | 0 | **0.02** | 0 |
| | FL698 | 0 | 0 | 0 | 0 | 0 | 0 | 0 | 0 | 0 | 0 | 0 | 0 | 0 | 0 | 0 | **0.05** | 0 | 0 |
| | FL701 | 0 | 0 | 0 | 0 | 0 | 0 | 0 | 0 | 0 | 0 | 0 | 0 | 0 | 0 | 0 | 0 | 0 | **0.04** |
| | FL723 | 0 | 0 | 0 | 0 | 0 | 0 | 0 | 0 | 0 | 0 | 0 | 0 | 0 | **0.05** | 0 | 0 | 0 | 0 |
| Jurong West | FL430 | 0 | **0.10** | 0 | 0 | **0.05** | 0 | 0 | 0 | 0 | 0 | 0 | 0 | 0 | **0.02** | 0 | 0 | 0 | 0 |
| | FL441 | 0 | 0 | 0 | 0 | 0 | **0.09** | 0 | 0 | 0 | **0.06** | 0 | 0 | 0 | 0 | 0 | 0 | 0 | 0 |
| | FL442 | 0 | 0 | 0 | 0 | 0 | 0 | 0 | 0 | 0 | 0 | **0.04** | 0 | 0 | 0 | 0 | 0 | 0 | 0 |
| | FL621 | 0 | 0 | 0 | **0.34** | 0 | 0 | 0 | 0 | 0 | 0 | 0 | 0 | **0.31** | 0 | 0 | 0 | 0 | 0 |
| | FL625 | 0 | 0 | 0 | 0 | 0 | 0 | **0.03** | 0 | 0 | 0 | 0 | 0 | 0 | 0 | 0 | 0 | 0 | 0 |
| | FL652 | 0 | **0.29** | 0 | 0 | 0 | **0.13** | 0 | 0 | 0 | **0.02** | **0.06** | **0.03** | 0 | 0 | **0.13** | 0 | **0.01** | 0 |
| | FL662 | 0 | 0 | 0 | 0 | 0 | 0 | 0 | 0 | 0 | 0 | 0 | 0 | 0 | 0 | 0 | **0.23** | 0 | 0 |
| | FL663 | 0 | 0 | 0 | 0 | 0 | 0 | 0 | 0 | 0 | 0 | 0 | 0 | 0 | **0.18** | 0 | 0 | 0 | 0 |
| | FL664 | 0 | 0 | 0 | 0 | 0 | 0 | 0 | 0 | 0 | 0 | **0.02** | 0 | 0 | 0 | 0 | 0 | 0 | 0 |
| | FL709 | 0 | 0 | 0 | 0 | 0 | 0 | 0 | 0 | 0 | 0 | 0 | 0 | 0 | 0 | **0.04** | 0 | 0 | 0 |
| | FL710 | 0 | 0 | 0 | 0 | 0 | 0 | 0 | 0 | **0.24** | 0 | 0 | 0 | 0 | 0 | 0 | 0 | 0 | **0.07** |
| | FL769 | 0 | 0 | 0 | 0 | 0 | **0.31** | 0 | 0 | 0 | 0 | 0 | 0 | 0 | 0 | **0.14** | 0 | 0 | 0 |
| Kallang | FL15 | 0 | 0 | 0 | 0 | 0 | 0 | 0 | 0 | 0 | 0 | **0.06** | 0 | 0 | 0 | 0 | 0 | 0 | 0 |
| | FL16 | 0 | 0 | 0 | 0 | 0 | 0 | 0 | 0 | 0 | 0 | 0 | **0.01** | 0 | **0.10** | 0 | 0 | 0 | 0 |
| | FL182 | 0 | 0 | 0 | 0 | 0 | 0 | 0 | 0 | 0 | 0 | 0 | 0 | 0 | 0 | 0 | **0.04** | 0 | 0 |
| | FL183 | 0 | 0 | 0 | 0 | 0 | 0 | 0 | 0 | 0 | 0 | 0 | 0 | 0 | 0 | 0 | **0.11** | 0 | 0 |
| | FL185 | 0 | 0 | 0 | **0.02** | 0 | **0.06** | 0 | 0 | **0.09** | 0 | 0 | **0.01** | 0 | 0 | **0.02** | 0 | 0 | **0.06** |
| Marine Parade | FL310 | 0 | 0 | 0 | 0 | 0 | 0 | 0 | 0 | 0 | 0 | 0 | 0 | 0 | 0 | 0 | 0 | **0.02** | **0.02** |
| Novena | FL68 | 0 | 0 | 0 | 0 | 0 | 0 | 0 | 0 | 0 | **0.28** | 0 | 0 | 0 | 0 | 0 | 0 | 0 | 0 |
| | FL70 | 0 | 0 | 0 | 0 | 0 | 0 | 0 | 0 | 0 | 0 | 0 | **0.04** | 0 | 0 | 0 | 0 | 0 | 0 |
| | FL166 | 0 | 0 | 0 | 0 | 0 | 0 | 0 | 0 | 0 | 0 | 0 | 0 | **0.03** | 0 | 0 | 0 | 0 | 0 |
| | FL217 | 0 | 0 | 0 | 0 | 0 | 0 | 0 | 0 | 0 | **0.14** | 0 | 0 | 0 | 0 | 0 | 0 | 0 | 0 |

Pasir Ris

Continued on next page

**Table 5 – continued from previous page**

| Donor | | Model 3 | | | | | | | | | Model 4 | | | | | | | | |
|---|---|---|---|---|---|---|---|---|---|---|---|---|---|---|---|---|---|---|---|
| Town | Sector | FL635 | FL636 | FL637 | FL650 | FL718 | FL740 | FL742 | FL748 | FL765 | FL635 | FL636 | FL637 | FL650 | FL718 | FL740 | FL742 | FL748 | FL765 |
| | FL256 | 0 | 0 | 0 | 0 | 0 | 0 | 0 | 0 | 0 | 0 | **0.02** | 0 | 0 | 0 | 0 | 0 | 0 | 0 |
| | FL321 | 0 | 0 | 0 | 0 | 0 | 0 | **0.13** | 0 | 0 | 0 | 0 | 0 | 0 | 0 | 0 | 0 | 0 | 0 |
| | FL348 | 0 | 0 | 0 | 0 | 0 | **0.10** | 0 | 0 | 0 | 0 | 0 | **0.13** | 0 | 0 | **0.08** | 0 | 0 | 0 |
| Queenstown | FL114 | 0 | 0 | 0 | 0 | 0 | 0 | 0 | 0 | 0 | 0 | 0 | **0.01** | 0 | 0 | 0 | 0 | 0 | **0.03** |
| | FL339 | 0 | 0 | 0 | 0 | 0 | 0 | 0 | 0 | 0 | 0 | 0 | 0 | 0 | 0 | 0 | 0 | 0 | **0.23** |
| | FL349 | 0 | 0 | 0 | 0 | 0 | 0 | 0 | 0 | 0 | 0 | **0.06** | **0.04** | 0 | 0 | 0 | 0 | 0 | 0 |
| Rochor | FL76 | 0 | 0 | 0 | 0 | 0 | 0 | 0 | 0 | 0 | 0 | 0 | 0 | 0 | 0 | 0 | **0.08** | 0 | 0 |
| Sengkang | FL497 | **0.26** | 0 | 0 | 0 | 0 | 0 | 0 | 0 | 0 | 0 | 0 | **0.04** | 0 | 0 | 0 | 0 | 0 | 0 |
| | FL535 | 0 | 0 | 0 | 0 | 0 | 0 | **0.03** | 0 | 0 | 0 | 0 | 0 | 0 | 0 | 0 | 0 | 0 | 0 |
| | FL733 | **0.20** | 0 | 0 | 0 | 0 | 0 | 0 | 0 | 0 | **0.09** | 0 | 0 | 0 | 0 | 0 | 0 | 0 | 0 |
| Serangoon | FL392 | 0 | 0 | 0 | 0 | 0 | 0 | 0 | 0 | 0 | 0 | 0 | 0 | 0 | 0 | 0 | **0.02** | 0 | 0 |
| | FL406 | 0 | 0 | **0.05** | 0 | 0 | 0 | 0 | 0 | 0 | 0 | 0 | 0 | 0 | 0 | 0 | 0 | 0 | 0 |
| Toa Payoh | FL164 | 0 | 0 | 0 | 0 | 0 | 0 | 0 | 0 | 0 | 0 | 0 | 0 | 0 | 0 | **0.15** | 0 | 0 | 0 |
| | FL58 | 0 | 0 | 0 | 0 | 0 | 0 | 0 | 0 | 0 | 0 | 0 | **0.01** | 0 | 0 | 0 | 0 | 0 | 0 |
| Toa Payoh | FL59 | **0.18** | 0 | 0 | 0 | 0 | **0.12** | 0 | 0 | **0.07** | **0.02** | 0 | 0 | 0 | 0 | **0.13** | 0 | 0 | 0 |
| | FL60 | 0 | 0 | 0 | 0 | 0 | 0 | 0 | 0 | 0 | 0 | 0 | 0 | 0 | 0 | 0 | 0 | 0 | **0.05** |
| Woodlands | FL253 | 0 | 0 | 0 | 0 | 0 | 0 | 0 | 0 | 0 | 0 | 0 | 0 | 0 | 0 | 0 | 0 | **0.06** | 0 |
| | FL575 | 0 | 0 | 0 | 0 | 0 | 0 | 0 | 0 | 0 | 0 | 0 | 0 | 0 | 0 | 0 | **0.01** | 0 | 0 |
| | FL598 | 0 | 0 | 0 | 0 | 0 | 0 | 0 | **0.05** | 0 | 0 | 0 | 0 | 0 | 0 | 0 | 0 | 0 | 0 |
| | FL599 | 0 | **0.02** | 0 | 0 | 0 | 0 | 0 | 0 | 0 | 0 | 0 | 0 | 0 | 0 | 0 | 0 | 0 | 0 |
| | FL600 | 0 | 0 | 0 | 0 | 0 | 0 | 0 | 0 | 0 | 0 | 0 | 0 | 0 | **0.01** | 0 | 0 | 0 | 0 |
| | FL607 | 0 | 0 | 0 | 0 | 0 | 0 | **0.25** | 0 | 0 | 0 | 0 | 0 | 0 | 0 | 0 | 0 | 0 | 0 |
| | FL619 | 0 | 0 | 0 | 0 | 0 | 0 | **0.04** | 0 | 0 | 0 | 0 | 0 | 0 | 0 | 0 | 0 | 0 | 0 |

Table 6: Donor weights for Choa Chu Kang sectors for models 3 and 4. Only sectors with any contribution to synthetic control weights for intervention sectors' synthetic controls are shown

| Donor | | Model 3 | | | | | | Model 4 | | | | | |
|---|---|---|---|---|---|---|---|---|---|---|---|---|---|
| Town | Sector | FL768 | FL803 | FL804 | FL806 | FL807 | FL826 | FL768 | FL803 | FL804 | FL806 | FL807 | FL826 |
| Ang Mo Kio | FL18 | 0 | 0 | 0 | **0.10** | 0 | 0 | 0 | 0 | 0 | 0 | 0 | 0 |
| | FL372 | 0 | 0 | 0 | 0 | 0 | 0 | 0 | **0.02** | 0 | 0 | 0 | 0 |
| | FL456 | 0 | 0 | 0 | 0 | 0 | 0 | 0 | 0 | **0.01** | **0.01** | 0 | 0 |
| | FL478 | 0 | 0 | 0 | 0 | 0 | 0 | 0 | **0.02** | 0 | 0 | 0 | 0 |
| | FL499 | 0 | 0 | 0 | **0.03** | **0.14** | 0 | 0 | 0 | 0 | 0 | 0 | 0 |
| Bedok | FL131 | 0 | 0 | 0 | 0 | 0 | 0 | 0 | 0 | 0 | **0.03** | 0 | 0 |
| | FL225 | 0 | 0 | 0 | 0 | 0 | 0 | 0 | 0 | 0 | **0.04** | **0.09** | 0 |
| | FL237 | 0 | 0 | 0 | 0 | 0 | 0 | 0 | **0.15** | **0.01** | 0 | **0.02** | **0.07** |
| Bishan | FL34 | **0.19** | **0.21** | **0.27** | 0 | 0 | 0 | 0 | 0 | **0.04** | 0 | 0 | 0 |
| | FL36 | 0 | 0 | 0 | 0 | 0 | 0 | 0 | 0 | **0.02** | 0 | 0 | 0 |
| | FL51 | 0 | 0 | 0 | 0 | 0 | 0 | 0 | **0.01** | 0 | 0 | 0 | 0 |
| Bukit Batok | FL434 | 0 | 0 | 0 | 0 | 0 | 0 | **0.05** | 0 | **0.10** | 0 | 0 | 0 |
| | FL435 | 0 | 0 | **0.19** | 0 | 0 | 0 | 0 | 0 | 0 | 0 | 0 | 0 |
| | FL680 | 0 | 0 | 0 | 0 | **0.03** | 0 | 0 | 0 | 0 | 0 | 0 | 0 |
| | FL686 | 0 | 0 | 0 | 0 | 0 | 0 | 0 | 0 | 0 | 0 | 0 | **0.11** |
| | FL699 | 0 | 0 | 0 | 0 | 0 | 0 | **0.03** | 0 | 0 | **0.07** | 0 | 0 |
| Bukit Merah | FL179 | 0 | 0 | 0 | 0 | 0 | 0 | 0 | **0.01** | 0 | 0 | 0 | 0 |
| | FL193 | 0 | 0 | 0 | 0 | 0 | 0 | 0 | 0 | 0 | 0 | **0.02** | 0 |
| | FL8 | 0 | 0 | 0 | 0 | 0 | 0 | 0 | 0 | 0 | 0 | 0 | **0.14** |
| Bukit Panjang | FL647 | 0 | 0 | 0 | 0 | **0.05** | 0 | 0 | 0 | 0 | 0 | 0 | 0 |
| | FL829 | 0 | 0 | 0 | 0 | 0 | 0 | 0 | 0 | 0 | **0.04** | 0 | 0 |
| Choa Chu Kang | FL729 | 0 | **0.03** | 0 | 0 | 0 | **0.09** | 0 | 0 | 0 | 0 | 0 | **0.01** |
| | FL764 | 0 | 0 | **0.02** | 0 | 0 | **0.20** | 0 | 0 | 0 | 0 | 0 | 0 |
| | FL811 | **0.07** | 0 | 0 | 0 | 0 | 0 | 0 | 0 | 0 | 0 | 0 | 0 |
| | FL818 | 0 | 0 | 0 | 0 | 0 | **0.03** | 0 | 0 | 0 | 0 | 0 | 0 |
| | FL823 | 0 | 0 | 0 | 0 | 0 | 0 | 0 | 0 | 0 | **0.05** | 0 | 0 |
| Clementi | FL670 | 0 | 0 | 0 | 0 | 0 | 0 | **0.02** | 0 | 0 | 0 | 0 | 0 |
| Geylang | FL19 | 0 | 0 | 0 | 0 | 0 | 0 | 0 | **0.04** | 0 | 0 | 0 | 0 |
| | FL200 | **0.02** | 0 | 0 | 0 | 0 | 0 | **0.01** | 0 | 0 | 0 | 0 | 0 |
| | FL204 | 0 | 0 | 0 | 0 | 0 | 0 | 0 | 0 | 0 | 0 | 0 | **0.04** |
| | FL81 | 0 | 0 | 0 | 0 | 0 | 0 | 0 | 0 | **0.01** | 0 | 0 | 0 |
| Hougang | FL367 | 0 | 0 | 0 | 0 | 0 | 0 | 0 | 0 | **0.15** | 0 | 0 | 0 |
| | FL410 | 0 | **0.10** | 0 | 0 | 0 | 0 | 0 | 0 | 0 | 0 | 0 | 0 |
| | FL451 | 0 | 0 | 0 | **0.29** | 0 | 0 | 0 | 0 | 0 | **0.08** | 0 | **0.15** |
| | FL452 | 0 | 0 | 0 | 0 | 0 | 0 | 0 | 0 | **0.10** | 0 | **0.03** | 0 |

Continued on next page

**Table 6 – continued from previous page**

| Donor | | Model 3 | | | | | | Model 4 | | | | | |
|---|---|---|---|---|---|---|---|---|---|---|---|---|---|
| Town | Sector | FL768 | FL803 | FL804 | FL806 | FL807 | FL826 | FL768 | FL803 | FL804 | FL806 | FL807 | FL826 |
| | FL464 | 0 | 0 | 0 | 0 | 0 | 0 | 0 | **0.05** | 0 | 0 | 0 | 0 |
| Jurong East | FL665 | **0.22** | **0.59** | 0 | 0 | 0 | 0 | 0 | **0.03** | 0 | 0 | 0 | 0 |
| | FL711 | 0 | 0 | 0 | 0 | 0 | 0 | 0 | **0.16** | **0.13** | 0 | 0 | 0 |
| | FL723 | 0 | 0 | 0 | 0 | 0 | 0 | 0 | **0.01** | 0 | 0 | 0 | 0 |
| | FL725 | 0 | 0 | 0 | 0 | 0 | 0 | 0 | **0.04** | 0 | 0 | 0 | 0 |
| | FL752 | 0 | 0 | 0 | 0 | 0 | 0 | 0 | 0 | 0 | 0 | **0.08** | 0 |
| Jurong West | FL431 | 0 | **0.02** | 0 | 0 | 0 | 0 | 0 | 0 | 0 | 0 | 0 | 0 |
| | FL441 | 0 | 0 | 0 | 0 | 0 | 0 | **0.02** | 0 | 0 | 0 | 0 | 0 |
| | FL442 | 0 | 0 | 0 | 0 | 0 | 0 | **0.07** | 0 | 0 | 0 | 0 | 0 |
| | FL652 | 0 | 0 | **0.12** | 0 | 0 | 0 | 0 | 0 | 0 | 0 | 0 | 0 |
| | FL662 | 0 | 0 | 0 | 0 | 0 | 0 | 0 | 0 | 0 | 0 | **0.01** | 0 |
| | FL709 | 0 | 0 | 0 | 0 | 0 | 0 | 0 | 0 | **0.02** | 0 | 0 | 0 |
| | FL710 | 0 | 0 | 0 | 0 | 0 | 0 | 0 | 0 | 0 | 0 | **0.19** | 0 |
| | FL769 | **0.32** | 0 | **0.20** | 0 | 0 | 0 | 0 | 0 | 0 | 0 | 0 | 0 |
| Kallang | FL14 | 0 | 0 | **0.15** | 0 | 0 | 0 | 0 | 0 | 0 | 0 | 0 | 0 |
| | FL185 | 0 | 0 | 0 | **0.06** | 0 | **0.07** | 0 | 0 | 0 | **0.06** | **0.01** | **0.10** |
| | FL26 | 0 | 0 | 0 | 0 | 0 | 0 | 0 | 0 | **0.10** | 0 | 0 | 0 |
| | FL85 | 0 | 0 | 0 | 0 | 0 | 0 | 0 | 0 | 0 | 0 | 0 | **0.04** |
| | FL93 | 0 | 0 | 0 | 0 | 0 | 0 | 0 | 0 | **0.06** | 0 | 0 | 0 |
| Marine Parade | FL310 | 0 | 0 | 0 | 0 | 0 | 0 | 0 | 0 | **0.08** | 0 | 0 | 0 |
| Novena | FL68 | 0 | 0 | 0 | 0 | 0 | 0 | 0 | 0 | 0 | 0 | **0.07** | 0 |
| | FL70 | 0 | 0 | 0 | 0 | 0 | 0 | 0 | 0 | 0 | 0 | 0 | **0.01** |
| Pasir Ris | FL166 | 0 | 0 | 0 | **0.40** | 0 | 0 | 0 | **0.11** | **0.09** | **0.17** | 0 | 0 |
| | FL228 | 0 | 0 | 0 | 0 | 0 | 0 | **0.08** | 0 | 0 | 0 | 0 | 0 |
| | FL256 | 0 | 0 | 0 | 0 | 0 | 0 | **0.12** | **0.04** | 0 | **0.02** | 0 | 0 |
| | FL257 | 0 | 0 | 0 | 0 | 0 | 0 | 0 | 0 | 0 | **0.02** | 0 | 0 |
| | FL272 | 0 | 0 | **0.04** | 0 | 0 | 0 | 0 | 0 | 0 | 0 | 0 | 0 |
| | FL273 | 0 | 0 | 0 | 0 | 0 | 0 | 0 | 0 | 0 | **0.08** | **0.02** | 0 |
| | FL293 | 0 | 0 | 0 | 0 | 0 | 0 | **0.08** | 0 | 0 | **0.07** | **0.12** | 0 |
| | FL321 | **0.03** | 0 | 0 | 0 | 0 | 0 | 0 | 0 | 0 | 0 | 0 | 0 |
| | FL336 | 0 | 0 | 0 | 0 | 0 | 0 | 0 | **0.02** | 0 | 0 | 0 | 0 |
| | FL348 | 0 | 0 | 0 | 0 | 0 | **0.13** | **0.01** | 0 | 0 | 0 | 0 | 0 |
| Queenstown | FL114 | 0 | 0 | 0 | 0 | 0 | 0 | **0.03** | 0 | 0 | 0 | 0 | 0 |
| | FL349 | 0 | 0 | 0 | **0.05** | 0 | 0 | 0 | 0 | 0 | 0 | 0 | 0 |
| Rochor | FL72 | 0 | 0 | 0 | 0 | 0 | 0 | 0 | 0 | **0.05** | 0 | 0 | 0 |
| | FL73 | 0 | 0 | 0 | 0 | 0 | 0 | **0.04** | **0.05** | 0 | 0 | 0 | 0 |
| Sembawang | FL527 | 0 | 0 | 0 | 0 | **0.14** | 0 | 0 | 0 | 0 | 0 | 0 | 0 |
| Sengkang | FL369 | 0 | 0 | 0 | 0 | 0 | 0 | 0 | 0 | 0 | **0.01** | 0 | 0 |
| | FL503 | 0 | 0 | 0 | 0 | 0 | 0 | **0.03** | 0 | 0 | 0 | 0 | 0 |
| | FL504 | 0 | 0 | 0 | 0 | 0 | 0 | **0.03** | 0 | 0 | 0 | 0 | 0 |
| | FL535 | **0.05** | 0 | 0 | 0 | 0 | 0 | 0 | 0 | 0 | 0 | 0 | 0 |
| | FL733 | 0 | 0 | 0 | 0 | **0.45** | 0 | 0 | 0 | 0 | 0 | **0.05** | 0 |
| | FL759 | 0 | 0 | 0 | 0 | 0 | 0 | **0.02** | 0 | 0 | 0 | 0 | 0 |
| Toa Payoh | FL164 | 0 | 0 | 0 | 0 | 0 | 0 | 0 | 0 | **0.02** | 0 | 0 | 0 |
| | FL319 | 0 | 0 | 0 | 0 | 0 | 0 | 0 | 0 | 0 | **0.05** | 0 | 0 |
| | FL359 | 0 | 0 | 0 | 0 | 0 | 0 | 0 | **0.19** | 0 | 0 | 0 | 0 |
| | FL42 | 0 | 0 | 0 | 0 | 0 | 0 | 0 | **0.05** | 0 | **0.01** | 0 | 0 |
| | FL58 | 0 | 0 | 0 | 0 | 0 | 0 | **0.13** | 0 | 0 | **0.17** | 0 | 0 |
| | FL59 | 0 | 0 | 0 | 0 | 0 | 0 | 0 | 0 | 0 | 0 | **0.13** | 0 |
| | FL61 | 0 | 0 | 0 | 0 | 0 | 0 | 0 | 0 | **0.01** | 0 | 0 | 0 |
| Woodlands | FL232 | 0 | 0 | 0 | 0 | 0 | 0 | 0 | 0 | 0 | 0 | 0 | **0.12** |
| | FL246 | 0 | 0 | 0 | 0 | 0 | 0 | **0.04** | 0 | 0 | 0 | 0 | 0 |
| | FL253 | 0 | 0 | 0 | 0 | 0 | **0.12** | 0 | 0 | 0 | 0 | 0 | 0 |
| | FL518 | 0 | 0 | 0 | 0 | 0 | 0 | **0.05** | 0 | 0 | 0 | 0 | 0 |
| | FL574 | 0 | 0 | 0 | 0 | 0 | **0.20** | 0 | 0 | 0 | 0 | 0 | **0.13** |
| | FL580 | 0 | 0 | 0 | 0 | 0 | 0 | 0 | 0 | 0 | **0.02** | 0 | **0.01** |
| | FL587 | 0 | 0 | 0 | **0.06** | 0 | 0 | 0 | 0 | 0 | 0 | 0 | 0 |
| | FL595 | 0 | 0 | 0 | 0 | 0 | 0 | **0.13** | 0 | 0 | 0 | 0 | 0 |
| | FL598 | 0 | 0 | 0 | 0 | 0 | **0.16** | 0 | 0 | 0 | 0 | 0 | **0.07** |
| | FL600 | **0.10** | **0.05** | 0 | 0 | 0 | 0 | 0 | 0 | 0 | 0 | 0 | 0 |
| | FL617 | 0 | 0 | 0 | **0.01** | 0 | 0 | 0 | 0 | 0 | 0 | 0 | 0 |
| | FL619 | 0 | 0 | 0 | 0 | **0.18** | 0 | 0 | 0 | 0 | 0 | **0.17** | 0 |

Table 7: Donor weights for Tampines sectors for model 2. Only sectors with any contribution to synthetic control weights for intervention sectors' synthetic controls are shown

| Donor | | Model 2 | | | | | | | | | | | | | | | | | | |
|---|---|---|---|---|---|---|---|---|---|---|---|---|---|---|---|---|---|---|---|---|
| Town | Sector | FL118 | FL119 | FL120 | FL121 | FL122 | FL165 | FL170 | FL171 | FL172 | FL304 | FL305 | FL314 | FL315 | FL316 | FL326 | FL327 | FL328 | FL332 | FL333 |
| Ang Mo Kio | FL18 | 0 | 0 | 0 | 0 | 0 | 0 | 0 | 0 | 0 | **0.02** | 0 | 0 | 0 | 0 | 0 | 0 | 0 | 0 | 0 |
| | FL477 | 0 | 0 | 0 | **0.23** | 0 | 0 | 0 | 0 | 0 | 0 | 0 | 0 | 0 | 0 | 0 | 0 | 0 | 0 | 0 |
| | FL488 | 0 | 0 | 0 | 0 | 0 | 0 | 0 | **0.32** | 0 | 0 | 0 | 0 | 0 | 0 | 0 | 0 | 0 | 0 | 0 |
| | FL499 | 0 | 0 | 0 | 0 | 0 | 0 | 0 | 0 | 0 | **0.09** | 0 | 0 | 0 | 0 | 0 | 0 | 0 | 0 | 0 |
| | FL500 | 0 | 0 | 0 | 0 | 0 | 0 | 0 | 0 | 0 | 0 | 0 | 0 | 0 | 0 | 0 | 0 | 0 | 0 | **0.23** |
| Bedok | FL216 | 0 | 0 | 0 | 0 | 0 | 0 | 0 | 0 | 0 | 0 | 0 | 0 | **0.19** | 0 | 0 | 0 | 0 | 0 | 0 |
| | FL225 | 0 | 0 | 0 | 0 | **0.10** | 0 | 0 | 0 | 0 | 0 | 0 | 0 | 0 | 0 | 0 | 0 | 0 | 0 | 0 |
| | FL231 | 0 | 0 | 0 | 0 | 0 | 0 | 0 | 0 | 0 | 0 | 0 | 0 | 0 | 0 | 0 | **0.17** | 0 | **0.34** | 0 |
| | FL237 | 0 | 0 | 0 | 0 | 0 | 0 | 0 | **0.11** | 0 | 0 | 0 | 0 | 0 | 0 | 0 | 0 | 0 | 0 | 0 |
| | FL238 | 0 | 0 | 0 | 0 | 0 | 0 | 0 | 0 | 0 | 0 | **0.17** | 0 | 0 | 0 | 0 | **0.15** | 0 | 0 | 0 |
| | FL239 | 0 | 0 | 0 | 0 | 0 | 0 | 0 | 0 | **0.15** | 0 | 0 | 0 | 0 | 0 | 0 | 0 | 0 | 0 | 0 |
| | FL240 | 0 | 0 | **0.27** | 0 | 0 | 0 | 0 | 0 | 0 | 0 | 0 | 0 | 0 | 0 | 0 | 0 | 0 | 0 | 0 |
| | FL242 | 0 | **0.28** | 0 | 0 | 0 | 0 | 0 | 0 | 0 | 0 | 0 | 0 | 0 | 0 | 0 | 0 | 0 | 0 | 0 |
| | FL259 | 0 | 0 | 0 | 0 | 0 | 0 | 0 | 0 | **0.11** | 0 | 0 | 0 | 0 | 0 | 0 | 0 | 0 | 0 | 0 |
| Bishan | FL34 | 0 | 0 | 0 | 0 | 0 | **0.29** | **0.44** | **0.19** | 0 | 0 | 0 | **0.58** | **0.21** | **0.60** | **0.38** | **0.20** | **0.31** | 0 | **0.04** |
| Bukit Batok | FL680 | **0.24** | 0 | 0 | 0 | 0 | 0 | 0 | 0 | 0 | 0 | 0 | 0 | 0 | 0 | 0 | 0 | 0 | 0 | 0 |
| | FL699 | 0 | 0 | 0 | 0 | 0 | **0.21** | **0.23** | 0 | 0 | 0 | 0 | **0.08** | 0 | 0 | 0 | 0 | 0 | 0 | 0 |
| Bukit Merah | FL152 | 0 | 0 | 0 | 0 | 0 | 0 | 0 | 0 | 0 | 0 | 0 | 0 | 0 | 0 | 0 | 0 | **0.11** | 0 | 0 |
| | FL193 | 0 | 0 | 0 | **0.19** | 0 | 0 | 0 | 0 | 0 | 0 | 0 | 0 | 0 | 0 | 0 | 0 | 0 | **0.08** | 0 |
| Choa Chu Kang | FL729 | 0 | 0 | 0 | 0 | **0.19** | **0.11** | **0.02** | 0 | 0 | 0 | 0 | 0 | **0.02** | 0 | 0 | **0.03** | 0 | 0 | 0 |
| | FL811 | 0 | 0 | 0 | 0 | 0 | 0 | 0 | 0 | 0 | 0 | 0 | 0 | 0 | 0 | 0 | **0.10** | 0 | 0 | 0 |
| Geylang | FL200 | 0 | **0.08** | 0 | 0 | 0 | 0 | 0 | 0 | **0.18** | 0 | 0 | 0 | 0 | 0 | 0 | **0.06** | 0 | **0.26** | 0 |
| | FL204 | 0 | 0 | 0 | 0 | **0.09** | 0 | 0 | 0 | 0 | 0 | 0 | 0 | 0 | 0 | 0 | 0 | 0 | 0 | 0 |
| Hougang | FL395 | 0 | 0 | 0 | 0 | 0 | 0 | 0 | 0 | 0 | **0.16** | 0 | 0 | 0 | 0 | 0 | 0 | 0 | 0 | 0 |
| | FL405 | **0.59** | 0 | 0 | **0.17** | 0 | 0 | 0 | 0 | 0 | 0 | **0.30** | 0 | 0 | 0 | 0 | 0 | 0 | 0 | 0 |
| | FL409 | 0 | 0 | 0 | 0 | 0 | 0 | 0 | 0 | 0 | 0 | 0 | 0 | 0 | 0 | 0 | 0 | 0 | 0 | **0.08** |
| | FL448 | 0 | 0 | 0 | 0 | 0 | 0 | 0 | 0 | 0 | 0 | 0 | 0 | 0 | 0 | 0 | **0.04** | 0 | 0 | 0 |
| | FL464 | 0 | 0 | **0.17** | 0 | 0 | 0 | 0 | 0 | 0 | 0 | 0 | 0 | 0 | 0 | 0 | 0 | 0 | 0 | 0 |
| Jurong West | FL437 | 0 | 0 | 0 | **0.10** | 0 | 0 | 0 | 0 | 0 | 0 | 0 | 0 | 0 | 0 | 0 | 0 | 0 | 0 | 0 |
| | FL441 | 0 | 0 | 0 | 0 | 0 | **0.13** | **0.12** | 0 | 0 | 0 | 0 | 0 | **0.06** | 0 | **0.12** | **0.02** | 0 | 0 | 0 |
| | FL442 | **0.16** | 0 | 0 | 0 | 0 | 0 | 0 | 0 | 0 | **0.19** | **0.06** | 0 | 0 | 0 | 0 | 0 | 0 | 0 | 0 |
| | FL621 | 0 | 0 | 0 | 0 | **0.10** | **0.26** | **0.19** | 0 | 0 | 0 | 0 | **0.35** | 0 | **0.37** | **0.40** | 0 | **0.52** | 0 | 0 |
| | FL709 | 0 | 0 | 0 | 0 | 0 | 0 | 0 | 0 | 0 | 0 | 0 | 0 | 0 | 0 | 0 | 0 | 0 | **0.17** | 0 |
| Kallang | FL13 | 0 | **0.01** | 0 | 0 | 0 | 0 | 0 | 0 | **0.16** | 0 | 0 | 0 | 0 | 0 | 0 | 0 | 0 | 0 | 0 |
| | FL185 | 0 | 0 | 0 | 0 | 0 | 0 | 0 | **0.15** | 0 | 0 | 0 | 0 | 0 | 0 | 0 | 0 | 0 | 0 | 0 |
| Marine Parade | FL310 | 0 | 0 | 0 | 0 | 0 | 0 | 0 | 0 | 0 | 0 | 0 | 0 | 0 | **0.02** | 0 | 0 | 0 | 0 | 0 |
| | FL311 | 0 | 0 | 0 | 0 | 0 | 0 | 0 | 0 | 0 | **0.38** | 0 | 0 | 0 | 0 | 0 | 0 | **0.04** | 0 | 0 |
| Novena | FL68 | 0 | 0 | 0 | 0 | 0 | 0 | 0 | 0 | 0 | 0 | 0 | 0 | 0 | 0 | 0 | 0 | 0 | 0 | **0.12** |
| Pasir Ris | FL166 | 0 | 0 | 0 | 0 | **0.13** | 0 | 0 | 0 | 0 | 0 | **0.06** | 0 | 0 | 0 | 0 | 0 | 0 | 0 | 0 |
| | FL223 | 0 | 0 | **0.06** | 0 | 0 | 0 | 0 | 0 | **0.16** | 0 | 0 | 0 | 0 | 0 | 0 | 0 | 0 | 0 | **0.06** |
| | FL227 | 0 | **0.27** | 0 | 0 | 0 | 0 | 0 | **0.22** | 0 | **0.13** | **0.38** | 0 | 0 | 0 | 0 | 0 | 0 | 0 | **0.17** |
| | FL228 | 0 | 0 | 0 | 0 | 0 | 0 | 0 | 0 | 0 | **0.04** | 0 | 0 | 0 | 0 | 0 | 0 | 0 | 0 | 0 |
| | FL235 | 0 | **0.37** | 0 | 0 | 0 | 0 | 0 | 0 | **0.20** | 0 | **0.03** | 0 | 0 | 0 | 0 | 0 | 0 | 0 | 0 |
| | FL256 | 0 | 0 | **0.40** | 0 | 0 | 0 | 0 | 0 | 0 | 0 | 0 | 0 | 0 | 0 | 0 | 0 | 0 | 0 | 0 |
| | FL257 | 0 | 0 | 0 | 0 | 0 | 0 | 0 | 0 | **0.05** | 0 | 0 | 0 | 0 | 0 | 0 | 0 | 0 | 0 | 0 |
| | FL348 | 0 | 0 | 0 | 0 | 0 | 0 | 0 | 0 | 0 | 0 | 0 | 0 | 0 | 0 | **0.10** | **0.23** | 0 | 0 | 0 |
| Sengkang | FL136 | 0 | 0 | 0 | 0 | **0.16** | 0 | 0 | 0 | 0 | 0 | 0 | 0 | 0 | 0 | 0 | 0 | **0.01** | 0 | 0 |
| | FL492 | 0 | 0 | 0 | 0 | 0 | 0 | 0 | 0 | 0 | 0 | 0 | 0 | **0.15** | 0 | 0 | 0 | 0 | 0 | 0 |
| | FL535 | 0 | 0 | 0 | **0.16** | 0 | 0 | 0 | 0 | 0 | 0 | 0 | 0 | 0 | 0 | 0 | 0 | 0 | 0 | **0.30** |
| | FL537 | 0 | 0 | 0 | 0 | 0 | 0 | 0 | 0 | 0 | 0 | 0 | 0 | **0.37** | 0 | 0 | 0 | 0 | 0 | 0 |
| Toa Payoh | FL319 | 0 | 0 | 0 | **0.15** | 0 | 0 | 0 | 0 | 0 | 0 | 0 | 0 | 0 | 0 | 0 | 0 | 0 | 0 | 0 |
| Woodlands | FL247 | 0 | 0 | **0.09** | 0 | 0 | 0 | 0 | 0 | 0 | 0 | 0 | 0 | 0 | 0 | 0 | 0 | 0 | 0 | 0 |
| | FL248 | 0 | 0 | 0 | 0 | **0.23** | 0 | 0 | 0 | 0 | 0 | 0 | 0 | 0 | 0 | 0 | 0 | 0 | 0 | 0 |
| | FL571 | **0.01** | 0 | 0 | 0 | 0 | 0 | 0 | 0 | 0 | 0 | 0 | 0 | 0 | 0 | 0 | 0 | 0 | 0 | 0 |
| | FL580 | 0 | 0 | 0 | 0 | 0 | 0 | 0 | 0 | 0 | 0 | 0 | 0 | 0 | 0 | 0 | 0 | 0 | **0.15** | 0 |

Table 8: Donor weights for Tampines sectors for model 2. Only sectors with any contribution to synthetic control weights for intervention sectors' synthetic controls are shown

| Donor | | Model 2 | | | | | | | | | | | | | | | | | | |
|---|---|---|---|---|---|---|---|---|---|---|---|---|---|---|---|---|---|---|---|---|
| Town | Sector | FL337 | FL340 | FL341 | FL342 | FL343 | FL344 | FL350 | FL353 | FL354 | FL355 | FL356 | FL357 | FL787 | FL837 | FL94 | FL95 | FL96 | FL97 | FL98 |
| Ang Mo Kio | FL454 | 0 | 0 | 0 | 0 | 0 | 0 | 0 | 0 | 0 | 0 | 0 | 0 | 0 | 0 | 0 | 0 | 0 | **0.09** | 0 |
| | FL472 | 0 | 0 | **0.07** | 0 | 0 | 0 | 0 | 0 | 0 | 0 | 0 | 0 | 0 | 0 | 0 | 0 | 0 | 0 | 0 |
| | FL500 | 0 | 0 | 0 | **0.06** | **0.32** | 0 | 0 | 0 | 0 | **0.30** | 0 | 0 | 0 | 0 | 0 | 0 | 0 | **0.05** | 0 |
| | FL505 | 0 | 0 | 0 | 0 | 0 | 0 | 0 | 0 | 0 | 0 | 0 | 0 | 0 | 0 | 0 | 0 | **0.17** | **0.07** | 0 |
| Bedok | FL211 | 0 | 0 | 0 | 0 | 0 | 0 | 0 | 0 | 0 | 0 | 0 | **0.06** | 0 | 0 | 0 | 0 | 0 | 0 | 0 |
| | FL225 | 0 | 0 | 0 | 0 | **0.18** | 0 | 0 | 0 | 0 | 0 | 0 | 0 | 0 | 0 | 0 | 0 | 0 | 0 | 0 |
| | FL231 | 0 | **0.14** | 0 | 0 | 0 | 0 | 0 | 0 | 0 | 0 | 0 | 0 | 0 | 0 | 0 | 0 | 0 | 0 | 0 |
| | FL238 | 0 | 0 | 0 | 0 | 0 | 0 | 0 | 0 | 0 | 0 | **0.14** | 0 | 0 | 0 | 0 | 0 | 0 | 0 | 0 |
| | FL259 | 0 | 0 | 0 | 0 | **0.04** | 0 | 0 | 0 | 0 | **0.10** | 0 | 0 | 0 | 0 | 0 | 0 | 0 | 0 | 0 |
| | FL275 | 0 | 0 | 0 | 0 | 0 | 0 | 0 | 0 | 0 | 0 | **0.18** | 0 | 0 | 0 | 0 | 0 | 0 | 0 | 0 |
| Bishan | FL34 | 0 | **0.14** | 0 | 0 | 0 | **0.60** | **0.47** | **0.20** | 0 | 0 | 0 | **0.18** | **0.52** | **0.48** | **0.43** | 0 | **0.36** | 0 | **0.30** |
| | FL51 | 0 | **0.04** | 0 | 0 | 0 | 0 | 0 | 0 | 0 | 0 | 0 | 0 | 0 | 0 | 0 | 0 | 0 | 0 | 0 |
| Bukit Merah | FL152 | 0 | 0 | 0 | 0 | 0 | 0 | 0 | 0 | 0 | 0 | 0 | 0 | 0 | 0 | **0.08** | 0 | 0 | 0 | 0 |
| | FL7 | 0 | 0 | 0 | 0 | **0.27** | 0 | 0 | 0 | 0 | 0 | 0 | 0 | 0 | 0 | 0 | 0 | 0 | 0 | 0 |
| Choa Chu Kang | FL729 | 0 | 0 | **0.07** | 0 | 0 | **0.02** | **0.33** | 0 | 0 | 0 | 0 | **0.32** | 0 | **0.04** | 0 | 0 | 0 | 0 | **0.05** |
| | FL811 | 0 | **0.07** | 0 | 0 | 0 | 0 | 0 | 0 | 0 | 0 | 0 | 0 | 0 | 0 | 0 | 0 | 0 | 0 | **0.01** |
| | FL818 | 0 | 0 | 0 | 0 | 0 | 0 | 0 | 0 | 0 | 0 | 0 | 0 | 0 | 0 | 0 | 0 | 0 | **0.09** | 0 |
| Clementi | FL640 | 0 | **0.02** | 0 | 0 | 0 | 0 | 0 | 0 | 0 | 0 | 0 | 0 | 0 | 0 | 0 | 0 | 0 | 0 | 0 |
| Geylang | FL200 | 0 | 0 | 0 | 0 | 0 | 0 | 0 | 0 | 0 | 0 | **0.08** | 0 | 0 | 0 | 0 | 0 | 0 | 0 | 0 |
| | FL204 | 0 | 0 | 0 | 0 | 0 | 0 | 0 | 0 | 0 | 0 | 0 | 0 | 0 | 0 | 0 | **0.09** | 0 | 0 | 0 |
| | FL81 | 0 | 0 | 0 | 0 | 0 | 0 | 0 | 0 | **0.22** | 0 | 0 | 0 | 0 | 0 | 0 | 0 | 0 | 0 | 0 |
| Hougang | FL367 | 0 | 0 | 0 | 0 | 0 | 0 | 0 | 0 | 0 | 0 | **0.02** | 0 | 0 | 0 | 0 | 0 | 0 | 0 | 0 |
| | FL375 | 0 | **0.01** | 0 | 0 | 0 | 0 | 0 | 0 | 0 | 0 | 0 | 0 | 0 | 0 | 0 | 0 | 0 | 0 | 0 |
| | FL410 | **0.23** | 0 | 0 | 0 | 0 | 0 | 0 | 0 | 0 | 0 | 0 | 0 | 0 | 0 | 0 | 0 | 0 | 0 | 0 |
| | FL449 | 0 | 0 | 0 | **0.22** | 0 | 0 | 0 | 0 | 0 | 0 | 0 | 0 | 0 | 0 | 0 | 0 | 0 | 0 | 0 |
| | FL452 | 0 | 0 | 0 | 0 | 0 | 0 | 0 | 0 | **0.04** | 0 | 0 | 0 | 0 | 0 | 0 | 0 | 0 | 0 | 0 |
| | FL489 | **0.08** | 0 | 0 | 0 | 0 | 0 | 0 | 0 | 0 | 0 | 0 | 0 | 0 | 0 | 0 | 0 | 0 | 0 | 0 |
| | FL495 | **0.18** | 0 | 0 | 0 | 0 | **0.17** | 0 | 0 | 0 | 0 | 0 | 0 | 0 | 0 | 0 | 0 | 0 | 0 | 0 |
| Jurong East | FL2 | 0 | 0 | 0 | 0 | 0 | **0.13** | 0 | 0 | 0 | 0 | 0 | 0 | 0 | 0 | 0 | 0 | 0 | 0 | 0 |
| | FL697 | 0 | 0 | 0 | 0 | 0 | 0 | 0 | 0 | 0 | **0.29** | 0 | 0 | 0 | 0 | 0 | 0 | 0 | **0.25** | 0 |
| Jurong West | FL431 | 0 | 0 | 0 | 0 | 0 | 0 | 0 | 0 | 0 | 0 | 0 | 0 | 0 | **0.07** | 0 | 0 | 0 | 0 | 0 |
| | FL441 | 0 | 0 | 0 | 0 | 0 | 0 | 0 | 0 | 0 | 0 | 0 | 0 | 0 | 0 | 0 | 0 | 0 | 0 | **0.20** |
| | FL442 | 0 | 0 | 0 | 0 | 0 | 0 | 0 | 0 | **0.05** | 0 | 0 | 0 | 0 | 0 | 0 | 0 | 0 | 0 | 0 |
| | FL621 | 0 | 0 | 0 | **0.07** | 0 | 0 | **0.21** | 0 | 0 | 0 | 0 | 0 | **0.48** | 0 | **0.47** | 0 | 0 | 0 | **0.21** |
| | FL694 | 0 | 0 | 0 | 0 | 0 | 0 | 0 | 0 | 0 | 0 | 0 | 0 | 0 | 0 | **0.01** | 0 | 0 | 0 | 0 |
| | FL777 | 0 | 0 | 0 | 0 | 0 | **0.03** | 0 | 0 | 0 | 0 | 0 | 0 | 0 | 0 | 0 | 0 | 0 | 0 | 0 |
| Kallang | FL13 | 0 | 0 | 0 | 0 | 0 | **0.04** | 0 | 0 | 0 | 0 | 0 | 0 | 0 | 0 | 0 | **0.01** | 0 | 0 | 0 |
| | FL185 | 0 | 0 | 0 | 0 | 0 | 0 | 0 | 0 | 0 | 0 | 0 | 0 | 0 | 0 | 0 | 0 | **0.13** | 0 | 0 |
| | FL26 | 0 | 0 | 0 | 0 | 0 | 0 | 0 | 0 | 0 | **0.12** | 0 | 0 | 0 | 0 | 0 | 0 | 0 | 0 | 0 |
| | FL93 | 0 | 0 | **0.21** | 0 | 0 | 0 | 0 | 0 | 0 | 0 | 0 | 0 | 0 | 0 | 0 | 0 | 0 | 0 | 0 |
| Marine Parade | FL311 | 0 | 0 | 0 | 0 | 0 | 0 | 0 | 0 | **0.26** | 0 | 0 | 0 | 0 | 0 | **0.02** | **0.21** | 0 | 0 | 0 |
| | FL318 | 0 | 0 | 0 | 0 | 0 | 0 | 0 | **0.03** | **0.02** | 0 | 0 | 0 | 0 | 0 | 0 | 0 | 0 | 0 | 0 |
| Pasir Ris | FL166 | 0 | 0 | 0 | 0 | 0 | 0 | 0 | 0 | 0 | **0.12** | 0 | 0 | 0 | 0 | 0 | 0 | 0 | 0 | 0 |
| | FL223 | 0 | 0 | 0 | 0 | **0.01** | 0 | 0 | 0 | 0 | 0 | **0.28** | 0 | 0 | 0 | 0 | 0 | 0 | 0 | 0 |
| | FL227 | 0 | 0 | 0 | 0 | 0 | 0 | 0 | 0 | **0.15** | 0 | 0 | 0 | 0 | 0 | 0 | 0 | 0 | 0 | 0 |
| | FL228 | 0 | 0 | **0.26** | **0.22** | **0.16** | 0 | 0 | 0 | 0 | 0 | 0 | 0 | 0 | 0 | 0 | 0 | 0 | 0 | 0 |
| | FL229 | 0 | 0 | 0 | 0 | 0 | 0 | 0 | 0 | **0.25** | **0.07** | **0.09** | 0 | 0 | 0 | 0 | 0 | 0 | **0.15** | 0 |
| | FL234 | 0 | 0 | 0 | 0 | 0 | 0 | 0 | 0 | 0 | 0 | 0 | 0 | 0 | 0 | 0 | **0.40** | 0 | 0 | 0 |
| | FL235 | **0.37** | 0 | 0 | 0 | 0 | 0 | 0 | 0 | 0 | 0 | 0 | 0 | 0 | 0 | 0 | **0.29** | 0 | 0 | 0 |
| | FL257 | 0 | 0 | 0 | **0.01** | 0 | 0 | 0 | 0 | 0 | 0 | 0 | 0 | 0 | 0 | 0 | 0 | 0 | 0 | 0 |
| | FL272 | 0 | 0 | 0 | 0 | **0.02** | 0 | 0 | **0.40** | 0 | 0 | 0 | 0 | 0 | 0 | 0 | 0 | 0 | 0 | 0 |
| | FL348 | 0 | 0 | 0 | 0 | 0 | 0 | 0 | 0 | 0 | 0 | 0 | 0 | 0 | 0 | 0 | 0 | 0 | 0 | **0.24** |
| | FL6 | **0.15** | 0 | 0 | 0 | 0 | 0 | 0 | 0 | 0 | 0 | **0.21** | 0 | 0 | 0 | 0 | 0 | 0 | 0 | 0 |
| Sengkang | FL132 | 0 | **0.27** | 0 | 0 | 0 | 0 | 0 | 0 | 0 | 0 | 0 | 0 | 0 | 0 | 0 | 0 | 0 | 0 | 0 |
| | FL136 | 0 | 0 | 0 | 0 | 0 | 0 | 0 | **0.36** | 0 | 0 | 0 | 0 | 0 | 0 | 0 | 0 | 0 | 0 | 0 |
| | FL492 | 0 | 0 | 0 | 0 | 0 | 0 | 0 | 0 | 0 | 0 | 0 | 0 | 0 | 0 | 0 | 0 | **0.04** | **0.08** | 0 |
| | FL497 | 0 | 0 | 0 | **0.16** | 0 | 0 | 0 | 0 | 0 | 0 | 0 | **0.27** | 0 | 0 | 0 | 0 | **0.21** | **0.22** | 0 |
| | FL535 | 0 | **0.30** | 0 | 0 | 0 | 0 | 0 | 0 | 0 | 0 | 0 | 0 | 0 | 0 | 0 | 0 | 0 | 0 | 0 |
| Toa Payoh | FL61 | 0 | 0 | 0 | 0 | 0 | 0 | 0 | 0 | 0 | 0 | 0 | **0.10** | 0 | 0 | 0 | 0 | **0.03** | 0 | 0 |
| Woodlands | FL226 | 0 | 0 | **0.29** | 0 | 0 | 0 | 0 | 0 | 0 | 0 | 0 | 0 | 0 | 0 | 0 | 0 | 0 | 0 | 0 |
| | FL252 | 0 | 0 | 0 | **0.11** | 0 | 0 | 0 | 0 | 0 | 0 | 0 | 0 | 0 | 0 | 0 | 0 | 0 | 0 | 0 |
| | FL508 | 0 | 0 | 0 | 0 | 0 | 0 | 0 | 0 | 0 | 0 | 0 | 0 | 0 | **0.26** | 0 | 0 | 0 | 0 | 0 |
| | FL513 | 0 | 0 | 0 | 0 | 0 | 0 | 0 | 0 | 0 | 0 | 0 | 0 | 0 | **0.15** | 0 | 0 | 0 | 0 | 0 |
| | FL571 | 0 | 0 | 0 | 0 | 0 | 0 | 0 | 0 | 0 | 0 | 0 | **0.07** | 0 | 0 | 0 | 0 | 0 | 0 | 0 |
| | FL581 | 0 | 0 | **0.09** | 0 | 0 | 0 | 0 | 0 | 0 | 0 | 0 | 0 | 0 | 0 | 0 | 0 | 0 | 0 | 0 |
| | FL608 | 0 | 0 | 0 | 0 | 0 | 0 | 0 | 0 | 0 | 0 | 0 | 0 | 0 | 0 | 0 | 0 | **0.07** | 0 | 0 |
| | FL611 | 0 | 0 | 0 | **0.14** | 0 | 0 | 0 | 0 | 0 | 0 | 0 | 0 | 0 | 0 | 0 | 0 | 0 | 0 | 0 |

Table 9: Donor weights for Tampines sectors for models 3 and 4. Only sectors with any contribution to synthetic control weights for intervention sectors' synthetic controls are shown

| Donor | | Model 3 | | | | | | | | | | | | | Model 4 | | | | | | | | | | | | |
|---|---|---|---|---|---|---|---|---|---|---|---|---|---|---|---|---|---|---|---|---|---|---|---|---|---|---|---|
| Town | Sector | FL118 | FL119 | FL120 | FL121 | FL122 | FL165 | FL170 | FL171 | FL172 | FL304 | FL305 | FL314 | FL315 | FL118 | FL119 | FL120 | FL121 | FL122 | FL165 | FL170 | FL171 | FL172 | FL304 | FL305 | FL314 | FL315 |
| Ang Mo Kio | FL18 | 0 | 0 | 0 | **0.40** | 0 | 0 | 0 | 0 | 0 | 0 | 0 | 0 | 0 | 0 | 0 | 0 | **0.15** | 0 | 0 | 0 | 0 | 0 | 0 | 0 | 0 | 0 |
| | FL362 | 0 | **0.01** | 0 | 0 | 0 | 0 | 0 | 0 | 0 | 0 | **0.10** | 0 | 0 | 0 | 0 | 0 | 0 | 0 | 0 | 0 | 0 | 0 | 0 | 0 | 0 | 0 |
| | FL456 | 0 | 0 | 0 | 0 | **0.11** | 0 | 0 | 0 | 0 | 0 | 0 | 0 | 0 | 0 | 0 | 0 | 0 | 0 | 0 | 0 | 0 | 0 | 0 | 0 | 0 | 0 |
| | FL471 | 0 | 0 | 0 | 0 | 0 | 0 | 0 | 0 | 0 | 0 | **0.06** | 0 | 0 | 0 | **0.02** | 0 | 0 | 0 | 0 | 0 | 0 | 0 | **0.07** | 0 | 0 | 0 |
| | FL472 | 0 | **0.21** | 0 | 0 | 0 | 0 | 0 | 0 | 0 | 0 | 0 | 0 | 0 | 0 | **0.12** | 0 | **0.03** | 0 | 0 | 0 | 0 | 0 | 0 | 0 | 0 | 0 |
| | FL477 | 0 | 0 | 0 | 0 | 0 | 0 | 0 | 0 | **0.12** | 0 | 0 | 0 | 0 | 0 | 0 | 0 | **0.04** | 0 | 0 | 0 | 0 | 0 | 0 | 0 | 0 | 0 |
| | FL488 | 0 | 0 | 0 | 0 | 0 | 0 | 0 | 0 | 0 | 0 | 0 | 0 | 0 | 0 | 0 | 0 | 0 | 0 | 0 | 0 | **0.25** | 0 | 0 | 0 | 0 | 0 |
| | FL500 | 0 | 0 | **0.05** | 0 | 0 | 0 | 0 | **0.13** | 0 | 0 | 0 | 0 | 0 | 0 | 0 | 0 | 0 | 0 | 0 | 0 | 0 | 0 | 0 | 0 | 0 | 0 |
| Bedok | FL216 | 0 | 0 | 0 | 0 | **0.01** | 0 | 0 | 0 | 0 | 0 | 0 | 0 | 0 | 0 | 0 | 0 | 0 | 0 | 0 | 0 | 0 | 0 | 0 | 0 | 0 | 0 |
| | FL237 | 0 | 0 | 0 | 0 | 0 | 0 | 0 | **0.15** | 0 | 0 | 0 | 0 | **0.08** | 0 | 0 | 0 | **0.03** | 0 | 0 | 0 | **0.03** | 0 | 0 | **0.07** | 0 | **0.07** |
| | FL239 | 0 | 0 | 0 | 0 | 0 | 0 | 0 | 0 | **0.03** | 0 | 0 | 0 | 0 | 0 | 0 | 0 | 0 | 0 | 0 | 0 | 0 | 0 | 0 | 0 | 0 | 0 |
| | FL240 | 0 | 0 | 0 | 0 | 0 | 0 | 0 | 0 | 0 | 0 | 0 | 0 | 0 | 0 | 0 | 0 | 0 | 0 | 0 | 0 | 0 | 0 | 0 | **0.02** | 0 | 0 |
| | FL242 | 0 | 0 | 0 | 0 | 0 | 0 | 0 | 0 | 0 | 0 | **0.29** | 0 | 0 | 0 | 0 | 0 | 0 | 0 | 0 | 0 | 0 | 0 | **0.01** | 0 | 0 | 0 |
| | FL275 | 0 | 0 | **0.33** | **0.17** | 0 | 0 | 0 | 0 | 0 | 0 | 0 | 0 | 0 | 0 | 0 | 0 | 0 | 0 | 0 | 0 | 0 | 0 | 0 | 0 | 0 | 0 |
| Bishan | FL34 | 0 | 0 | 0 | 0 | 0 | **0.21** | **0.38** | **0.31** | 0 | 0 | 0 | **0.58** | **0.15** | 0 | 0 | 0 | 0 | 0 | **0.21** | **0.38** | **0.24** | 0 | 0 | 0 | **0.58** | **0.18** |
| | FL36 | 0 | 0 | 0 | 0 | 0 | 0 | 0 | 0 | 0 | 0 | 0 | 0 | 0 | 0 | 0 | 0 | 0 | 0 | 0 | 0 | 0 | 0 | 0 | **0.07** | 0 | 0 |
| Bukit Batok | FL434 | 0 | 0 | 0 | 0 | 0 | **0.13** | 0 | 0 | 0 | 0 | 0 | 0 | 0 | 0 | 0 | 0 | 0 | 0 | **0.13** | 0 | 0 | 0 | 0 | 0 | 0 | 0 |
| | FL680 | **0.22** | 0 | 0 | 0 | 0 | 0 | 0 | 0 | 0 | 0 | 0 | 0 | 0 | **0.29** | 0 | 0 | 0 | 0 | 0 | 0 | 0 | 0 | 0 | **0.03** | 0 | 0 |
| | FL686 | 0 | 0 | 0 | 0 | 0 | 0 | 0 | 0 | 0 | 0 | 0 | 0 | 0 | 0 | 0 | 0 | 0 | 0 | 0 | 0 | 0 | 0 | 0 | 0 | 0 | **0.01** |
| | FL699 | 0 | 0 | 0 | 0 | 0 | **0.21** | **0.24** | 0 | 0 | 0 | 0 | **0.08** | 0 | 0 | 0 | 0 | 0 | 0 | **0.21** | **0.24** | 0 | 0 | 0 | 0 | **0.08** | **0.03** |
| | FL731 | 0 | 0 | 0 | 0 | 0 | **0.15** | **0.03** | 0 | 0 | 0 | 0 | 0 | 0 | 0 | 0 | 0 | 0 | 0 | **0.15** | **0.03** | **0.12** | 0 | 0 | 0 | 0 | 0 |
| Bukit Merah | FL161 | 0 | 0 | 0 | 0 | 0 | 0 | 0 | 0 | 0 | 0 | 0 | 0 | 0 | 0 | 0 | 0 | 0 | 0 | 0 | 0 | 0 | **0.12** | 0 | 0 | 0 | 0 |
| | FL193 | 0 | 0 | 0 | 0 | 0 | 0 | 0 | 0 | 0 | 0 | 0 | 0 | 0 | 0 | 0 | 0 | **0.02** | 0 | 0 | 0 | 0 | 0 | 0 | 0 | 0 | 0 |
| | FL194 | 0 | 0 | 0 | 0 | 0 | 0 | 0 | 0 | 0 | 0 | 0 | 0 | 0 | 0 | 0 | 0 | 0 | 0 | 0 | 0 | 0 | **0.07** | 0 | 0 | 0 | 0 |
| Choa Chu Kang | FL729 | 0 | 0 | 0 | 0 | 0 | 0 | 0 | 0 | 0 | 0 | 0 | 0 | **0.10** | 0 | 0 | 0 | **0.05** | **0.10** | 0 | 0 | 0 | **0.12** | 0 | 0 | 0 | **0.01** |
| | FL764 | 0 | 0 | 0 | 0 | 0 | 0 | 0 | 0 | 0 | 0 | 0 | 0 | 0 | 0 | 0 | 0 | 0 | 0 | 0 | 0 | 0 | 0 | 0 | 0 | 0 | **0.07** |
| | FL811 | 0 | 0 | 0 | 0 | 0 | 0 | 0 | 0 | 0 | 0 | 0 | 0 | 0 | 0 | 0 | 0 | 0 | 0 | 0 | 0 | 0 | **0.07** | 0 | 0 | 0 | 0 |
| | FL818 | 0 | 0 | 0 | 0 | 0 | 0 | 0 | 0 | 0 | 0 | 0 | 0 | 0 | 0 | 0 | 0 | 0 | **0.28** | 0 | 0 | 0 | 0 | 0 | 0 | 0 | 0 |
| | FL823 | 0 | 0 | 0 | 0 | 0 | 0 | 0 | 0 | 0 | 0 | 0 | 0 | 0 | 0 | 0 | **0.30** | **0.15** | 0 | 0 | 0 | 0 | 0 | 0 | 0 | 0 | 0 |
| Geylang | FL200 | 0 | 0 | 0 | 0 | 0 | 0 | 0 | 0 | **0.20** | 0 | 0 | 0 | 0 | 0 | 0 | 0 | 0 | 0 | 0 | 0 | 0 | 0 | 0 | 0 | 0 | 0 |
| | FL204 | 0 | 0 | 0 | 0 | 0 | 0 | 0 | 0 | 0 | 0 | 0 | 0 | 0 | 0 | 0 | 0 | 0 | **0.13** | 0 | 0 | **0.02** | **0.01** | 0 | 0 | 0 | **0.02** |
| Hougang | FL395 | 0 | 0 | 0 | 0 | 0 | 0 | 0 | 0 | 0 | **0.23** | 0 | 0 | 0 | 0 | 0 | 0 | 0 | 0 | 0 | 0 | 0 | 0 | 0 | 0 | 0 | 0 |
| | FL405 | **0.27** | 0 | 0 | 0 | 0 | 0 | 0 | 0 | 0 | 0 | **0.30** | 0 | 0 | **0.18** | 0 | 0 | 0 | 0 | 0 | 0 | 0 | 0 | 0 | **0.13** | 0 | 0 |
| | FL451 | 0 | 0 | 0 | **0.13** | 0 | 0 | 0 | 0 | **0.20** | 0 | 0 | 0 | **0.19** | 0 | 0 | 0 | 0 | 0 | 0 | 0 | 0 | 0 | 0 | 0 | 0 | **0.07** |
| | FL463 | 0 | 0 | 0 | 0 | 0 | 0 | 0 | 0 | 0 | 0 | 0 | 0 | 0 | 0 | 0 | 0 | 0 | 0 | 0 | 0 | 0 | 0 | 0 | **0.05** | 0 | 0 |
| | FL465 | 0 | 0 | 0 | 0 | 0 | 0 | 0 | 0 | 0 | 0 | 0 | 0 | 0 | 0 | 0 | 0 | 0 | 0 | 0 | 0 | 0 | 0 | 0 | **0.02** | 0 | 0 |
| | FL495 | 0 | 0 | 0 | 0 | 0 | 0 | 0 | 0 | **0.06** | 0 | 0 | 0 | 0 | 0 | **0.03** | 0 | 0 | 0 | 0 | 0 | 0 | **0.01** | 0 | 0 | 0 | 0 |
| Jurong East | FL697 | 0 | 0 | 0 | 0 | 0 | 0 | 0 | 0 | 0 | 0 | 0 | 0 | 0 | 0 | 0 | 0 | 0 | **0.19** | 0 | 0 | 0 | 0 | 0 | 0 | 0 | 0 |
| | FL698 | 0 | 0 | 0 | 0 | 0 | 0 | 0 | 0 | 0 | 0 | 0 | 0 | 0 | 0 | 0 | **0.19** | 0 | 0 | 0 | 0 | 0 | 0 | 0 | 0 | 0 | 0 |
| | FL711 | 0 | 0 | 0 | 0 | 0 | 0 | 0 | 0 | 0 | 0 | 0 | 0 | 0 | 0 | 0 | **0.03** | 0 | 0 | 0 | 0 | 0 | 0 | 0 | 0 | 0 | 0 |
| | FL752 | 0 | 0 | 0 | 0 | 0 | 0 | 0 | 0 | 0 | 0 | 0 | 0 | 0 | 0 | 0 | 0 | 0 | 0 | 0 | 0 | 0 | 0 | 0 | **0.04** | 0 | 0 |
| Jurong West | FL442 | 0 | 0 | 0 | 0 | 0 | 0 | 0 | 0 | 0 | **0.19** | 0 | 0 | 0 | **0.07** | **0.17** | **0.12** | **0.03** | 0 | 0 | 0 | 0 | 0 | **0.03** | **0.18** | 0 | 0 |
| | FL621 | 0 | 0 | 0 | 0 | 0 | **0.23** | **0.19** | **0.20** | 0 | 0 | 0 | **0.35** | 0 | 0 | 0 | 0 | 0 | **0.03** | **0.23** | **0.19** | 0 | 0 | 0 | 0 | **0.35** | **0.16** |
| | FL643 | 0 | 0 | 0 | 0 | 0 | 0 | 0 | 0 | 0 | 0 | **0.04** | 0 | 0 | 0 | 0 | 0 | 0 | 0 | 0 | 0 | 0 | 0 | 0 | 0 | 0 | 0 |
| | FL652 | 0 | 0 | 0 | 0 | 0 | 0 | 0 | 0 | 0 | 0 | 0 | 0 | 0 | 0 | 0 | 0 | 0 | 0 | 0 | 0 | 0 | **0.01** | 0 | 0 | 0 | 0 |
| | FL709 | 0 | 0 | 0 | 0 | 0 | 0 | 0 | 0 | 0 | 0 | 0 | 0 | 0 | 0 | 0 | 0 | 0 | 0 | 0 | 0 | 0 | 0 | **0.09** | 0 | 0 | 0 |
| Kallang | FL13 | 0 | 0 | 0 | 0 | 0 | 0 | 0 | 0 | **0.14** | 0 | 0 | 0 | 0 | 0 | 0 | 0 | 0 | 0 | 0 | 0 | 0 | 0 | 0 | 0 | 0 | 0 |
| | FL185 | 0 | 0 | 0 | 0 | 0 | 0 | 0 | 0 | 0 | 0 | 0 | 0 | 0 | 0 | 0 | 0 | 0 | **0.01** | 0 | 0 | **0.11** | 0 | 0 | 0 | 0 | 0 |
| | FL26 | 0 | 0 | 0 | 0 | **0.32** | 0 | 0 | 0 | 0 | 0 | 0 | 0 | 0 | 0 | 0 | 0 | 0 | **0.06** | 0 | 0 | 0 | 0 | 0 | 0 | 0 | 0 |
| Marine Parade | FL311 | 0 | 0 | 0 | 0 | 0 | 0 | 0 | 0 | 0 | **0.34** | 0 | 0 | 0 | 0 | 0 | 0 | 0 | 0 | 0 | 0 | 0 | 0 | **0.03** | 0 | 0 | 0 |
| Novena | FL68 | 0 | 0 | 0 | 0 | 0 | 0 | 0 | 0 | 0 | 0 | 0 | 0 | 0 | 0 | 0 | 0 | 0 | 0 | 0 | 0 | 0 | 0 | 0 | 0 | 0 | **0.05** |
| Pasir Ris | FL223 | 0 | 0 | 0 | 0 | 0 | 0 | 0 | 0 | **0.25** | 0 | 0 | 0 | 0 | 0 | 0 | 0 | 0 | 0 | 0 | 0 | 0 | 0 | 0 | 0 | 0 | 0 |
| | FL227 | **0.09** | **0.37** | 0 | 0 | 0 | 0 | 0 | 0 | 0 | 0 | 0 | 0 | 0 | **0.05** | **0.32** | **0.02** | 0 | 0 | 0 | 0 | 0 | 0 | **0.08** | **0.21** | 0 | 0 |
| | FL228 | 0 | 0 | **0.01** | 0 | 0 | 0 | 0 | 0 | 0 | 0 | 0 | 0 | 0 | 0 | 0 | 0 | 0 | 0 | 0 | 0 | 0 | 0 | 0 | 0 | 0 | 0 |
| | FL229 | 0 | 0 | 0 | 0 | 0 | 0 | 0 | 0 | 0 | 0 | **0.21** | 0 | 0 | 0 | 0 | 0 | 0 | 0 | 0 | 0 | 0 | 0 | 0 | 0 | 0 | **0.02** |
| | FL235 | **0.43** | **0.40** | 0 | **0.20** | 0 | 0 | 0 | 0 | 0 | 0 | 0 | 0 | 0 | **0.19** | **0.08** | 0 | 0 | 0 | 0 | 0 | 0 | **0.04** | **0.01** | 0 | 0 | 0 |
| | FL256 | 0 | 0 | **0.36** | 0 | 0 | 0 | 0 | 0 | 0 | 0 | 0 | 0 | 0 | 0 | 0 | **0.35** | 0 | 0 | 0 | 0 | 0 | 0 | **0.14** | 0 | 0 | 0 |
| | FL288 | 0 | 0 | 0 | **0.10** | 0 | 0 | 0 | 0 | 0 | 0 | 0 | 0 | 0 | 0 | 0 | 0 | **0.03** | 0 | 0 | 0 | 0 | 0 | 0 | 0 | 0 | 0 |
| | FL336 | 0 | 0 | 0 | 0 | 0 | 0 | 0 | 0 | 0 | 0 | 0 | 0 | 0 | 0 | 0 | 0 | 0 | 0 | 0 | 0 | 0 | 0 | **0.02** | 0 | 0 | 0 |
| Queenstown | FL114 | 0 | 0 | 0 | 0 | 0 | 0 | 0 | 0 | 0 | 0 | 0 | 0 | 0 | 0 | 0 | 0 | 0 | 0 | 0 | 0 | 0 | **0.11** | 0 | 0 | 0 | 0 |
| | FL352 | 0 | 0 | 0 | 0 | 0 | 0 | 0 | 0 | 0 | 0 | 0 | 0 | 0 | 0 | 0 | 0 | 0 | 0 | 0 | 0 | 0 | **0.09** | 0 | 0 | 0 | 0 |
| Rochor | FL76 | 0 | 0 | 0 | 0 | 0 | 0 | 0 | 0 | 0 | 0 | 0 | 0 | 0 | **0.05** | 0 | 0 | 0 | 0 | 0 | 0 | 0 | 0 | **0.01** | **0.06** | 0 | 0 |

Continued on next page

**Table 9 – continued from previous page**

| Donor | | Model 3 | | | | | | | | | | | | | Model 4 | | | | | | | | | | | | |
|---|---|---|---|---|---|---|---|---|---|---|---|---|---|---|---|---|---|---|---|---|---|---|---|---|---|---|---|
| Town | Sector | FL118 | FL119 | FL120 | FL121 | FL122 | FL165 | FL170 | FL171 | FL172 | FL304 | FL305 | FL314 | FL315 | FL118 | FL119 | FL120 | FL121 | FL122 | FL165 | FL170 | FL171 | FL172 | FL304 | FL305 | FL314 | FL315 |
| Sembawang | FL527 | 0 | 0 | 0 | 0 | 0 | 0 | 0 | 0 | 0 | 0 | 0 | 0 | 0 | 0 | 0 | 0 | 0 | 0 | 0 | 0 | 0 | **0.21** | 0 | 0 | 0 | 0 |
| | FL132 | 0 | 0 | 0 | 0 | 0 | 0 | 0 | 0 | 0 | 0 | 0 | 0 | 0 | 0 | 0 | 0 | 0 | **0.06** | 0 | 0 | 0 | 0 | 0 | 0 | 0 | 0 |
| | FL133 | 0 | 0 | 0 | 0 | 0 | 0 | 0 | 0 | 0 | 0 | 0 | 0 | 0 | 0 | 0 | 0 | 0 | **0.06** | 0 | 0 | 0 | 0 | 0 | 0 | 0 | 0 |
| | FL136 | 0 | 0 | 0 | 0 | 0 | 0 | 0 | 0 | 0 | 0 | 0 | 0 | 0 | 0 | 0 | 0 | 0 | 0 | 0 | 0 | 0 | 0 | 0 | 0 | 0 | **0.01** |
| Sengkang | FL390 | 0 | 0 | 0 | 0 | 0 | 0 | 0 | 0 | 0 | 0 | 0 | 0 | 0 | 0 | 0 | 0 | 0 | **0.04** | 0 | 0 | 0 | 0 | 0 | 0 | 0 | 0 |
| | FL497 | 0 | 0 | 0 | 0 | **0.07** | 0 | 0 | 0 | 0 | 0 | 0 | 0 | **0.11** | 0 | 0 | 0 | 0 | 0 | 0 | 0 | 0 | 0 | 0 | 0 | 0 | **0.02** |
| | FL535 | 0 | 0 | 0 | 0 | 0 | 0 | 0 | 0 | 0 | 0 | 0 | 0 | 0 | 0 | 0 | 0 | **0.05** | **0.02** | 0 | 0 | 0 | 0 | 0 | **0.01** | 0 | 0 |
| | FL537 | 0 | 0 | 0 | 0 | 0 | 0 | 0 | 0 | 0 | 0 | 0 | 0 | 0 | 0 | 0 | 0 | 0 | 0 | 0 | 0 | 0 | 0 | 0 | 0 | 0 | **0.01** |
| | FL392 | 0 | 0 | 0 | 0 | 0 | 0 | 0 | 0 | 0 | **0.02** | 0 | 0 | 0 | 0 | 0 | 0 | 0 | 0 | 0 | 0 | 0 | 0 | 0 | 0 | 0 | 0 |
| Serangoon | FL393 | 0 | 0 | 0 | 0 | 0 | 0 | 0 | 0 | 0 | 0 | 0 | 0 | 0 | 0 | 0 | 0 | **0.11** | 0 | 0 | 0 | 0 | 0 | 0 | 0 | 0 | 0 |
| | FL406 | 0 | 0 | 0 | 0 | **0.32** | 0 | 0 | **0.22** | 0 | **0.11** | 0 | 0 | 0 | 0 | 0 | 0 | 0 | 0 | 0 | 0 | **0.03** | 0 | 0 | 0 | 0 | 0 |
| | FL164 | 0 | 0 | 0 | 0 | 0 | **0.07** | **0.16** | 0 | 0 | 0 | 0 | 0 | **0.21** | 0 | 0 | 0 | 0 | 0 | **0.07** | **0.16** | 0 | 0 | 0 | 0 | 0 | **0.10** |
| | FL359 | 0 | 0 | 0 | 0 | 0 | 0 | 0 | 0 | 0 | 0 | 0 | 0 | 0 | 0 | 0 | 0 | **0.10** | 0 | 0 | 0 | 0 | 0 | 0 | 0 | 0 | 0 |
| | FL40 | 0 | 0 | 0 | 0 | **0.07** | 0 | 0 | 0 | 0 | 0 | 0 | 0 | 0 | 0 | 0 | 0 | 0 | 0 | 0 | 0 | 0 | 0 | 0 | 0 | 0 | 0 |
| Toa Payoh | FL42 | 0 | 0 | 0 | 0 | 0 | 0 | 0 | 0 | 0 | 0 | 0 | 0 | 0 | 0 | 0 | 0 | 0 | 0 | 0 | 0 | **0.06** | 0 | 0 | 0 | 0 | 0 |
| | FL44 | 0 | 0 | 0 | 0 | 0 | 0 | 0 | 0 | 0 | 0 | 0 | 0 | 0 | 0 | 0 | 0 | 0 | 0 | 0 | 0 | 0 | 0 | 0 | **0.03** | 0 | 0 |
| | FL59 | 0 | 0 | 0 | 0 | 0 | 0 | 0 | 0 | 0 | 0 | 0 | 0 | 0 | 0 | 0 | 0 | 0 | 0 | 0 | 0 | **0.01** | 0 | 0 | 0 | 0 | 0 |
| | FL60 | 0 | 0 | 0 | 0 | 0 | 0 | 0 | 0 | 0 | 0 | 0 | 0 | 0 | 0 | 0 | 0 | 0 | 0 | 0 | 0 | **0.04** | 0 | 0 | 0 | 0 | 0 |
| | FL61 | 0 | 0 | 0 | 0 | 0 | 0 | 0 | 0 | 0 | 0 | 0 | 0 | 0 | 0 | 0 | 0 | 0 | 0 | 0 | 0 | 0 | 0 | 0 | 0 | 0 | **0.07** |
| | FL226 | 0 | 0 | 0 | 0 | 0 | 0 | 0 | 0 | 0 | 0 | 0 | 0 | **0.16** | 0 | 0 | 0 | 0 | 0 | 0 | 0 | 0 | 0 | 0 | 0 | 0 | **0.01** |
| Woodlands | FL246 | 0 | 0 | 0 | 0 | 0 | 0 | 0 | 0 | 0 | 0 | 0 | 0 | 0 | 0 | 0 | 0 | 0 | 0 | 0 | 0 | 0 | 0 | **0.22** | 0 | 0 | 0 |
| | FL247 | 0 | 0 | **0.24** | 0 | 0 | 0 | 0 | 0 | 0 | 0 | 0 | 0 | 0 | 0 | **0.06** | 0 | 0 | 0 | 0 | 0 | 0 | 0 | 0 | 0 | 0 | 0 |
| | FL248 | 0 | 0 | 0 | 0 | 0 | 0 | 0 | 0 | 0 | 0 | 0 | 0 | 0 | 0 | 0 | 0 | 0 | **0.01** | 0 | 0 | 0 | 0 | 0 | 0 | 0 | 0 |
| | FL249 | 0 | 0 | 0 | 0 | 0 | 0 | 0 | 0 | 0 | 0 | 0 | 0 | 0 | 0 | 0 | 0 | 0 | 0 | 0 | 0 | 0 | **0.09** | 0 | 0 | 0 | 0 |
| | FL250 | 0 | 0 | 0 | 0 | 0 | 0 | 0 | 0 | 0 | 0 | 0 | 0 | 0 | 0 | 0 | 0 | 0 | 0 | 0 | 0 | 0 | 0 | 0 | **0.02** | 0 | 0 |
| | FL252 | 0 | 0 | 0 | 0 | **0.10** | 0 | 0 | 0 | 0 | 0 | 0 | 0 | 0 | 0 | 0 | 0 | 0 | 0 | 0 | 0 | 0 | 0 | 0 | 0 | 0 | 0 |
| | FL508 | 0 | 0 | 0 | 0 | 0 | 0 | 0 | 0 | 0 | 0 | 0 | 0 | 0 | 0 | 0 | 0 | **0.13** | 0 | 0 | 0 | 0 | 0 | 0 | 0 | 0 | 0 |
| | FL514 | 0 | 0 | 0 | 0 | 0 | 0 | 0 | 0 | 0 | 0 | 0 | 0 | 0 | 0 | 0 | 0 | 0 | 0 | 0 | 0 | 0 | 0 | **0.01** | 0 | 0 | 0 |
| | FL518 | 0 | 0 | 0 | 0 | 0 | 0 | 0 | 0 | 0 | 0 | 0 | 0 | 0 | 0 | 0 | 0 | 0 | 0 | 0 | 0 | 0 | 0 | **0.14** | 0 | 0 | 0 |
| | FL571 | 0 | 0 | 0 | 0 | 0 | 0 | 0 | 0 | 0 | **0.11** | 0 | 0 | 0 | 0 | 0 | 0 | 0 | 0 | 0 | 0 | 0 | 0 | 0 | 0 | 0 | 0 |
| Woodlands | FL574 | 0 | 0 | 0 | 0 | 0 | 0 | 0 | 0 | 0 | 0 | 0 | 0 | 0 | 0 | 0 | 0 | 0 | 0 | 0 | 0 | 0 | 0 | 0 | 0 | 0 | **0.05** |
| | FL579 | 0 | **0.01** | 0 | 0 | 0 | 0 | 0 | 0 | 0 | 0 | 0 | 0 | 0 | 0 | **0.06** | 0 | 0 | 0 | 0 | 0 | 0 | 0 | 0 | **0.05** | 0 | 0 |
| | FL580 | 0 | 0 | 0 | 0 | 0 | 0 | 0 | 0 | 0 | 0 | 0 | 0 | 0 | 0 | 0 | 0 | **0.09** | 0 | 0 | 0 | **0.09** | 0 | 0 | 0 | 0 | **0.04** |
| | FL592 | 0 | 0 | 0 | 0 | 0 | 0 | 0 | 0 | 0 | 0 | 0 | 0 | 0 | 0 | 0 | 0 | 0 | 0 | 0 | 0 | 0 | **0.06** | 0 | 0 | 0 | 0 |
| | FL599 | 0 | 0 | 0 | 0 | 0 | 0 | 0 | 0 | 0 | 0 | 0 | 0 | 0 | 0 | 0 | 0 | 0 | 0 | 0 | 0 | 0 | 0 | **0.07** | 0 | 0 | 0 |
| | FL600 | 0 | 0 | 0 | 0 | 0 | 0 | 0 | 0 | 0 | 0 | 0 | 0 | 0 | **0.16** | **0.13** | 0 | 0 | **0.01** | 0 | 0 | 0 | 0 | **0.01** | 0 | 0 | 0 |
| | FL606 | 0 | 0 | 0 | 0 | 0 | 0 | 0 | 0 | 0 | 0 | 0 | 0 | 0 | 0 | 0 | 0 | 0 | 0 | 0 | 0 | 0 | 0 | **0.05** | 0 | 0 | 0 |
| | FL608 | 0 | 0 | 0 | 0 | 0 | 0 | 0 | 0 | 0 | 0 | 0 | 0 | 0 | 0 | 0 | 0 | 0 | **0.01** | 0 | 0 | 0 | 0 | 0 | 0 | 0 | 0 |
| | FL611 | 0 | 0 | 0 | 0 | 0 | 0 | 0 | 0 | 0 | 0 | 0 | 0 | 0 | 0 | 0 | 0 | 0 | 0 | 0 | 0 | 0 | 0 | 0 | 0 | 0 | **0.01** |

Table 10: Donor weights for Tampines sectors for models 3 and 4. Only sectors with any contribution to synthetic control weights for intervention sectors' synthetic controls are shown

| Donor | | Model 3 | | | | | | | | | | | | | Model 4 | | | | | | | | | | | | |
|---|---|---|---|---|---|---|---|---|---|---|---|---|---|---|---|---|---|---|---|---|---|---|---|---|---|---|---|
| Town | Sector | FL316 | FL326 | FL327 | FL328 | FL332 | FL333 | FL337 | FL340 | FL341 | FL342 | FL343 | FL344 | FL350 | FL316 | FL326 | FL327 | FL328 | FL332 | FL333 | FL337 | FL340 | FL341 | FL342 | FL343 | FL344 | FL350 |
| Ang Mo Kio | FL372 | 0 | 0 | 0 | 0 | 0 | **0.27** | 0 | **0.18** | 0 | 0 | 0 | 0 | 0 | 0 | 0 | 0 | 0 | 0 | **0.27** | 0 | **0.18** | 0 | 0 | 0 | 0 | 0 |
| | FL471 | 0 | 0 | 0 | 0 | 0 | 0 | 0 | **0.20** | 0 | 0 | 0 | 0 | 0 | 0 | 0 | 0 | 0 | 0 | 0 | 0 | **0.20** | 0 | 0 | 0 | 0 | 0 |
| | FL216 | 0 | 0 | 0 | 0 | 0 | **0.01** | 0 | 0 | 0 | 0 | 0 | 0 | 0 | 0 | 0 | 0 | 0 | 0 | **0.01** | 0 | 0 | 0 | 0 | 0 | 0 | 0 |
| | FL237 | 0 | 0 | **0.02** | 0 | 0 | 0 | 0 | 0 | 0 | **0.17** | **0.17** | 0 | 0 | 0 | 0 | **0.02** | 0 | 0 | 0 | 0 | 0 | 0 | **0.17** | **0.17** | 0 | 0 |
| Bedok | FL238 | 0 | 0 | **0.15** | 0 | 0 | 0 | 0 | 0 | 0 | 0 | 0 | 0 | 0 | 0 | 0 | **0.15** | 0 | 0 | 0 | 0 | 0 | 0 | 0 | 0 | 0 | 0 |
| | FL275 | 0 | 0 | **0.14** | 0 | 0 | 0 | 0 | 0 | 0 | 0 | 0 | 0 | 0 | 0 | 0 | **0.14** | 0 | 0 | 0 | 0 | 0 | 0 | 0 | 0 | 0 | 0 |
| | FL303 | 0 | 0 | 0 | 0 | 0 | 0 | 0 | **0.09** | 0 | 0 | 0 | 0 | 0 | 0 | 0 | 0 | 0 | 0 | 0 | 0 | **0.09** | 0 | 0 | 0 | 0 | 0 |
| Bishan | FL34 | **0.51** | **0.24** | **0.12** | **0.11** | 0 | **0.03** | 0 | 0 | 0 | 0 | **0.09** | 0 | **0.47** | **0.51** | **0.24** | **0.12** | **0.11** | 0 | **0.03** | 0 | 0 | 0 | 0 | **0.09** | 0 | **0.47** |
| | FL47 | 0 | 0 | 0 | 0 | 0 | 0 | 0 | **0.01** | 0 | 0 | 0 | 0 | 0 | 0 | 0 | 0 | 0 | 0 | 0 | 0 | **0.01** | 0 | 0 | 0 | 0 | 0 |
| | FL434 | 0 | **0.15** | 0 | 0 | 0 | 0 | 0 | 0 | 0 | 0 | **0.13** | 0 | 0 | 0 | **0.15** | 0 | 0 | 0 | 0 | 0 | 0 | 0 | 0 | **0.13** | 0 | 0 |
| Bukit Batok | FL699 | 0 | 0 | 0 | **0.20** | 0 | **0.01** | 0 | 0 | 0 | 0 | 0 | 0 | 0 | 0 | 0 | 0 | **0.20** | 0 | **0.01** | 0 | 0 | 0 | 0 | 0 | 0 | 0 |
| | FL731 | 0 | **0.21** | 0 | 0 | 0 | 0 | 0 | 0 | 0 | 0 | 0 | 0 | 0 | 0 | **0.21** | 0 | 0 | 0 | 0 | 0 | 0 | 0 | 0 | 0 | 0 | 0 |
| Choa Chu Kang | FL729 | 0 | 0 | 0 | **0.07** | 0 | 0 | 0 | 0 | 0 | 0 | 0 | 0 | **0.33** | 0 | 0 | 0 | **0.07** | 0 | 0 | 0 | 0 | 0 | 0 | 0 | 0 | **0.33** |
| Geylang | FL200 | 0 | 0 | 0 | 0 | **0.18** | 0 | 0 | 0 | 0 | 0 | 0 | 0 | 0 | 0 | 0 | 0 | 0 | **0.18** | 0 | 0 | 0 | 0 | 0 | 0 | 0 | 0 |
| | FL409 | 0 | 0 | 0 | 0 | 0 | **0.20** | 0 | **0.07** | 0 | 0 | 0 | 0 | 0 | 0 | 0 | 0 | 0 | 0 | **0.20** | 0 | **0.07** | 0 | 0 | 0 | 0 | 0 |
| | FL410 | 0 | 0 | 0 | 0 | 0 | 0 | **0.32** | 0 | 0 | 0 | 0 | 0 | 0 | 0 | 0 | 0 | 0 | 0 | 0 | **0.32** | 0 | 0 | 0 | 0 | 0 | 0 |
| | FL415 | 0 | 0 | 0 | 0 | 0 | 0 | 0 | 0 | **0.22** | 0 | 0 | 0 | 0 | 0 | 0 | 0 | 0 | 0 | 0 | 0 | 0 | **0.22** | 0 | 0 | 0 | 0 |
| Hougang | FL449 | 0 | 0 | 0 | 0 | 0 | 0 | 0 | 0 | 0 | **0.23** | 0 | 0 | 0 | 0 | 0 | 0 | 0 | 0 | 0 | 0 | 0 | 0 | **0.23** | 0 | 0 | 0 |
| | FL452 | 0 | 0 | 0 | 0 | 0 | 0 | 0 | 0 | **0.10** | 0 | 0 | 0 | 0 | 0 | 0 | 0 | 0 | 0 | 0 | 0 | 0 | **0.10** | 0 | 0 | 0 | 0 |

Continued on next page

**Table 10 – continued from previous page**

| Donor | | Model 3 | | | | | | | | | | | | | Model 4 | | | | | | | | | | | | |
|---|---|---|---|---|---|---|---|---|---|---|---|---|---|---|---|---|---|---|---|---|---|---|---|---|---|---|---|
| Town | Sector | FL316 | FL326 | FL327 | FL328 | FL332 | FL333 | FL337 | FL340 | FL341 | FL342 | FL343 | FL344 | FL350 | FL316 | FL326 | FL327 | FL328 | FL332 | FL333 | FL337 | FL340 | FL341 | FL342 | FL343 | FL344 | FL350 |
| | FL490 | 0 | 0 | 0 | 0 | 0 | 0 | 0 | 0 | **0.42** | 0 | 0 | 0 | 0 | 0 | 0 | 0 | 0 | 0 | 0 | 0 | 0 | **0.42** | 0 | 0 | 0 | 0 |
| | FL495 | 0 | 0 | 0 | 0 | 0 | 0 | **0.17** | 0 | 0 | 0 | 0 | 0 | 0 | 0 | 0 | 0 | 0 | 0 | 0 | **0.17** | 0 | 0 | 0 | 0 | 0 | 0 |
| Jurong East | FL697 | 0 | 0 | 0 | 0 | **0.16** | **0.19** | 0 | 0 | 0 | 0 | 0 | 0 | 0 | 0 | 0 | 0 | 0 | **0.16** | **0.19** | 0 | 0 | 0 | 0 | 0 | 0 | 0 |
| Jurong West | FL441 | 0 | 0 | **0.09** | 0 | **0.06** | 0 | 0 | **0.16** | 0 | 0 | 0 | 0 | 0 | 0 | 0 | **0.09** | 0 | **0.06** | 0 | 0 | **0.16** | 0 | 0 | 0 | 0 | 0 |
| | FL621 | **0.38** | **0.25** | 0 | **0.25** | 0 | 0 | 0 | 0 | 0 | 0 | 0 | 0 | **0.21** | **0.38** | **0.25** | 0 | **0.25** | 0 | 0 | 0 | 0 | 0 | 0 | 0 | 0 | **0.21** |
| Kallang | FL181 | 0 | 0 | 0 | 0 | 0 | 0 | 0 | **0.13** | 0 | 0 | 0 | 0 | 0 | 0 | 0 | 0 | 0 | 0 | 0 | 0 | **0.13** | 0 | 0 | 0 | 0 | 0 |
| | FL185 | 0 | 0 | 0 | 0 | 0 | 0 | 0 | 0 | 0 | 0 | **0.19** | 0 | 0 | 0 | 0 | 0 | 0 | 0 | 0 | 0 | 0 | 0 | 0 | **0.19** | 0 | 0 |
| Marine Parade | FL310 | 0 | 0 | **0.04** | **0.02** | 0 | 0 | 0 | 0 | 0 | 0 | 0 | 0 | 0 | 0 | 0 | **0.04** | **0.02** | 0 | 0 | 0 | 0 | 0 | 0 | 0 | 0 | 0 |
| | FL311 | 0 | 0 | **0.12** | 0 | 0 | 0 | 0 | 0 | 0 | 0 | 0 | 0 | 0 | 0 | 0 | **0.12** | 0 | 0 | 0 | 0 | 0 | 0 | 0 | 0 | 0 | 0 |
| Pasir Ris | FL227 | 0 | 0 | 0 | 0 | 0 | **0.22** | 0 | 0 | 0 | 0 | 0 | 0 | 0 | 0 | 0 | 0 | 0 | 0 | **0.22** | 0 | 0 | 0 | 0 | 0 | 0 | 0 |
| | FL228 | 0 | 0 | 0 | 0 | 0 | 0 | 0 | 0 | 0 | **0.22** | **0.05** | 0 | 0 | 0 | 0 | 0 | 0 | 0 | 0 | 0 | 0 | 0 | **0.22** | **0.05** | 0 | 0 |
| | FL235 | 0 | 0 | 0 | 0 | 0 | 0 | **0.49** | 0 | 0 | 0 | 0 | 0 | 0 | 0 | 0 | 0 | 0 | 0 | 0 | **0.49** | 0 | 0 | 0 | 0 | 0 | 0 |
| | FL294 | 0 | 0 | 0 | 0 | **0.18** | 0 | 0 | 0 | **0.26** | 0 | 0 | 0 | 0 | 0 | 0 | 0 | 0 | **0.18** | 0 | 0 | 0 | **0.26** | 0 | 0 | 0 | 0 |
| | FL348 | 0 | **0.01** | 0 | 0 | 0 | 0 | 0 | 0 | 0 | 0 | 0 | 0 | 0 | 0 | **0.01** | 0 | 0 | 0 | 0 | 0 | 0 | 0 | 0 | 0 | 0 | 0 |
| | FL6 | 0 | 0 | 0 | 0 | 0 | 0 | **0.02** | 0 | 0 | 0 | 0 | 0 | 0 | 0 | 0 | 0 | 0 | 0 | 0 | **0.02** | 0 | 0 | 0 | 0 | 0 | 0 |
| Sengkang | FL132 | 0 | 0 | 0 | 0 | 0 | 0 | 0 | **0.15** | 0 | 0 | 0 | 0 | 0 | 0 | 0 | 0 | 0 | 0 | 0 | 0 | **0.15** | 0 | 0 | 0 | 0 | 0 |
| | FL497 | **0.03** | 0 | 0 | 0 | **0.32** | 0 | 0 | 0 | 0 | **0.10** | **0.30** | 0 | 0 | **0.03** | 0 | 0 | 0 | **0.32** | 0 | 0 | 0 | 0 | **0.10** | **0.30** | 0 | 0 |
| Serangoon | FL406 | 0 | 0 | 0 | 0 | 0 | **0.09** | 0 | 0 | 0 | 0 | 0 | 0 | 0 | 0 | 0 | 0 | 0 | 0 | **0.09** | 0 | 0 | 0 | 0 | 0 | 0 | 0 |
| Toa Payoh | FL164 | **0.08** | **0.14** | **0.32** | 0 | 0 | 0 | 0 | 0 | 0 | **0.08** | 0 | 0 | 0 | **0.08** | **0.14** | **0.32** | 0 | 0 | 0 | 0 | 0 | 0 | **0.08** | 0 | 0 | 0 |
| | FL58 | 0 | 0 | 0 | 0 | 0 | 0 | 0 | **0.02** | 0 | 0 | 0 | 0 | 0 | 0 | 0 | 0 | 0 | 0 | 0 | 0 | **0.02** | 0 | 0 | 0 | 0 | 0 |
| | FL59 | 0 | 0 | 0 | 0 | **0.10** | 0 | 0 | 0 | 0 | 0 | 0 | 0 | 0 | 0 | 0 | 0 | 0 | **0.10** | 0 | 0 | 0 | 0 | 0 | 0 | 0 | 0 |
| | FL61 | 0 | 0 | 0 | 0 | 0 | 0 | 0 | 0 | 0 | **0.12** | 0 | 0 | 0 | 0 | 0 | 0 | 0 | 0 | 0 | 0 | 0 | 0 | **0.12** | 0 | 0 | 0 |
| Woodlands | FL262 | 0 | 0 | 0 | **0.32** | 0 | 0 | 0 | 0 | 0 | 0 | **0.07** | 0 | 0 | 0 | 0 | 0 | **0.32** | 0 | 0 | 0 | 0 | 0 | 0 | **0.07** | 0 | 0 |
| | FL580 | 0 | 0 | 0 | 0 | 0 | 0 | 0 | 0 | 0 | **0.10** | 0 | 0 | 0 | 0 | 0 | 0 | 0 | 0 | 0 | 0 | 0 | 0 | **0.10** | 0 | 0 | 0 |
| | FL581 | 0 | 0 | 0 | 0 | 0 | 0 | 0 | 0 | 0 | 0 | 0 | 0 | 0 | 0 | 0 | 0 | 0 | 0 | 0 | 0 | 0 | 0 | 0 | 0 | 0 | 0 |
| | FL586 | 0 | 0 | 0 | **0.03** | 0 | 0 | 0 | 0 | 0 | 0 | 0 | 0 | 0 | 0 | 0 | 0 | **0.03** | 0 | 0 | 0 | 0 | 0 | 0 | 0 | 0 | 0 |

Table 11: Donor weights for Tampines sectors for models 3 and 4. Only sectors with any contribution to synthetic control weights for intervention sectors' synthetic controls are shown

| Donor | | Model 3 | | | | | | | | | | | | Model 4 | | | | | | | | | | | |
|---|---|---|---|---|---|---|---|---|---|---|---|---|---|---|---|---|---|---|---|---|---|---|---|---|---|
| Town | Sector | FL353 | FL354 | FL355 | FL356 | FL357 | FL787 | FL837 | FL94 | FL95 | FL96 | FL97 | FL98 | FL353 | FL354 | FL355 | FL356 | FL357 | FL787 | FL837 | FL94 | FL95 | FL96 | FL97 | FL98 |
| Ang Mo Kio | FL360 | 0 | 0 | 0 | 0 | 0 | 0 | 0 | 0 | 0 | 0 | 0 | 0 | 0 | 0 | 0 | 0 | 0 | 0 | 0 | 0 | 0 | 0 | **0.05** | 0 |
| | FL386 | 0 | **0.01** | 0 | 0 | 0 | 0 | 0 | 0 | 0 | 0 | 0 | 0 | 0 | 0 | 0 | 0 | 0 | 0 | 0 | 0 | 0 | 0 | 0 | 0 |
| | FL471 | 0 | 0 | 0 | 0 | 0 | 0 | 0 | 0 | 0 | 0 | 0 | 0 | 0 | 0 | 0 | **0.05** | 0 | 0 | 0 | 0 | 0 | 0 | 0 | 0 |
| | FL472 | 0 | 0 | 0 | 0 | 0 | 0 | 0 | 0 | 0 | 0 | 0 | 0 | 0 | 0 | 0 | **0.05** | 0 | 0 | 0 | 0 | 0 | 0 | 0 | 0 |
| | FL478 | 0 | 0 | 0 | 0 | 0 | 0 | 0 | 0 | 0 | 0 | 0 | 0 | 0 | 0 | 0 | 0 | 0 | 0 | **0.02** | 0 | 0 | 0 | 0 | 0 |
| | FL488 | 0 | 0 | 0 | 0 | 0 | 0 | 0 | 0 | 0 | 0 | 0 | 0 | 0 | 0 | **0.01** | **0.01** | 0 | 0 | 0 | 0 | 0 | 0 | 0 | 0 |
| | FL500 | **0.01** | 0 | 0 | 0 | **0.27** | 0 | 0 | 0 | 0 | 0 | 0 | 0 | 0 | 0 | **0.03** | 0 | 0 | 0 | 0 | 0 | 0 | **0.04** | 0 | 0 |
| Bedok | FL131 | 0 | 0 | 0 | 0 | 0 | 0 | 0 | 0 | 0 | 0 | 0 | 0 | 0 | **0.01** | **0.03** | 0 | 0 | 0 | 0 | 0 | 0 | 0 | 0 | 0 |
| | FL231 | 0 | 0 | 0 | 0 | 0 | 0 | 0 | **0.10** | 0 | 0 | 0 | 0 | 0 | 0 | 0 | 0 | 0 | 0 | 0 | 0 | 0 | 0 | 0 | 0 |
| | FL237 | 0 | 0 | **0.09** | 0 | **0.01** | 0 | **0.04** | 0 | 0 | 0 | 0 | 0 | 0 | 0 | **0.06** | 0 | 0 | 0 | 0 | 0 | 0 | 0 | 0 | 0 |
| | FL240 | 0 | 0 | 0 | 0 | 0 | 0 | 0 | 0 | 0 | 0 | 0 | 0 | 0 | **0.04** | 0 | 0 | 0 | 0 | 0 | 0 | 0 | 0 | 0 | 0 |
| | FL259 | 0 | **0.44** | 0 | 0 | 0 | 0 | 0 | 0 | 0 | 0 | 0 | 0 | 0 | **0.19** | 0 | 0 | 0 | 0 | 0 | 0 | 0 | 0 | 0 | 0 |
| | FL303 | 0 | 0 | 0 | 0 | 0 | 0 | 0 | 0 | 0 | 0 | 0 | 0 | **0.07** | 0 | 0 | 0 | 0 | 0 | 0 | 0 | 0 | 0 | 0 | 0 |
| Bishan | FL34 | 0 | 0 | 0 | 0 | **0.08** | **0.52** | **0.19** | **0.27** | 0 | **0.27** | 0 | **0.39** | 0 | 0 | 0 | 0 | **0.21** | **0.52** | **0.33** | **0.37** | 0 | **0.25** | 0 | **0.27** |
| | FL51 | 0 | 0 | 0 | 0 | 0 | 0 | 0 | 0 | 0 | 0 | 0 | 0 | 0 | 0 | 0 | 0 | 0 | 0 | 0 | 0 | 0 | **0.03** | 0 | 0 |
| Bukit Batok | FL434 | 0 | 0 | 0 | 0 | 0 | 0 | 0 | 0 | 0 | 0 | 0 | 0 | 0 | 0 | **0.04** | 0 | 0 | 0 | 0 | **0.02** | 0 | 0 | 0 | 0 |
| | FL699 | 0 | 0 | 0 | 0 | 0 | 0 | 0 | 0 | 0 | 0 | 0 | 0 | 0 | 0 | 0 | 0 | 0 | **0.02** | 0 | **0.06** | 0 | 0 | 0 | **0.05** |
| | FL731 | 0 | 0 | 0 | 0 | 0 | 0 | **0.22** | **0.17** | 0 | 0 | 0 | 0 | 0 | 0 | 0 | 0 | 0 | 0 | **0.25** | **0.01** | 0 | 0 | 0 | 0 |
| Bukit Merah | FL147 | 0 | 0 | 0 | 0 | **0.08** | 0 | 0 | 0 | 0 | 0 | 0 | 0 | 0 | 0 | **0.04** | 0 | **0.03** | 0 | 0 | 0 | 0 | 0 | 0 | 0 |
| | FL152 | 0 | 0 | 0 | 0 | 0 | 0 | 0 | 0 | 0 | 0 | 0 | 0 | 0 | 0 | 0 | 0 | 0 | 0 | 0 | **0.05** | 0 | 0 | 0 | 0 |
| | FL161 | 0 | 0 | 0 | 0 | 0 | 0 | 0 | 0 | 0 | 0 | 0 | 0 | 0 | 0 | 0 | 0 | 0 | 0 | **0.18** | 0 | 0 | 0 | 0 | 0 |
| | FL194 | 0 | 0 | 0 | 0 | 0 | 0 | 0 | 0 | 0 | 0 | 0 | 0 | 0 | 0 | 0 | **0.04** | 0 | 0 | 0 | 0 | **0.06** | 0 | 0 | 0 |
| Choa Chu Kang | FL729 | 0 | 0 | 0 | 0 | **0.29** | 0 | 0 | 0 | 0 | **0.06** | 0 | 0 | **0.25** | 0 | **0.08** | 0 | **0.28** | 0 | **0.01** | 0 | 0 | **0.02** | **0.06** | **0.10** |
| | FL811 | 0 | 0 | 0 | 0 | 0 | 0 | 0 | 0 | 0 | 0 | 0 | 0 | 0 | **0.04** | 0 | **0.13** | 0 | 0 | 0 | 0 | 0 | **0.01** | 0 | 0 |
| | FL818 | 0 | 0 | 0 | 0 | 0 | 0 | 0 | 0 | 0 | 0 | 0 | 0 | 0 | 0 | **0.01** | 0 | 0 | 0 | 0 | 0 | 0 | **0.06** | **0.07** | 0 |
| | FL823 | 0 | 0 | 0 | 0 | 0 | 0 | 0 | 0 | 0 | 0 | 0 | 0 | 0 | 0 | 0 | 0 | **0.04** | 0 | 0 | 0 | **0.10** | 0 | 0 | 0 |
| Clementi | FL671 | 0 | 0 | **0.01** | 0 | 0 | 0 | 0 | 0 | 0 | 0 | 0 | 0 | 0 | 0 | 0 | 0 | 0 | 0 | 0 | 0 | 0 | 0 | 0 | 0 |
| Geylang | FL200 | 0 | 0 | 0 | 0 | 0 | 0 | 0 | 0 | 0 | 0 | **0.09** | 0 | 0 | 0 | 0 | 0 | 0 | 0 | 0 | 0 | 0 | 0 | **0.05** | 0 |
| | FL204 | 0 | 0 | 0 | **0.10** | 0 | 0 | 0 | 0 | 0 | 0 | 0 | 0 | 0 | 0 | **0.03** | 0 | 0 | 0 | 0 | 0 | **0.14** | **0.07** | **0.06** | 0 |
| | FL83 | 0 | 0 | 0 | 0 | 0 | 0 | 0 | 0 | 0 | 0 | 0 | 0 | 0 | 0 | 0 | **0.03** | 0 | 0 | 0 | **0.01** | 0 | 0 | 0 | 0 |
| Hougang | FL451 | 0 | 0 | 0 | 0 | 0 | 0 | 0 | 0 | 0 | 0 | **0.21** | 0 | 0 | 0 | 0 | 0 | 0 | 0 | 0 | 0 | 0 | 0 | **0.17** | 0 |
| | FL452 | 0 | 0 | 0 | **0.06** | 0 | 0 | 0 | 0 | 0 | 0 | 0 | 0 | 0 | **0.03** | 0 | 0 | 0 | 0 | 0 | 0 | 0 | 0 | 0 | 0 |

Continued on next page

**Table 11 – continued from previous page**

| Donor | | Model 3 | | | | | | | | | | | | Model 4 | | | | | | | | | | | |
|---|---|---|---|---|---|---|---|---|---|---|---|---|---|---|---|---|---|---|---|---|---|---|---|---|---|
| Town | Sector | FL353 | FL354 | FL355 | FL356 | FL357 | FL787 | FL837 | FL94 | FL95 | FL96 | FL97 | FL98 | FL353 | FL354 | FL355 | FL356 | FL357 | FL787 | FL837 | FL94 | FL95 | FL96 | FL97 | FL98 |
| | FL495 | 0 | 0 | 0 | **0.14** | 0 | 0 | 0 | 0 | 0 | 0 | 0 | 0 | 0 | 0 | 0 | **0.01** | 0 | 0 | 0 | 0 | 0 | 0 | 0 | 0 |
| | FL696 | 0 | 0 | 0 | 0 | 0 | 0 | 0 | 0 | 0 | 0 | 0 | 0 | 0 | 0 | 0 | 0 | 0 | 0 | **0.05** | 0 | 0 | 0 | 0 | 0 |
| Jurong East | FL697 | 0 | 0 | **0.07** | 0 | 0 | 0 | 0 | 0 | 0 | **0.06** | **0.19** | 0 | 0 | 0 | **0.16** | 0 | 0 | 0 | 0 | 0 | 0 | **0.19** | **0.22** | 0 |
| | FL698 | 0 | 0 | 0 | 0 | 0 | 0 | 0 | 0 | 0 | 0 | 0 | 0 | 0 | 0 | 0 | 0 | 0 | 0 | 0 | 0 | **0.01** | 0 | 0 | 0 |
| | FL724 | 0 | 0 | 0 | 0 | 0 | 0 | 0 | 0 | 0 | 0 | 0 | 0 | 0 | 0 | 0 | 0 | 0 | 0 | 0 | 0 | **0.05** | 0 | 0 | 0 |
| | FL752 | 0 | 0 | 0 | 0 | 0 | 0 | 0 | 0 | 0 | 0 | 0 | 0 | 0 | **0.02** | 0 | 0 | 0 | 0 | 0 | 0 | 0 | 0 | 0 | 0 |
| Jurong West | FL431 | 0 | 0 | 0 | 0 | 0 | 0 | 0 | 0 | 0 | 0 | 0 | 0 | 0 | 0 | **0.01** | 0 | 0 | 0 | 0 | 0 | 0 | 0 | 0 | 0 |
| | FL441 | 0 | 0 | 0 | 0 | 0 | 0 | 0 | 0 | 0 | 0 | 0 | 0 | 0 | 0 | 0 | 0 | 0 | 0 | 0 | 0 | 0 | 0 | 0 | **0.01** |
| | FL621 | 0 | 0 | 0 | 0 | 0 | **0.48** | 0 | **0.22** | 0 | 0 | 0 | **0.36** | 0 | 0 | 0 | 0 | **0.04** | **0.46** | 0 | **0.20** | 0 | 0 | 0 | **0.14** |
| Kallang | FL16 | 0 | 0 | 0 | 0 | 0 | 0 | 0 | 0 | 0 | 0 | 0 | 0 | 0 | 0 | 0 | 0 | 0 | 0 | 0 | 0 | **0.02** | 0 | 0 | 0 |
| | FL185 | 0 | 0 | 0 | 0 | 0 | 0 | 0 | 0 | 0 | **0.07** | 0 | 0 | 0 | 0 | **0.01** | 0 | 0 | 0 | **0.01** | 0 | 0 | **0.05** | 0 | 0 |
| | FL26 | 0 | 0 | **0.24** | 0 | 0 | 0 | 0 | 0 | 0 | 0 | **0.15** | 0 | 0 | 0 | **0.09** | 0 | 0 | 0 | 0 | 0 | 0 | **0.09** | **0.06** | 0 |
| | FL85 | 0 | 0 | 0 | 0 | 0 | 0 | 0 | 0 | 0 | 0 | 0 | 0 | 0 | 0 | **0.05** | 0 | 0 | 0 | 0 | 0 | 0 | 0 | 0 | 0 |
| Marine Parade | FL310 | **0.02** | 0 | **0.02** | 0 | 0 | 0 | 0 | 0 | 0 | 0 | 0 | 0 | 0 | 0 | 0 | 0 | 0 | 0 | 0 | 0 | 0 | 0 | 0 | 0 |
| | FL311 | 0 | **0.30** | 0 | **0.41** | 0 | 0 | **0.10** | 0 | 0 | 0 | 0 | 0 | 0 | **0.02** | 0 | 0 | 0 | 0 | 0 | 0 | 0 | 0 | 0 | 0 |
| Novena | FL68 | 0 | 0 | 0 | 0 | 0 | 0 | 0 | 0 | 0 | 0 | 0 | 0 | 0 | 0 | **0.03** | 0 | 0 | 0 | 0 | 0 | 0 | **0.02** | 0 | 0 |
| Pasir Ris | FL166 | 0 | 0 | 0 | 0 | **0.10** | 0 | 0 | 0 | 0 | 0 | 0 | 0 | 0 | 0 | **0.02** | 0 | **0.09** | 0 | 0 | 0 | 0 | 0 | 0 | 0 |
| | FL223 | 0 | **0.15** | 0 | 0 | 0 | 0 | 0 | 0 | 0 | 0 | 0 | 0 | 0 | 0 | 0 | 0 | 0 | 0 | 0 | 0 | 0 | 0 | 0 | 0 |
| | FL227 | 0 | **0.09** | 0 | 0 | 0 | 0 | 0 | 0 | 0 | 0 | 0 | 0 | 0 | **0.06** | 0 | 0 | 0 | 0 | 0 | 0 | 0 | 0 | 0 | 0 |
| | FL235 | 0 | 0 | 0 | 0 | 0 | 0 | 0 | 0 | 0 | 0 | 0 | 0 | 0 | 0 | 0 | 0 | 0 | 0 | 0 | 0 | **0.10** | 0 | 0 | 0 |
| | FL255 | 0 | 0 | 0 | **0.13** | 0 | 0 | 0 | 0 | 0 | 0 | 0 | 0 | 0 | 0 | 0 | 0 | 0 | 0 | 0 | 0 | 0 | 0 | 0 | 0 |
| | FL256 | 0 | 0 | 0 | 0 | 0 | 0 | 0 | 0 | **0.19** | 0 | 0 | 0 | 0 | **0.07** | 0 | 0 | 0 | 0 | 0 | 0 | 0 | 0 | 0 | 0 |
| | FL264 | 0 | 0 | 0 | **0.17** | 0 | 0 | 0 | 0 | 0 | 0 | 0 | **0.25** | 0 | 0 | 0 | 0 | 0 | 0 | 0 | 0 | 0 | 0 | 0 | **0.15** |
| | FL272 | 0 | 0 | 0 | 0 | 0 | 0 | 0 | 0 | 0 | 0 | 0 | 0 | **0.28** | 0 | 0 | 0 | 0 | 0 | 0 | 0 | 0 | 0 | 0 | 0 |
| | FL293 | 0 | 0 | 0 | 0 | 0 | 0 | **0.08** | 0 | 0 | 0 | 0 | 0 | 0 | 0 | 0 | 0 | 0 | 0 | 0 | 0 | 0 | 0 | 0 | 0 |
| | FL336 | 0 | 0 | 0 | 0 | 0 | 0 | 0 | 0 | 0 | 0 | 0 | 0 | 0 | 0 | 0 | 0 | 0 | 0 | 0 | 0 | **0.05** | 0 | 0 | 0 |
| | FL348 | 0 | 0 | 0 | 0 | 0 | 0 | **0.06** | 0 | 0 | 0 | 0 | 0 | 0 | 0 | **0.09** | 0 | **0.03** | 0 | 0 | **0.03** | 0 | 0 | 0 | **0.10** |
| Queenstown | FL114 | 0 | 0 | 0 | 0 | 0 | 0 | 0 | 0 | 0 | 0 | 0 | 0 | 0 | 0 | 0 | **0.13** | 0 | 0 | 0 | 0 | 0 | 0 | 0 | 0 |
| | FL352 | 0 | 0 | 0 | 0 | 0 | 0 | 0 | 0 | 0 | 0 | 0 | 0 | 0 | 0 | 0 | **0.10** | 0 | 0 | 0 | 0 | 0 | 0 | 0 | 0 |
| Rochor | FL73 | 0 | 0 | 0 | 0 | 0 | 0 | 0 | 0 | 0 | 0 | 0 | 0 | 0 | **0.02** | 0 | 0 | 0 | 0 | 0 | 0 | 0 | 0 | 0 | 0 |
| | FL76 | 0 | 0 | 0 | 0 | 0 | 0 | 0 | 0 | 0 | 0 | 0 | 0 | 0 | **0.04** | 0 | **0.07** | 0 | 0 | 0 | 0 | 0 | 0 | 0 | 0 |
| Sembawang | FL527 | 0 | 0 | 0 | 0 | 0 | 0 | 0 | 0 | 0 | 0 | 0 | 0 | 0 | 0 | 0 | **0.03** | 0 | 0 | 0 | 0 | 0 | 0 | 0 | 0 |
| Sengkang | FL133 | **0.10** | 0 | 0 | 0 | 0 | 0 | 0 | 0 | 0 | 0 | 0 | 0 | **0.28** | 0 | 0 | **0.02** | **0.01** | 0 | 0 | 0 | 0 | 0 | 0 | 0 |
| | FL136 | **0.56** | 0 | 0 | 0 | 0 | 0 | 0 | 0 | 0 | 0 | 0 | 0 | **0.11** | 0 | 0 | 0 | **0.09** | 0 | 0 | 0 | 0 | 0 | 0 | 0 |
| | FL497 | 0 | 0 | 0 | 0 | 0 | 0 | 0 | 0 | 0 | **0.20** | **0.13** | 0 | 0 | 0 | 0 | 0 | 0 | 0 | 0 | 0 | 0 | **0.01** | **0.03** | 0 |
| | FL538 | 0 | 0 | 0 | 0 | 0 | 0 | 0 | 0 | **0.16** | 0 | 0 | 0 | 0 | 0 | 0 | 0 | 0 | 0 | 0 | 0 | 0 | 0 | 0 | 0 |
| | FL733 | 0 | 0 | 0 | 0 | 0 | 0 | 0 | 0 | 0 | 0 | 0 | 0 | 0 | 0 | 0 | **0.05** | 0 | 0 | 0 | 0 | 0 | 0 | 0 | 0 |
| Serangoon | FL392 | 0 | 0 | 0 | 0 | 0 | 0 | 0 | 0 | **0.33** | 0 | 0 | 0 | 0 | 0 | 0 | 0 | 0 | 0 | 0 | 0 | **0.05** | 0 | 0 | 0 |
| | FL393 | 0 | 0 | 0 | 0 | 0 | 0 | 0 | 0 | 0 | 0 | 0 | 0 | 0 | 0 | 0 | 0 | 0 | 0 | 0 | 0 | **0.09** | 0 | 0 | 0 |
| | FL406 | **0.30** | 0 | 0 | 0 | 0 | 0 | 0 | 0 | 0 | 0 | 0 | 0 | 0 | 0 | 0 | 0 | 0 | 0 | 0 | 0 | 0 | 0 | 0 | 0 |
| Toa Payoh | FL164 | 0 | 0 | **0.36** | 0 | 0 | 0 | 0 | **0.24** | 0 | **0.34** | **0.23** | 0 | 0 | 0 | **0.11** | 0 | 0 | 0 | 0 | **0.24** | 0 | **0.12** | **0.16** | **0.18** |
| | FL319 | 0 | 0 | 0 | 0 | 0 | 0 | 0 | 0 | 0 | 0 | 0 | 0 | 0 | 0 | **0.05** | 0 | 0 | 0 | 0 | 0 | 0 | 0 | 0 | 0 |
| | FL40 | 0 | 0 | 0 | 0 | 0 | 0 | 0 | 0 | **0.02** | 0 | 0 | 0 | 0 | 0 | 0 | 0 | 0 | 0 | 0 | 0 | 0 | 0 | 0 | 0 |
| | FL42 | 0 | 0 | **0.18** | 0 | 0 | 0 | 0 | 0 | 0 | 0 | 0 | 0 | 0 | 0 | **0.04** | 0 | 0 | 0 | 0 | 0 | 0 | 0 | 0 | 0 |
| | FL58 | 0 | 0 | 0 | 0 | **0.07** | 0 | 0 | 0 | 0 | 0 | 0 | 0 | 0 | 0 | 0 | 0 | 0 | 0 | 0 | 0 | 0 | **0.02** | 0 | 0 |
| | FL59 | 0 | 0 | **0.03** | 0 | 0 | 0 | 0 | 0 | 0 | 0 | 0 | 0 | 0 | 0 | 0 | 0 | 0 | 0 | 0 | 0 | 0 | 0 | 0 | 0 |
| | FL61 | 0 | 0 | 0 | 0 | 0 | 0 | 0 | 0 | 0 | 0 | 0 | 0 | 0 | **0.06** | **0.02** | 0 | **0.10** | 0 | 0 | 0 | 0 | **0.02** | **0.03** | 0 |
| Woodlands | FL232 | 0 | 0 | 0 | 0 | 0 | 0 | **0.31** | 0 | 0 | 0 | 0 | 0 | 0 | 0 | 0 | 0 | 0 | 0 | **0.14** | 0 | 0 | 0 | 0 | 0 |
| | FL246 | 0 | 0 | 0 | 0 | 0 | 0 | 0 | 0 | 0 | 0 | 0 | 0 | 0 | 0 | 0 | **0.13** | 0 | 0 | 0 | 0 | 0 | 0 | 0 | 0 |
| | FL248 | **0.01** | 0 | 0 | 0 | 0 | 0 | 0 | 0 | 0 | 0 | 0 | 0 | 0 | 0 | 0 | **0.14** | **0.07** | 0 | 0 | 0 | 0 | 0 | **0.02** | 0 |
| | FL250 | 0 | 0 | 0 | 0 | 0 | 0 | 0 | 0 | 0 | 0 | 0 | 0 | 0 | **0.37** | 0 | 0 | 0 | 0 | 0 | 0 | 0 | 0 | 0 | 0 |
| | FL253 | 0 | 0 | 0 | 0 | 0 | 0 | 0 | 0 | **0.26** | 0 | 0 | 0 | 0 | 0 | 0 | 0 | 0 | 0 | 0 | 0 | 0 | 0 | 0 | 0 |
| | FL563 | 0 | 0 | 0 | 0 | 0 | 0 | 0 | 0 | 0 | 0 | 0 | 0 | 0 | 0 | 0 | **0.01** | 0 | 0 | 0 | 0 | 0 | 0 | 0 | 0 |
| | FL580 | 0 | 0 | 0 | 0 | 0 | 0 | 0 | 0 | 0 | 0 | 0 | 0 | 0 | 0 | 0 | 0 | 0 | 0 | **0.01** | 0 | 0 | 0 | **0.02** | 0 |
| | FL581 | 0 | 0 | 0 | 0 | 0 | 0 | 0 | 0 | 0 | 0 | 0 | 0 | 0 | 0 | 0 | 0 | 0 | 0 | 0 | 0 | **0.01** | 0 | 0 | 0 |
| | FL587 | 0 | 0 | 0 | 0 | **0.12** | 0 | 0 | 0 | 0 | 0 | 0 | 0 | 0 | 0 | 0 | 0 | 0 | 0 | 0 | 0 | 0 | 0 | 0 | 0 |
| | FL596 | 0 | 0 | 0 | 0 | 0 | 0 | 0 | 0 | 0 | 0 | 0 | 0 | 0 | 0 | 0 | 0 | 0 | 0 | 0 | 0 | **0.05** | 0 | 0 | 0 |
| | FL600 | 0 | 0 | 0 | 0 | 0 | 0 | 0 | 0 | **0.02** | 0 | 0 | 0 | 0 | **0.01** | 0 | 0 | 0 | 0 | 0 | 0 | **0.26** | 0 | 0 | 0 |
| | FL619 | 0 | 0 | 0 | 0 | 0 | 0 | 0 | 0 | **0.02** | 0 | 0 | 0 | 0 | 0 | 0 | 0 | 0 | 0 | 0 | 0 | 0 | 0 | 0 | 0 |

Table 12: Donor weights for Yishun sectors for model 2. Only sectors with any contribution to synthetic control weights for intervention sectors' synthetic controls are shown

| Donor | | Model 2 | | | | | | | | | | | | | | | | |
|---|---|---|---|---|---|---|---|---|---|---|---|---|---|---|---|---|---|---|
| Town | Sector | FL278 | FL283 | FL285 | FL290 | FL291 | FL295 | FL296 | FL297 | FL298 | FL420 | FL421 | FL517 | FL523 | FL528 | FL540 | FL557 | FL558 |
| Ang Mo Kio | FL18 | 0 | 0 | 0 | 0 | 0 | 0 | 0 | 0 | 0 | 0 | 0 | 0 | 0 | 0 | 0 | 0 | **0.14** |
| | FL454 | **0.02** | **0.11** | 0 | 0 | 0 | 0 | 0 | 0 | **0.01** | 0 | 0 | 0 | 0 | 0 | 0 | 0 | **0.02** |
| | FL456 | 0 | 0 | 0 | 0 | 0 | 0 | 0 | 0 | **0.04** | 0 | 0 | 0 | 0 | **0.33** | 0 | 0 | 0 |
| | FL471 | 0 | **0.01** | 0 | 0 | 0 | 0 | 0 | 0 | 0 | 0 | 0 | 0 | 0 | **0.08** | 0 | 0 | **0.04** |
| | FL478 | **0.04** | 0 | 0 | 0 | 0 | 0 | 0 | 0 | 0 | 0 | 0 | 0 | 0 | 0 | 0 | 0 | 0 |
| | FL499 | 0 | 0 | 0 | 0 | 0 | 0 | **0.31** | 0 | 0 | 0 | 0 | 0 | 0 | 0 | **0.26** | 0 | **0.20** |
| | FL500 | 0 | 0 | 0 | 0 | **0.17** | 0 | 0 | 0 | 0 | 0 | 0 | 0 | 0 | 0 | 0 | 0 | 0 |
| Bedok | FL239 | 0 | 0 | 0 | 0 | 0 | **0.14** | 0 | 0 | 0 | 0 | 0 | 0 | 0 | 0 | 0 | 0 | 0 |
| | FL303 | 0 | 0 | 0 | **0.03** | 0 | 0 | 0 | 0 | 0 | 0 | 0 | 0 | 0 | 0 | 0 | 0 | 0 |
| Bishan | FL34 | 0 | 0 | 0 | 0 | 0 | 0 | 0 | **0.46** | 0 | 0 | 0 | 0 | 0 | 0 | 0 | 0 | 0 |
| | FL36 | 0 | 0 | 0 | 0 | 0 | 0 | 0 | 0 | 0 | 0 | 0 | 0 | 0 | **0.11** | 0 | 0 | 0 |
| Bukit Batok | FL434 | 0 | 0 | 0 | **0.31** | 0 | 0 | 0 | 0 | 0 | 0 | 0 | 0 | 0 | 0 | 0 | 0 | 0 |
| Bukit Merah | FL144 | 0 | 0 | 0 | 0 | 0 | 0 | 0 | 0 | 0 | 0 | 0 | 0 | 0 | **0.22** | 0 | 0 | 0 |
| | FL160 | 0 | 0 | 0 | 0 | 0 | 0 | 0 | 0 | 0 | 0 | **0.15** | 0 | 0 | 0 | 0 | 0 | 0 |
| | FL174 | 0 | 0 | 0 | 0 | 0 | **0.12** | 0 | 0 | 0 | 0 | 0 | 0 | 0 | 0 | 0 | 0 | 0 |
| | FL179 | 0 | 0 | 0 | 0 | 0 | 0 | 0 | 0 | **0.02** | 0 | **0.09** | 0 | 0 | 0 | 0 | 0 | 0 |
| | FL193 | **0.21** | 0 | 0 | 0 | 0 | 0 | 0 | 0 | 0 | **0.13** | 0 | 0 | 0 | 0 | **0.47** | 0 | 0 |
| | FL194 | 0 | 0 | **0.09** | 0 | 0 | 0 | 0 | 0 | 0 | 0 | **0.19** | 0 | 0 | 0 | 0 | 0 | 0 |
| | FL7 | 0 | 0 | 0 | **0.29** | 0 | 0 | 0 | 0 | 0 | 0 | 0 | 0 | 0 | 0 | 0 | 0 | 0 |
| Bukit Panjang | FL659 | 0 | **0.01** | **0.14** | 0 | 0 | 0 | 0 | 0 | 0 | 0 | 0 | 0 | 0 | 0 | 0 | 0 | 0 |
| | FL829 | 0 | 0 | 0 | **0.13** | **0.12** | 0 | 0 | 0 | 0 | 0 | 0 | **0.35** | 0 | 0 | 0 | 0 | 0 |
| Choa Chu Kang | FL729 | 0 | 0 | 0 | 0 | 0 | 0 | **0.22** | 0 | **0.24** | 0 | 0 | 0 | 0 | 0 | 0 | 0 | 0 |
| | FL764 | 0 | 0 | 0 | 0 | 0 | 0 | 0 | 0 | **0.25** | **0.02** | 0 | **0.18** | 0 | 0 | 0 | 0 | 0 |
| | FL811 | 0 | 0 | 0 | 0 | 0 | **0.17** | 0 | 0 | 0 | **0.16** | 0 | 0 | 0 | 0 | 0 | 0 | 0 |
| | FL818 | 0 | 0 | 0 | 0 | **0.11** | 0 | 0 | 0 | 0 | 0 | 0 | 0 | 0 | 0 | **0.20** | **0.30** | 0 |
| | FL823 | 0 | 0 | 0 | 0 | 0 | **0.10** | 0 | 0 | 0 | 0 | **0.30** | 0 | 0 | 0 | 0 | 0 | 0 |
| Geylang | FL200 | **0.31** | 0 | 0 | 0 | 0 | 0 | 0 | 0 | 0 | 0 | 0 | 0 | **0.06** | 0 | 0 | 0 | 0 |
| Hougang | FL375 | 0 | 0 | 0 | 0 | 0 | 0 | 0 | 0 | 0 | 0 | 0 | 0 | **0.29** | 0 | 0 | 0 | 0 |
| | FL451 | 0 | 0 | 0 | 0 | 0 | 0 | 0 | 0 | 0 | 0 | 0 | 0 | **0.39** | 0 | 0 | 0 | 0 |
| Jurong East | FL697 | 0 | 0 | 0 | 0 | 0 | 0 | 0 | 0 | **0.21** | 0 | 0 | 0 | 0 | 0 | 0 | 0 | 0 |
| Jurong West | FL441 | 0 | 0 | 0 | 0 | **0.08** | 0 | 0 | 0 | 0 | 0 | 0 | 0 | 0 | 0 | 0 | 0 | 0 |
| | FL621 | 0 | 0 | 0 | 0 | **0.43** | 0 | 0 | **0.38** | 0 | 0 | 0 | 0 | 0 | 0 | 0 | 0 | 0 |
| | FL664 | 0 | 0 | 0 | 0 | 0 | 0 | 0 | 0 | 0 | **0.09** | 0 | 0 | 0 | 0 | 0 | 0 | 0 |
| Kallang | FL14 | 0 | 0 | 0 | 0 | 0 | **0.10** | 0 | 0 | **0.14** | 0 | 0 | 0 | 0 | 0 | 0 | 0 | 0 |
| | FL181 | 0 | 0 | **0.15** | 0 | 0 | 0 | 0 | 0 | 0 | 0 | 0 | 0 | 0 | 0 | 0 | 0 | 0 |
| | FL182 | 0 | 0 | **0.06** | 0 | 0 | 0 | 0 | 0 | 0 | 0 | 0 | 0 | 0 | 0 | 0 | 0 | 0 |
| | FL185 | 0 | 0 | 0 | 0 | 0 | 0 | 0 | 0 | 0 | 0 | 0 | 0 | 0 | 0 | **0.06** | 0 | 0 |
| | FL26 | 0 | **0.01** | 0 | 0 | 0 | 0 | 0 | 0 | 0 | 0 | 0 | **0.10** | 0 | 0 | 0 | 0 | 0 |
| | FL85 | 0 | 0 | 0 | 0 | 0 | 0 | 0 | 0 | 0 | 0 | 0 | 0 | 0 | 0 | 0 | **0.10** | 0 |
| Marine Parade | FL322 | 0 | 0 | 0 | 0 | 0 | **0.14** | 0 | 0 | 0 | 0 | 0 | 0 | 0 | 0 | 0 | 0 | 0 |
| Pasir Ris | FL166 | 0 | 0 | **0.01** | 0 | 0 | 0 | **0.10** | 0 | 0 | 0 | 0 | 0 | 0 | 0 | 0 | 0 | **0.07** |
| | FL217 | 0 | 0 | 0 | 0 | 0 | 0 | 0 | 0 | 0 | 0 | 0 | 0 | **0.05** | 0 | 0 | 0 | 0 |
| | FL272 | 0 | 0 | **0.07** | 0 | 0 | 0 | 0 | 0 | 0 | 0 | 0 | 0 | 0 | 0 | 0 | 0 | **0.31** |
| | FL321 | 0 | **0.02** | 0 | 0 | 0 | 0 | 0 | 0 | 0 | **0.09** | 0 | 0 | 0 | 0 | 0 | 0 | 0 |
| | FL323 | 0 | 0 | 0 | 0 | 0 | 0 | **0.20** | 0 | 0 | 0 | 0 | 0 | 0 | 0 | 0 | 0 | 0 |
| | FL336 | 0 | 0 | 0 | 0 | 0 | 0 | 0 | 0 | 0 | 0 | **0.08** | 0 | 0 | 0 | 0 | 0 | 0 |
| | FL348 | 0 | **0.02** | 0 | **0.25** | 0 | 0 | 0 | 0 | 0 | 0 | 0 | 0 | 0 | 0 | 0 | 0 | 0 |
| Queenstown | FL339 | 0 | 0 | 0 | 0 | 0 | 0 | 0 | 0 | **0.01** | 0 | 0 | 0 | 0 | 0 | 0 | 0 | 0 |
| Rochor | FL71 | 0 | 0 | 0 | 0 | 0 | 0 | 0 | 0 | 0 | 0 | **0.15** | 0 | 0 | 0 | 0 | **0.07** | 0 |
| | FL73 | **0.03** | 0 | 0 | 0 | 0 | 0 | 0 | 0 | 0 | 0 | 0 | 0 | 0 | 0 | 0 | 0 | 0 |
| | FL76 | 0 | 0 | 0 | 0 | 0 | 0 | 0 | 0 | 0 | 0 | 0 | 0 | 0 | **0.09** | 0 | 0 | 0 |
| Sembawang | FL527 | **0.31** | **0.82** | **0.05** | 0 | 0 | 0 | **0.02** | 0 | 0 | 0 | 0 | 0 | 0 | 0 | 0 | **0.04** | **0.09** |
| Sengkang | FL133 | 0 | 0 | **0.09** | 0 | 0 | 0 | 0 | 0 | 0 | 0 | 0 | 0 | 0 | 0 | 0 | 0 | 0 |
| | FL497 | 0 | 0 | 0 | 0 | 0 | 0 | 0 | **0.16** | 0 | 0 | 0 | 0 | 0 | 0 | 0 | 0 | 0 |
| Serangoon | FL461 | 0 | 0 | 0 | 0 | 0 | 0 | 0 | 0 | 0 | 0 | **0.04** | 0 | 0 | 0 | 0 | 0 | 0 |
| | FL480 | 0 | 0 | 0 | 0 | 0 | 0 | 0 | 0 | 0 | 0 | 0 | 0 | 0 | **0.13** | 0 | 0 | 0 |
| Toa Payoh | FL42 | 0 | 0 | 0 | 0 | 0 | 0 | 0 | 0 | 0 | 0 | 0 | 0 | 0 | **0.03** | 0 | **0.30** | 0 |
| | FL58 | 0 | 0 | 0 | 0 | 0 | 0 | **0.15** | 0 | 0 | 0 | 0 | 0 | 0 | 0 | 0 | 0 | 0 |
| | FL59 | 0 | 0 | 0 | 0 | 0 | 0 | 0 | 0 | 0 | 0 | 0 | **0.20** | **0.20** | 0 | 0 | **0.03** | 0 |
| | FL60 | 0 | 0 | 0 | 0 | 0 | 0 | 0 | 0 | 0 | 0 | 0 | **0.15** | 0 | 0 | 0 | **0.16** | 0 |
| | FL65 | 0 | 0 | 0 | 0 | 0 | 0 | 0 | 0 | 0 | 0 | 0 | **0.02** | 0 | 0 | 0 | 0 | 0 |
| Woodlands | FL226 | 0 | 0 | 0 | 0 | 0 | 0 | 0 | 0 | **0.02** | 0 | 0 | 0 | 0 | 0 | 0 | 0 | 0 |
| | FL245 | 0 | 0 | 0 | 0 | 0 | **0.23** | 0 | 0 | 0 | **0.21** | 0 | 0 | 0 | 0 | 0 | 0 | 0 |
| | FL572 | 0 | 0 | 0 | 0 | 0 | 0 | 0 | 0 | **0.07** | 0 | 0 | 0 | 0 | 0 | 0 | 0 | 0 |
| | FL580 | **0.07** | 0 | 0 | 0 | **0.09** | 0 | 0 | 0 | 0 | 0 | 0 | 0 | 0 | 0 | 0 | 0 | **0.13** |
| | FL596 | 0 | 0 | 0 | 0 | 0 | 0 | 0 | 0 | 0 | **0.15** | 0 | 0 | 0 | 0 | 0 | 0 | 0 |
| | FL599 | 0 | 0 | **0.34** | 0 | 0 | 0 | 0 | 0 | 0 | **0.13** | 0 | 0 | 0 | 0 | 0 | 0 | 0 |

Table 13: Donor weights for Yishun sectors for model 2. Only sectors with any contribution to synthetic control weights for intervention sectors' synthetic controls are shown

| Donor | | Model 2 | | | | | | | | | | | | | | |
|---|---|---|---|---|---|---|---|---|---|---|---|---|---|---|---|---|
| Town | Sector | FL564 | FL565 | FL569 | FL573 | FL576 | FL582 | FL584 | FL585 | FL585b | FL588 | FL589 | FL602 | FL609 | FL610 | FL618 |
| Ang Mo Kio | FL18 | 0 | 0 | 0 | 0 | 0 | 0 | 0 | 0 | 0 | 0 | 0 | **0.16** | **0.08** | 0 | 0 |
| | FL454 | 0 | 0 | 0 | 0 | 0 | 0 | **0.15** | **0.04** | **0.05** | **0.23** | 0 | **0.05** | 0 | 0 | 0 |
| | FL471 | 0 | 0 | 0 | 0 | 0 | 0 | 0 | 0 | **0.16** | 0 | 0 | 0 | 0 | 0 | 0 |
| | FL477 | 0 | 0 | 0 | 0 | 0 | **0.03** | 0 | 0 | 0 | 0 | 0 | **0.09** | 0 | 0 | 0 |
| | FL488 | 0 | 0 | 0 | 0 | 0 | 0 | 0 | 0 | 0 | 0 | 0 | 0 | **0.34** | 0 | 0 |
| | FL499 | 0 | 0 | 0 | 0 | 0 | 0 | 0 | 0 | 0 | 0 | 0 | **0.03** | **0.13** | 0 | 0 |
| | FL500 | 0 | 0 | **0.19** | 0 | **0.35** | 0 | 0 | 0 | 0 | 0 | 0 | 0 | 0 | 0 | 0 |
| | FL505 | 0 | 0 | 0 | 0 | **0.09** | 0 | 0 | 0 | 0 | 0 | 0 | 0 | 0 | 0 | 0 |
| Bedok | FL231 | 0 | 0 | 0 | 0 | 0 | 0 | 0 | 0 | 0 | 0 | 0 | 0 | 0 | **0.10** | 0 |
| | FL259 | 0 | 0 | 0 | 0 | 0 | 0 | 0 | 0 | 0 | 0 | 0 | 0 | 0 | 0 | **0.02** |
| Bishan | FL34 | 0 | 0 | 0 | 0 | 0 | **0.16** | **0.23** | 0 | 0 | 0 | 0 | 0 | 0 | **0.25** | 0 |
| Bukit Batok | FL699 | 0 | 0 | 0 | 0 | 0 | 0 | 0 | 0 | 0 | 0 | **0.41** | 0 | 0 | 0 | 0 |
| | FL731 | 0 | 0 | 0 | 0 | 0 | 0 | 0 | 0 | 0 | 0 | 0 | 0 | 0 | **0.10** | 0 |
| Bukit Merah | FL144 | 0 | 0 | 0 | 0 | 0 | 0 | 0 | 0 | 0 | 0 | 0 | 0 | **0.21** | 0 | 0 |
| | FL152 | 0 | 0 | 0 | 0 | 0 | 0 | 0 | 0 | 0 | 0 | 0 | 0 | 0 | **0.10** | 0 |
| | FL161 | **0.38** | 0 | 0 | 0 | 0 | 0 | 0 | 0 | 0 | 0 | 0 | 0 | 0 | 0 | 0 |
| | FL193 | 0 | 0 | 0 | 0 | 0 | 0 | 0 | 0 | 0 | 0 | 0 | 0 | 0 | 0 | 0 |
| Bukit Panjang | FL647 | 0 | 0 | 0 | 0 | 0 | 0 | 0 | 0 | 0 | **0.26** | 0 | 0 | 0 | 0 | 0 |
| | FL648 | 0 | 0 | 0 | **0.20** | 0 | 0 | 0 | 0 | 0 | 0 | 0 | 0 | 0 | 0 | 0 |
| | FL659 | **0.21** | 0 | 0 | 0 | 0 | 0 | 0 | 0 | 0 | 0 | 0 | 0 | 0 | 0 | 0 |
| | FL829 | 0 | 0 | 0 | 0 | 0 | 0 | 0 | 0 | **0.14** | **0.12** | 0 | 0 | 0 | 0 | 0 |
| Choa Chu Kang | FL729 | 0 | 0 | 0 | 0 | 0 | **0.09** | 0 | 0 | 0 | 0 | **0.12** | **0.21** | **0.10** | 0 | 0 |
| | FL811 | 0 | 0 | 0 | 0 | 0 | **0.21** | **0.08** | **0.12** | 0 | 0 | 0 | 0 | **0.14** | 0 | 0 |
| | FL814 | 0 | 0 | 0 | 0 | 0 | 0 | **0.08** | 0 | 0 | 0 | 0 | 0 | 0 | 0 | 0 |
| | FL818 | 0 | 0 | 0 | **0.32** | 0 | 0 | **0.23** | **0.04** | **0.27** | **0.20** | 0 | 0 | 0 | 0 | 0 |
| | FL823 | 0 | 0 | 0 | 0 | 0 | 0 | 0 | 0 | 0 | **0.01** | 0 | 0 | 0 | 0 | 0 |
| Geylang | FL19 | **0.01** | 0 | 0 | 0 | 0 | 0 | 0 | 0 | 0 | 0 | 0 | 0 | 0 | 0 | 0 |
| Hougang | FL367 | 0 | 0 | 0 | 0 | 0 | 0 | 0 | **0.15** | 0 | 0 | 0 | 0 | 0 | 0 | 0 |
| | FL408 | 0 | 0 | 0 | 0 | 0 | 0 | 0 | 0 | 0 | 0 | 0 | 0 | 0 | 0 | **0.18** |
| | FL451 | 0 | 0 | **0.32** | 0 | 0 | 0 | 0 | 0 | 0 | 0 | **0.26** | 0 | 0 | 0 | 0 |
| Jurong East | FL696 | 0 | 0 | 0 | **0.05** | 0 | 0 | 0 | 0 | 0 | 0 | 0 | 0 | 0 | 0 | 0 |
| | FL697 | 0 | 0 | 0 | **0.14** | 0 | 0 | 0 | 0 | 0 | 0 | 0 | 0 | 0 | 0 | 0 |
| Jurong West | FL441 | 0 | 0 | **0.22** | 0 | 0 | **0.30** | **0.07** | 0 | 0 | 0 | 0 | 0 | 0 | 0 | **0.36** |
| | FL621 | 0 | 0 | 0 | 0 | 0 | 0 | 0 | **0.24** | 0 | 0 | **0.20** | 0 | 0 | 0 | 0 |
| | FL652 | 0 | 0 | 0 | 0 | 0 | 0 | 0 | 0 | 0 | 0 | 0 | 0 | 0 | **0.06** | 0 |
| | FL694 | 0 | 0 | 0 | 0 | 0 | 0 | 0 | **0.32** | **0.03** | 0 | 0 | 0 | 0 | 0 | 0 |
| | FL769 | 0 | 0 | 0 | 0 | 0 | 0 | 0 | **0.09** | 0 | 0 | 0 | 0 | 0 | 0 | 0 |
| Marine Parade | FL311 | 0 | 0 | **0.22** | 0 | 0 | 0 | 0 | 0 | 0 | 0 | 0 | 0 | 0 | 0 | 0 |
| Pasir Ris | FL217 | 0 | 0 | 0 | 0 | **0.22** | 0 | 0 | 0 | 0 | 0 | 0 | 0 | 0 | 0 | 0 |
| | FL227 | 0 | 0 | 0 | 0 | 0 | 0 | 0 | 0 | 0 | 0 | 0 | 0 | 0 | 0 | **0.04** |
| | FL228 | 0 | 0 | 0 | 0 | 0 | 0 | 0 | 0 | 0 | **0.07** | 0 | 0 | 0 | 0 | 0 |
| | FL264 | 0 | 0 | 0 | 0 | **0.09** | 0 | 0 | 0 | 0 | 0 | 0 | 0 | 0 | 0 | 0 |
| | FL293 | 0 | 0 | 0 | 0 | 0 | 0 | **0.16** | 0 | 0 | **0.10** | 0 | 0 | 0 | 0 | 0 |
| | FL321 | 0 | 0 | 0 | 0 | 0 | 0 | 0 | 0 | 0 | 0 | 0 | 0 | 0 | 0 | **0.01** |
| | FL348 | 0 | 0 | 0 | 0 | 0 | 0 | 0 | 0 | 0 | **0.02** | 0 | **0.31** | 0 | 0 | 0 |
| Rochor | FL76 | **0.14** | 0 | 0 | 0 | 0 | 0 | 0 | 0 | 0 | 0 | 0 | 0 | 0 | 0 | 0 |
| Sembawang | FL527 | **0.16** | **0.54** | **0.06** | **0.11** | 0 | 0 | 0 | 0 | 0 | 0 | 0 | **0.14** | 0 | 0 | **0.02** |
| Sengkang | FL136 | 0 | **0.45** | 0 | 0 | **0.02** | 0 | 0 | 0 | 0 | 0 | 0 | 0 | 0 | 0 | 0 |
| | FL368 | 0 | 0 | 0 | 0 | 0 | **0.08** | 0 | 0 | 0 | 0 | **0.02** | **0.01** | 0 | 0 | 0 |
| | FL497 | 0 | 0 | 0 | **0.18** | 0 | 0 | 0 | 0 | 0 | 0 | 0 | 0 | 0 | 0 | 0 |
| | FL535 | 0 | 0 | 0 | 0 | 0 | **0.13** | 0 | 0 | 0 | 0 | 0 | 0 | 0 | 0 | 0 |
| | FL733 | 0 | 0 | 0 | 0 | 0 | 0 | 0 | 0 | 0 | 0 | 0 | 0 | 0 | 0 | 0 |
| Serangoon | FL393 | 0 | 0 | 0 | 0 | 0 | 0 | 0 | 0 | 0 | 0 | 0 | 0 | 0 | 0 | **0.06** |
| | FL480 | 0 | 0 | 0 | 0 | 0 | 0 | 0 | 0 | 0 | 0 | 0 | 0 | 0 | 0 | **0.18** |
| | FL501 | 0 | 0 | 0 | 0 | 0 | 0 | 0 | 0 | 0 | 0 | 0 | 0 | 0 | 0 | **0.13** |
| Toa Payoh | FL164 | 0 | 0 | 0 | 0 | 0 | 0 | 0 | 0 | 0 | 0 | 0 | 0 | 0 | **0.39** | 0 |
| | FL359 | 0 | 0 | 0 | 0 | **0.18** | 0 | 0 | 0 | 0 | 0 | 0 | 0 | 0 | 0 | 0 |
| Woodlands | FL508 | **0.06** | **0.01** | 0 | 0 | **0.06** | 0 | 0 | 0 | 0 | 0 | 0 | 0 | 0 | 0 | 0 |
| | FL575 | 0 | 0 | 0 | 0 | 0 | 0 | 0 | 0 | **0.04** | 0 | 0 | 0 | 0 | 0 | 0 |
| | FL580 | 0 | 0 | 0 | 0 | 0 | 0 | 0 | 0 | **0.30** | 0 | 0 | 0 | 0 | 0 | 0 |
| | FL598 | 0 | 0 | 0 | 0 | 0 | 0 | 0 | 0 | 0 | 0 | 0 | 0 | 0 | 0 | 0 |
| | FL599 | **0.04** | 0 | 0 | 0 | 0 | 0 | 0 | 0 | 0 | 0 | 0 | 0 | 0 | 0 | 0 |
| | FL617 | 0 | 0 | 0 | 0 | 0 | 0 | 0 | 0 | 0 | 0 | **0.01** | 0 | 0 | 0 | 0 |

Table 14: Donor weights for Yishun sectors for models 3 and 4. Only sectors with any contribution to synthetic control weights for intervention sectors' synthetic controls are shown

| Donor | | Model 3 | | | | | | | | | | | | Model 4 | | | | | | | | | | | |
|---|---|---|---|---|---|---|---|---|---|---|---|---|---|---|---|---|---|---|---|---|---|---|---|---|---|
| Town | Sector | FL278 | FL283 | FL285 | FL290 | FL291 | FL295 | FL296 | FL297 | FL298 | FL420 | FL421 | FL517 | FL278 | FL283 | FL285 | FL290 | FL291 | FL295 | FL296 | FL297 | FL298 | FL420 | FL421 | FL517 |
| Ang Mo Kio | FL17 | 0 | 0 | 0 | 0 | 0 | 0 | 0 | 0 | 0 | 0 | 0 | 0 | 0 | 0 | 0 | 0 | 0 | 0 | 0 | 0 | **0.01** | 0 | 0 | 0 |
| | FL18 | 0 | 0 | 0 | **0.07** | 0 | 0 | 0 | 0 | 0 | 0 | 0 | 0 | 0 | 0 | **0.08** | 0 | 0 | 0 | 0 | 0 | 0 | 0 | 0 | 0 |
| | FL360 | 0 | 0 | 0 | 0 | 0 | 0 | 0 | 0 | 0 | 0 | 0 | 0 | 0 | 0 | 0 | 0 | 0 | **0.01** | 0 | 0 | 0 | **0.11** | 0 | 0 |
| | FL362 | 0 | 0 | 0 | 0 | 0 | **0.12** | 0 | 0 | 0 | 0 | 0 | 0 | 0 | 0 | 0 | 0 | 0 | 0 | 0 | 0 | 0 | 0 | 0 | 0 |
| | FL386 | 0 | 0 | 0 | 0 | 0 | **0.18** | 0 | 0 | 0 | 0 | 0 | 0 | 0 | 0 | 0 | 0 | 0 | 0 | 0 | 0 | 0 | 0 | 0 | 0 |
| | FL454 | 0 | 0 | 0 | 0 | 0 | **0.01** | 0 | 0 | **0.01** | 0 | 0 | 0 | 0 | 0 | 0 | 0 | 0 | 0 | 0 | 0 | 0 | 0 | 0 | 0 |
| | FL456 | 0 | 0 | 0 | 0 | 0 | 0 | 0 | 0 | 0 | 0 | 0 | 0 | 0 | 0 | 0 | 0 | 0 | 0 | 0 | 0 | **0.03** | 0 | 0 | 0 |
| | FL477 | 0 | 0 | 0 | 0 | 0 | 0 | 0 | 0 | 0 | 0 | 0 | 0 | 0 | 0 | 0 | 0 | 0 | 0 | 0 | 0 | **0.02** | 0 | 0 | 0 |
| | FL478 | 0 | 0 | 0 | 0 | 0 | 0 | 0 | 0 | 0 | 0 | 0 | 0 | 0 | 0 | **0.01** | **0.04** | 0 | 0 | 0 | 0 | 0 | 0 | 0 | 0 |
| | FL487 | 0 | 0 | 0 | 0 | 0 | 0 | 0 | 0 | 0 | 0 | 0 | 0 | 0 | 0 | **0.03** | 0 | 0 | 0 | 0 | 0 | 0 | 0 | 0 | 0 |
| | FL488 | 0 | 0 | 0 | 0 | 0 | 0 | 0 | 0 | 0 | 0 | 0 | 0 | 0 | 0 | 0 | **0.11** | **0.01** | 0 | **0.08** | 0 | 0 | 0 | 0 | 0 |
| | FL499 | 0 | 0 | 0 | **0.04** | 0 | 0 | **0.14** | 0 | **0.07** | 0 | 0 | 0 | 0 | 0 | 0 | 0 | 0 | 0 | 0 | 0 | 0 | 0 | 0 | 0 |
| | FL500 | 0 | 0 | 0 | 0 | 0 | 0 | **0.22** | 0 | 0 | 0 | 0 | 0 | 0 | 0 | 0 | 0 | **0.08** | 0 | **0.05** | 0 | 0 | 0 | 0 | 0 |
| | FL505 | 0 | 0 | 0 | 0 | 0 | 0 | 0 | 0 | 0 | 0 | 0 | 0 | 0 | 0 | 0 | 0 | 0 | **0.05** | 0 | 0 | 0 | 0 | 0 | 0 |
| Bedok | FL211 | 0 | 0 | 0 | 0 | 0 | 0 | 0 | 0 | 0 | 0 | 0 | 0 | 0 | 0 | 0 | 0 | 0 | 0 | 0 | 0 | 0 | 0 | 0 | **0.02** |
| | FL230 | 0 | 0 | 0 | 0 | 0 | 0 | 0 | 0 | 0 | 0 | 0 | 0 | 0 | 0 | 0 | **0.03** | 0 | 0 | 0 | 0 | 0 | 0 | 0 | 0 |
| | FL237 | 0 | 0 | 0 | 0 | 0 | 0 | 0 | **0.01** | 0 | 0 | 0 | 0 | 0 | 0 | 0 | 0 | 0 | 0 | **0.02** | **0.01** | 0 | 0 | 0 | 0 |
| | FL241 | 0 | 0 | 0 | 0 | 0 | 0 | 0 | 0 | 0 | 0 | 0 | 0 | 0 | 0 | 0 | 0 | 0 | 0 | 0 | 0 | 0 | 0 | **0.08** | 0 |
| | FL303 | 0 | 0 | 0 | 0 | 0 | 0 | 0 | 0 | 0 | 0 | 0 | 0 | 0 | 0 | 0 | **0.03** | 0 | 0 | 0 | 0 | 0 | 0 | 0 | 0 |
| Bishan | FL34 | 0 | 0 | 0 | 0 | 0 | 0 | 0 | **0.47** | 0 | 0 | 0 | 0 | 0 | 0 | 0 | **0.15** | **0.10** | 0 | **0.10** | **0.47** | 0 | 0 | 0 | 0 |
| | FL47 | **0.24** | 0 | 0 | 0 | 0 | 0 | 0 | 0 | 0 | 0 | 0 | 0 | **0.25** | 0 | 0 | 0 | 0 | 0 | 0 | 0 | 0 | 0 | 0 | 0 |
| | FL51 | 0 | 0 | 0 | 0 | 0 | 0 | 0 | 0 | 0 | 0 | 0 | 0 | 0 | 0 | 0 | 0 | 0 | 0 | 0 | 0 | 0 | 0 | 0 | **0.12** |
| Bukit Batok | FL435 | 0 | 0 | 0 | 0 | 0 | 0 | 0 | 0 | 0 | 0 | 0 | 0 | 0 | 0 | 0 | 0 | 0 | 0 | 0 | 0 | **0.01** | 0 | 0 | 0 |
| | FL680 | 0 | 0 | 0 | 0 | 0 | 0 | 0 | 0 | 0 | 0 | **0.07** | 0 | 0 | 0 | 0 | 0 | 0 | 0 | 0 | 0 | 0 | 0 | 0 | 0 |
| | FL686 | 0 | 0 | 0 | 0 | 0 | 0 | 0 | 0 | 0 | 0 | 0 | 0 | 0 | 0 | 0 | **0.04** | 0 | 0 | 0 | 0 | 0 | 0 | 0 | 0 |
| | FL699 | 0 | 0 | 0 | 0 | 0 | 0 | 0 | 0 | 0 | 0 | 0 | 0 | 0 | 0 | 0 | **0.20** | 0 | 0 | **0.04** | 0 | 0 | 0 | 0 | 0 |
| | FL731 | 0 | 0 | 0 | 0 | **0.10** | 0 | 0 | 0 | 0 | 0 | 0 | 0 | 0 | 0 | 0 | 0 | **0.09** | 0 | 0 | 0 | 0 | 0 | 0 | 0 |
| Bukit Merah | FL147 | 0 | 0 | 0 | 0 | 0 | 0 | 0 | 0 | 0 | 0 | 0 | 0 | 0 | 0 | 0 | 0 | 0 | **0.05** | 0 | 0 | 0 | 0 | 0 | **0.02** |
| | FL152 | 0 | 0 | 0 | 0 | 0 | **0.08** | 0 | 0 | 0 | 0 | 0 | 0 | 0 | 0 | 0 | 0 | 0 | **0.03** | 0 | 0 | 0 | 0 | 0 | 0 |
| | FL160 | 0 | 0 | 0 | 0 | 0 | 0 | 0 | 0 | 0 | 0 | **0.06** | 0 | 0 | 0 | 0 | 0 | 0 | **0.02** | 0 | 0 | 0 | **0.06** | 0 | 0 |
| | FL174 | 0 | 0 | 0 | 0 | 0 | 0 | 0 | 0 | 0 | 0 | 0 | 0 | 0 | 0 | 0 | 0 | 0 | 0 | 0 | 0 | 0 | 0 | 0 | **0.01** |
| | FL176 | 0 | 0 | 0 | 0 | 0 | 0 | 0 | 0 | 0 | 0 | 0 | 0 | 0 | 0 | 0 | 0 | 0 | 0 | 0 | 0 | 0 | **0.09** | 0 | 0 |
| | FL179 | 0 | 0 | 0 | 0 | 0 | 0 | 0 | 0 | **0.03** | 0 | 0 | 0 | 0 | 0 | **0.02** | 0 | 0 | 0 | 0 | 0 | 0 | 0 | 0 | 0 |
| | FL193 | **0.05** | 0 | 0 | 0 | 0 | 0 | 0 | 0 | 0 | **0.16** | 0 | **0.13** | **0.01** | **0.05** | 0 | 0 | 0 | 0 | 0 | 0 | 0 | **0.14** | 0 | 0 |
| | FL194 | 0 | 0 | 0 | 0 | 0 | 0 | 0 | 0 | 0 | **0.12** | **0.24** | 0 | 0 | 0 | 0 | 0 | 0 | 0 | 0 | 0 | 0 | 0 | **0.12** | 0 |
| | FL7 | 0 | 0 | 0 | **0.27** | 0 | 0 | 0 | 0 | 0 | 0 | 0 | 0 | 0 | 0 | 0 | **0.14** | 0 | 0 | 0 | 0 | **0.04** | 0 | 0 | 0 |
| | FL8 | 0 | 0 | 0 | **0.07** | 0 | 0 | 0 | 0 | 0 | 0 | 0 | 0 | 0 | 0 | 0 | 0 | 0 | 0 | 0 | 0 | 0 | 0 | 0 | 0 |
| Bukit Panjang | FL647 | 0 | 0 | 0 | 0 | 0 | 0 | 0 | 0 | 0 | 0 | 0 | **0.09** | 0 | 0 | 0 | **0.01** | 0 | 0 | **0.01** | 0 | 0 | 0 | 0 | **0.04** |
| | FL648 | 0 | 0 | 0 | 0 | 0 | 0 | 0 | 0 | 0 | 0 | **0.07** | 0 | 0 | 0 | 0 | 0 | 0 | **0.02** | 0 | 0 | 0 | 0 | 0 | 0 |
| | FL659 | 0 | 0 | **0.14** | 0 | 0 | 0 | 0 | 0 | 0 | 0 | **0.17** | 0 | 0 | 0 | **0.09** | 0 | 0 | 0 | 0 | 0 | 0 | 0 | 0 | 0 |
| | FL829 | 0 | 0 | 0 | 0 | 0 | 0 | 0 | 0 | 0 | 0 | 0 | 0 | 0 | 0 | 0 | 0 | 0 | 0 | **0.05** | 0 | 0 | 0 | 0 | 0 |
| Choa Chu Kang | FL729 | 0 | 0 | 0 | **0.02** | 0 | 0 | 0 | 0 | **0.27** | 0 | 0 | **0.09** | 0 | 0 | 0 | 0 | 0 | 0 | 0 | 0 | **0.11** | 0 | 0 | 0 |
| | FL764 | 0 | 0 | 0 | 0 | 0 | 0 | 0 | 0 | **0.17** | 0 | 0 | **0.25** | 0 | 0 | 0 | 0 | 0 | 0 | **0.08** | 0 | **0.02** | 0 | 0 | **0.06** |
| | FL811 | 0 | 0 | **0.04** | 0 | 0 | **0.12** | 0 | 0 | 0 | 0 | 0 | 0 | 0 | 0 | 0 | 0 | 0 | **0.02** | 0 | 0 | 0 | 0 | 0 | 0 |
| | FL814 | 0 | 0 | 0 | 0 | 0 | **0.12** | 0 | 0 | 0 | **0.15** | 0 | **0.23** | 0 | 0 | 0 | 0 | 0 | **0.04** | 0 | 0 | 0 | 0 | 0 | **0.14** |
| | FL818 | 0 | 0 | 0 | 0 | 0 | 0 | 0 | 0 | 0 | 0 | 0 | **0.21** | 0 | 0 | 0 | 0 | 0 | 0 | 0 | 0 | **0.10** | 0 | 0 | **0.01** |
| | FL823 | 0 | 0 | **0.25** | 0 | 0 | **0.09** | 0 | 0 | **0.05** | 0 | **0.18** | 0 | 0 | 0 | **0.09** | 0 | 0 | **0.01** | 0 | 0 | 0 | 0 | **0.01** | 0 |
| Clementi | FL640 | 0 | 0 | 0 | 0 | 0 | 0 | 0 | 0 | **0.17** | 0 | 0 | 0 | 0 | 0 | 0 | 0 | **0.04** | 0 | 0 | 0 | **0.01** | 0 | 0 | 0 |
| Geylang | FL19 | 0 | 0 | 0 | 0 | 0 | 0 | 0 | 0 | 0 | 0 | 0 | 0 | 0 | 0 | 0 | 0 | 0 | 0 | 0 | 0 | 0 | 0 | **0.15** | 0 |
| | FL200 | **0.48** | **0.10** | 0 | 0 | 0 | 0 | 0 | 0 | 0 | 0 | 0 | 0 | **0.42** | **0.10** | 0 | 0 | 0 | 0 | 0 | 0 | 0 | 0 | 0 | 0 |
| | FL204 | 0 | 0 | 0 | 0 | 0 | 0 | 0 | 0 | 0 | 0 | 0 | 0 | 0 | 0 | 0 | 0 | 0 | **0.11** | 0 | 0 | 0 | 0 | **0.05** | **0.07** |
| | FL81 | 0 | 0 | 0 | 0 | 0 | 0 | 0 | 0 | 0 | 0 | 0 | 0 | 0 | 0 | **0.02** | 0 | 0 | 0 | 0 | 0 | 0 | 0 | 0 | 0 |
| Hougang | FL367 | 0 | 0 | 0 | 0 | 0 | 0 | 0 | 0 | 0 | 0 | 0 | 0 | 0 | 0 | 0 | 0 | 0 | **0.01** | 0 | 0 | 0 | 0 | 0 | 0 |
| | FL375 | 0 | **0.19** | 0 | 0 | 0 | 0 | 0 | 0 | 0 | 0 | 0 | 0 | 0 | **0.15** | 0 | 0 | 0 | 0 | 0 | 0 | 0 | 0 | 0 | 0 |
| | FL451 | **0.12** | **0.17** | 0 | **0.26** | 0 | 0 | **0.17** | 0 | 0 | 0 | 0 | 0 | **0.10** | **0.13** | 0 | **0.02** | 0 | 0 | **0.01** | 0 | 0 | 0 | 0 | 0 |
| | FL482 | 0 | 0 | 0 | 0 | 0 | 0 | 0 | 0 | 0 | 0 | 0 | 0 | 0 | 0 | 0 | 0 | 0 | **0.02** | 0 | 0 | 0 | 0 | 0 | 0 |
| | FL489 | 0 | 0 | 0 | 0 | 0 | 0 | 0 | 0 | 0 | 0 | 0 | 0 | 0 | 0 | 0 | 0 | 0 | **0.01** | 0 | 0 | 0 | 0 | 0 | 0 |
| Jurong East | FL697 | 0 | **0.25** | 0 | 0 | **0.36** | 0 | 0 | 0 | 0 | 0 | 0 | 0 | 0 | **0.16** | 0 | 0 | **0.22** | 0 | 0 | 0 | 0 | 0 | 0 | 0 |
| | FL701 | 0 | 0 | 0 | 0 | 0 | 0 | 0 | 0 | 0 | 0 | 0 | 0 | **0.02** | **0.14** | 0 | 0 | 0 | 0 | 0 | 0 | 0 | 0 | 0 | 0 |
| | FL724 | 0 | 0 | 0 | 0 | 0 | 0 | 0 | 0 | 0 | 0 | 0 | 0 | 0 | 0 | 0 | 0 | 0 | **0.01** | 0 | 0 | 0 | 0 | 0 | 0 |
| | FL725 | 0 | 0 | **0.05** | 0 | 0 | 0 | 0 | 0 | 0 | 0 | 0 | 0 | 0 | 0 | **0.01** | 0 | 0 | **0.01** | 0 | 0 | 0 | 0 | **0.04** | 0 |
| | FL752 | 0 | 0 | 0 | 0 | 0 | 0 | 0 | 0 | 0 | 0 | 0 | 0 | 0 | 0 | 0 | 0 | 0 | 0 | 0 | 0 | 0 | 0 | **0.05** | 0 |
| Jurong West | FL430 | 0 | 0 | 0 | 0 | 0 | 0 | 0 | 0 | **0.02** | 0 | 0 | 0 | 0 | 0 | 0 | 0 | 0 | 0 | 0 | 0 | 0 | 0 | 0 | **0.01** |
| | FL441 | 0 | 0 | 0 | 0 | 0 | 0 | 0 | 0 | 0 | 0 | 0 | 0 | **0.03** | 0 | 0 | 0 | 0 | 0 | 0 | 0 | 0 | 0 | 0 | 0 |

Continued on next page

**Table 14 – continued from previous page**

| Donor | | Model 3 | | | | | | | | | | | | Model 4 | | | | | | | | | | | |
|---|---|---|---|---|---|---|---|---|---|---|---|---|---|---|---|---|---|---|---|---|---|---|---|---|---|
| Town | Sector | FL278 | FL283 | FL285 | FL290 | FL291 | FL295 | FL296 | FL297 | FL298 | FL420 | FL421 | FL517 | FL278 | FL283 | FL285 | FL290 | FL291 | FL295 | FL296 | FL297 | FL298 | FL420 | FL421 | FL517 |
| | FL442 | 0 | 0 | 0 | 0 | 0 | 0 | 0 | 0 | 0 | 0 | 0 | 0 | 0 | 0 | **0.03** | 0 | 0 | 0 | 0 | 0 | 0 | 0 | 0 | 0 |
| | FL621 | 0 | 0 | 0 | **0.25** | 0 | 0 | **0.11** | **0.34** | 0 | 0 | 0 | 0 | 0 | 0 | 0 | 0 | **0.07** | 0 | **0.06** | **0.34** | 0 | 0 | 0 | 0 |
| | FL652 | 0 | 0 | 0 | 0 | 0 | 0 | 0 | 0 | 0 | 0 | 0 | 0 | **0.02** | **0.04** | 0 | 0 | 0 | 0 | 0 | 0 | 0 | 0 | 0 | 0 |
| | FL662 | 0 | 0 | 0 | 0 | 0 | 0 | 0 | 0 | 0 | 0 | 0 | 0 | 0 | 0 | 0 | 0 | 0 | 0 | 0 | 0 | **0.03** | 0 | 0 | 0 |
| | FL663 | 0 | 0 | 0 | 0 | 0 | 0 | 0 | 0 | **0.11** | 0 | 0 | 0 | 0 | 0 | 0 | 0 | 0 | 0 | 0 | 0 | **0.03** | 0 | 0 | 0 |
| | FL710 | 0 | 0 | 0 | 0 | 0 | 0 | 0 | 0 | 0 | 0 | 0 | 0 | 0 | 0 | 0 | 0 | 0 | 0 | 0 | 0 | 0 | **0.08** | 0 | 0 |
| Kallang | FL14 | 0 | 0 | 0 | 0 | 0 | 0 | 0 | 0 | **0.11** | 0 | 0 | 0 | 0 | 0 | 0 | 0 | 0 | 0 | 0 | 0 | **0.02** | 0 | 0 | 0 |
| | FL16 | 0 | 0 | 0 | 0 | 0 | 0 | 0 | 0 | 0 | 0 | 0 | 0 | 0 | 0 | **0.07** | 0 | 0 | 0 | 0 | 0 | 0 | 0 | 0 | 0 |
| | FL181 | 0 | 0 | 0 | 0 | 0 | 0 | 0 | 0 | 0 | 0 | 0 | 0 | 0 | 0 | **0.05** | 0 | 0 | 0 | 0 | 0 | **0.03** | 0 | 0 | 0 |
| | FL182 | 0 | 0 | 0 | 0 | 0 | 0 | 0 | 0 | 0 | 0 | 0 | 0 | 0 | 0 | **0.01** | 0 | 0 | 0 | 0 | 0 | 0 | 0 | 0 | 0 |
| | FL183 | 0 | 0 | 0 | 0 | 0 | 0 | 0 | 0 | 0 | 0 | 0 | 0 | 0 | 0 | 0 | 0 | 0 | 0 | 0 | 0 | 0 | 0 | 0 | **0.14** |
| | FL185 | 0 | 0 | 0 | 0 | **0.10** | 0 | **0.05** | 0 | 0 | 0 | 0 | 0 | 0 | 0 | 0 | 0 | **0.06** | 0 | 0 | 0 | **0.01** | 0 | 0 | 0 |
| | FL26 | 0 | 0 | 0 | 0 | 0 | 0 | 0 | 0 | 0 | 0 | 0 | 0 | 0 | 0 | 0 | 0 | **0.06** | 0 | 0 | 0 | **0.02** | 0 | 0 | **0.03** |
| | FL85 | 0 | 0 | 0 | 0 | 0 | 0 | 0 | 0 | 0 | 0 | 0 | 0 | 0 | 0 | 0 | 0 | 0 | 0 | **0.01** | 0 | 0 | 0 | 0 | 0 |
| Novena | FL68 | 0 | 0 | 0 | 0 | 0 | 0 | 0 | 0 | 0 | 0 | 0 | 0 | 0 | 0 | 0 | 0 | 0 | 0 | 0 | 0 | 0 | 0 | 0 | **0.13** |
| Pasir Ris | FL166 | 0 | 0 | 0 | 0 | 0 | 0 | **0.03** | 0 | 0 | 0 | 0 | 0 | 0 | 0 | 0 | 0 | 0 | 0 | 0 | 0 | **0.04** | 0 | 0 | 0 |
| | FL223 | 0 | 0 | 0 | 0 | 0 | 0 | 0 | 0 | 0 | 0 | **0.05** | 0 | 0 | 0 | 0 | 0 | 0 | 0 | 0 | 0 | 0 | 0 | 0 | 0 |
| | FL234 | 0 | 0 | 0 | 0 | 0 | **0.04** | 0 | 0 | 0 | 0 | **0.03** | 0 | 0 | 0 | 0 | 0 | 0 | 0 | 0 | 0 | 0 | 0 | 0 | 0 |
| | FL256 | 0 | 0 | 0 | 0 | 0 | 0 | 0 | 0 | 0 | 0 | 0 | 0 | 0 | 0 | 0 | 0 | 0 | **0.05** | 0 | 0 | **0.01** | 0 | **0.01** | 0 |
| | FL288 | 0 | 0 | 0 | 0 | 0 | 0 | 0 | 0 | 0 | 0 | 0 | 0 | 0 | 0 | 0 | **0.05** | 0 | 0 | 0 | 0 | 0 | 0 | 0 | 0 |
| | FL293 | 0 | 0 | 0 | 0 | 0 | 0 | 0 | 0 | 0 | 0 | 0 | 0 | 0 | 0 | 0 | 0 | 0 | 0 | 0 | 0 | 0 | **0.02** | 0 | 0 |
| | FL321 | 0 | 0 | **0.07** | **0.02** | 0 | 0 | 0 | 0 | 0 | 0 | 0 | 0 | 0 | 0 | 0 | 0 | 0 | 0 | 0 | 0 | 0 | 0 | 0 | 0 |
| | FL323 | 0 | 0 | 0 | 0 | 0 | 0 | 0 | 0 | 0 | 0 | 0 | 0 | 0 | 0 | 0 | 0 | 0 | 0 | **0.08** | 0 | 0 | 0 | 0 | 0 |
| | FL336 | 0 | 0 | 0 | 0 | 0 | 0 | 0 | 0 | 0 | 0 | 0 | 0 | 0 | 0 | **0.04** | 0 | 0 | 0 | 0 | 0 | 0 | 0 | **0.07** | 0 |
| | FL348 | 0 | 0 | 0 | 0 | 0 | 0 | 0 | 0 | 0 | 0 | 0 | 0 | 0 | 0 | 0 | **0.03** | 0 | 0 | 0 | 0 | 0 | 0 | 0 | 0 |
| | FL6 | 0 | 0 | 0 | 0 | 0 | 0 | 0 | 0 | 0 | 0 | **0.13** | 0 | 0 | 0 | 0 | 0 | 0 | 0 | 0 | 0 | 0 | 0 | **0.01** | 0 |
| Queenstown | FL108 | 0 | 0 | 0 | 0 | 0 | 0 | 0 | 0 | 0 | 0 | 0 | 0 | 0 | 0 | 0 | 0 | 0 | 0 | 0 | 0 | 0 | 0 | **0.09** | 0 |
| | FL31 | 0 | 0 | 0 | 0 | 0 | 0 | 0 | 0 | 0 | 0 | 0 | 0 | 0 | 0 | 0 | 0 | 0 | 0 | **0.05** | 0 | **0.10** | 0 | 0 | **0.04** |
| | FL352 | 0 | 0 | 0 | 0 | 0 | 0 | 0 | 0 | 0 | 0 | 0 | 0 | 0 | 0 | 0 | 0 | 0 | 0 | 0 | 0 | 0 | 0 | **0.02** | 0 |
| Rochor | FL76 | 0 | 0 | **0.06** | 0 | 0 | 0 | 0 | 0 | 0 | 0 | 0 | 0 | 0 | 0 | 0 | 0 | 0 | 0 | 0 | 0 | 0 | 0 | 0 | 0 |
| Sembawang | FL527 | 0 | 0 | **0.06** | 0 | 0 | 0 | 0 | 0 | 0 | 0 | 0 | 0 | 0 | 0 | 0 | 0 | 0 | 0 | 0 | 0 | 0 | 0 | 0 | 0 |
| Sengkang | FL136 | 0 | 0 | 0 | 0 | 0 | 0 | 0 | 0 | 0 | 0 | 0 | 0 | 0 | 0 | 0 | **0.02** | 0 | 0 | 0 | 0 | **0.09** | 0 | 0 | 0 |
| | FL368 | 0 | 0 | 0 | 0 | 0 | 0 | 0 | 0 | 0 | 0 | 0 | 0 | 0 | 0 | 0 | 0 | 0 | **0.05** | 0 | 0 | 0 | 0 | 0 | 0 |
| | FL492 | 0 | 0 | 0 | 0 | 0 | 0 | 0 | 0 | 0 | 0 | 0 | 0 | 0 | 0 | 0 | 0 | 0 | 0 | **0.02** | 0 | 0 | 0 | 0 | 0 |
| | FL497 | 0 | **0.11** | 0 | 0 | **0.30** | 0 | 0 | **0.13** | 0 | 0 | 0 | 0 | **0.03** | **0.06** | 0 | **0.07** | **0.13** | 0 | 0 | **0.13** | 0 | 0 | 0 | 0 |
| | FL537 | 0 | 0 | 0 | 0 | 0 | 0 | 0 | 0 | 0 | 0 | 0 | 0 | 0 | 0 | 0 | 0 | 0 | 0 | 0 | 0 | 0 | **0.05** | 0 | 0 |
| | FL538 | 0 | 0 | 0 | 0 | 0 | 0 | 0 | 0 | 0 | 0 | 0 | 0 | 0 | 0 | 0 | **0.05** | 0 | 0 | 0 | 0 | 0 | 0 | 0 | 0 |
| Serangoon | FL392 | 0 | 0 | 0 | 0 | 0 | 0 | 0 | 0 | 0 | 0 | 0 | 0 | 0 | 0 | 0 | 0 | 0 | 0 | 0 | 0 | 0 | 0 | **0.02** | 0 |
| | FL393 | 0 | 0 | 0 | 0 | 0 | 0 | 0 | 0 | 0 | 0 | 0 | 0 | 0 | 0 | 0 | 0 | 0 | **0.02** | 0 | 0 | 0 | 0 | 0 | 0 |
| | FL406 | 0 | 0 | **0.21** | 0 | 0 | 0 | 0 | 0 | 0 | **0.14** | 0 | 0 | 0 | 0 | 0 | 0 | 0 | 0 | 0 | 0 | 0 | **0.01** | 0 | 0 |
| | FL460 | 0 | 0 | 0 | 0 | 0 | 0 | 0 | 0 | 0 | 0 | 0 | 0 | 0 | 0 | 0 | 0 | 0 | **0.12** | 0 | 0 | 0 | 0 | 0 | 0 |
| | FL461 | 0 | 0 | 0 | 0 | 0 | 0 | 0 | 0 | 0 | 0 | 0 | 0 | 0 | 0 | 0 | 0 | 0 | 0 | 0 | 0 | **0.04** | 0 | **0.14** | 0 |
| | FL480 | 0 | 0 | 0 | 0 | 0 | 0 | 0 | 0 | 0 | 0 | 0 | 0 | 0 | 0 | 0 | 0 | 0 | **0.07** | 0 | 0 | 0 | 0 | 0 | 0 |
| | FL501 | 0 | 0 | 0 | 0 | 0 | 0 | 0 | 0 | 0 | 0 | 0 | 0 | 0 | 0 | 0 | 0 | 0 | **0.02** | 0 | 0 | 0 | 0 | 0 | 0 |
| Toa Payoh | FL164 | 0 | 0 | 0 | 0 | 0 | 0 | 0 | **0.05** | 0 | 0 | 0 | 0 | 0 | 0 | 0 | 0 | 0 | 0 | **0.15** | **0.05** | 0 | 0 | 0 | 0 |
| | FL359 | 0 | 0 | 0 | 0 | 0 | 0 | 0 | 0 | 0 | 0 | 0 | 0 | 0 | 0 | 0 | 0 | 0 | 0 | 0 | 0 | **0.10** | 0 | 0 | 0 |
| | FL38 | 0 | 0 | 0 | 0 | 0 | 0 | 0 | 0 | 0 | 0 | 0 | 0 | 0 | 0 | 0 | 0 | 0 | 0 | 0 | 0 | **0.05** | 0 | 0 | 0 |
| | FL58 | 0 | 0 | 0 | 0 | 0 | 0 | 0 | 0 | 0 | 0 | 0 | 0 | 0 | 0 | 0 | 0 | 0 | 0 | **0.02** | 0 | 0 | 0 | 0 | 0 |
| | FL59 | **0.11** | **0.17** | 0 | 0 | 0 | 0 | 0 | 0 | 0 | 0 | 0 | 0 | **0.13** | **0.15** | 0 | 0 | 0 | **0.03** | 0 | 0 | **0.01** | 0 | 0 | **0.10** |
| | FL61 | 0 | 0 | 0 | 0 | **0.15** | 0 | **0.15** | 0 | 0 | 0 | 0 | 0 | 0 | 0 | 0 | **0.01** | **0.11** | 0 | **0.12** | 0 | **0.02** | 0 | 0 | **0.04** |
| | FL65 | 0 | 0 | 0 | 0 | 0 | 0 | 0 | 0 | 0 | 0 | 0 | 0 | 0 | 0 | **0.14** | 0 | 0 | **0.15** | 0 | 0 | 0 | **0.16** | **0.02** | 0 |
| Woodlands | FL245 | 0 | 0 | 0 | 0 | 0 | **0.23** | 0 | 0 | 0 | **0.24** | 0 | 0 | 0 | 0 | 0 | 0 | 0 | **0.06** | 0 | 0 | 0 | 0 | 0 | 0 |
| | FL261 | 0 | 0 | 0 | 0 | 0 | 0 | 0 | 0 | 0 | 0 | 0 | 0 | 0 | 0 | 0 | 0 | 0 | 0 | 0 | 0 | 0 | **0.08** | 0 | 0 |
| | FL417 | 0 | 0 | 0 | 0 | 0 | 0 | 0 | 0 | 0 | 0 | 0 | 0 | 0 | 0 | **0.05** | 0 | 0 | 0 | 0 | 0 | 0 | 0 | 0 | 0 |
| | FL514 | 0 | 0 | 0 | 0 | 0 | 0 | 0 | 0 | 0 | 0 | 0 | 0 | 0 | 0 | **0.01** | 0 | 0 | 0 | 0 | 0 | 0 | 0 | 0 | 0 |
| | FL518 | 0 | 0 | 0 | 0 | 0 | 0 | 0 | 0 | 0 | 0 | 0 | 0 | 0 | 0 | **0.04** | 0 | 0 | 0 | 0 | 0 | 0 | 0 | **0.06** | 0 |
| | FL572 | 0 | 0 | 0 | 0 | 0 | 0 | 0 | 0 | 0 | 0 | 0 | 0 | 0 | 0 | 0 | 0 | 0 | 0 | 0 | 0 | **0.03** | 0 | 0 | 0 |
| | FL574 | 0 | 0 | 0 | 0 | 0 | 0 | 0 | 0 | 0 | 0 | 0 | 0 | 0 | 0 | 0 | 0 | 0 | 0 | **0.05** | 0 | 0 | 0 | 0 | 0 |
| | FL580 | 0 | 0 | 0 | 0 | 0 | 0 | 0 | 0 | 0 | 0 | 0 | 0 | 0 | 0 | 0 | 0 | **0.03** | 0 | 0 | 0 | 0 | 0 | 0 | 0 |
| | FL581 | 0 | 0 | 0 | 0 | 0 | 0 | 0 | 0 | 0 | 0 | 0 | 0 | 0 | 0 | 0 | 0 | 0 | 0 | 0 | 0 | 0 | **0.09** | 0 | 0 |
| | FL596 | 0 | 0 | **0.02** | 0 | 0 | 0 | 0 | 0 | 0 | 0 | 0 | 0 | 0 | 0 | 0 | 0 | 0 | 0 | 0 | 0 | 0 | **0.07** | 0 | 0 |
| | FL598 | 0 | 0 | 0 | 0 | 0 | 0 | **0.13** | 0 | 0 | 0 | 0 | 0 | 0 | 0 | 0 | 0 | 0 | 0 | 0 | 0 | **0.03** | 0 | 0 | 0 |
| | FL599 | 0 | 0 | 0 | 0 | 0 | 0 | 0 | 0 | 0 | 0 | 0 | 0 | 0 | 0 | **0.12** | 0 | 0 | 0 | 0 | 0 | 0 | 0 | 0 | 0 |
| | FL600 | 0 | 0 | **0.10** | 0 | 0 | 0 | 0 | 0 | 0 | 0 | 0 | 0 | 0 | 0 | **0.07** | 0 | 0 | 0 | 0 | 0 | 0 | 0 | **0.05** | 0 |
| | FL601 | 0 | 0 | 0 | 0 | 0 | **0.01** | 0 | 0 | 0 | 0 | 0 | 0 | 0 | 0 | 0 | 0 | 0 | 0 | 0 | 0 | 0 | 0 | 0 | 0 |
| | FL607 | 0 | 0 | 0 | 0 | 0 | 0 | 0 | 0 | 0 | **0.19** | 0 | 0 | 0 | 0 | 0 | 0 | 0 | 0 | 0 | 0 | 0 | 0 | 0 | 0 |
| | FL619 | 0 | 0 | 0 | 0 | 0 | 0 | 0 | 0 | 0 | 0 | 0 | 0 | 0 | 0 | 0 | **0.02** | 0 | **0.02** | 0 | 0 | 0 | 0 | 0 | 0 |
| | FL830 | 0 | 0 | 0 | 0 | 0 | 0 | 0 | 0 | 0 | 0 | 0 | 0 | 0 | 0 | 0 | 0 | 0 | 0 | 0 | 0 | 0 | **0.03** | 0 | 0 |

Table 15: Donor weights for Yishun sectors for models 3 and 4. Only sectors with any contribution to synthetic control weights for intervention sectors' synthetic controls are shown

| Donor | | Model 3 | | | | | | | | | | | Model 4 | | | | | | | | | | |
|---|---|---|---|---|---|---|---|---|---|---|---|---|---|---|---|---|---|---|---|---|---|---|---|
| Town | Sector | FL523 | FL528 | FL540 | FL557 | FL558 | FL564 | FL565 | FL568 | FL569 | FL573 | FL576 | FL523 | FL528 | FL540 | FL557 | FL558 | FL564 | FL565 | FL568 | FL569 | FL573 | FL576 |
| Ang Mo Kio | FL18 | 0 | 0 | 0 | 0 | **0.16** | 0 | 0 | 0 | 0 | 0 | 0 | 0 | 0 | 0 | 0 | **0.03** | 0 | 0 | 0 | 0 | 0 | 0 |
| | FL360 | 0 | **0.09** | 0 | 0 | 0 | 0 | 0 | 0 | 0 | 0 | 0 | 0 | 0 | 0 | 0 | 0 | **0.04** | 0 | 0 | 0 | 0 | **0.02** |
| | FL362 | 0 | **0.11** | 0 | 0 | 0 | 0 | 0 | 0 | 0 | 0 | 0 | 0 | **0.14** | 0 | 0 | 0 | 0 | 0 | 0 | 0 | 0 | 0 |
| | FL372 | 0 | 0 | 0 | 0 | 0 | 0 | 0 | 0 | 0 | 0 | 0 | 0 | 0 | 0 | 0 | 0 | 0 | 0 | 0 | 0 | 0 | **0.04** |
| | FL386 | 0 | 0 | 0 | 0 | 0 | 0 | 0 | 0 | 0 | 0 | **0.07** | 0 | 0 | 0 | 0 | 0 | 0 | 0 | 0 | 0 | 0 | 0 |
| | FL454 | 0 | 0 | 0 | 0 | 0 | 0 | 0 | **0.10** | 0 | 0 | 0 | 0 | 0 | 0 | 0 | 0 | 0 | 0 | 0 | 0 | 0 | 0 |
| | FL455 | 0 | 0 | **0.08** | 0 | 0 | **0.30** | 0 | 0 | 0 | 0 | 0 | 0 | 0 | 0 | 0 | 0 | 0 | 0 | 0 | 0 | 0 | 0 |
| | FL456 | 0 | **0.32** | 0 | 0 | 0 | 0 | 0 | 0 | 0 | 0 | 0 | 0 | **0.16** | 0 | 0 | 0 | 0 | 0 | 0 | 0 | 0 | 0 |
| | FL471 | 0 | 0 | 0 | 0 | 0 | **0.01** | 0 | 0 | 0 | 0 | 0 | 0 | 0 | 0 | 0 | 0 | 0 | 0 | 0 | 0 | 0 | 0 |
| | FL472 | 0 | 0 | 0 | 0 | 0 | 0 | 0 | 0 | 0 | 0 | 0 | 0 | **0.01** | 0 | 0 | **0.03** | **0.04** | 0 | 0 | 0 | 0 | 0 |
| | FL477 | 0 | **0.06** | 0 | 0 | 0 | 0 | 0 | 0 | 0 | 0 | 0 | 0 | 0 | 0 | 0 | 0 | **0.07** | 0 | 0 | 0 | 0 | 0 |
| | FL478 | 0 | 0 | 0 | 0 | 0 | 0 | 0 | 0 | 0 | 0 | 0 | 0 | 0 | 0 | 0 | **0.03** | 0 | 0 | 0 | 0 | 0 | 0 |
| | FL487 | 0 | 0 | 0 | 0 | 0 | 0 | 0 | 0 | 0 | 0 | 0 | 0 | **0.03** | 0 | 0 | 0 | 0 | 0 | 0 | 0 | 0 | 0 |
| | FL500 | 0 | 0 | 0 | **0.06** | 0 | 0 | 0 | 0 | 0 | 0 | **0.27** | 0 | 0 | 0 | 0 | 0 | 0 | 0 | 0 | 0 | 0 | **0.21** |
| Bedok | FL131 | 0 | 0 | 0 | 0 | 0 | 0 | 0 | 0 | 0 | 0 | 0 | 0 | 0 | 0 | **0.03** | 0 | 0 | 0 | 0 | 0 | 0 | 0 |
| | FL230 | 0 | 0 | 0 | 0 | 0 | 0 | 0 | 0 | 0 | 0 | 0 | 0 | 0 | 0 | 0 | **0.03** | 0 | 0 | 0 | 0 | 0 | 0 |
| | FL237 | 0 | 0 | 0 | 0 | 0 | 0 | 0 | 0 | 0 | 0 | 0 | 0 | 0 | 0 | **0.01** | **0.03** | 0 | 0 | 0 | 0 | 0 | **0.02** |
| | FL240 | 0 | 0 | 0 | 0 | 0 | 0 | 0 | 0 | 0 | 0 | 0 | 0 | 0 | 0 | 0 | 0 | **0.03** | 0 | 0 | 0 | 0 | 0 |
| | FL259 | 0 | 0 | 0 | 0 | 0 | 0 | 0 | 0 | 0 | 0 | 0 | 0 | 0 | 0 | **0.04** | 0 | **0.11** | 0 | 0 | 0 | 0 | 0 |
| Bishan | FL36 | 0 | **0.12** | 0 | 0 | 0 | **0.13** | 0 | 0 | 0 | 0 | 0 | 0 | **0.04** | 0 | 0 | 0 | **0.08** | 0 | 0 | 0 | 0 | 0 |
| | FL47 | **0.08** | 0 | 0 | **0.09** | 0 | 0 | **0.27** | 0 | 0 | 0 | 0 | **0.18** | 0 | **0.28** | **0.03** | 0 | 0 | **0.17** | 0 | 0 | 0 | 0 |
| Bukit Batok | FL434 | 0 | 0 | 0 | 0 | 0 | 0 | 0 | 0 | 0 | 0 | 0 | 0 | 0 | 0 | **0.01** | 0 | **0.02** | 0 | 0 | 0 | 0 | 0 |
| | FL435 | 0 | 0 | 0 | 0 | 0 | **0.06** | 0 | 0 | 0 | 0 | 0 | 0 | 0 | 0 | 0 | 0 | 0 | 0 | 0 | 0 | 0 | 0 |
| | FL699 | 0 | 0 | 0 | 0 | 0 | 0 | 0 | 0 | 0 | 0 | **0.09** | 0 | 0 | 0 | 0 | 0 | 0 | 0 | 0 | 0 | 0 | **0.10** |
| | FL731 | 0 | 0 | 0 | **0.31** | 0 | 0 | 0 | 0 | 0 | 0 | 0 | 0 | 0 | 0 | **0.17** | 0 | 0 | 0 | 0 | 0 | 0 | 0 |
| Bukit Merah | FL161 | 0 | 0 | **0.21** | 0 | 0 | 0 | 0 | 0 | 0 | 0 | 0 | 0 | 0 | 0 | 0 | 0 | **0.08** | 0 | 0 | 0 | 0 | 0 |
| | FL193 | **0.10** | 0 | 0 | 0 | 0 | 0 | **0.08** | 0 | **0.11** | **0.06** | 0 | **0.04** | 0 | **0.23** | 0 | 0 | 0 | **0.09** | **0.10** | **0.11** | **0.01** | 0 |
| Bukit Panjang | FL647 | 0 | 0 | 0 | 0 | 0 | 0 | 0 | 0 | 0 | 0 | 0 | 0 | 0 | 0 | 0 | 0 | 0 | 0 | 0 | 0 | 0 | **0.01** |
| | FL648 | 0 | 0 | 0 | 0 | 0 | 0 | 0 | **0.32** | 0 | **0.17** | 0 | 0 | 0 | 0 | 0 | 0 | 0 | 0 | 0 | 0 | **0.06** | 0 |
| | FL659 | 0 | 0 | 0 | **0.13** | **0.22** | 0 | 0 | 0 | 0 | 0 | 0 | 0 | 0 | 0 | **0.05** | **0.05** | **0.11** | 0 | 0 | 0 | 0 | 0 |
| Choa Chu Kang | FL729 | 0 | 0 | 0 | 0 | **0.09** | 0 | 0 | 0 | 0 | 0 | 0 | 0 | 0 | 0 | 0 | 0 | 0 | 0 | 0 | 0 | 0 | 0 |
| | FL811 | 0 | 0 | 0 | 0 | 0 | 0 | 0 | 0 | 0 | 0 | 0 | 0 | 0 | 0 | 0 | 0 | **0.04** | 0 | 0 | 0 | **0.11** | 0 |
| | FL818 | 0 | 0 | **0.01** | 0 | 0 | 0 | 0 | 0 | 0 | **0.32** | **0.19** | 0 | 0 | 0 | 0 | 0 | 0 | 0 | 0 | 0 | **0.17** | 0 |
| Clementi | FL670 | 0 | 0 | 0 | 0 | 0 | 0 | 0 | 0 | 0 | 0 | 0 | 0 | **0.04** | 0 | 0 | 0 | 0 | 0 | 0 | 0 | 0 | 0 |
| | FL671 | 0 | 0 | 0 | 0 | 0 | 0 | 0 | 0 | 0 | 0 | 0 | 0 | 0 | 0 | 0 | 0 | **0.01** | 0 | 0 | 0 | 0 | 0 |
| Geylang | FL200 | **0.28** | 0 | **0.01** | **0.11** | 0 | 0 | 0 | 0 | **0.35** | 0 | 0 | **0.23** | 0 | 0 | **0.10** | 0 | 0 | 0 | **0.12** | **0.29** | 0 | 0 |
| Hougang | FL367 | 0 | 0 | 0 | 0 | 0 | 0 | 0 | 0 | 0 | 0 | 0 | 0 | **0.09** | 0 | 0 | 0 | 0 | 0 | 0 | 0 | **0.05** | **0.11** |
| | FL375 | **0.11** | 0 | 0 | 0 | 0 | 0 | 0 | 0 | **0.05** | 0 | 0 | 0 | 0 | 0 | 0 | 0 | 0 | **0.10** | **0.15** | **0.08** | 0 | 0 |
| | FL451 | 0 | 0 | **0.52** | 0 | 0 | 0 | **0.23** | 0 | **0.34** | 0 | 0 | **0.06** | 0 | **0.22** | 0 | 0 | 0 | **0.13** | **0.11** | **0.26** | 0 | **0.02** |
| | FL452 | 0 | 0 | 0 | 0 | 0 | 0 | 0 | 0 | 0 | 0 | 0 | 0 | 0 | 0 | 0 | 0 | **0.02** | 0 | 0 | 0 | 0 | 0 |
| | FL465 | 0 | 0 | 0 | 0 | 0 | 0 | 0 | 0 | 0 | 0 | 0 | 0 | **0.01** | 0 | 0 | 0 | 0 | 0 | 0 | 0 | 0 | 0 |
| | FL483 | 0 | 0 | 0 | 0 | 0 | 0 | 0 | 0 | 0 | 0 | 0 | 0 | 0 | 0 | 0 | **0.03** | 0 | 0 | 0 | 0 | 0 | 0 |
| | FL490 | 0 | 0 | 0 | 0 | 0 | 0 | 0 | 0 | 0 | 0 | 0 | 0 | 0 | 0 | 0 | **0.03** | 0 | 0 | 0 | 0 | 0 | 0 |
| Jurong East | FL668 | 0 | 0 | 0 | 0 | 0 | 0 | 0 | 0 | 0 | 0 | 0 | 0 | **0.04** | 0 | 0 | 0 | 0 | 0 | 0 | 0 | 0 | 0 |
| | FL669 | 0 | 0 | 0 | 0 | 0 | 0 | 0 | 0 | 0 | 0 | 0 | 0 | 0 | 0 | 0 | **0.07** | 0 | 0 | 0 | 0 | 0 | 0 |
| | FL701 | **0.16** | 0 | 0 | 0 | 0 | 0 | 0 | 0 | 0 | 0 | 0 | **0.10** | 0 | **0.04** | 0 | 0 | 0 | 0 | 0 | 0 | 0 | 0 |
| | FL723 | 0 | 0 | 0 | 0 | 0 | 0 | 0 | 0 | 0 | 0 | 0 | 0 | 0 | 0 | 0 | **0.07** | 0 | 0 | 0 | 0 | 0 | 0 |
| | FL752 | 0 | 0 | 0 | 0 | 0 | 0 | 0 | 0 | 0 | 0 | 0 | 0 | 0 | 0 | 0 | 0 | **0.01** | 0 | 0 | 0 | 0 | 0 |
| Jurong West | FL430 | 0 | 0 | 0 | 0 | 0 | 0 | 0 | 0 | 0 | 0 | 0 | 0 | 0 | 0 | 0 | **0.01** | 0 | 0 | 0 | 0 | 0 | 0 |
| | FL431 | 0 | 0 | 0 | 0 | 0 | 0 | 0 | 0 | 0 | 0 | **0.19** | 0 | 0 | 0 | **0.05** | 0 | 0 | 0 | 0 | 0 | 0 | **0.09** |
| | FL441 | 0 | 0 | 0 | 0 | 0 | 0 | 0 | 0 | 0 | 0 | 0 | **0.03** | 0 | 0 | 0 | 0 | 0 | **0.07** | 0 | 0 | 0 | 0 |
| | FL442 | 0 | 0 | 0 | 0 | 0 | 0 | 0 | 0 | 0 | 0 | 0 | 0 | **0.04** | 0 | 0 | 0 | 0 | 0 | 0 | 0 | 0 | 0 |
| | FL643 | 0 | 0 | 0 | 0 | 0 | 0 | 0 | 0 | 0 | 0 | 0 | 0 | **0.05** | 0 | 0 | 0 | 0 | 0 | 0 | 0 | 0 | 0 |
| | FL652 | **0.09** | 0 | 0 | 0 | 0 | 0 | **0.17** | 0 | 0 | 0 | 0 | **0.07** | 0 | **0.11** | 0 | **0.02** | 0 | **0.17** | 0 | **0.06** | **0.03** | **0.02** |
| | FL662 | 0 | 0 | 0 | 0 | 0 | 0 | 0 | 0 | 0 | 0 | 0 | 0 | 0 | 0 | 0 | 0 | **0.05** | 0 | 0 | 0 | **0.20** | 0 |
| | FL709 | 0 | 0 | 0 | 0 | 0 | 0 | 0 | 0 | 0 | 0 | **0.05** | 0 | 0 | 0 | 0 | 0 | 0 | 0 | 0 | 0 | 0 | 0 |
| | FL710 | 0 | 0 | 0 | 0 | 0 | 0 | 0 | 0 | 0 | 0 | 0 | 0 | 0 | 0 | 0 | **0.02** | 0 | 0 | 0 | 0 | 0 | 0 |
| | FL769 | 0 | 0 | 0 | **0.22** | 0 | 0 | 0 | 0 | 0 | 0 | 0 | 0 | 0 | 0 | 0 | 0 | 0 | 0 | 0 | 0 | 0 | 0 |
| Kallang | FL13 | 0 | 0 | 0 | 0 | 0 | 0 | 0 | 0 | 0 | 0 | 0 | 0 | 0 | 0 | 0 | **0.04** | 0 | 0 | 0 | 0 | 0 | 0 |
| | FL14 | 0 | 0 | 0 | 0 | 0 | 0 | 0 | 0 | 0 | 0 | 0 | 0 | **0.05** | 0 | 0 | 0 | 0 | 0 | 0 | 0 | 0 | 0 |
| | FL181 | 0 | **0.17** | 0 | 0 | 0 | 0 | 0 | 0 | 0 | 0 | 0 | 0 | **0.01** | 0 | 0 | 0 | 0 | 0 | 0 | 0 | 0 | 0 |
| | FL185 | 0 | 0 | 0 | 0 | 0 | 0 | 0 | 0 | 0 | 0 | 0 | 0 | 0 | 0 | 0 | 0 | 0 | **0.02** | 0 | 0 | 0 | 0 |
| | FL85 | 0 | 0 | 0 | 0 | 0 | **0.06** | 0 | 0 | 0 | 0 | 0 | 0 | 0 | 0 | 0 | 0 | 0 | 0 | 0 | 0 | 0 | 0 |
| | FL88 | 0 | 0 | 0 | 0 | 0 | 0 | 0 | 0 | 0 | 0 | 0 | 0 | 0 | 0 | 0 | 0 | 0 | 0 | 0 | 0 | 0 | **0.01** |
| Marine Parade | FL311 | 0 | 0 | 0 | 0 | 0 | 0 | 0 | 0 | 0 | 0 | 0 | 0 | 0 | 0 | 0 | 0 | 0 | 0 | 0 | 0 | 0 | **0.01** |
| Novena | FL68 | 0 | 0 | 0 | 0 | 0 | 0 | 0 | 0 | 0 | 0 | 0 | 0 | 0 | 0 | **0.01** | **0.02** | 0 | 0 | 0 | 0 | **0.01** | 0 |
| Pasir Ris | FL217 | 0 | 0 | 0 | 0 | 0 | 0 | 0 | 0 | 0 | 0 | 0 | 0 | 0 | 0 | 0 | 0 | 0 | 0 | 0 | 0 | 0 | **0.06** |

Continued on next page

**Table 15 – continued from previous page**

| Donor | | Model 3 | | | | | | | | | | | Model 4 | | | | | | | | | | |
|---|---|---|---|---|---|---|---|---|---|---|---|---|---|---|---|---|---|---|---|---|---|---|---|
| Town | Sector | FL523 | FL528 | FL540 | FL557 | FL558 | FL564 | FL565 | FL568 | FL569 | FL573 | FL576 | FL523 | FL528 | FL540 | FL557 | FL558 | FL564 | FL565 | FL568 | FL569 | FL573 | FL576 |
| | FL228 | 0 | 0 | 0 | 0 | 0 | **0.11** | 0 | 0 | 0 | 0 | **0.14** | 0 | 0 | 0 | 0 | 0 | 0 | 0 | 0 | 0 | 0 | 0 |
| | FL229 | 0 | **0.01** | 0 | 0 | 0 | **0.25** | 0 | 0 | 0 | 0 | 0 | 0 | 0 | 0 | 0 | 0 | **0.02** | 0 | 0 | 0 | 0 | 0 |
| | FL255 | 0 | 0 | 0 | 0 | 0 | 0 | 0 | 0 | 0 | 0 | 0 | 0 | **0.01** | 0 | 0 | 0 | **0.03** | 0 | 0 | 0 | 0 | 0 |
| | FL256 | 0 | 0 | 0 | 0 | 0 | 0 | 0 | 0 | 0 | 0 | 0 | 0 | **0.01** | 0 | 0 | 0 | 0 | 0 | 0 | 0 | 0 | 0 |
| | FL272 | 0 | 0 | 0 | 0 | **0.14** | 0 | 0 | 0 | 0 | 0 | 0 | 0 | 0 | 0 | 0 | 0 | 0 | 0 | 0 | 0 | 0 | 0 |
| | FL321 | 0 | 0 | 0 | 0 | 0 | 0 | 0 | 0 | 0 | 0 | 0 | 0 | 0 | 0 | 0 | **0.01** | 0 | 0 | 0 | 0 | 0 | 0 |
| | FL329 | 0 | 0 | 0 | 0 | 0 | 0 | 0 | 0 | 0 | 0 | 0 | 0 | 0 | 0 | 0 | **0.01** | 0 | 0 | 0 | 0 | 0 | 0 |
| | FL336 | 0 | 0 | 0 | 0 | 0 | 0 | 0 | 0 | 0 | 0 | 0 | 0 | 0 | 0 | 0 | **0.08** | 0 | 0 | 0 | 0 | 0 | 0 |
| Queenstown | FL114 | 0 | 0 | 0 | 0 | 0 | 0 | 0 | 0 | 0 | 0 | 0 | **0.10** | 0 | 0 | 0 | 0 | 0 | 0 | **0.40** | **0.09** | 0 | 0 |
| Rochor | FL73 | 0 | 0 | 0 | 0 | 0 | 0 | 0 | 0 | 0 | 0 | 0 | 0 | **0.05** | 0 | 0 | 0 | 0 | 0 | 0 | 0 | 0 | 0 |
| | FL76 | 0 | 0 | 0 | 0 | 0 | 0 | 0 | 0 | 0 | 0 | 0 | 0 | **0.01** | 0 | 0 | 0 | **0.06** | 0 | 0 | 0 | 0 | 0 |
| Sembawang | FL527 | 0 | 0 | **0.17** | 0 | 0 | **0.08** | 0 | **0.21** | 0 | 0 | 0 | 0 | 0 | 0 | 0 | 0 | 0 | 0 | 0 | 0 | 0 | 0 |
| Sengkang | FL369 | 0 | 0 | 0 | 0 | 0 | 0 | 0 | 0 | 0 | 0 | 0 | 0 | 0 | 0 | 0 | 0 | 0 | 0 | 0 | 0 | 0 | **0.01** |
| | FL492 | 0 | 0 | 0 | 0 | 0 | 0 | 0 | 0 | 0 | 0 | 0 | 0 | 0 | 0 | 0 | 0 | **0.04** | 0 | 0 | 0 | 0 | **0.01** |
| | FL497 | **0.19** | 0 | 0 | 0 | 0 | 0 | 0 | 0 | **0.14** | **0.18** | 0 | **0.13** | 0 | **0.10** | 0 | 0 | **0.02** | **0.02** | **0.12** | **0.11** | **0.10** | **0.02** |
| | FL537 | 0 | 0 | 0 | 0 | 0 | 0 | 0 | 0 | 0 | 0 | 0 | 0 | 0 | 0 | 0 | **0.06** | 0 | 0 | 0 | 0 | 0 | 0 |
| | FL538 | 0 | 0 | 0 | 0 | **0.10** | 0 | 0 | 0 | 0 | 0 | 0 | 0 | 0 | 0 | 0 | 0 | 0 | 0 | 0 | 0 | 0 | 0 |
| | FL732 | 0 | 0 | 0 | 0 | 0 | 0 | 0 | 0 | 0 | 0 | 0 | 0 | 0 | 0 | 0 | 0 | **0.01** | 0 | 0 | 0 | 0 | 0 |
| | FL733 | 0 | 0 | 0 | 0 | 0 | 0 | 0 | 0 | 0 | **0.02** | 0 | 0 | 0 | 0 | 0 | 0 | **0.02** | 0 | 0 | 0 | 0 | 0 |
| Serangoon | FL480 | 0 | **0.09** | 0 | 0 | 0 | 0 | 0 | 0 | 0 | 0 | 0 | 0 | **0.03** | 0 | 0 | 0 | 0 | 0 | 0 | 0 | 0 | 0 |
| Toa Payoh | FL164 | 0 | 0 | 0 | 0 | 0 | 0 | 0 | 0 | 0 | 0 | 0 | 0 | 0 | 0 | 0 | 0 | 0 | 0 | 0 | 0 | **0.18** | **0.14** |
| | FL319 | 0 | 0 | 0 | 0 | 0 | 0 | 0 | 0 | 0 | 0 | 0 | 0 | **0.07** | 0 | **0.28** | 0 | 0 | 0 | 0 | 0 | 0 | 0 |
| | FL359 | 0 | 0 | 0 | 0 | 0 | 0 | 0 | 0 | 0 | 0 | 0 | 0 | 0 | 0 | 0 | **0.05** | 0 | 0 | 0 | 0 | 0 | **0.06** |
| | FL42 | 0 | 0 | 0 | 0 | 0 | 0 | 0 | 0 | 0 | 0 | 0 | 0 | 0 | 0 | **0.12** | 0 | 0 | 0 | 0 | 0 | 0 | 0 |
| | FL59 | 0 | 0 | 0 | **0.08** | 0 | 0 | **0.25** | 0 | 0 | **0.06** | 0 | **0.07** | 0 | **0.02** | **0.07** | 0 | 0 | **0.23** | 0 | 0 | **0.08** | 0 |
| Woodlands | FL226 | 0 | 0 | 0 | 0 | **0.12** | 0 | 0 | 0 | 0 | 0 | 0 | 0 | 0 | 0 | 0 | 0 | 0 | 0 | 0 | 0 | 0 | 0 |
| | FL245 | 0 | 0 | 0 | 0 | **0.05** | 0 | 0 | 0 | 0 | 0 | 0 | 0 | 0 | 0 | 0 | 0 | 0 | 0 | 0 | 0 | 0 | 0 |
| | FL250 | 0 | 0 | 0 | 0 | 0 | 0 | 0 | 0 | 0 | 0 | 0 | 0 | 0 | 0 | 0 | 0 | **0.05** | 0 | 0 | 0 | 0 | 0 |
| | FL252 | 0 | 0 | 0 | 0 | 0 | 0 | 0 | 0 | 0 | 0 | 0 | 0 | 0 | 0 | 0 | 0 | 0 | 0 | 0 | 0 | 0 | **0.01** |
| | FL254 | 0 | 0 | 0 | 0 | 0 | 0 | 0 | 0 | 0 | 0 | 0 | 0 | **0.12** | 0 | 0 | 0 | 0 | 0 | 0 | 0 | 0 | 0 |
| | FL261 | 0 | 0 | 0 | 0 | 0 | 0 | 0 | 0 | 0 | 0 | 0 | 0 | 0 | 0 | 0 | **0.05** | 0 | 0 | 0 | 0 | 0 | 0 |
| | FL417 | 0 | 0 | 0 | 0 | 0 | 0 | 0 | 0 | 0 | 0 | 0 | 0 | 0 | 0 | **0.01** | 0 | 0 | 0 | 0 | 0 | 0 | 0 |
| | FL508 | 0 | 0 | 0 | 0 | 0 | 0 | 0 | 0 | 0 | 0 | 0 | 0 | 0 | 0 | 0 | 0 | **0.02** | 0 | 0 | 0 | 0 | 0 |
| | FL518 | 0 | 0 | 0 | 0 | 0 | 0 | 0 | 0 | 0 | 0 | 0 | 0 | 0 | 0 | 0 | 0 | **0.03** | 0 | 0 | 0 | 0 | 0 |
| | FL572 | 0 | **0.03** | 0 | 0 | 0 | 0 | 0 | 0 | 0 | 0 | 0 | 0 | 0 | 0 | 0 | 0 | 0 | 0 | 0 | 0 | 0 | 0 |
| | FL574 | 0 | 0 | 0 | 0 | 0 | 0 | 0 | 0 | 0 | 0 | 0 | 0 | 0 | 0 | 0 | **0.11** | 0 | 0 | 0 | 0 | 0 | 0 |
| | FL580 | 0 | 0 | 0 | 0 | **0.10** | 0 | 0 | 0 | 0 | 0 | 0 | 0 | 0 | 0 | 0 | **0.08** | 0 | 0 | 0 | 0 | 0 | 0 |
| | FL591 | 0 | 0 | 0 | 0 | 0 | 0 | 0 | 0 | 0 | 0 | 0 | 0 | 0 | 0 | 0 | **0.03** | 0 | 0 | 0 | 0 | 0 | 0 |
| | FL595 | 0 | 0 | 0 | 0 | 0 | 0 | 0 | 0 | 0 | **0.18** | 0 | 0 | 0 | 0 | 0 | 0 | 0 | 0 | 0 | 0 | 0 | 0 |
| | FL598 | 0 | 0 | 0 | 0 | 0 | 0 | 0 | **0.38** | 0 | 0 | 0 | 0 | 0 | 0 | 0 | 0 | 0 | 0 | 0 | 0 | 0 | 0 |
| | FL601 | 0 | 0 | 0 | 0 | 0 | 0 | 0 | 0 | 0 | 0 | 0 | 0 | 0 | 0 | 0 | 0 | 0 | 0 | 0 | 0 | 0 | **0.01** |
| | FL608 | 0 | 0 | 0 | 0 | 0 | 0 | 0 | 0 | 0 | 0 | 0 | 0 | 0 | 0 | 0 | 0 | **0.01** | 0 | 0 | 0 | 0 | 0 |
| | FL619 | 0 | 0 | 0 | 0 | 0 | 0 | 0 | 0 | 0 | 0 | 0 | 0 | 0 | 0 | 0 | **0.02** | 0 | 0 | 0 | 0 | 0 | 0 |

Table 16: Donor weights for Yishun sectors for models 3 and 4. Only sectors with any contribution to synthetic control weights for intervention sectors' synthetic controls are shown

| Donor | | Model 3 | | | | | | | | | | Model 4 | | | | | | | | | |
|---|---|---|---|---|---|---|---|---|---|---|---|---|---|---|---|---|---|---|---|---|---|
| Town | Sector | FL582 | FL584 | FL585 | FL585b | FL588 | FL589 | FL602 | FL609 | FL610 | FL618 | FL582 | FL584 | FL585 | FL585b | FL588 | FL589 | FL602 | FL609 | FL610 | FL618 |
| Ang Mo Kio | FL18 | 0 | 0 | 0 | 0 | 0 | 0 | 0 | **0.08** | 0 | 0 | 0 | 0 | 0 | 0 | 0 | 0 | 0 | 0 | 0 | 0 |
| | FL360 | 0 | 0 | 0 | 0 | 0 | 0 | 0 | 0 | 0 | 0 | 0 | 0 | **0.08** | 0 | 0 | 0 | 0 | **0.06** | 0 | 0 |
| | FL372 | 0 | 0 | 0 | 0 | 0 | 0 | 0 | 0 | 0 | 0 | 0 | **0.10** | 0 | 0 | 0 | 0 | 0 | **0.04** | 0 | 0 |
| | FL454 | **0.03** | 0 | **0.11** | **0.07** | 0 | 0 | **0.02** | 0 | 0 | **0.07** | 0 | 0 | 0 | 0 | 0 | 0 | 0 | 0 | 0 | 0 |
| | FL456 | 0 | 0 | 0 | 0 | 0 | 0 | 0 | 0 | 0 | 0 | 0 | 0 | 0 | 0 | 0 | 0 | **0.04** | 0 | 0 | 0 |
| | FL471 | 0 | 0 | 0 | 0 | 0 | 0 | 0 | 0 | 0 | **0.05** | 0 | 0 | 0 | 0 | 0 | 0 | 0 | 0 | 0 | 0 |
| | FL477 | 0 | 0 | 0 | **0.20** | 0 | 0 | **0.14** | 0 | 0 | 0 | 0 | 0 | 0 | **0.12** | 0 | 0 | **0.14** | 0 | 0 | 0 |
| | FL488 | 0 | 0 | 0 | 0 | 0 | 0 | 0 | **0.28** | 0 | 0 | 0 | 0 | 0 | 0 | 0 | 0 | 0 | **0.14** | 0 | 0 |
| | FL499 | 0 | 0 | 0 | 0 | **0.04** | 0 | 0 | 0 | 0 | 0 | 0 | 0 | 0 | 0 | 0 | 0 | 0 | 0 | 0 | 0 |
| | FL500 | 0 | 0 | 0 | 0 | 0 | 0 | 0 | 0 | 0 | 0 | 0 | 0 | 0 | 0 | 0 | 0 | 0 | **0.01** | 0 | 0 |
| | FL505 | 0 | 0 | 0 | 0 | 0 | 0 | 0 | 0 | 0 | 0 | 0 | 0 | 0 | 0 | 0 | 0 | 0 | **0.12** | 0 | 0 |
| Bedok | FL211 | 0 | 0 | 0 | 0 | 0 | 0 | 0 | 0 | 0 | 0 | 0 | **0.04** | 0 | 0 | 0 | 0 | 0 | 0 | 0 | **0.06** |
| | FL216 | 0 | 0 | 0 | 0 | 0 | 0 | 0 | 0 | 0 | 0 | 0 | **0.06** | 0 | 0 | 0 | 0 | 0 | 0 | 0 | 0 |
| | FL225 | 0 | 0 | 0 | 0 | 0 | 0 | 0 | 0 | 0 | 0 | 0 | 0 | 0 | 0 | 0 | 0 | 0 | 0 | 0 | **0.04** |
| | FL231 | 0 | 0 | 0 | 0 | 0 | 0 | 0 | 0 | **0.26** | 0 | 0 | 0 | 0 | 0 | 0 | 0 | 0 | 0 | 0 | 0 |
| | FL237 | 0 | 0 | 0 | 0 | 0 | 0 | 0 | 0 | 0 | 0 | **0.01** | 0 | **0.01** | **0.11** | 0 | 0 | 0 | 0 | 0 | 0 |

Continued on next page

**Table 16 – continued from previous page**

| Donor | | Model 3 | | | | | | | | | | Model 4 | | | | | | | | | |
|---|---|---|---|---|---|---|---|---|---|---|---|---|---|---|---|---|---|---|---|---|---|
| Town | Sector | FL582 | FL584 | FL585 | FL585b | FL588 | FL589 | FL602 | FL609 | FL610 | FL618 | FL582 | FL584 | FL585 | FL585b | FL588 | FL589 | FL602 | FL609 | FL610 | FL618 |
| | FL303 | 0 | 0 | 0 | 0 | 0 | 0 | 0 | 0 | 0 | 0 | **0.02** | **0.01** | 0 | 0 | 0 | 0 | 0 | 0 | 0 | 0 |
| Bishan | FL34 | **0.12** | 0 | 0 | 0 | 0 | **0.20** | **0.01** | 0 | **0.10** | 0 | 0 | 0 | 0 | 0 | 0 | **0.19** | 0 | 0 | **0.24** | 0 |
| Bukit Batok | FL686 | 0 | 0 | 0 | 0 | 0 | 0 | 0 | 0 | 0 | 0 | 0 | 0 | 0 | 0 | 0 | **0.01** | 0 | 0 | 0 | 0 |
| | FL699 | 0 | 0 | 0 | 0 | 0 | 0 | 0 | 0 | 0 | 0 | 0 | 0 | 0 | 0 | **0.08** | **0.13** | 0 | 0 | **0.23** | 0 |
| | FL731 | 0 | 0 | 0 | 0 | 0 | 0 | 0 | 0 | **0.33** | 0 | 0 | 0 | 0 | 0 | 0 | 0 | 0 | 0 | 0 | 0 |
| Bukit Merah | FL144 | 0 | 0 | 0 | 0 | 0 | 0 | 0 | **0.13** | 0 | 0 | 0 | 0 | 0 | 0 | 0 | 0 | 0 | 0 | 0 | 0 |
| | FL146 | 0 | 0 | 0 | 0 | 0 | 0 | 0 | **0.01** | 0 | 0 | 0 | 0 | 0 | 0 | 0 | 0 | 0 | 0 | 0 | 0 |
| | FL147 | 0 | 0 | 0 | 0 | 0 | 0 | 0 | 0 | 0 | 0 | **0.03** | 0 | 0 | 0 | 0 | 0 | 0 | 0 | 0 | **0.12** |
| | FL160 | 0 | 0 | 0 | 0 | 0 | 0 | 0 | 0 | 0 | 0 | 0 | 0 | 0 | 0 | 0 | 0 | 0 | 0 | 0 | **0.01** |
| | FL174 | 0 | 0 | 0 | 0 | **0.20** | 0 | 0 | 0 | 0 | 0 | 0 | **0.05** | 0 | 0 | 0 | 0 | 0 | 0 | 0 | 0 |
| | FL176 | 0 | 0 | 0 | 0 | 0 | 0 | 0 | 0 | 0 | 0 | 0 | **0.01** | **0.02** | 0 | 0 | 0 | 0 | 0 | 0 | 0 |
| | FL179 | 0 | 0 | 0 | 0 | 0 | 0 | 0 | 0 | 0 | 0 | **0.02** | 0 | 0 | 0 | 0 | 0 | 0 | 0 | 0 | 0 |
| | FL193 | 0 | **0.13** | 0 | 0 | 0 | 0 | 0 | 0 | 0 | 0 | 0 | **0.02** | 0 | 0 | 0 | 0 | 0 | 0 | 0 | 0 |
| Bukit Panjang | FL647 | 0 | **0.05** | 0 | 0 | **0.13** | 0 | 0 | 0 | 0 | 0 | 0 | 0 | 0 | 0 | **0.18** | 0 | 0 | 0 | 0 | 0 |
| | FL648 | 0 | 0 | **0.05** | 0 | 0 | 0 | 0 | 0 | 0 | 0 | 0 | 0 | **0.01** | 0 | 0 | 0 | 0 | 0 | 0 | 0 |
| | FL829 | 0 | 0 | **0.07** | 0 | 0 | 0 | 0 | 0 | 0 | **0.08** | 0 | 0 | 0 | 0 | **0.04** | 0 | 0 | 0 | 0 | 0 |
| Choa Chu Kang | FL729 | **0.02** | 0 | 0 | **0.03** | 0 | 0 | **0.23** | **0.16** | 0 | 0 | 0 | 0 | 0 | 0 | 0 | 0 | **0.06** | **0.01** | 0 | 0 |
| | FL764 | 0 | 0 | 0 | 0 | 0 | 0 | 0 | **0.03** | 0 | 0 | 0 | 0 | **0.09** | 0 | 0 | 0 | 0 | 0 | 0 | 0 |
| | FL811 | **0.20** | 0 | 0 | 0 | 0 | 0 | 0 | **0.25** | 0 | 0 | **0.15** | **0.04** | **0.01** | 0 | 0 | 0 | 0 | **0.06** | 0 | 0 |
| | FL814 | 0 | **0.27** | 0 | 0 | 0 | 0 | 0 | 0 | 0 | 0 | 0 | **0.05** | 0 | 0 | **0.14** | 0 | 0 | 0 | 0 | 0 |
| | FL818 | **0.16** | **0.10** | **0.17** | **0.27** | **0.04** | 0 | 0 | 0 | 0 | 0 | 0 | **0.10** | 0 | **0.11** | **0.05** | 0 | 0 | 0 | 0 | 0 |
| | FL823 | 0 | 0 | 0 | 0 | 0 | 0 | 0 | 0 | 0 | 0 | 0 | 0 | 0 | 0 | 0 | 0 | 0 | **0.06** | 0 | 0 |
| Clementi | FL428 | 0 | 0 | 0 | 0 | 0 | 0 | 0 | 0 | 0 | 0 | 0 | 0 | 0 | 0 | **0.03** | 0 | 0 | 0 | 0 | 0 |
| | FL640 | 0 | 0 | 0 | 0 | 0 | 0 | 0 | 0 | 0 | 0 | 0 | **0.07** | 0 | 0 | 0 | 0 | 0 | 0 | 0 | 0 |
| Geylang | FL200 | 0 | 0 | 0 | 0 | **0.04** | 0 | 0 | 0 | 0 | 0 | 0 | 0 | 0 | 0 | 0 | 0 | **0.04** | 0 | 0 | 0 |
| | FL204 | 0 | 0 | 0 | 0 | 0 | 0 | 0 | 0 | 0 | 0 | 0 | 0 | 0 | 0 | **0.02** | 0 | 0 | 0 | 0 | 0 |
| | FL81 | 0 | 0 | 0 | 0 | 0 | 0 | 0 | 0 | 0 | 0 | 0 | 0 | 0 | 0 | 0 | 0 | 0 | 0 | 0 | **0.06** |
| | FL83 | 0 | 0 | 0 | 0 | 0 | 0 | 0 | 0 | 0 | 0 | **0.02** | 0 | 0 | 0 | 0 | 0 | 0 | 0 | **0.04** | 0 |
| Hougang | FL367 | 0 | 0 | 0 | 0 | 0 | 0 | 0 | 0 | 0 | **0.06** | 0 | 0 | **0.04** | 0 | 0 | 0 | 0 | 0 | 0 | **0.02** |
| | FL451 | 0 | 0 | 0 | 0 | **0.21** | **0.06** | 0 | 0 | 0 | **0.35** | **0.14** | 0 | 0 | 0 | 0 | **0.02** | 0 | 0 | 0 | 0 |
| | FL452 | 0 | 0 | 0 | 0 | 0 | 0 | 0 | 0 | 0 | 0 | 0 | 0 | **0.09** | **0.06** | 0 | 0 | 0 | 0 | 0 | 0 |
| Jurong East | FL696 | 0 | 0 | 0 | 0 | 0 | 0 | 0 | 0 | 0 | 0 | 0 | 0 | 0 | 0 | 0 | 0 | **0.08** | 0 | 0 | 0 |
| | FL697 | 0 | **0.32** | 0 | 0 | 0 | 0 | 0 | 0 | 0 | 0 | **0.19** | **0.14** | **0.05** | **0.14** | 0 | 0 | **0.11** | 0 | 0 | 0 |
| | FL698 | 0 | 0 | 0 | 0 | 0 | 0 | 0 | 0 | 0 | 0 | **0.03** | 0 | 0 | 0 | 0 | 0 | 0 | 0 | 0 | 0 |
| | FL701 | 0 | 0 | 0 | 0 | 0 | 0 | 0 | 0 | 0 | 0 | 0 | 0 | 0 | 0 | 0 | 0 | **0.03** | 0 | 0 | 0 |
| Jurong West | FL431 | 0 | 0 | 0 | 0 | 0 | 0 | 0 | 0 | 0 | 0 | 0 | 0 | 0 | 0 | 0 | 0 | 0 | **0.03** | 0 | **0.10** |
| | FL441 | **0.15** | **0** | 0 | 0 | 0 | 0 | 0 | 0 | 0 | 0 | 0 | 0 | 0 | 0 | 0 | 0 | 0 | 0 | 0 | **0.02** |
| | FL442 | 0 | 0 | 0 | 0 | 0 | 0 | 0 | 0 | 0 | 0 | **0.10** | 0 | 0 | 0 | 0 | 0 | 0 | 0 | 0 | 0 |
| | FL621 | 0 | 0 | 0 | 0 | **0.18** | **0.59** | 0 | 0 | **0.18** | 0 | 0 | 0 | 0 | 0 | 0 | **0.55** | 0 | 0 | **0.06** | 0 |
| | FL652 | 0 | 0 | 0 | 0 | 0 | 0 | 0 | 0 | **0.12** | 0 | 0 | 0 | 0 | 0 | 0 | 0 | 0 | 0 | **0.04** | 0 |
| | FL653 | 0 | 0 | 0 | 0 | 0 | 0 | 0 | 0 | 0 | 0 | 0 | 0 | 0 | 0 | 0 | 0 | 0 | 0 | 0 | **0.06** |
| | FL662 | 0 | 0 | 0 | 0 | 0 | 0 | 0 | 0 | 0 | 0 | **0.02** | 0 | 0 | 0 | 0 | 0 | 0 | 0 | 0 | 0 |
| | FL694 | 0 | 0 | **0.29** | 0 | 0 | 0 | 0 | 0 | 0 | 0 | 0 | **0** | **0.11** | 0 | 0 | 0 | 0 | 0 | 0 | 0 |
| | FL710 | 0 | 0 | 0 | 0 | 0 | 0 | 0 | 0 | 0 | 0 | 0 | 0 | 0 | 0 | **0.07** | 0 | 0 | 0 | 0 | 0 |
| | FL769 | **0.10** | 0 | 0 | 0 | 0 | 0 | 0 | 0 | 0 | 0 | 0 | 0 | 0 | 0 | 0 | 0 | 0 | 0 | 0 | 0 |
| Kallang | FL183 | 0 | 0 | 0 | 0 | 0 | 0 | 0 | **0.04** | 0 | 0 | 0 | 0 | 0 | 0 | 0 | 0 | 0 | 0 | 0 | 0 |
| | FL185 | 0 | **0.04** | 0 | 0 | 0 | 0 | 0 | 0 | 0 | 0 | 0 | **0.07** | 0 | 0 | **0.12** | 0 | **0.02** | 0 | 0 | 0 |
| | FL26 | 0 | 0 | 0 | 0 | 0 | 0 | 0 | 0 | 0 | 0 | 0 | 0 | 0 | 0 | 0 | 0 | 0 | **0.02** | 0 | **0.02** |
| | FL85 | 0 | 0 | 0 | 0 | 0 | 0 | 0 | 0 | 0 | 0 | 0 | 0 | 0 | 0 | **0.03** | 0 | 0 | 0 | 0 | 0 |
| | FL93 | 0 | 0 | 0 | 0 | 0 | 0 | 0 | **0.03** | 0 | 0 | 0 | 0 | 0 | 0 | 0 | 0 | 0 | **0.02** | 0 | 0 |
| Marine Parade | FL310 | 0 | 0 | 0 | 0 | 0 | 0 | 0 | 0 | 0 | 0 | 0 | 0 | 0 | 0 | 0 | 0 | **0.05** | 0 | 0 | 0 |
| | FL318 | 0 | 0 | 0 | 0 | 0 | 0 | 0 | 0 | 0 | 0 | 0 | 0 | 0 | **0.10** | 0 | 0 | 0 | 0 | 0 | 0 |
| Novena | FL70 | 0 | 0 | 0 | 0 | 0 | 0 | 0 | 0 | 0 | 0 | 0 | **0.07** | 0 | 0 | 0 | 0 | 0 | 0 | 0 | 0 |
| Pasir Ris | FL256 | 0 | 0 | 0 | 0 | 0 | 0 | 0 | 0 | 0 | 0 | 0 | 0 | **0.01** | 0 | 0 | 0 | 0 | 0 | **0.06** | 0 |
| | FL293 | 0 | **0.09** | 0 | 0 | 0 | 0 | 0 | 0 | 0 | 0 | 0 | **0.05** | 0 | 0 | 0 | 0 | 0 | 0 | 0 | 0 |
| | FL321 | 0 | 0 | 0 | 0 | **0.16** | 0 | 0 | 0 | 0 | 0 | 0 | 0 | 0 | 0 | 0 | 0 | 0 | 0 | 0 | **0.02** |
| | FL348 | 0 | 0 | 0 | 0 | 0 | 0 | **0.36** | 0 | 0 | 0 | 0 | 0 | 0 | 0 | 0 | 0 | **0.20** | 0 | 0 | 0 |
| Queenstown | FL31 | 0 | 0 | 0 | 0 | 0 | 0 | 0 | 0 | 0 | 0 | 0 | 0 | 0 | 0 | **0.03** | 0 | 0 | 0 | 0 | 0 |
| | FL347 | 0 | 0 | 0 | 0 | 0 | 0 | 0 | 0 | 0 | 0 | 0 | 0 | **0.11** | 0 | 0 | 0 | 0 | 0 | 0 | 0 |
| | FL349 | 0 | 0 | 0 | 0 | 0 | 0 | 0 | 0 | 0 | 0 | 0 | 0 | 0 | 0 | **0.05** | 0 | 0 | 0 | 0 | 0 |
| Sembawang | FL527 | 0 | 0 | **0.01** | 0 | 0 | 0 | **0.24** | 0 | 0 | **0.02** | 0 | 0 | **0.02** | 0 | 0 | 0 | **0.05** | 0 | 0 | 0 |
| Sengkang | FL133 | 0 | 0 | 0 | 0 | 0 | 0 | 0 | 0 | 0 | 0 | 0 | 0 | 0 | 0 | 0 | 0 | 0 | **0.06** | 0 | 0 |
| | FL136 | 0 | 0 | 0 | 0 | 0 | 0 | 0 | 0 | 0 | 0 | **0.04** | 0 | 0 | 0 | 0 | 0 | 0 | **0.08** | 0 | 0 |
| | FL368 | 0 | 0 | 0 | 0 | 0 | 0 | 0 | 0 | 0 | 0 | **0.01** | 0 | 0 | 0 | 0 | 0 | 0 | 0 | 0 | 0 |
| | FL369 | 0 | 0 | 0 | 0 | 0 | 0 | 0 | 0 | 0 | **0.11** | 0 | 0 | 0 | 0 | 0 | 0 | 0 | 0 | 0 | 0 |
| | FL492 | 0 | 0 | 0 | **0.13** | 0 | 0 | 0 | 0 | 0 | 0 | 0 | 0 | 0 | **0.04** | 0 | 0 | 0 | **0.08** | 0 | **0.11** |
| | FL497 | 0 | 0 | 0 | 0 | 0 | 0 | 0 | 0 | 0 | 0 | 0 | 0 | **0.02** | 0 | 0 | 0 | 0 | 0 | 0 | 0 |
| | FL535 | **0.22** | 0 | 0 | 0 | 0 | 0 | 0 | 0 | 0 | 0 | **0.01** | 0 | 0 | 0 | 0 | 0 | 0 | 0 | 0 | **0.01** |
| | FL537 | 0 | 0 | 0 | 0 | 0 | 0 | 0 | 0 | 0 | 0 | 0 | **0.01** | 0 | 0 | **0** | **0.02** | 0 | 0 | 0 | 0 |
| | FL733 | 0 | 0 | 0 | 0 | 0 | 0 | 0 | 0 | 0 | 0 | **0.03** | 0 | 0 | **0.02** | 0 | 0 | 0 | 0 | 0 | **0.04** |
| | FL759 | 0 | 0 | 0 | 0 | 0 | 0 | 0 | 0 | 0 | 0 | 0 | 0 | 0 | 0 | **0.03** | 0 | 0 | 0 | 0 | 0 |
| | FL393 | 0 | 0 | 0 | 0 | 0 | 0 | 0 | 0 | 0 | 0 | 0 | 0 | 0 | 0 | 0 | 0 | 0 | 0 | 0 | **0.05** |

Continued on next page

Serangoon

**Table 16 – continued from previous page**

| Donor | | Model 3 | | | | | | | | | | Model 4 | | | | | | | | | |
|---|---|---|---|---|---|---|---|---|---|---|---|---|---|---|---|---|---|---|---|---|---|
| Town | Sector | FL582 | FL584 | FL585 | FL585b | FL588 | FL589 | FL602 | FL609 | FL610 | FL618 | FL582 | FL584 | FL585 | FL585b | FL588 | FL589 | FL602 | FL609 | FL610 | FL618 |
| | FL460 | 0 | 0 | 0 | 0 | 0 | 0 | 0 | 0 | 0 | **0.09** | 0 | 0 | 0 | 0 | 0 | 0 | 0 | 0 | 0 | **0.06** |
| | FL461 | 0 | 0 | 0 | 0 | 0 | 0 | 0 | 0 | 0 | 0 | 0 | 0 | 0 | 0 | 0 | 0 | 0 | **0.01** | 0 | 0 |
| | FL480 | 0 | 0 | 0 | 0 | 0 | 0 | 0 | 0 | 0 | **0.17** | 0 | 0 | 0 | 0 | 0 | 0 | 0 | 0 | 0 | **0.07** |
| | FL501 | 0 | 0 | 0 | 0 | 0 | 0 | 0 | 0 | 0 | 0 | 0 | 0 | 0 | 0 | 0 | 0 | 0 | 0 | 0 | **0.04** |
| Toa Payoh | FL164 | 0 | 0 | 0 | 0 | 0 | **0.15** | 0 | 0 | 0 | 0 | 0 | **0.07** | 0 | 0 | **0.02** | **0.08** | 0 | 0 | **0.29** | 0 |
| | FL319 | 0 | 0 | 0 | 0 | 0 | 0 | 0 | 0 | 0 | 0 | 0 | 0 | **0.20** | 0 | 0 | 0 | 0 | 0 | 0 | 0 |
| | FL42 | 0 | 0 | 0 | 0 | 0 | 0 | 0 | 0 | 0 | 0 | 0 | 0 | 0 | 0 | 0 | 0 | **0.09** | 0 | 0 | 0 |
| | FL58 | 0 | 0 | 0 | 0 | 0 | 0 | 0 | 0 | 0 | 0 | 0 | 0 | 0 | 0 | **0.04** | 0 | 0 | 0 | 0 | **0.02** |
| | FL59 | 0 | 0 | 0 | 0 | 0 | 0 | 0 | 0 | 0 | 0 | 0 | 0 | 0 | 0 | 0 | 0 | 0 | 0 | **0.04** | 0 |
| | FL61 | 0 | 0 | 0 | 0 | 0 | 0 | 0 | 0 | 0 | 0 | **0.16** | 0 | 0 | 0 | 0 | 0 | 0 | **0.02** | 0 | 0 |
| Woodlands | FL232 | 0 | 0 | 0 | 0 | 0 | 0 | 0 | 0 | 0 | 0 | 0 | 0 | 0 | 0 | 0 | 0 | **0.08** | 0 | 0 | 0 |
| | FL246 | 0 | 0 | 0 | 0 | 0 | 0 | 0 | 0 | 0 | 0 | 0 | 0 | 0 | 0 | 0 | 0 | 0 | **0.14** | 0 | 0 |
| | FL250 | 0 | 0 | 0 | 0 | 0 | 0 | 0 | 0 | 0 | 0 | **0.01** | 0 | 0 | **0.06** | 0 | 0 | 0 | **0.03** | 0 | **0.04** |
| | FL518 | 0 | 0 | 0 | 0 | 0 | 0 | 0 | 0 | 0 | 0 | 0 | 0 | 0 | 0 | **0.02** | 0 | 0 | 0 | 0 | 0 |
| | FL575 | 0 | 0 | 0 | 0 | 0 | 0 | 0 | 0 | 0 | 0 | 0 | 0 | 0 | 0 | 0 | 0 | 0 | **0.01** | 0 | 0 |
| | FL580 | 0 | 0 | **0.13** | **0.31** | 0 | 0 | 0 | 0 | 0 | 0 | 0 | 0 | **0.12** | **0.24** | **0.03** | 0 | 0 | 0 | 0 | 0 |
| | FL591 | 0 | 0 | 0 | 0 | 0 | 0 | 0 | 0 | 0 | 0 | 0 | 0 | 0 | 0 | 0 | 0 | 0 | 0 | 0 | **0.01** |
| | FL595 | 0 | 0 | **0.15** | 0 | 0 | 0 | 0 | 0 | 0 | 0 | 0 | 0 | 0 | 0 | 0 | 0 | 0 | 0 | 0 | 0 |
| | FL598 | 0 | 0 | 0 | 0 | 0 | 0 | 0 | 0 | 0 | 0 | 0 | **0.02** | 0 | 0 | 0 | 0 | 0 | 0 | 0 | 0 |
| | FL601 | 0 | 0 | 0 | 0 | 0 | 0 | 0 | 0 | 0 | 0 | 0 | 0 | 0 | 0 | 0 | 0 | 0 | **0.01** | 0 | 0 |

### 4 Placebo tests for intervention efficacy

To further assess the credibility and robustness of our estimated intervention effects of *Wolbachia* interventions on suppressing *Aedes aegypti* populations, we conducted 2 placebo and 1 permutation test.

#### 4.1 In-time placebo tests

We reran the synthetic control weighting scheme for the intervention group using the same donor pool with a placebo intervention date in a pre-specified time period before actual *Wolbachia* releases took place. We then evaluated the estimated intervention effect after the placebo intervention has occurred but before the actual *Wolbachia* releases took place. If the synthetic control generated closely reproduces the dengue incidence rate of intervention sites i.e. the root mean square error (RMSE) between observed dengue incidence rate and synthetic control should be close to 0. This provides evidence that the estimated gap computed after the post intervention date is the impact of *Wolbachia* releases rather than the synthetic controls having a lack of predictive power due to poor fit or pre-trends in dengue incidences which were occurring at the intervention sites (Abadie et al., 2015).

We took 2 dates as the start date for the placebo intervention – 13 weeks and 24 weeks prior to respective intervention start date for all the 4 study sites and **M2–4** were rerun. The RMSE between the synthetic control and intervention sites was then computed as:

$$RMSE_i = \left( \sum_{t=T^*-tp}^{T^*} \frac{\alpha_{it}^2}{tp} \right)^{1/2}$$

for $i = J+1, J+2, J+3, ..., J+N_{tr}$, where $tp = \{13, 24\}$ and $T^*$ corresponds to actual intervention start date for each $i$,

In Table 17, we report the root mean square prediction error between the synthetic control and outcome variable during the testing period. This hovers between 0.07 – 0.12 GAI over the placebo intervention periods of 13 and 24 weeks till before *Wolbachia* releases occurred, and is negligible compared to overall intervention efficacies as reported in the subsequent subsections. We further visualized the gaps for the 13 and 24 week placebo intervention periods across time. All synthetic controls constructed using the 3 weighting schemes for the four intervention sites followed that of actual observations closely in the post-placebo intervention periods (Figure 5,6).

| **Pseudo-intervention period** | **Bukit Batok** | **Choa Chu Kang** | **Tampines** | **Yishun** |
|---|---|---|---|---|
| Model 2 | | | | |
| 13 weeks | 0.09 | 0.07 | 0.10 | 0.10 |
| 24 weeks | 0.09 | 0.08 | 0.10 | 0.09 |
| Model 3 | | | | |
| 13 weeks | 0.09 | 0.08 | 0.10 | 0.12 |
| 24 weeks | 0.09 | 0.09 | 0.12 | 0.11 |
| Model 4 | | | | |
| 13 weeks | 0.08 | 0.07 | 0.10 | 0.13 |
| 24 weeks | 0.08 | 0.09 | 0.10 | 0.11 |

Table 17: RMSPE during the pseudo-intervention period of 13 weeks and 24 weeks prior to actual intervention aggregated to intervention town.

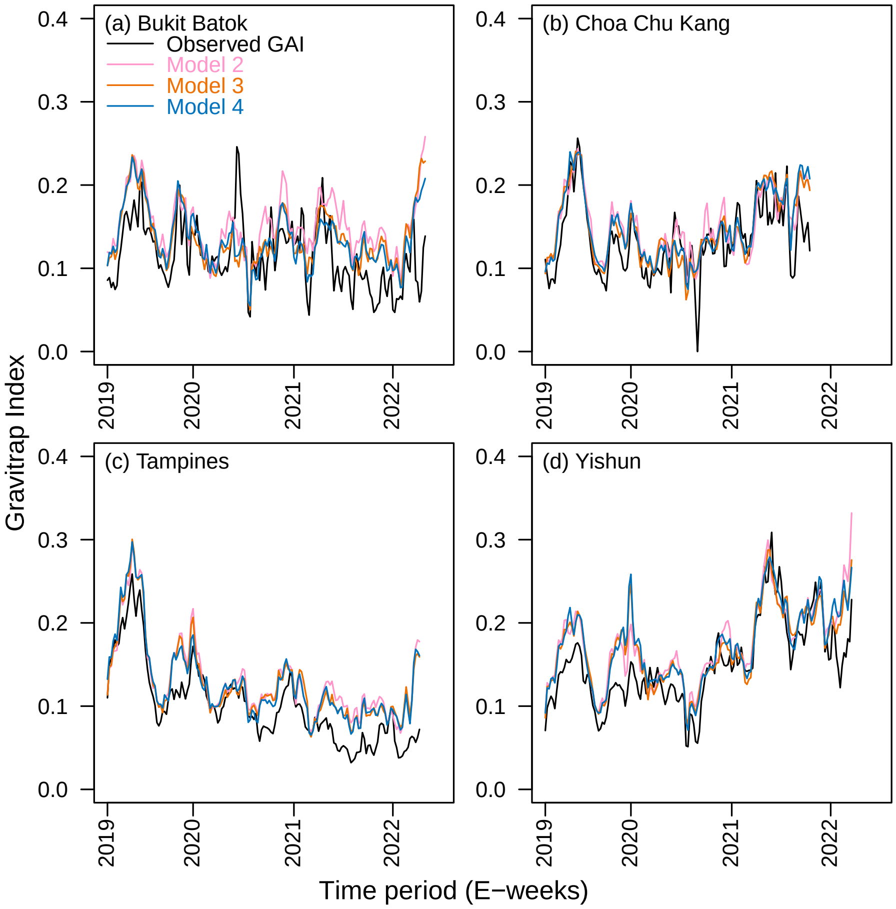

Figure 5: Gap of female *Aedes aegypti* abundance between synthetic control and intervention sites for all synthetic control weighting schemes 13 weeks prior to actual intervention date for each sector aggregated by (a) Bukit Batok (b) Choa Chu Kang (c) Tampines and (d) Yishun. The intervention sectors are aggregated to towns only during their respective pre-intervention period.

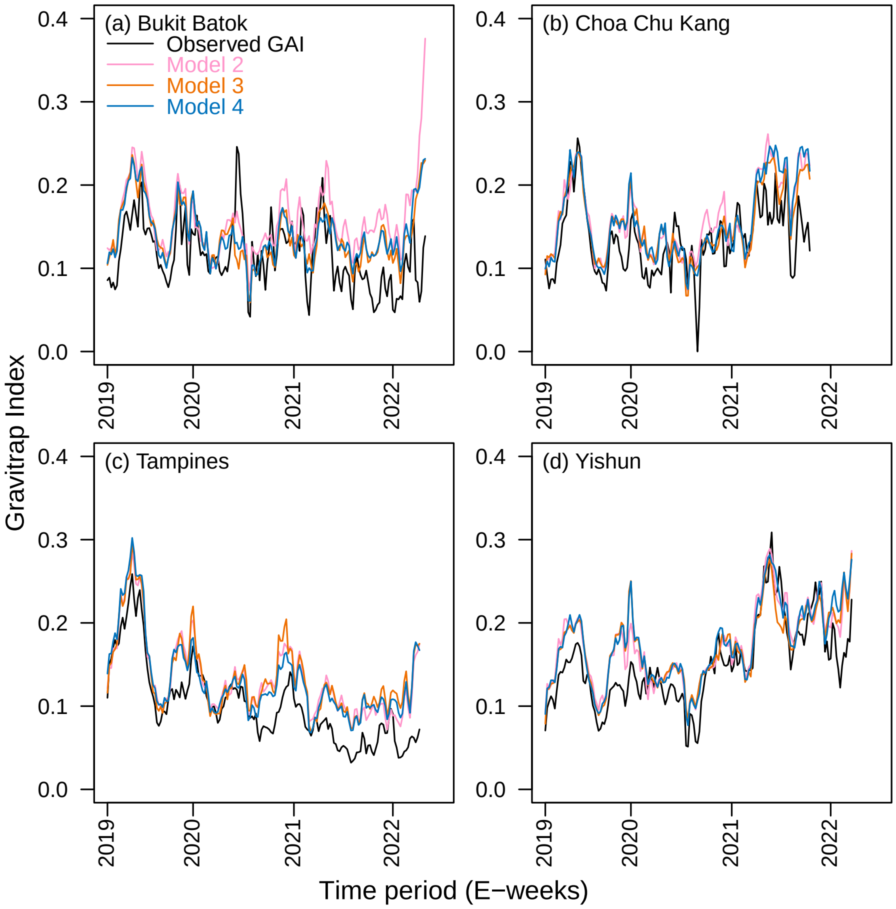

Figure 6: Gap of female *Aedes aegypti* abundance between synthetic control and intervention sites for all synthetic control weighting schemes 24 weeks prior to actual intervention date in (a) Bukit Batok (b) Choa Chu Kang (c) Tampines and (d) Yishun. The intervention sectors are aggregated to towns only during their respective pre-intervention period.

##### 4.2 In-space placebo tests

We further conducted placebo tests by reassigning the treatment in the data to control units while removing the actual intervention sites from the construction of synthetic controls for the placebo-intervention units. For each synthetic control weighting scheme, we iteratively took a control site as the placebo-intervention site and used the remaining control sites as the donor pool to construct the synthetic control. Thereafter, we estimated the placebo-intervention effect for the placebo-intervention site from EW1 of 2020 till EW26 2022 (period where *Wolbachia* releases occurred).

This procedure allowed us to obtain estimates for the intervention efficacy in regions that did not experience *Wolbachia* releases. Applying this idea to each region in the donor pool allows us to compare the estimated effect of *Wolbachia* release on the actual intervention sites to the distribution of placebo effects obtained from the construct of synthetic controls in the placebo-intervention regions. We will deem the effect of *Wolbachia* release as significant if the estimated effect for treated units is unusually large relative to the distribution of placebo effects (Abadie et al., 2015).

Table 18 shows that ratio of post to pre-RMSPE for intervention sectors is far higher than that for sectors which never received *Wolbachia* release.

| Model | 2 | 3 | 4 |
|---|---|---|---|
| Intervention Arm | | | |
| Bukit Batok | 3.51 | 3.67 | 4.80 |
| Choa Chu Kang | 4.28 | 4.17 | 5.53 |
| Tampines | 2.67 | 2.73 | 3.66 |
| Yishun | 3.83 | 4.14 | 5.13 |
| **Average (Intervention)** | 3.40 | 3.53 | 4.57 |
| Control Arm | | | |
| Ang Mo Kio | 0.50 | 0.48 | 0.38 |
| Bedok | 0.36 | 0.39 | 0.29 |
| Bishan | 0.41 | 0.39 | 0.30 |
| Bukit Batok | 0.42 | 0.38 | 0.34 |
| Bukit Merah | 0.33 | 0.36 | 0.25 |
| Bukit Panjang | 0.27 | 0.29 | 0.23 |
| Choa Chu Kang | 0.39 | 0.43 | 0.31 |
| Clementi | 0.27 | 0.27 | 0.20 |
| Geylang | 0.35 | 0.38 | 0.26 |
| Hougang | 0.42 | 0.38 | 0.27 |
| Jurong East | 0.51 | 0.48 | 0.40 |
| Jurong West | 0.44 | 0.45 | 0.35 |
| Kallang | 0.40 | 0.38 | 0.33 |
| Marine Parade | 0.48 | 0.38 | 0.27 |
| Novena | 0.29 | 0.34 | 0.20 |
| Outram | 0.32 | 0.37 | 0.18 |
| Pasir Ris | 0.42 | 0.43 | 0.31 |
| Queenstown | 0.31 | 0.23 | 0.17 |
| Rochor | 0.33 | 0.34 | 0.26 |
| Sembawang | 0.30 | 0.33 | 0.28 |
| Sengkang | 0.52 | 0.52 | 0.32 |
| Serangoon | 0.44 | 0.42 | 0.32 |
| Toa Payoh | 0.40 | 0.41 | 0.26 |
| Woodlands | 0.43 | 0.40 | 0.28 |
| **Average (Control)** | 0.42 | 0.41 | 0.30 |
| $\frac{\text{Avg Intervention}}{\text{Avg Control}}$ | 8.17 | 8.71 | 15.26 |

Table 18: Post/Pre-RMPSE for intervention arm and donor pool townships, aggregated by sector and by epidemiological week for endpoint of *Aedes aegypti* abundance across models 2 – 4 in post-intervention time

### 5 Robustness checks for other entomological outcomes

#### 5.1 Robustness checks for *Aedes albopictus* abundance

We performed the above stated in-space and in-time placebo checks as well as visual inspection for *Aedes albopictus* abundance using the model which performed best on previous assessment checks for the primary entomological endpoint of *Ae. aegypti* abundance (**M4**).

##### 5.1.1 Visual inspection for model fit

The model captures the *Aedes albopictus* abundance trends in the pre-intervention period for all the towns.

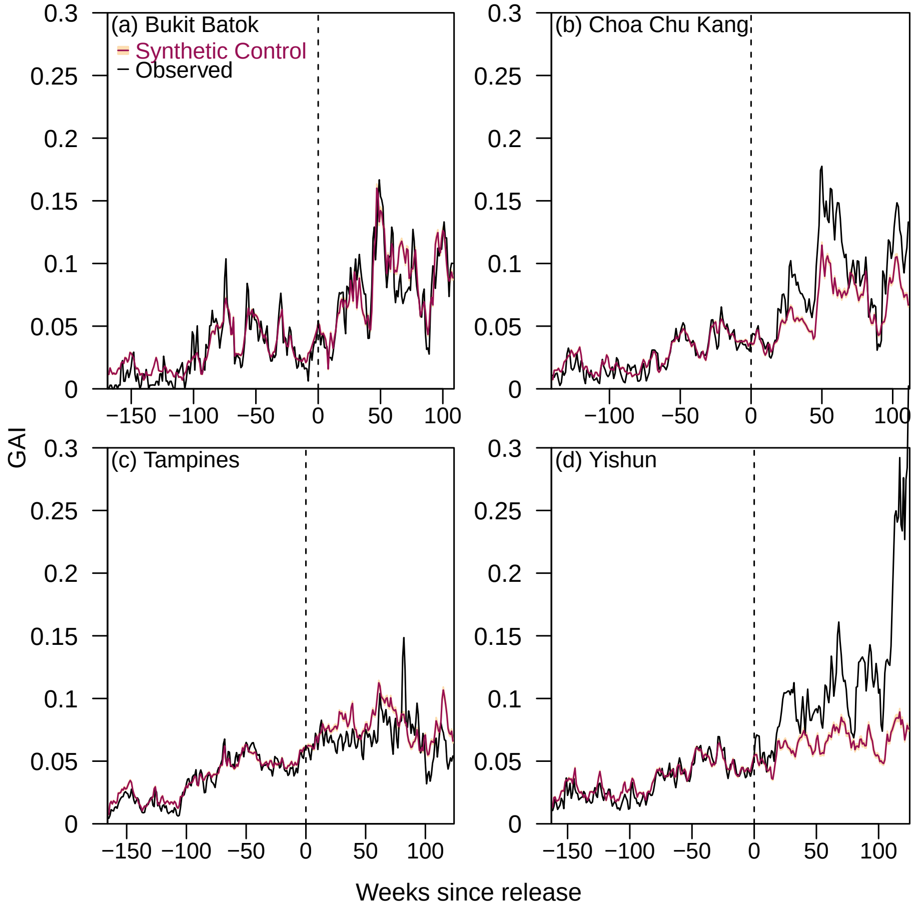

Figure 7: Observed and synthetic control *Aedes albopictus* abundance in (a) Bukit Batok (b) Choa Chu Kang (c) Tampines and (d) Yishun.

##### 5.1.2 In-space placebo checks for *Aedes albopictus* abundance

Table 19 shows the distribution of post to pre-RMSPE of the intervention arm versus the donor pool. We see that ratio of post to pre-RMSPE for intervention towns is higher than that for donor pool towns which never received *Wolbachia* release.

| Township | Post/Pre-RMPSE |
|---|---|
| Intervention Arm | |
| Bukit Batok | 2.53 |
| Choa Chu Kang | 4.84 |
| Tampines | 3.52 |
| Yishun | 6.69 |
| **Average** | 4.72 |
| Donor Pool | |
| Ang Mo Kio | 0.26 |
| Bedok | 0.25 |
| Bishan | 0.29 |
| Bukit Batok | 0.22 |
| Bukit Merah | 0.27 |
| Bukit Panjang | 0.21 |
| CCK | 0.26 |
| Clementi | 0.14 |
| Geylang | 0.15 |
| Hougang | 0.24 |
| Jurong East | 0.28 |
| Jurong West | 0.30 |
| Kallang | 0.25 |
| Marine Parade | 0.14 |
| Novena | 0.30 |
| Outram | 0.21 |
| Queenstown | 0.20 |
| Rochor | 0.21 |
| Sembawang | 0.13 |
| Senkang | 0.29 |
| Serangoon | 0.19 |
| Toa Payoh | 0.20 |
| Woodlands | 0.25 |
| **Average (Control)** | 0.24 |
| $\frac{\text{Avg Intervention}}{\text{Avg Control}}$ | 19.67 |

Table 19: Post/Pre-RMPSE for intervention arm and donor pool townships, aggregated by sector and by epidemiological week for endpoint of *Aedes albopictus* abundance for model 4.

###### 5.1.3 In-time placebo for *Aedes albopictus* abundance

Table 20 quantifies the gap between synthetic control and observed *Ae. albopictus* abundance, and shows that the average gap between the two in the pre-intervention period is 0.03, indicating a good model fit.

| **Validation period** | **Bukit Batok** | **Choa Chu Kang** | **Tampines** | **Yishun** |
|---|---|---|---|---|
| 13 | 0.03 | 0.02 | 0.03 | 0.02 |
| 24 | 0.02 | 0.03 | 0.03 | 0.03 |

Table 20: RMSPE during the validation period in the pre-intervention time for all intervention sectors aggregated by township for 13 and 24 weeks

##### 5.2 Robustness checks for *Aedes aegypti* abundance in non-release, adjacent sectors

###### 5.2.1 Visual inspection for model fit

The model captures the *Aedes aegypti* abundance trends in the pre-intervention period for all the towns. The pre-intervention period here is the period before sector becomes a non-release, adjacent sector, and the post-intervention period is during which the sector is non-release, adjacent sector.

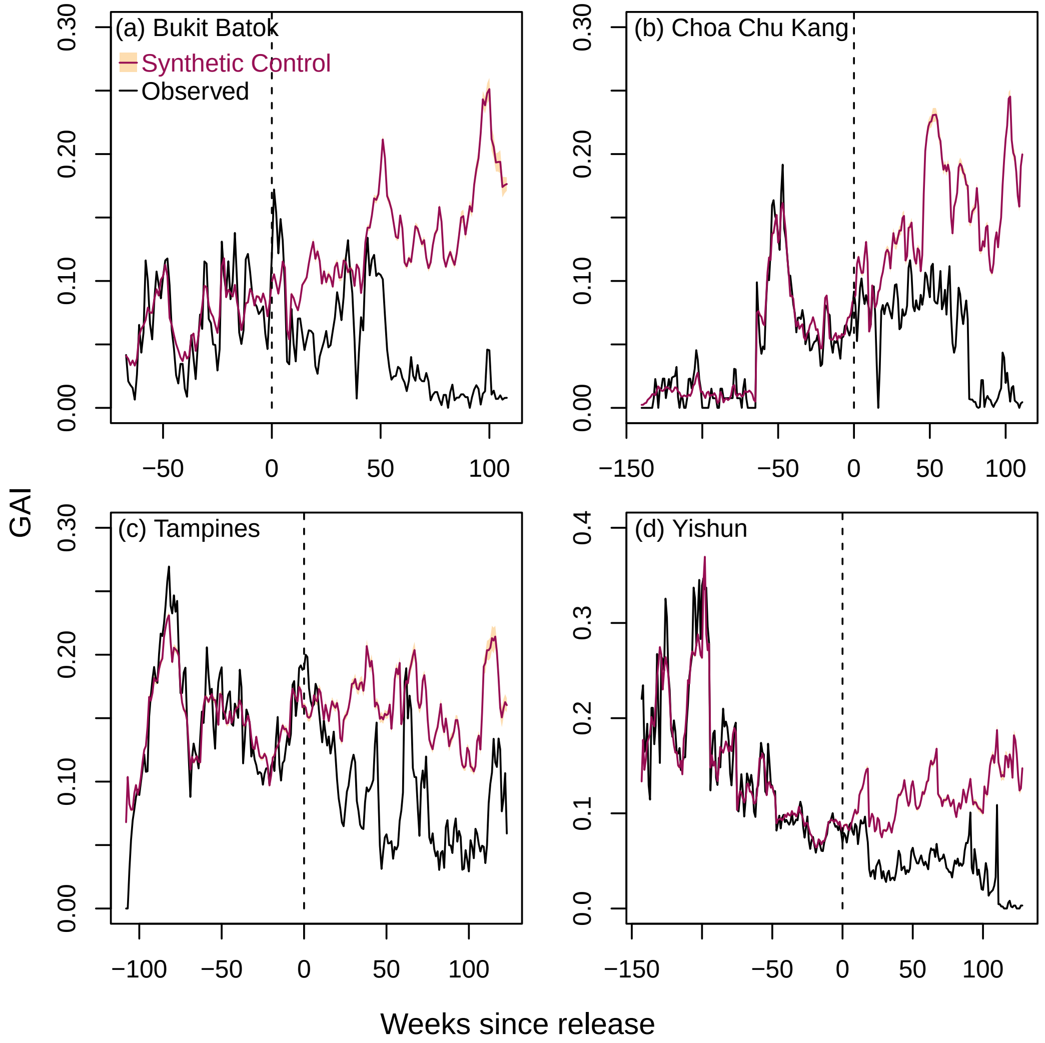

Figure 8: Observed and synthetic control *Aedes albopictus* abundance in (a) Bukit Batok (b) Choa Chu Kang (c) Tampines and (d) Yishun. The sectors are aggregated only until the sector is a non-release, adjacent sector.

###### 5.2.2 In-time placebo check for spillover locations

| Sector ID | RMSPE (13 weeks) | RMSPE (24 Weeks) |
|---|---|---|
| *Average* | 0.065 | 0.070 |
| FL122 | 0.087 | 0.168 |
| FL168 | 0.032 | 0.030 |
| FL172 | 0.095 | 0.116 |
| FL264 | 0.060 | 0.080 |
| FL281 | 0.024 | 0.026 |
| FL283 | 0.016 | 0.020 |
| FL290 | 0.042 | 0.100 |
| FL291 | 0.015 | 0.049 |
| FL292 | 0.110 | 0.107 |
| FL295 | 0.077 | 0.066 |
| FL296 | 0.042 | 0.036 |
| FL305 | 0.096 | 0.077 |
| FL312 | 0.022 | 0.049 |
| FL333 | 0.076 | 0.088 |
| FL337 | 0.260 | 0.272 |
| FL338 | 0.033 | 0.041 |
| FL341 | 0.107 | 0.083 |
| FL342 | 0.065 | 0.081 |
| FL343 | 0.027 | 0.033 |
| FL354 | 0.217 | 0.133 |
| FL355 | 0.047 | 0.040 |
| FL356 | 0.076 | 0.107 |
| FL357 | 0.062 | 0.076 |
| FL539 | 0.016 | 0.010 |
| FL564 | 0.085 | 0.058 |
| FL565 | 0.013 | 0.017 |
| FL566 | 0.015 | 0.012 |
| FL567 | 0.024 | 0.018 |
| FL583 | 0.033 | 0.038 |
| FL585b | 0.052 | 0.057 |
| FL589 | 0.055 | 0.093 |
| FL602 | 0.060 | 0.050 |
| FL634 | 0.079 | 0.103 |
| FL636 | 0.055 | 0.053 |
| FL637 | 0.106 | 0.068 |
| FL650 | 0.051 | 0.065 |
| FL729 | 0.067 | 0.060 |
| FL737 | 0.060 | 0.064 |
| FL741 | 0.007 | 0.016 |
| FL749 | 0.026 | 0.020 |
| FL751 | 0.007 | 0.008 |
| FL792 | 0.012 | 0.020 |
| FL798 | 0.075 | 0.079 |
| FL799 | 0.019 | 0.019 |
| FL826 | 0.074 | 0.078 |

| | | |
|---|---|---|
| FL837 | 0.250 | 0.193 |
| FL94 | 0.056 | 0.063 |
| FL95 | 0.097 | 0.188 |
| FL97 | 0.075 | 0.063 |
| FL98 | 0.108 | 0.086 |

Table 21: In-time placebo checks with accompanying root mean squared prediction errors (RMSPE) for spillover locations, 13 and 24 weeks before sectors become non-release, adjacent sectors.

### 6 Estimated intervention efficacy across entomological endpoints

Below, we report the intervention efficacy of *Wolbachia* interventions on *Aedes aegypti* and *Aedes albopictus* abundance using the best model (**M4**) by event time, calendar time and sector aggregates, to complement main-text figures on intervention efficacy.

#### 6.1 Estimated intervention efficacies on *Aedes aegypti* abundance for best model

| **Time period** | **Bukit Batok** | **Choa Chu Kang** | **Tampines** | **Yishun** |
|---|---|---|---|---|
| | Intervention Efficacies (Time from start of intervention) | | | |
| 1 – 6 months | 53.45% (52.42% – 54.43%) | 59.44% (58.5% – 60.34%) | 53.98% (53% – 54.91%) | 58.37% (57.4% – 59.29%) |
| 7 – 12 months | 93.3% (93.14% – 93.46%) | 83.43% (82.98% – 83.85%) | 69.05% (68.17% – 69.88%) | 88.02% (87.55% – 88.45%) |
| 13 – 18 months | 95.69% (95.61% – 95.77%) | 90.56% (90.34% – 90.76%) | 82.37% (82.01% – 82.72%) | 92.91% (92.69% – 93.12%) |
| 19 – 24 months | 95.33% (95.23% – 95.43%) | 95% (94.88% – 95.11%) | 85.05% (84.71% – 85.36%) | 91.54% (91.22% – 91.83%) |
| > 24 months | 94.93% (94.84% – 95.02%) | 96.5% (96.44% – 96.57%) | 91.26% (91.05% – 91.45%) | 90.88% (90.49% – 91.24%) |
| | Intervention Efficacies (Calendar Time) | | | |
| 2020 EW 27 – 52 | 71.09% (70.3% – 71.84%) | 70.63% (69.75% – 71.45%) | 48.52% (46.81% – 50.11%) | 70.33% (68.9% – 71.64%) |
| 2021 EW 1 – 26 | 93.65% (93.52% – 93.78%) | 84.74% (84.37% – 85.1%) | 61.54% (60.7% – 62.33%) | 78.55% (77.77% – 79.28%) |
| 2021 EW 27 – 52 | 95.82% (95.73% – 95.9%) | 87.79% (87.51% – 88.05%) | 83.41% (83.03% – 83.78%) | 86.81% (86.38% – 87.2%) |
| 2022 EW 1 – 26 | 91.69% (91.52% – 91.86%) | 94.08% (93.96% – 94.2%) | 86.52% (86.23% – 86.78%) | 85.78% (85.42% – 86.12%) |

Table 22: Intervention efficacy of *Wolbachia* interventions in reducing female *Aedes aegypti* abundance aggregated by sector and each township based on calendar time and time from start of intervention.

| Sector ID | Mean IE during intervention period | Sector ID | Mean IE during intervention period |
|---|---|---|---|
| FL118 | 46.92(45.5 – 48.27) | FL528 | 65.34(65.01 – 65.67) |
| FL119 | 75.67(74.98 – 76.32) | FL540 | 94.65(94.13 – 95.08) |
| FL120 | 69.4(68.44 – 70.32) | FL557 | 94.93(94.8 – 95.04) |
| FL121 | 82.3(82.03 – 82.56) | FL558 | 93.08(92.93 – 93.21) |
| FL122 | 92.1(91.97 – 92.23) | FL564 | 88.43(88.26 – 88.6) |
| FL165 | 61.57(60.79 – 62.32) | FL565 | 95.2(94.77 – 95.56) |
| FL170 | 76.63(76.17 – 77.08) | FL569 | 85.69(84.5 – 86.7) |
| FL171 | 92.1(91.89 – 92.29) | FL573 | 76.85(76.11 – 77.54) |
| FL172 | 89.42(89.28 – 89.55) | FL576 | 74.21(73.98 – 74.44) |
| FL278 | 89.44(88.78 – 90.02) | FL582 | 72.47(72.12 – 72.81) |
| FL283 | 94.21(93.79 – 94.58) | FL584 | 89.37(89.14 – 89.6) |
| FL285 | 68.85(68.48 – 69.21) | FL585 | 94.26(94.12 – 94.39) |
| FL290 | 15.64(14.89 – 16.38) | FL585b | 95.84(95.72 – 95.96) |
| FL291 | 39.34(37.89 – 40.72) | FL588 | 77.21(76.86 – 77.54) |
| FL295 | 64.01(63.7 – 64.32) | FL589 | 72.21(71.8 – 72.61) |
| FL296 | 56.56(55.72 – 57.36) | FL602 | 78.18(77.75 – 78.59) |
| FL297 | 78.88(77.91 – 79.77) | FL609 | 80.04(79.75 – 80.32) |
| FL298 | 60.96(60.4 – 61.51) | FL610 | 82.79(82.46 – 83.1) |
| FL304 | 86.76(86.48 – 87.03) | FL618 | 18.59(17.68 – 19.48) |
| FL305 | 58.27(57.64 – 58.88) | FL633 | 40.14(38.79 – 41.42) |
| FL314 | 86.71(86.31 – 87.08) | FL634 | 75.13(74.71 – 75.52) |
| FL315 | 30.64(29.6 – 31.64) | FL635 | 80.49(79.84 – 81.1) |
| FL316 | 56.25(55.32 – 57.15) | FL636 | 73.95(73.51 – 74.37) |
| FL326 | 59.1(58.08 – 60.07) | FL637 | 82.93(82.72 – 83.14) |
| FL327 | 57.67(57.21 – 58.13) | FL641 | 90.38(90.24 – 90.53) |
| FL328 | 44.1(43.28 – 44.9) | FL650 | 83.06(82.62 – 83.47) |
| FL332 | 88.29(87.89 – 88.67) | FL655 | 78.8(78.31 – 79.26) |
| FL333 | 89.58(89.33 – 89.81) | FL718 | 78.18(77.68 – 78.66) |
| FL337 | 24.21(20.82 – 27.32) | FL740 | 71.7(70.46 – 72.84) |
| FL340 | 79.04(78.56 – 79.5) | FL742 | 90.99(90.85 – 91.14) |
| FL341 | 86.87(86.6 – 87.14) | FL748 | 80.7(80.28 – 81.09) |
| FL342 | 94.6(94.5 – 94.7) | FL765 | 86.5(86.19 – 86.8) |
| FL343 | 31.13(29.8 – 32.41) | FL768 | 88.04(87.81 – 88.27) |
| FL344 | 74.08(73.54 – 74.6) | FL787 | 89.59(89.33 – 89.83) |
| FL350 | 98.53(98.46 – 98.59) | FL803 | 88.12(87.78 – 88.44) |
| FL353 | 84.38(84.17 – 84.58) | FL804 | 91.42(91.18 – 91.64) |
| FL354 | 55.43(54.76 – 56.08) | FL806 | 81.02(80.53 – 81.48) |
| FL355 | 90.9(90.71 – 91.08) | FL807 | 78.86(78.51 – 79.2) |
| FL356 | 79.53(79.2 – 79.84) | FL826 | 83.17(82.92 – 83.41) |
| FL357 | 80.77(80.09 – 81.4) | FL833 | 90.95(90.74 – 91.14) |
| FL420 | 82.15(81.91 – 82.38) | FL833b | 29.52(28.63 – 30.4) |
| FL421 | 80.47(80.18 – 80.76) | FL837 | 61.7(60.86 – 62.51) |
| FL424 | 89.35(89.12 – 89.58) | FL94 | 74.9(74.45 – 75.32) |
| FL425 | 87.82(87.43 – 88.19) | FL95 | 69.27(68.82 – 69.71) |
| FL426 | 88.76(88.56 – 88.95) | FL96 | 92.73(92.52 – 92.93) |
| FL517 | 54.14(53.62 – 54.64) | FL97 | 88.63(88.33 – 88.92) |
| FL523 | 78.13(76.64 – 79.45) | FL98 | 80.09(79.83 – 80.35) |

Table 23: Intervention efficacy of *Wolbachia* interventions in reducing female *Aedes aegypti* aggregated by sector across their respective intervention time

#### 6.2 Estimated intervention efficacies on *Aedes aegypti* abundance for other models

| Time period | Bukit Batok | Choa Chu Kang | Tampines | Yishun |
|---|---|---|---|---|
| | Intervention Efficacies (Time from start of intervention) | | | |
| 11 – 6 months | 49.08% (46.99% – 51.01%) | 61.01% (59.36% – 62.53%) | 50.27% (47.98% – 52.37%) | 58.32% (56.54% – 59.96%) |
| 7 – 12 months | 92.51% (92.14% – 92.85%) | 83.28% (82.56% – 83.95%) | 67.9% (66.33% – 69.32%) | 86.79% (86.07% – 87.44%) |
| 13 – 18 months | 95.38% (95.16% – 95.58%) | 90.02% (89.58% – 90.42%) | 80.39% (79.58% – 81.15%) | 91.89% (91.46% – 92.29%) |
| 19 – 24 months | 95.48% (95.28% – 95.67%) | 95% (94.73% – 95.25%) | 83.71% (83.06% – 84.31%) | 91.05% (90.54% – 91.52%) |
| > 24 months | 94.9% (94.7% – 95.08%) | 96.23% (96.07% – 96.38%) | 90.56% (90.18% – 90.92%) | 91.29% (90.75% – 91.77%) |
| | Intervention Efficacies (Calendar Time) | | | |
| 2020 EW 27 – 52 | 66.06% (64.5% – 67.49%) | 70.02% (68.55% – 71.36%) | 49.84% (47.18% – 52.25%) | 69.2% (67.35% – 70.85%) |
| 2021 EW 1 – 26 | 93.14% (92.8% – 93.44%) | 84.38% (83.75% – 84.95%) | 59.94% (58.26% – 61.49%) | 76.02% (74.9% – 77.05%) |
| 2021 EW 27 – 52 | 95.61% (95.39% – 95.8%) | 87.08% (86.47% – 87.64%) | 81.38% (80.56% – 82.14%) | 85.53% (84.73% – 86.26%) |
| 2022 EW 1 – 26 | 91.89% (91.54% – 92.21%) | 94.24% (93.96% – 94.49%) | 84.99% (84.34% – 85.58%) | 85.22% (84.48% – 85.89%) |

Table 24: Intervention efficacy of *Wolbachia* interventions in reducing female *Aedes aegypti* abundance aggregated by sector and each township based on calendar time and time from start of intervention using **Model 2**.

| Time period | Bukit Batok | Choa Chu Kang | Tampines | Yishun |
|---|---|---|---|---|
| | Intervention Efficacies (Time from start of intervention) | | | |
| 1 – 6 months | 45.12% (42.6% – 47.43%) | 58.02% (56.33% – 59.6%) | 53.78% (52.02% – 55.42%) | 58.15% (56.46% – 59.72%) |
| 7 – 12 months | 91.43% (90.9% – 91.9%) | 82.35% (81.5% – 83.12%) | 67.42% (65.65% – 69.03%) | 87.52% (86.74% – 88.21%) |
| 13 – 18 months | 94.95% (94.67% – 95.19%) | 89.85% (89.42% – 90.24%) | 80.47% (79.63% – 81.24%) | 92.2% (91.76% – 92.6%) |
| 19 – 24 months | 95.31% (95.09% – 95.51%) | 94.76% (94.49% – 95.01%) | 82.84% (82.12% – 83.5%) | 91.26% (90.74% – 91.71%) |
| > 24 months | 94.57% (94.36% – 94.76%) | 96.21% (96.07% – 96.35%) | 89.82% (89.44% – 90.17%) | 90.49% (89.85% – 91.06%) |
| | Intervention Efficacies (Calendar Time) | | | |
| 2020 EW 27 – 52 | 64.02% (62% – 65.84%) | 69.3% (67.8% – 70.66%) | 50.57% (47.83% – 53.04%) | 69.4% (67.26% – 71.27%) |
| 2021 EW 1 – 26 | 92.14% (91.7% – 92.54%) | 83.93% (83.18% – 84.62%) | 62.33% (60.82% – 63.73%) | 77.48% (76.15% – 78.67%) |
| 2021 EW 27 – 52 | 95.19% (94.93% – 95.43%) | 86.5% (85.9% – 87.04%) | 80.91% (80.04% – 81.71%) | 85.79% (84.99% – 86.51%) |
| 2022 EW 1 – 26 | 91.56% (91.2% – 91.89%) | 93.91% (93.64% – 94.16%) | 84.42% (83.76% – 85.03%) | 85.37% (84.71% – 85.98%) |

Table 25: Intervention efficacy of *Wolbachia* interventions in reducing female *Aedes aegypti* abundance aggregated by sector and each township based on calendar time and time from start of intervention using **Model 3**.

#### 6.3 Estimated intervention efficacies on *Aedes albopictus* abundance by event time for best model

| Time period | Bukit Batok | Choa Chu Kang | Tampines | Yishun |
|---|---|---|---|---|
| | Intervention Efficacies (Time from start of intervention) | | | |
| 1 – 6 months | 1.59% (-0.78% – 3.86%) | -13.13% (-16.05% – -10.35%) | 6.43% (4.61% – 8.17%) | -28.25% (-31.52% – -25.14%) |
| 7 – 12 months | -10.41% (-12.82% – -8.09%) | -51.21% (-54.81% – -47.78%) | 16.42% (14.75% – 18.04%) | -47.19% (-51.85% – -42.82%) |
| 13 – 18 months | 12.42% (10.57% – 14.19%) | -38.86% (-42.12% – -35.74%) | 17.76% (16.09% – 19.36%) | -54.69% (-59.37% – -50.29%) |
| 19 – 24 months | 3.46% (1.28% – 5.54%) | -23.81% (-26.93% – -20.85%) | -10.59% (-13.4% – -7.91%) | -77.2% (-83.01% – -71.75%) |
| > 24 months | -3.77% (-6.46% – -1.2%) | -47.99% (-51.37% – -44.76%) | 24.71% (22.69% – 26.62%) | -170.74% (-179.44% – -162.57%) |
| | Intervention Efficacies (Calendar time) | | | |
| 2020 EW 27 – 52 | -2.27% (-4.81% – 0.15%) | -44.33% (-48.49% – -40.39%) | 3.72% (0.8% – 6.46%) | -122.77% (-131.08% – -115.03%) |
| 2021 EW 1 – 26 | -6.91% (-9.19% – -4.72%) | -50.58% (-54.02% – -47.28%) | -12.61% (-14.84% – -10.47%) | -73.97% (-78.92% – -69.28%) |
| 2021 EW 27 – 52 | 11.76% (9.86% – 13.57%) | -25.28% (-28.34% – -22.36%) | 9.87% (8.02% – 11.65%) | -40.69% (-45.07% – -36.57%) |
| 2022 EW 1 – 26 | 3.21% (1.01% – 5.32%) | -27.01% (-29.98% – -24.17%) | 29.27% (27.86% – 30.62%) | -43.3% (-47.19% – -39.61%) |

Table 26: Intervention efficacy of *Wolbachia* interventions in reducing female *Aedes albopictus* abundance aggregated by sector and each township based on calendar time and time from start of intervention.

| Sector ID | Mean IE during intervention period | Sector ID | Mean IE during intervention period |
|---|---|---|---|
| FL118 | -7.13(-9.5 – -4.86) | FL528 | -38.12(-41.64 – -34.77) |
| FL119 | -164.79(-172.3 – -157.69) | FL540 | 35.66(33.93 – 37.31) |
| FL120 | 14.47(12.45 – 16.39) | FL557 | -71.7(-75.98 – -67.63) |
| FL121 | 45.07(43.48 – 46.58) | FL558 | -223.82(-235.87 – -212.61) |
| FL122 | 9.92(8.59 – 11.21) | FL564 | -143.38(-154.43 – -133.24) |
| FL165 | 35.15(33.81 – 36.44) | FL565 | -33.65(-43.47 – -25.09) |
| FL170 | 35.8(34.47 – 37.07) | FL569 | -518.16(-542.84 – -495.32) |
| FL171 | 54.23(53.65 – 54.79) | FL573 | -65.03(-71.66 – -58.89) |
| FL172 | -48.01(-52.36 – -43.91) | FL576 | -19.01(-20.59 – -17.48) |
| FL278 | -202.87(-211.5 – -194.7) | FL582 | 0.51(-2.51 – 3.36) |
| FL283 | -18.02(-21.77 – -14.49) | FL584 | 38.57(37.06 – 40.01) |
| FL285 | 8.23(6.3 – 10.08) | FL585 | 52.08(50.96 – 53.14) |
| FL290 | 8.64(6.81 – 10.4) | FL585b | 68.3(67.16 – 69.37) |
| FL291 | -68.81(-71.65 – -66.08) | FL588 | -39.22(-43.05 – -35.59) |
| FL295 | -19.66(-21.5 – -17.88) | FL589 | -29.29(-31.34 – -27.3) |
| FL296 | -93.63(-101.81 – -86.08) | FL602 | 67.49(66.33 – 68.57) |
| FL297 | 12.48(10.82 – 14.08) | FL609 | 19.94(18.31 – 21.51) |
| FL298 | 15.32(13.43 – 17.14) | FL610 | 5.69(2.97 – 8.25) |
| FL304 | 20.92(19.11 – 22.66) | FL618 | -2.12(-3.46 – -0.81) |
| FL305 | 23.27(21.77 – 24.72) | FL633 | 11.69(10.39 – 12.95) |
| FL314 | 34.83(33.81 – 35.82) | FL634 | 16.19(14.65 – 17.66) |
| FL315 | -21.93(-23.54 – -20.37) | FL635 | -12.62(-15.3 – -10.06) |
| FL316 | 13.61(12.22 – 14.96) | FL636 | 20.93(19.24 – 22.54) |
| FL326 | 0.24(-1.71 – 2.13) | FL637 | -13.2(-15.39 – -11.08) |
| FL327 | -27.22(-29.22 – -25.27) | FL641 | 5.93(4.22 – 7.58) |
| FL328 | -10.04(-12.16 – -7.99) | FL650 | -39.83(-42.79 – -37) |
| FL332 | 13.38(10.24 – 16.31) | FL655 | 3.59(0.47 – 6.52) |
| FL333 | 11.09(9.85 – 12.31) | FL718 | -36.65(-40 – -33.46) |
| FL337 | -22.49(-28.93 – -16.65) | FL740 | -49.83(-53.04 – -46.75) |
| FL340 | 12.22(10.93 – 13.47) | FL742 | 0.11(-1.89 – 2.03) |
| FL341 | 37.13(36.16 – 38.07) | FL748 | 0.14(-2.06 – 2.25) |
| FL342 | 23.98(22.5 – 25.42) | FL765 | -9.13(-12.07 – -6.34) |
| FL343 | 58.65(58.04 – 59.23) | FL768 | -47.9(-51.37 – -44.59) |
| FL344 | 62.45(61.68 – 63.19) | FL787 | 50.96(48.7 – 53.04) |
| FL350 | 48.11(46.89 – 49.28) | FL803 | -145.5(-153.35 – -138.13) |
| FL353 | -33.19(-35.07 – -31.36) | FL804 | -67.1(-71.52 – -62.9) |
| FL354 | 22.1(20.43 – 23.7) | FL806 | -17.15(-20.37 – -14.1) |
| FL355 | 16.13(14.99 – 17.24) | FL807 | -38.38(-42.92 – -34.12) |
| FL356 | -15.02(-18.01 – -12.17) | FL826 | -24.47(-27.18 – -21.88) |
| FL357 | 47.25(46.19 – 48.26) | FL833 | -2.2(-4.62 – 0.12) |
| FL420 | -61.66(-66.57 – -57.03) | FL833b | 14.6(13.03 – 16.13) |
| FL421 | -54.04(-60.05 – -48.46) | FL837 | 80.29(79.66 – 80.88) |
| FL424 | 21.65(20.14 – 23.1) | FL94 | 5.12(3.62 – 6.57) |
| FL425 | -6.74(-9.54 – -4.09) | FL95 | -35.27(-38.61 – -32.08) |
| FL426 | -27.04(-29.51 – -24.66) | FL96 | 34.27(33.28 – 35.23) |
| FL517 | -25.93(-28.55 – -23.42) | FL97 | 19.17(17.7 – 20.59) |
| FL523 | -2.88(-5.87 – -0.05) | FL98 | 34(32.66 – 35.3) |

Table 27: Intervention efficacy of *Wolbachia* interventions in reducing female *Aedes albopictus* aggregated by sector across their respective intervention time

##### 6.4 Estimated intervention efficacies on *Aedes aegypti* abundance in non-release, adjacent sectors by event time for best model

| **Time period** | Intervention Efficacies (Time from start of intervention) |
|---|---|
| 1 - 6 months | 18% (16.21% – 19.71%) |
| 7 - 12 months | 47.32% (46.04% – 48.54%) |
| 13 - 18 months | 60.56% (59.64% – 61.43%) |
| 19 - 24 months | 77.15% (76.54% – 77.74%) |
| > 24 months | 79.35% (78.65% – 80%) |
| **Time period** | Intervention Efficacies (Calendar time) |
| 2020 EW 3 – 26 | -1.64% (-3.8% – 0.44%) |
| 2020 EW 27 – 52 | 29.77% (27.97% – 31.48%) |
| 2021 EW 1 – 26 | 42.68% (41.39% – 43.92%) |
| 2021 EW 27 – 52 | 63.81% (63.01% – 64.59%) |
| 2022 EW 1 – 26 | 71.44% (70.64% – 72.2%) |

Table 28: Spillover intervention efficacy of *Wolbachia* interventions in reducing female *Aedes aegypti* abundance aggregated by sector, calendar time and time from start of intervention
